# Multimorbidity Profiles in Patient Population from Central China: A Study Based on Electronic Health Records

**DOI:** 10.1101/2025.10.11.25337808

**Authors:** Weihao Shao, Yue Zhang, Yunyuan Kong, Enying Gong, Xiaoxia Wei, Xuejun Yin, Maigeng Zhou, Jiajuan Yang, Chi Hu, Xunliang Tong, Luzhao Feng, Zuolin Lu, Yaolong Chen, Ruitai Shao, Chen Wang

## Abstract

Comprehensive, life-course multimorbidity data derived from linked outpatient and inpatient electronic health records (EHRs) remain scarce globally. We analyzed integrated EHRs (2016-2023) from approximately 3.2 million individuals in Yichang, a prefecture-level city in Central China, to characterize lifetime disease co-occurrence by identifying both the most frequent combinations and significant non-random associations across all ages. Multimorbidity was defined as the presence of ≥ 2 distinct lifetime conditions. We identified the 50 most common disease triads and constructed disease networks using partial correlation analysis, ranking hub conditions with the Multimorbidity Coefficient (MMC). Overall, 74.5% of the population experienced multimorbidity (mean 5.29 conditions; women 5.59, men 4.98), with the burden rising steeply with age. Triad analysis revealed a clear life-course trajectory, beginning with respiratory clusters in childhood and diverging by sex in young adulthood, female gynecological versus male musculoskeletal/urological clusters, followed by cardiometabolic and cardiovascular dominance in mid-to-late life. Gastritis (K29) and sleep disorders (G47) were notably frequent components in adult triads. Network analysis identified K29, heart failure (I50), hypoproteinaemia (E88), anaemia (D64), and dermatitis (L30) as the top five hubs. Hub importance also varied by sex, with conditions such as osteoporosis (M81) being more central for women and benign prostatic hyperplasia (N40) for men. This study details a high lifetime multimorbidity burden and reveals a distinctive architecture characterized by a diverse, multi-system core where digestive, cardiometabolic, and systemic conditions co-dominate. Mapping these constellations provides critical insights for clinical anticipation, public health prevention, and research into shared pathways.

Graphical abstract

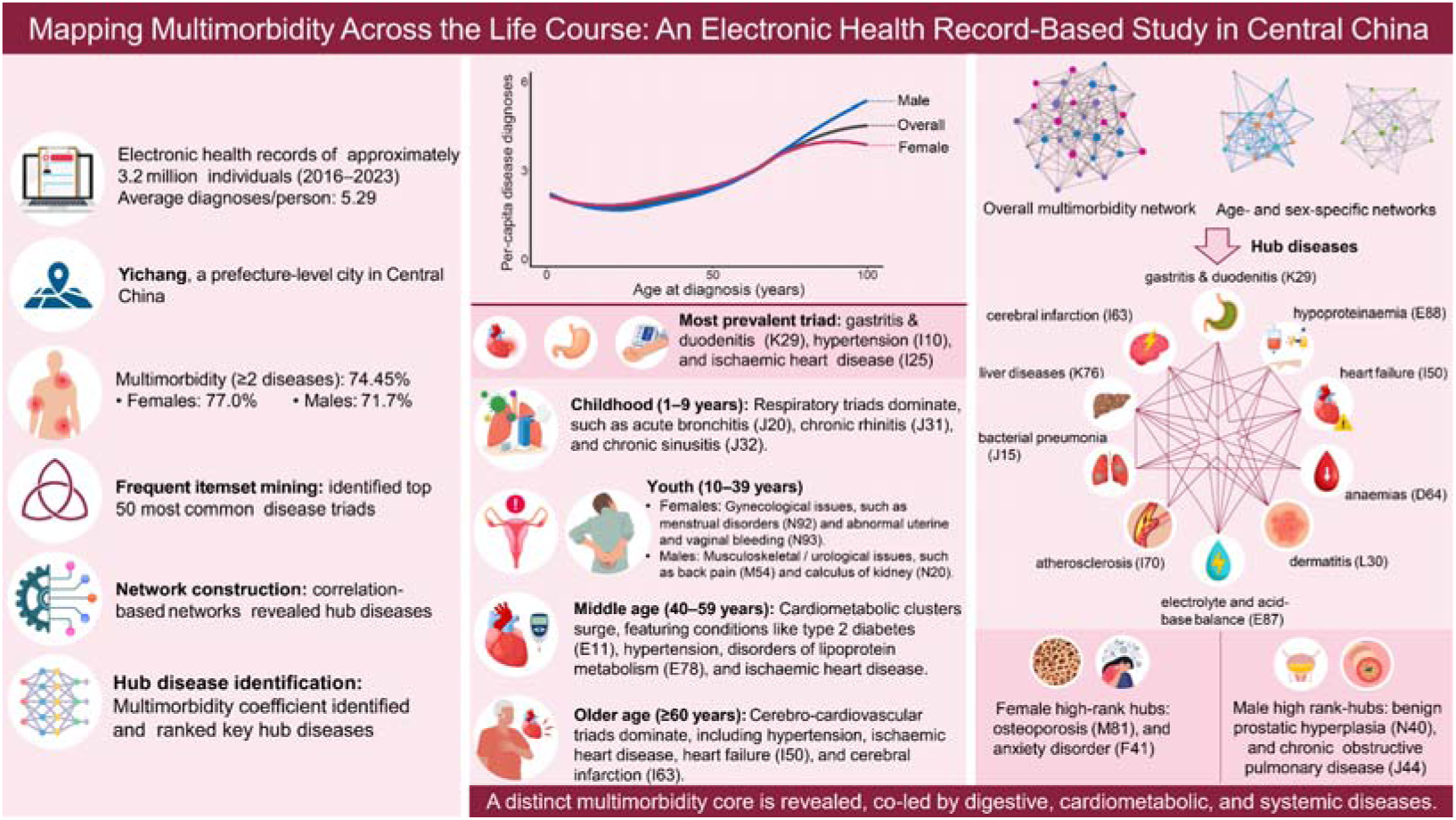

## 1. Introduction

Multimorbidity, defined as the co-occurrence of two or more chronic conditions within an individual, has become a significant concern in the healthcare system due to its association with diminished quality of life, functional impairments, and increased mortality risk^[1, 2]^. Furthermore, individuals with multimorbidity often experience greater healthcare utilization, including higher medical expenses, prolonged hospital stays, and more frequent medical consultations^[3]^.

Addressing this growing challenge requires a deeper understanding of multimorbidity. Therefore, the core research objectives focus on identifying common patterns or “constellations” of coexisting diseases, unraveling the mechanisms that drive their concurrence, and ultimately using this knowledge to develop targeted intervention and prevention strategies. The emergence of multimorbidity often stems from non-random associations between diseases. To achieve a mechanistic understanding of disease co-occurrence, it is essential to broaden the definition of multimorbidity to encompass any two or more diseases that an individual has experienced over their lifetime, regardless of whether they are chronic^[4]^. This expanded perspective is crucial for understanding non-random disease co-occurrences, as causal links may be temporally separated, and acute, treatable, or even self-limiting diseases can produce long-term consequences^[4, 5]^.

Previous research has investigated the core diseases within multimorbidity networks as well as multimorbidity patterns. However, many of these studies have relied on predefined lists of chronic diseases or expert consensus, and some have used self-reported data, which may lack accuracy^[6]^. Additionally, the study populations have frequently been limited to hospitalized or middle-aged and elderly individuals^[7–9]^, limiting the exploration of a broader spectrum of diseases and more diverse populations.

To address these gaps, we analyzed a comprehensive administrative dataset comprising outpatient and inpatient records from a city in Central China. By examining the most prevalent disease combinations and non-random associations within this population, we aimed to uncover the characteristics of multimorbidity among patients seeking medical care in this region. This approach leverages real-world data to provide insights into the complex interplay of diseases, thereby supporting the development of targeted interventions and informing healthcare policy to improve patient outcomes.

## 2. Methods

### 2.1 Data Source and Study Population

This retrospective population-based study utilized electronic health records (EH R) from Yichang City, Hubei Province, located in Central China. Yichang is a prefecture-level city, ranking within the top 25% of Chinese cities in terms of socioeconomic development level (http://www.yichang.gov.cn/zfxxgk/list.html?de pid=877&catid=150). The dataset comprised administrative health information, including both outpatient and inpatient encounters, from 462 secondary and tertiary hospitals within the city, covering the period from January 1, 2016, to Dec ember 31, 2023. The study population included all individuals with at least on e medical encounter during this period. Individuals whose only records pertained to routine health check-ups were excluded.

### 2.2 Health Condition Selection and Coding

All health conditions were coded using the International Classification of Diseases, Tenth Revision (ICD-10)^[10]^. Three–digit ICD-10 codes were used to specify diagnoses. To focus the analysis on diagnosed disease conditions, we excluded records primarily corresponding to the following ICD-10 chapters: Chapter XV (Pregnancy, childbirth and the puerperium; O00–O99), Chapter XVIII (Symptoms, signs and abnormal clinical and laboratory findings, not elsewhere classified; R00–R99), Chapter XX (External causes of morbidity and mortality; V00–Y98), Chapter XXI (Factors influencing health status and contact with health services; Z00–Z99), and Chapter XXII (Codes for special purposes; U00–U99).

### 2.3 Definition of Multimorbidity

Following established approaches designed to capture the cumulative burden of disease and potential causal pathways over time, multimorbidity was defined as the presence of two or more distinct health conditions (excluding those specified above) recorded for an individual at any point during the study period^[4, 5]^.

### 2.4 Analysis of Non-random Disease Associations and Network Construction

Network analysis was used to investigate non-random patterns of disease co-occurrence beyond chance. Prior to constructing the network, health conditions with a prevalence below 0.02% in the study population were excluded to focus on more common conditions and ensure robust estimates of association. Pairwise associations among the remaining conditions were quantified using partial correlation analysis, which statistically controls for the influence of other included conditions (Supplementary Methods, Section S1 online). A positive partial correlation coefficient indicated that two conditions co-occurred more frequently within the same individual than expected by chance, with the magnitude of the coefficient reflecting the strength of this non-random association.

Disease co-occurrence network was then constructed, in which nodes represented the included health conditions (defined by ICD-10 codes). An edge connecting two conditions was included in the final network only if the partial correlation coefficient was positive and the associated *P*-value remained statistically significant (*P* < 0.05) after adjustment for multiple comparisons using the Benjamini-Hochberg (BH) procedure to control the false discovery rate (FDR)^[11]^.

To identify the most central conditions within this network, we applied the Multimorbidity Coefficient (MMC)^[5, 12]^ (Supplementary Methods, Section S2 online). This approach was selected over simpler metrics such as degree (the number of connections) because MMC accounts for both the number and strength of non-random co-occurrences, providing a more nuanced measure of a condition’s cumulative positive association burden. Specifically, the MMC for a given condition was calculated as the sum of all positive partial correlation coefficients (φ >0) linking it to any other condition in the analysis, regardless of statistical significance. Conditions with the highest MMC values were designated as core conditions within the multimorbidity network.

### 2.5 Statistical Analysis

The age of the study population was summarized as mean ± standard deviation (SD). We calculated age-specific per-capita disease diagnoses to quantify diagnostic burden. Network analysis, including partial correlation and the Multimorbidity Coefficient (MMC), was applied to identify non-random disease associations and core conditions as described above. In addition, frequent itemset mining (Apriori algorithm) was used to identify the top 50 most prevalent disease triads, which were reported with prevalence and 95% confidence intervals, overall and stratified by sex and predefined age groups. To assess the robustness of our network findings, we performed sensitivity analyses by varying the statistical thresholds for network edge inclusion. These analyses confirmed the stability of the identified main hub conditions (Supplementary Methods, Section S3 online). All analyses were conducted using R software (Version 4.4.3, R Foundation for Statistical Computing, Vienna, Austria.) and relevant packages.

### 2.6 Ethical Considerations

This study was conducted in accordance with the Declaration of Helsinki and approved by ethical approval from the Ethics Committee of the School of Population Medicine and Public Health, Chinese Academy of Medical Sciences & Peking Union Medical College (Approval No: CAMS&PUMC-IEC-2022-076). The study used fully anonymized administrative EHR data. As the data were de-identified and analyzed retrospectively for secondary purposes, the requirement for individual informed consent was waived by the Ethics Committee. Patient confidentiality was maintained throughout the study.

## 3. Results

This study analyzed EHRs collected between January 1, 2016 and December 31, 2023, and identified 3,184,662 unique individuals. The population included 1,648,711 females (51.77%) and 1,535,951 males (48.23%). The overall mean age was 44.79 ± 20.67 years. Patient encounters were recorded in both inpatient and outpatient settings; 1,042,439 individuals (32.73%) had at least one inpatient record and 3,043,405 (95.56%) had at least one outpatient record during the observation window (note: these settings are not mutually exclusive). Based on the unique conditions documented per person during the study period, the mean number of diagnosed conditions was 5.29 ± 6.00. Furthermore, 2,371,063 individuals (74.45% overall; 77.02% of females, 71.70% of males) had two or more unique conditions recorded during this timeframe (Table 1).

**Table 1.** Sociodemographic characteristics of the patients.

|  | Population (%) | Age, years | Number of diagnosed conditions | Number of individuals with two or more diagnosed conditions (%) |
| --- | --- | --- | --- | --- |
| All | 3,184,662 (100) | 44.79±20.67 | 5.29±6.00 | 2371063 (74.45) |
| Sex |  |  |  |  |
| Female | 1,648,711 (51.77) | 44.60±20.48 | 5.59±6.08 | 1,269,801 (77.02) |
| Male | 1,535,951 (48.23) | 45.01±20.87 | 4.98±5.91 | 1,101,262 (71.70) |
| Age groups, males and females (years) |  |  |  |  |
| 1~9 | 217,765 (6.84) | 4.70±2.55 | 3.97±3.38 | 161,269 (74.06) |
| 10~19 | 212,386 (6.67) | 14.89±2.96 | 2.88±2.79 | 129,011 (60.74) |
| 20~29 | 361,375 (11.35) | 24.88±2.88 | 3.94±4.11 | 243,311 (67.33) |
| 30~39 | 427,350 (13.42) | 34.38±2.85 | 4.23±4.44 | 297,038 (69.51) |
| 40~49 | 496,115 (15.58) | 44.87±2.87 | 5.24±5.48 | 373,627 (75.31) |
| 50~59 | 634,963 (19.94) | 54.36±2.83 | 5.43±5.84 | 485,603 (76.48) |
| 60~69 | 483,507 (15.18) | 64.37±2.84 | 6.84±7.34 | 391,589 (80.99) |
| 70~79 | 255,272 (8.02) | 73.71±2.80 | 8.00±8.66 | 212,233 (83.14) |
| $\geq 80$ | 95,929 (3.01) | 84.12±3.53 | 7.81±9.08 | 77,382 (80.67) |
| Ethnicity |  |  |  |  |
| Han | 2,866,258 (90.00) | 45.10±20.60 | 5.30±6.07 | 2,123,363 (74.08) |
| Minority | 318,404 (10.00) | 42.04±21.14 | 5.26±5.36 | 247,700 (77.79) |
| Diagnostic scenario<sup>a</sup> |  |  |  |  |
| Inpatient | 1,042,439 (32.73) | 51.80±20.79 | 4.36±5.26 | 684,798 (65.69) |
| Outpatient | 3,043,405 (95.56) | 44.41±20.48 | 4.15±4.24 | 2,165,631 (71.16) |

<sup>a</sup> Percentages for Diagnostic scenario represent the proportion of the total study population ( $n = 3,184,662$ ) with at least one record from the respective setting; categories are not mutually exclusive.

The burden of diagnosed conditions increased substantially with age (Figs. 1 – 2). The overall average number of unique conditions recorded per person during the 2016–2023 study period was 5.73; females (6.03) averaged more conditions than males (5.41). Circulatory system diseases (Chapter 9) contributed most to the average per-capita diagnoses overall (0.97 diagnoses/person) and particularly in males (1.03 diagnoses/person, 1.13 times the female contribution). Females, conversely, had markedly higher contributions from musculoskeletal system diseases (Chapter 13; 0.84 diagnoses/person, 1.2 times the male contribution) and genitourinary diseases (Chapter 14; 0.77 vs. 0.39 diagnoses/person) (Fig. 1).

**Fig. 1.**
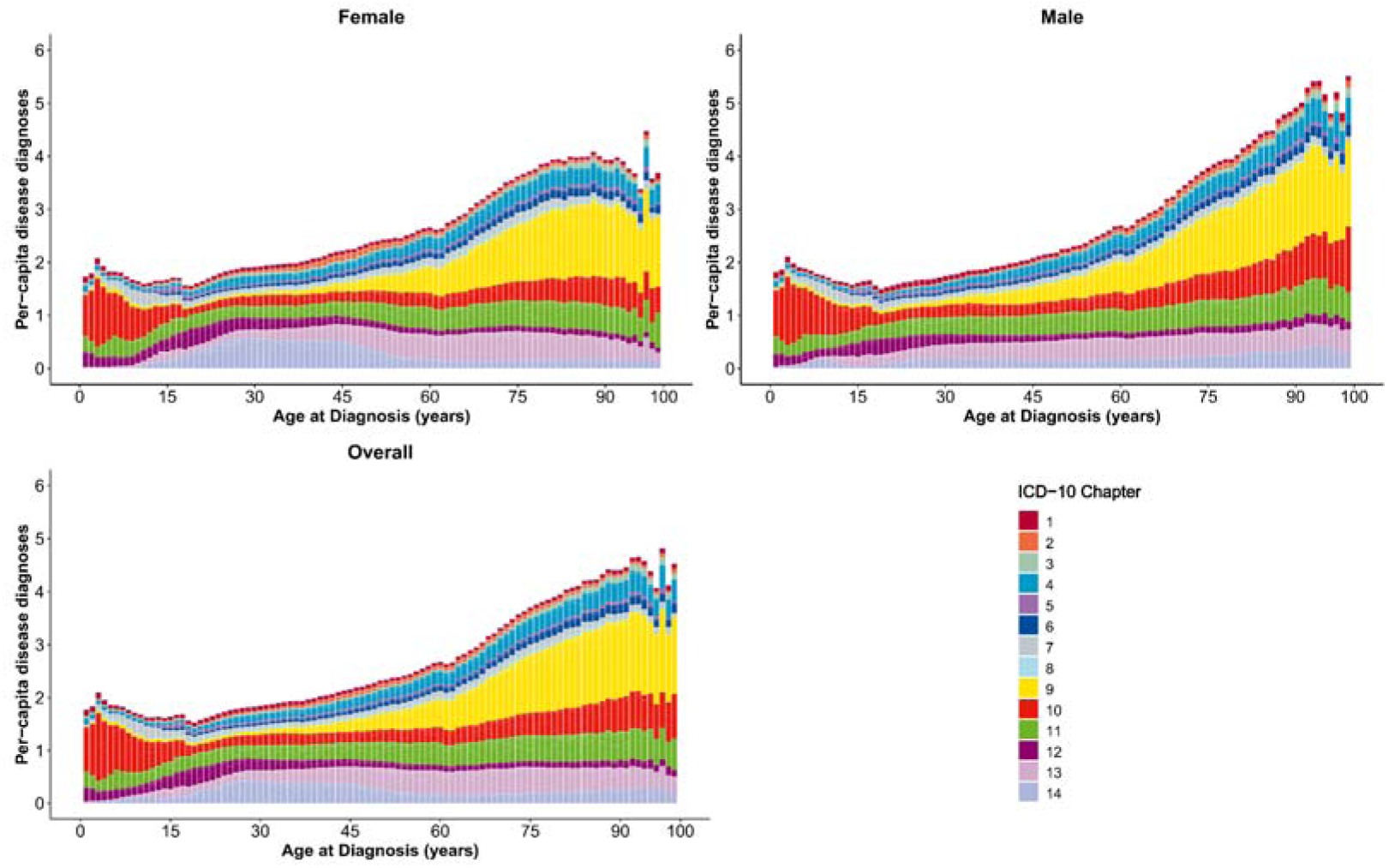
Stacked bar chart of per-capita disease diagnoses according to ICD-10 among patients. The x-axis represents age (from 1 to 100 years, unit: years), and the y-axis represents the per-capita disease diagnoses. The color of each bar represents disease systematic chapters.

**Fig. 2.**
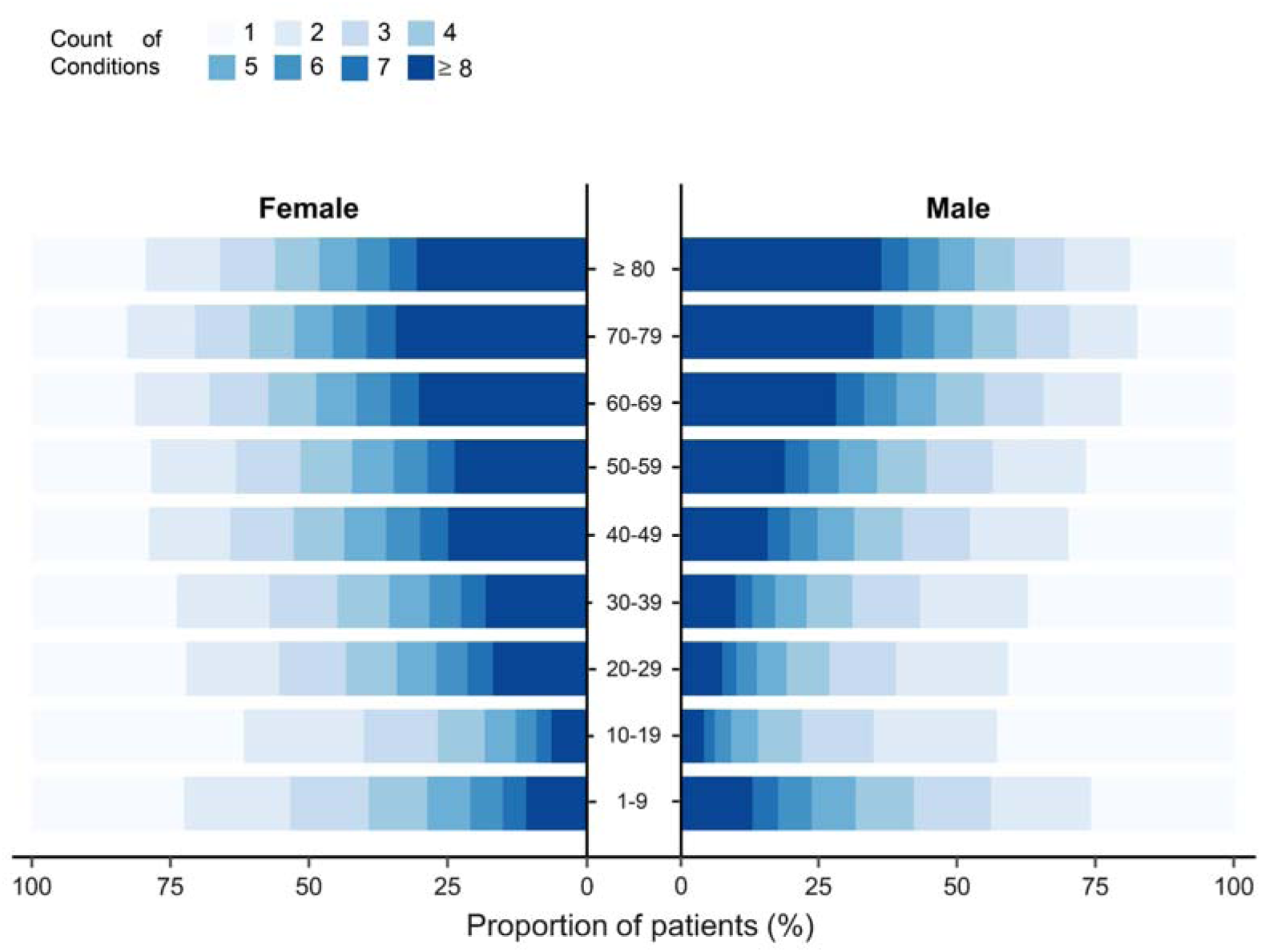
Proportion of patients by number of unique conditions recorded during the 2016-2023 study period, stratified by sex and age group. Note that observed patterns in younger age groups are influenced by the study’s time window.

Consistent with the rising average burden, the prevalence of multimorbidity (≥2 unique conditions recorded) increased sharply across age groups, with the vast majority of older adults (≥80 years) having multiple conditions (Fig. 2). Crucially, the apparent pattern of higher condition counts in the 1–9 year age group compared to the 10–19 year group is likely an artifact resulting from the limited 2016–2023 observation window (Fig. 2). Left-censoring of pre-2016 diagnoses for older children prevents a reliable comparison of cumulative condition counts between these specific youngest age groups. Females exhibited a higher prevalence of multimorbidity (≥2 conditions) than males across most adult age groups (20–79 years), although this pattern reversed in the oldest age group (≥80 years: 79.9% female vs 81.6% male) (Fig. 2).

### 3.1 Common Disease Triads Across the Life Course

Analysis of the 50 most frequent disease triads revealed distinct patterns across age groups (Tables S1–S12 online). Overall, {I10 (essential hypertension), I25 (chronic ischaemic heart disease), K29 (gastritis and duodenitis)} was the most prevalent triad (2.34%; Table S1 online), ranking highest in both sexes (Tables S2–S3 online). Age-stratified analysis showed a clear life-course trajectory with notable sex differences, visualized by the changing frequency of conditions within top triads across age groups (Fig. 3; Figs. S1–S2 online).

**Fig. 3.**
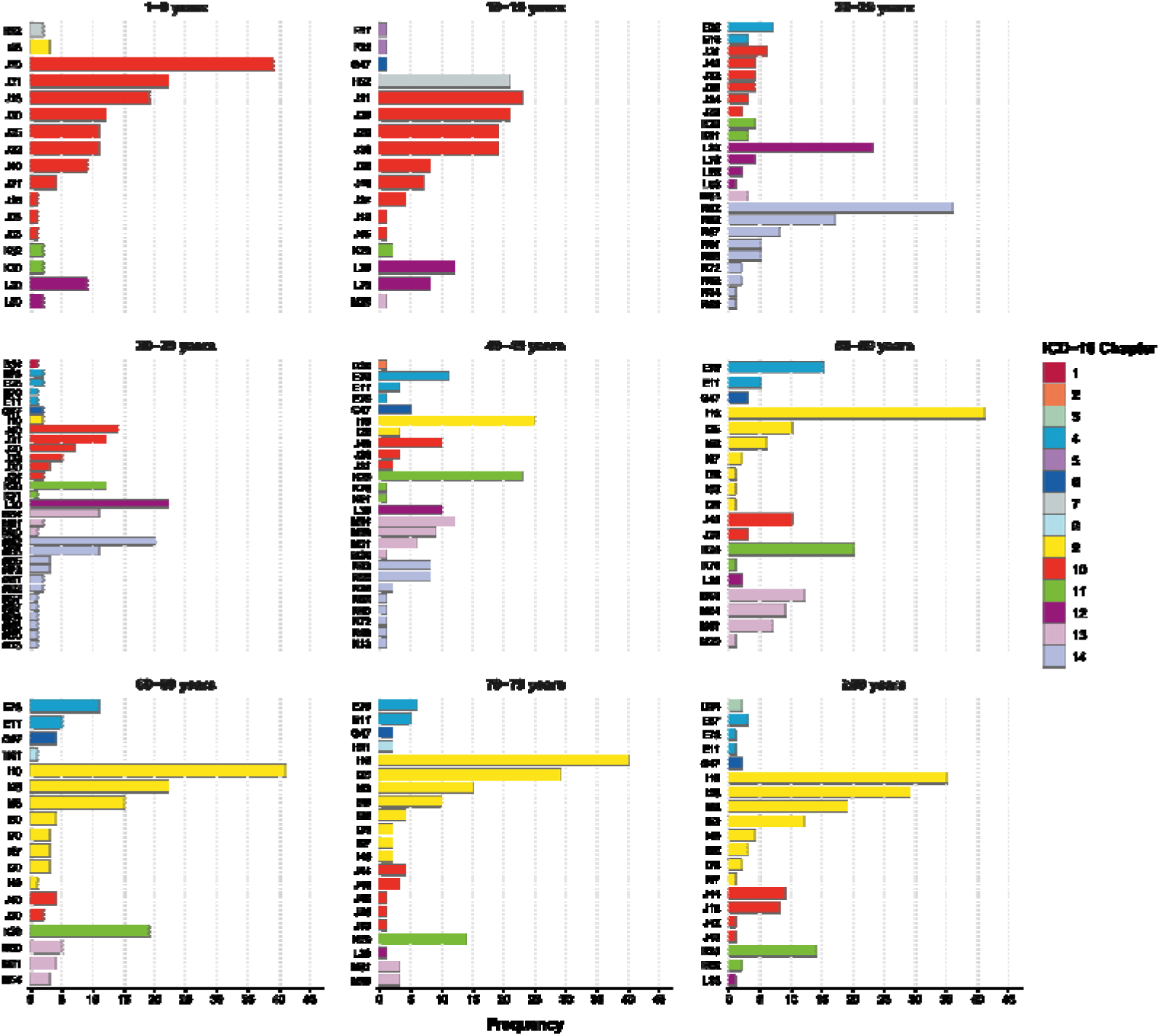
Number of times a health condition occurs in the 50 most common triads in the overall population. The graph shows the number of times a health condition occurs in the 50 most common triads by selected age groups. The x-axis shows the frequency of occurrence. For clarity, diseases are represented by their ICD-10 codes; a complete list of codes and their corresponding full disease names is provided in Table S25 online. An interactive version of this figure, with features for exploring full disease details, is available online at: https://pumc-multimorbidity.shinyapps.io/triad-frequency-visualization/

Respiratory conditions dominated childhood triads in both sexes (Table. S4 online). In adolescence (10-19 years), alongside common upper respiratory issues (e.g., J30–J32) and (H52) refractive errors, females distinctively showed early emergence of gynecological triads (involving N92 Excessive/frequent menstruation), while mental health combinations also appeared, more prominently in females (Table S5 online; Figs. S1–S2 online). Young adulthood (20–39 years) showed marked sex divergence: female top triads were dominated by gynaecological issues, notably N92 (excessive, frequent and irregular menstruation) and N93 (other abnormal uterine and vaginal bleeding), whereas male triads more often featured musculoskeletal and urological combinations, notably M54 (back pain), N13 (obstructive uropathy), and N20 (calculus of kidney) (Tables S6–S7; Figs. S1–S2 online).

A significant shift occurred in middle age (40–59 years) with the rise of cardiometabolic clusters {E11 (type 2 diabetes mellitus), E78 (disorders of lipoprotein metabolism), I10, I25} in both sexes (Tables S8–S9 online). However, male triads showed clearer cardiometabolic/cardiovascular focus, while female triads frequently involved persistent gynecological issues, prominent musculoskeletal conditions, particularly (M50/M51) cervical disorders, and (K29) gastritis/duodenitis alongside cardiometabolic components (Tables S8–S9 online; Figs. S1–S2 online). This trend is evident, with I10 frequency rising from age ≥40 years and E78 peaking at 50–59 years (Fig. 3; Figs. S1–S2 online).

Cardiovascular triads {I10, I25, I50 (heart failure), I63 (cerebral infarction)} became overwhelmingly dominant in older adults (≥60 years) for both sexes (Tables S10-S12 online), with (I10, I25, I50) highly prevalent in the group aged 80 years and older (Table S12 online). Sex differences remained: (K29) gastritis/duodenitis and (M50/M51) cervical issues were more frequent components in female triads, while (J44) Chronic Obstructive Pulmonary Disease (COPD) and (N40) Benign Prostatic Hyperplasia (BPH) were prominent in male triads (Tables S10–S12 online; Figs. S1–S2 online). K29 was a notably frequent component across most adult age groups, particularly prominent in female triads from middle age onwards, while (G47) sleep disorders also featured consistently in both sexes (Fig. 3; Figs. S1–S2 online).

### 3.2 Disease Co-occurrence Network Analysis

Disease co-occurrence networks were constructed based on significant positive partial correlations (FDR-adjusted *P* < 0.05), controlling for the influence of other conditions.

Comprehensive networks including edges with a partial correlation coefficient (φ) ≥ 0.01 and FDR-adjusted *P* < 0.05 were generated for the overall population (545 nodes, 4,175 edges), females (532 nodes, 4,134 edges), and males (512 nodes, 3,976 edges). Detailed visualizations of these networks, along with versions filtered using stricter significance levels (FDR-adjusted *P* < 0.01) or higher correlation thresholds (φ ≥ 0.05 or φ ≥ 0.1) to illustrate the impact on network density and highlight stronger associations (Figs. S3–S5 online). For clarity in the main text, the overall population network is presented filtered for associations with φ ≥ 0.05 and FDR-adjusted *P* < 0.05 (Fig. 4). Comparing the comprehensive networks reveals minor differences in overall size, with the female network encompassing slightly more diseases and associations than the male network within this population (Fig. S3–S5 online). The network structures evolved significantly across the life course, as illustrated in visualizations constructed for distinct age groups (using φ ≥ 0.05) (Fig. S6 online).

**Fig. 4.**
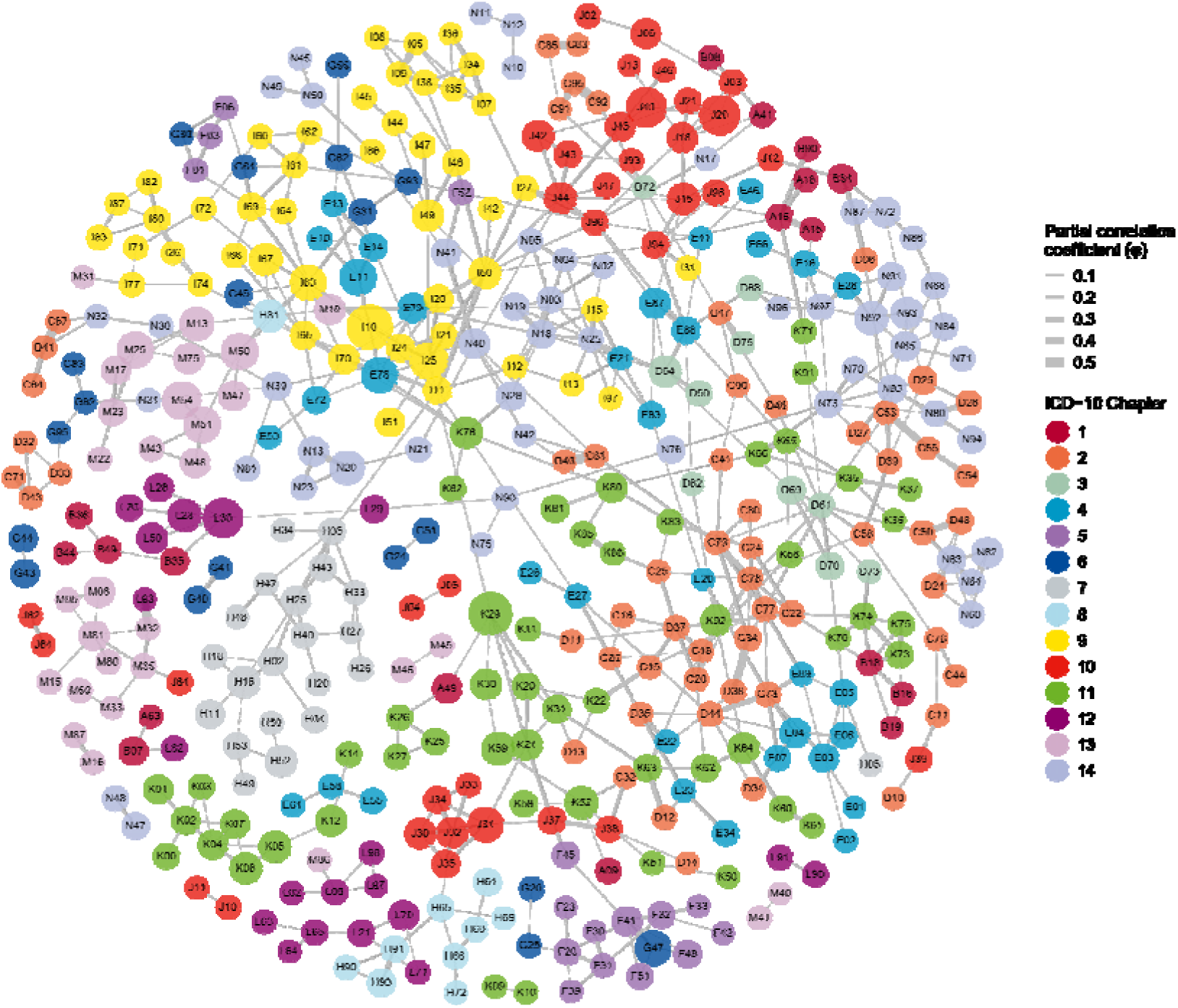
Multimorbidity network in overall population. Nodes represent individual ICD-10 conditions, with size proportional to prevalence and color denoting the ICD-10 chapter. Edges depict statistically significant (FDR-adjusted *P* < 0.05) positive partial correlations (φ ≥ 0.05) between conditions, controlling for all other diseases. Edge width is scaled to the magnitude of φ. For a full list of codes and names, see Table S25 online. An interactive version of this network is available at: https://pumc-multimorbidity.shinyapps.io/multimorbidity-network/

In the overall population, network analysis identified K29, I50, E88 (hypoproteinaemia), D64 (other anaemias), and L30 (other dermatitis) as the top five hub conditions (Figs. 4–5). A comprehensive list of the top 100 hub conditions for the overall population is provided separately (Table S13 online). Among the top five, K29 (rank = 1, MMC = 2.47), the leading digestive hub, showed strong associations (φ ≥ 0.05) with K21 (gastro-oesophageal reflux disease; φ = 0.098), K31 (other diseases of stomach/duodenum; φ = 0.089), and K20 (oesophagitis; φ = 0.084). E88 (rank = 2, MMC = 2.37), a key metabolic hub, connected strongly with E87 (φ = 0.130), J94 (other pleural conditions; φ = 0.106) and J96 (respiratory failure; φ = 0.081). I50 (rank = 3, MMC = 2.30), the top circulatory hub, was strongly linked to I11 (hypertensive heart disease; φ = 0.209), I20 (angina pectoris; φ = 0.189), and I48 (atrial fibrillation/flutter; φ = 0.141). Other top 5 hubs included D64 (rank = 4, MMC = 2.26), primarily linked to D50 (iron deficiency anaemia; φ = 0.159) and E88 (φ = 0.149), and L30 (rank = 5, MMC = 2.21), associated with L23 (allergic contact dermatitis; φ = 0.074) and L28 (lichen simplex chronicus; φ = 0.073). To further illustrate the local connectivity patterns, we visualized the subnetworks for each of the top 20 hub conditions in the overall population (Fig. S7 online; an interactive version is available at: https://pumc-multimorbidity.shinyapps.io/hub-disease-network/).

**Fig. 5.**
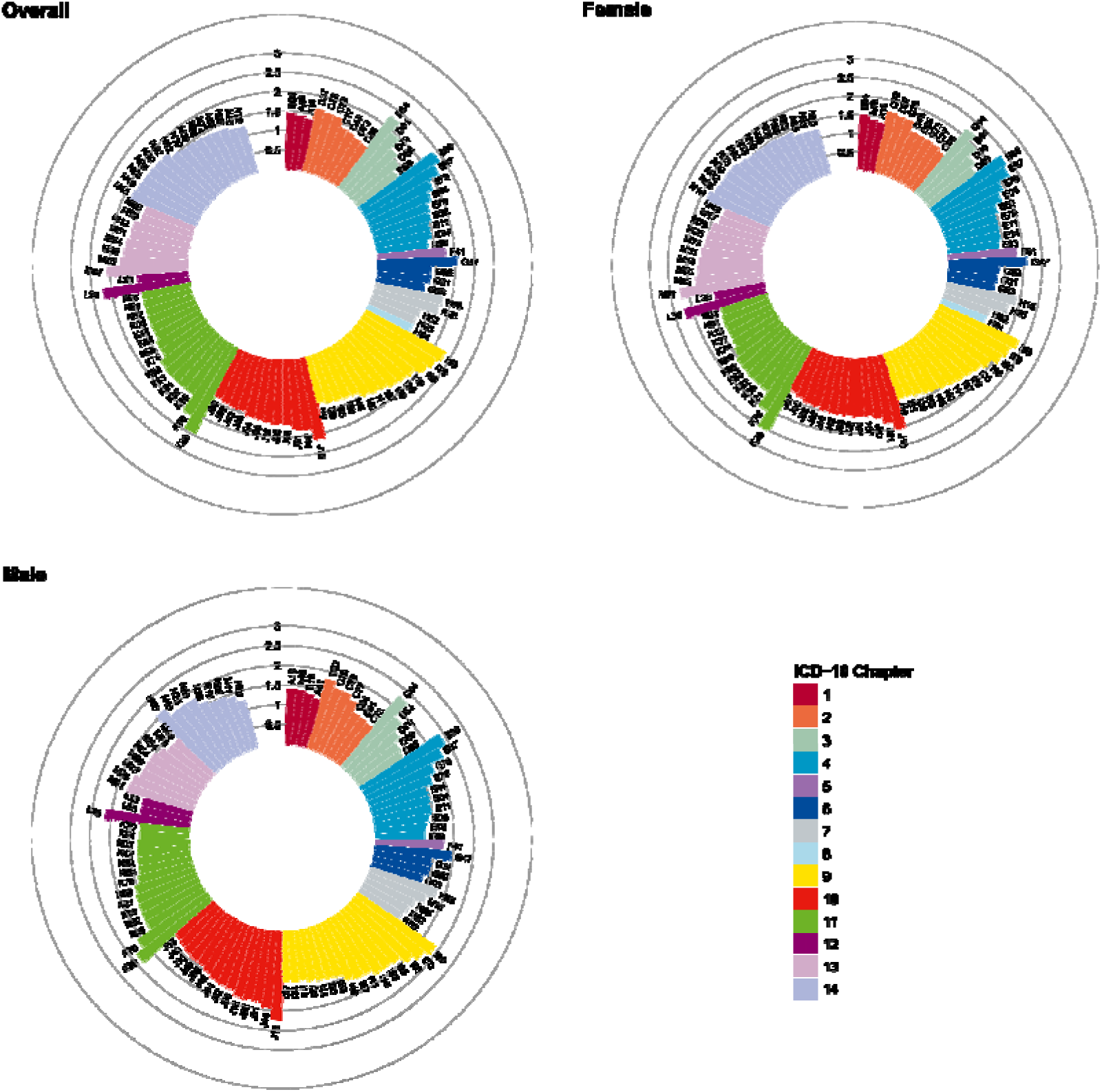
Multimorbidity Coefficient (MMC) of the top 100 conditions in the overall population, females, and males. The MMC quantifies a condition’s centrality in the disease network by summing the strength of all its positive partial correlations (φ > 0) with all other conditions, after controlling for confounders. A higher MMC (longer bar) indicates a more influential hub disease, distinct from simple co-occurrence frequency or degree centrality. Bar color corresponds to the ICD-10 chapter. A complete list of codes and names is in Table S25 online. An interactive version of this figure, which includes a tool to explore the robustness of these rankings via sensitivity analysis, is available at: https://pumc-multimorbidity.shinyapps.io/mmc-visualization/

Sex-stratified analyses revealed distinct patterns in network structure and hub conditions (Fig. 5; Tables S14–S15 online; Fig. S8–S9 online). While K29 (rank = 1, MMC = 2.53) and E88 (rank = 2, MMC = 2.32) were top hubs in females, E88 (rank = 1, MMC = 2.37) and I50 (rank = 2, MMC = 2.37) ranked highest in males, with I50 strongly associated with I11 (φ = 0.219) and I20 (φ = 0.181). Among females, M81 (osteoporosis; rank = 5, MMC = 2.22) was a prominent musculoskeletal hub, strongly linked to M80 (osteoporosis with fracture; φ = 0.135). F41 (other anxiety disorders; rank = 19, MMC = 1.82) also featured as a hub, connected to G47 (sleep disorders; φ = 0.196) and F32 (depressive episode; φ = 0.186) (Tables S14 online). Key hubs more prominent in males included N40 (rank = 11, MMC = 2.05), strongly associated with N41 (prostatitis; φ = 0.164) and N39 (other urinary disorders; φ = 0.119). Respiratory hubs like J44 (rank = 12, MMC = 1.97) and the sequelae hub I69 (sequelae of cerebrovascular disease; rank = 17, MMC = 1.82) were also notable in males, with J44 linked to I27 (other pulmonary heart diseases; φ = 0.153) and J43 (emphysema; φ = 0.152), and I69 linked to G81 (hemiplegia; φ = 0.189) (Table S15 online).

Network structures and their corresponding hub diseases evolved significantly across the life course (Fig. S6 online). In childhood (1–9 years), hubs were dominated by respiratory conditions such as J31 (rhinitis). By middle age (40–49 years), K29 emerged as the primary hub, alongside cardiovascular diseases like I50. In late life (≥80 years), the landscape shifted to chronic and degenerative diseases, with I70 (atherosclerosis), M81, and I69 becoming top hubs (Tables S16–S24 online).

## 4. Discussion

This study, based on EHRs from nearly 3.2 million individuals who accessed healthcare in Yichang City between 2016 and 2023, systematically characterized health condition co-occurrence across the life course in this Central China region. We found that 74.45% of individuals were recorded with two or more distinct health conditions over their lifetime (excluding records from specific ICD-10 chapters). This proportion increased significantly with age. We examined condition co-occurrence patterns from two perspectives of frequency (common combinations) and non-random association (network structure).

The results showed that females had a slightly higher mean number of diagnosed conditions (5.59) compared with males (4.98), and the overall health burden escalated throughout the life course. This is consistent with evidence that multimorbidity prevalence is higher among women, reflecting both biological susceptibility (e.g., autoimmune and osteo-metabolic conditions) and systemic factors such as more frequent healthcare utilization, which can increase ascertainment. Practically, this supports sex-responsive case-finding strategies, such as integrating anaemia and osteoporosis screening for women presenting with cardiometabolic diseases^[13, 14]^. These findings highlight that multimorbidity a prevalent public health phenomenon in this region, particularly within the context of China’s rapidly aging population ^[15]^.The proportion of individuals with two or more health conditions in our study (74.45%) was considerably higher than estimates from some Chinese studies based on predefined chronic disease lists or self-reported data^[16, 17]^, which may underestimate the burden due to methodological limitations^[18]^. However, considering our broad definition encompassing any lifetime health condition and the comprehensiveness of EHR data^[19]^, our estimates are more comparable to those from UK studies using similar EHR-based approaches despite differences in population structure and healthcare systems^[5, 20, 21]^.

Our finding that gastritis/duodenitis (K29) was the top-ranked hub is particularly noteworthy. This aligns with a study comparing inpatients in Shaanxi, China, with those in the UK, which also identified gastritis and duodenitis as a shared core condition, suggesting its importance may extend beyond our study region^[8]^. Importantly, the prominence of these hubs (notably gastritis, hypoproteinaemia (E88), heart failure (I50), and other anaemias (D64) remained stable across sensitivity analyses using stricter statistical thresholds, confirming the robustness of our findings.

Frequent itemset mining revealed distinct life-course patterns in disease combinations: respiratory conditions predominated in childhood; genitourinary and musculoskeletal intersections emerged in young adulthood; cardiometabolic clusters, including type 2 diabetes mellitus (E11), disorders of lipoprotein metabolism (E78), essential hypertension (I10), and chronic ischaemic heart disease (I25), increased markedly in middle age; and cardiovascular combinations, such as essential hypertension (I10), chronic ischaemic heart disease (I25), heart failure (I50), and cerebral infarction (I63),dominated in older age. This trajectory aligns with the chronic disease accumulation observed in longitudinal studies^[5, 22, 23]^ and corresponds with the biological logic of risk factor accumulation and physiological decline with aging^[24–26]^. Notably, gastritis and duodenitis and sleep disorders (G47) were frequent components of common triads across most adult age groups. Consistent with findings in other populations^[8]^, our results underscore the potential key role of gastritis and duodenitis as a central condition within disease co-occurrence networks.

The persistent frequency and centrality of gastritis and duodenitis and sleep disorders in adults warrants further discussion. The prominence of gastritis and duodenitis as the top hub may be driven by regional factors, including dietary habits, high prevalence of Helicobacter pylori infection, and psychosocial stress^[27, 28]^. Its hub role could indicate that it functions as a common soil for, or consequence of, multiple systemic conditions, explaining its broad connectivity. Similarly, the centrality of sleep disorders likely reflects the complex, bidirectional relationship between sleep health and chronic health disease, where sleep disorders can be both consequence of illnesses (e.g., pain- or anxiety-induced insomnia) and risk factors exacerbating exacerbates cardiovascular, metabolic, and mental health issues^[29]^.

Network analysis identified gastritis and duodenitis, heart failure, hypoproteinaemia, other anaemias, and other dermatitis as the top five core nodes in the overall disease network. The prominence of these hubs, particularly gastritis and duodenitis and heart failure, is consistent with findings from other studies^[30]^. Identifying these core nodes provides further insights into the disease co-occurrence network: heart failure and its strong links to hypertensive heart disease (I11), angina pectoris (I20), and atrial fibrillation/flutter (I48) represent core of cardiovascular disease progression, while K29 and its neighbors, including gastro-oesophageal reflux disease (K21), other diseases of stomach/duodenum (K31), and oesophagitis (K20), form an upper gastrointestinal disorder cluster. Systematic comparison with prior research highlights the innovations. Previous large-scale studies in China have often relied on distinct data sources, such as national surveys (e.g., CHARLS), inpatient cohorts, or regional EHRs focused primarily on the elderly^[6, 31, 32]^. While valuable, these approaches have produced a somewhat fragmented perspective, consistently pointing to the dominance of a cardiometabolic core, with hypertension as a foundational “gateway” disease^[8, 32]^. By contrast, large-scale EHR studies in Western populations frequently identify two major clusters of comparable importance: cardiometabolic conditions and mental health disorders, with the latter often serving as precursors in younger adults^[5, 33]^.

Our study contributes uniquely to this landscape. Methodologically, the use of comprehensive, all-age, linked clinical EHRs from a large city in Central China helps to fill a critical data gap. More importantly, our findings reveal a distinct multimorbidity architecture. While confirming the centrality of cardiometabolic hubs such as heart failure our network analysis also elevates digestive and systemic conditions reflecting nutritional and inflammatory status, namely hypoproteinaemia and other anaemias, to a similar level of prominence. This diverse, multi-system core contrasts both with the cardiometabolic-dominant model common in Chinese studies and the cardiometabolic–mental health pattern prevalent in Western research, suggesting a unique, locally shaped health landscape.

Observed sex differences also align with prior research^[34, 35]^. For example, osteoporosis (M81) was more central in the female network, while BPH (N40) was more prominent in the male network. These findings reflect known sex-specific disease predilections. Anxiety disorders (F41) being more central in the female network aligns with the higher prevalence of anxiety among women globally^[36]^. COPD (J44) and sequelae of cerebrovascular disease (I69) were more central in the male network, potentially linked to differential risks such as smoking and occupational exposures^[37, 38]^. These sex-specific patterns are consistent with findings from other studies on sex-specific multimorbidity clusters^[39]^.

Strengths of this study include its large sample size, broad life course coverage, and integration of outpatient and inpatient EHR data, in contrast to studies that focus on narrower populations. We incorporated a wide range of health conditions and applied complementary analytical methods (frequency mining and non-random association analysis). The use of partial correlation in network analysis controlled for confounding, and the lifetime definition allowed capture of cumulative disease burden.

Certain limitations should be considered. First, the retrospective design and the nature of EHR data preclude establishing causal inference between conditions. Our analysis was cross-sectional within the 2016– 2023 observation window, and we could not determine the precise sequence of disease onset due to left-censoring of diagnoses made before 2016. This particularly affects estimates of cumulative lifetime burden in older individuals. Second, focusing on healthcare utilizers may introduce selection bias, potentially overestimating multimorbidity prevalence compared with the general population, which includes individuals who do not seek care. However, this approach is well-suited to characterizing the burden on the healthcare system. Third, reliance on EHRs introduces information bias from variable data quality and record completeness, although quality control measures are in place. Fourth, the single-city setting restricts the generalizability of these multimorbidity profiles to other regions with different demographics or healthcare systems. Finally, the EHRs did not systematically record socioeconomic status (e.g., income, education, occupation), urban-rural residence, or detailed lifestyle factors. This absence precluded analysis of socioeconomic gradients and urban–rural differences in multimorbidity, limiting mechanistic interpretation by preventing control for these potential confounders. Furthermore, coded categories such as hypoproteinaemia, other anaemias, and dermatitis are heterogeneous groupings; this, together with differences in healthcare utilization, may inflate their recorded centrality. They should therefore be interpreted as contextual “recorded” hubs rather than causal drivers.

Despite limitations, our findings hold considerable value. The identified age- and sex-specific multimorbidity patterns have implications across multiple domains: guiding proactive clinical practice, shaping public health strategies, informing health policy and setting agendas for future research. Understanding these multimorbidity constellations offers opportunities to mitigate adverse outcomes associated with common disease clusters.

For clinical practice, the findings support proactive, patient-centered care by enabling clinicians to anticipate and screen for age- and sex-specific comorbidities. Identification of hubs such as hypoproteinaemia and other anaemias should serve as a clinical alert, prompting comprehensive evaluation for underlying illness rather than isolated symptomatic treatment. This facilitates personalized management, such as integrating osteoporosis care for women and addressing urological symptoms for men alongside their other chronic conditions to improve quality of life.

For public health and health service organizations, the findings can guide targeted prevention programs and resource allocation toward high-impact clusters, such as cardiometabolic conditions in middle-aged populations. For medication management, the data highlight the need to move beyond simple polypharmacy counts to optimizing regimens by considering treatment synergies and potential adverse drug–disease interactions (e.g., cardiovascular disease with gastritis). Finally, these results provide a blueprint for organizing health services around common patient profiles, such as integrated clinics for heart failure (I50) and its web of associated conditions.

To build on this work and address its limitations, several future research directions are proposed. First, to overcome the constraints of retrospective design and establish causal pathways, prospective cohort studies are needed. Second, to enhance generalizability beyond a single city, multi-center studies across diverse Chinese regions (e.g., coastal, western, and northeastern cities) should compare multimorbidity architectures. Third, to address the absence of socioeconomic data, future work could, under strict ethical and privacy-preserving protocols, link EHRs with socioeconomic databases or incorporate validated instruments for collecting such data during clinical encounters. Finally, building on our network map, future research should develop and validate prognostic models that combine conventional risk factors with network-based features (e.g., the MMC) to improve risk stratification for outcomes such as hospitalization and mortality, advancing the field from mapping to forecasting.

In Conclusion, utilizing large-scale EHR data, this study systematically characterized health condition co-occurrence profiles across the life course in a large Central Chinese population. Analysis of frequent combinations and non-random associations revealed evolving age- and sex-specific disease patterns and a distinct multimorbidity architecture, characterized by a diverse, multi-system core co-dominated by digestive, cardiometabolic, and systemic conditions. These findings advance understanding of the regional health landscape and provide an, empirical foundation for optimizing healthcare services, shaping precise public health strategies, and guiding future research.

## Acknowledgments

This work was supported by the non-profit Central Research Institute Fund of Chinese Academy of Medical Sciences (2021-RC330-004); the Fundamental Research Fund for Central Public Welfare Research Institutes of the Chinese Academy of Medical Sciences (2022-ZHCH330-01); the Disciplines Construction Project: Population Medicine (WH10022022010); and the Special Research Fund for Central Universities, Peking Union Medical College (3332025142). We gratefully acknowledge the support and collaboration of the Yichang Center for Disease Control and Prevention, whose assistance in data access and logistical coordination made this study possible.

## Author contributions

Ruitai Shao and Weihao Shao conceived and designed the study, with Ruitai Shao and Weihao Shao securing the funding; Weihao Shao conducted the data analysis and drafted the manuscript; Yaolong Chen and Zuolin Lu provided critical, constructive revisions of the manuscript; Yue Zhang, Yunyuan Kong, Enying Gong, Xuejun Yin, Xiaoxia Wei, Maigeng Zhou, Jiajuan Yang, Chi Hu, Xunliang Tong, Luzhao Feng, and Chen Wang contributed to data acquisition, preprocessing, and interpretation of findings; and all authors approved the final version of the manuscript and agree to be accountable for all aspects of the work.

## Data availability

SummaryLJlevel data supporting the findings of this study are provided in the Supplementary Materials. Access to the full, de-identified individual-level dataset can be made available upon reasonable request to the corresponding author. Requests will be evaluated in accordance with our institution’s data-sharing policies and applicable ethical and privacy regulations. A formal data-use agreement will be required to ensure secure handling of the dataset.

## Appendix A. Supplementary material

Supplementary materials to this article can be found online.

