## Supplementary_Materials for "Multimorbidity Profiles in Patient Population from Central China: A Study Based on Electronic Health Records"

Supplementary Methods

Section S1. Methods for identifying non-random disease associations and measuring the strength of association between health conditions.

To quantify the strength of the non-random association between pairs of health conditions within the network, we selected the partial correlation coefficient. While other metrics are sometimes used to assess co-occurrence, they were deemed less suitable for our specific goal of identifying direct associations adjusted for the presence of other conditions. For instance, the standard Pearson correlation coefficient measures the linear association between two conditions but does not account for the potential confounding influence of other conditions within the complex web of multimorbidity. Measures like the Odds Ratio (OR) or Relative Risk (RR) quantify the likelihood or risk of co-occurrence but can be sensitive to disease prevalence; specifically, RR tends to overestimate relationships involving rare diseases and underestimate those between highly prevalent ones. Furthermore, integrating ORs directly into a network framework to calculate cumulative association strength while controlling for all other variables simultaneously presents challenges. Similarity indices like the Salton Cosine Index (SCI), often used in information retrieval and sometimes adapted for co-occurrence, measure the overlap relative to the magnitude of individual occurrences but, similar to Pearson correlation, typically do not inherently control for the influence of other conditions in the analysis^[1]^. Therefore, we chose partial correlation (*ρij*∣*k*) because it specifically addresses the limitation of standard correlation by estimating the association strength between two conditions (*i* and *j*) while statistically removing the linear effects of all other included conditions (*k*), thus providing a more accurate measure of the direct pairwise relationship within the overall network^[2]^.

In this study, we employed partial correlation analysis to quantify the strength of non-random association between pairs of health conditions, while statistically controlling for the potential influence of all other health conditions included in the analysis.

First, health conditions with a prevalence below 0.02% in the study population were excluded from this part of the analysis to focus on more common conditions and ensure robust estimates of association. Subsequently, using R software (Version 4.4.3) and relevant packages, the partial correlation coefficients (*ρij*∣*k*) were calculated for all possible N*(N-1)/2 pairs among the remaining N health conditions. This coefficient assesses the correlation between two conditions, *i* and *j*, after accounting for the effects of all other conditions (*k*) in the dataset^[3]^. The formula for the partial correlation is:

$$\rho_{\left( ij \mid k \right)}=\frac{\rho_{ij}-\rho_{ik}\rho_{jk}}{\sqrt{\left( 1-\rho_{ik}^{2} \right)\times\left( \left( 1-\rho_{jk}^{2} \right) \right)}}$$

Where *ρ ij* is the Pearson correlation coefficient between conditions *i* and *j*, *ρik* is the Pearson correlation between conditions *i* and *k*, and *ρjk* is the Pearson correlation between conditions *j* and *k*. The Pearson correlation coefficient (*ρij* ) itself is calculated as:

$$\rho_{ij}=\frac{\left( N\times C_{ij} \right)-\left( C_{i}\times C_{\cdot j} \right)}{\sqrt{C_{i}\times C_{\cdot j}\times\left( N-C_{i\cdot} \right)\times\left( N-C_{\cdot j} \right)}}$$

where:

*N* = total number of individuals in dataset

*C_ij_* = number of individuals who had ever been diagnosed with both conditions i and j

*C_i_*· = number of individuals who had ever been diagnosed with condition i

*C_j_* = number of individuals who had ever been diagnosed with condition j

A pair of health conditions was considered to have a significant non-random positive association only if the partial correlation coefficient was positive and the associated *P*-value remained statistically significant after adjustment for multiple comparisons using the Benjamini-Hochberg (BH) procedure to control the False Discovery Rate (FDR) at less than 0.05 (FDR < 0.05). Condition pairs meeting these criteria were included as edges in the subsequent construction of the disease co-occurrence network.

Section S2. Quantifying Disease Centrality using the Multimorbidity Coefficient (MMC)

To identify central ('hub') health conditions within the disease co-occurrence network, this study utilized the Multimorbidity Coefficient (MMC) to quantify the centrality of each disease node. Compared to 'degree centrality', a common metric in network analysis that simply counts the number of connections a node has, the MMC provides a more nuanced and comprehensive measure^[4]^.

Specifically, while 'degree' only reflects how many connections a disease node has (i.e., how many other conditions it is significantly associated with), the MMC considers both the number of connections and the strength of these connections. It is calculated by summing the values of all positive partial correlation coefficients for the edges connected to a specific disease node. Consequently, the MMC better reflects the cumulative burden of non-random positive associations for a condition within the network. A condition with a high MMC value indicates that it is associated with many other conditions and/or that these associations are strong, potentially suggesting a central role in multimorbidity patterns or involvement in shared risk factors or pathophysiological pathways.

The calculation method for the MMC is as follows:

For a given health condition *i*, the MMC is calculated using the formula:

$$\mathrm{MMC}_{i}=\sum_{\begin{aligned} j,k\neq i \end{aligned}} \rho_{ij\mid k}\cdot\mathbb{I}(\rho_{ij\mid k}>0)$$

Where:

- $\rho ij|k$ is the partial correlation coefficient between condition *i* and condition *j*, controlling for all other conditions *k*.
- $\sum_{\begin{aligned} j,k\neq i \end{aligned}} \rho_{ij\mid k}$ indicates summation over all *j* and *k* that are different from *i*.
- $\mathbb{I}(\rho_{ij\mid k}>0)$ is an indicator function that takes the value 1 if the partial correlation coefficient $\rho_{ij\mid k}$is greater than 0 (i.e., positive), and 0 otherwise. This ensures that only the strengths of positive correlations are summed for the MMC.

Therefore, the MMC can also be interpreted as the number of positive connections (edges) for a condition multiplied by the average positive partial correlation strength per connection.

Section S3. Sensitivity Analyses

To evaluate the robustness of our findings, particularly the identification of hub diseases, we conducted sensitivity analyses by assessing the stability of the Multimorbidity Coefficient (MMC) rankings under different statistical criteria for defining an edge in the network. We systematically varied two key statistical thresholds for network edge inclusion: the significance level for the False Discovery Rate (with α set to 0.1, 0.05, and 0.01) and the minimum partial correlation coefficient (with φ ranging from 0 to 0.05). An interactive visualization of MMC sensitivity to these parameters is also available at: https://pumc-multimorbidity.shinyapps.io/mmc-visualization/

Supplementary Results

Supplementary Table S1. Prevalence of the 50 most common triads in the overall study population

| Condition triads | Prevalence (95% Confidence interval) |
| --- | --- |
| (I10, I25, K29) | 2.34% (2.32% - 2.35%) |
| (I10, I25, I63) | 1.99% (1.98% - 2.01%) |
| (I10, I25, I50) | 1.79% (1.77% - 1.80%) |
| (E78, I10, I25) | 1.74% (1.72% - 1.75%) |
| (I10, I63, K29) | 1.68% (1.66% - 1.69%) |
| (E11, I10, I25) | 1.64% (1.62% - 1.65%) |
| (E78, I10, K29) | 1.51% (1.50% - 1.52%) |
| (E78, I10, I63) | 1.43% (1.41% - 1.44%) |
| (I10, J40, K29) | 1.41% (1.39% - 1.42%) |
| (E11, E78, I10) | 1.39% (1.38% - 1.41%) |
| (I25, I50, K29) | 1.30% (1.29% - 1.31%) |
| (E11, I10, K29) | 1.27% (1.26% - 1.29%) |
| (E11, I10, I63) | 1.26% (1.25% - 1.28%) |
| (I25, I63, K29) | 1.25% (1.24% - 1.26%) |
| (I10, I25, J40) | 1.23% (1.21% - 1.24%) |
| (I10, K29, M50) | 1.20% (1.19% - 1.21%) |
| (I10, I50, K29) | 1.20% (1.19% - 1.21%) |
| (I10, I20, I25) | 1.19% (1.18% - 1.20%) |
| (G47, I10, I25) | 1.17% (1.16% - 1.18%) |
| (I10, I63, I67) | 1.14% (1.13% - 1.15%) |
| (I10, I63, M50) | 1.12% (1.11% - 1.13%) |
| (I10, K29, M51) | 1.11% (1.10% - 1.12%) |
| (G47, I10, K29) | 1.11% (1.10% - 1.12%) |
| (I10, K29, M54) | 1.10% (1.08% - 1.11%) |
| (E78, I25, K29) | 1.06% (1.05% - 1.07%) |
| (I10, I25, I49) | 1.05% (1.04% - 1.06%) |
| (I10, J20, K29) | 1.05% (1.04% - 1.06%) |
| (I10, I25, M50) | 1.02% (1.01% - 1.03%) |
| (I10, K29, L30) | 1.00% (0.99% - 1.01%) |
| (G47, I10, I63) | 1.00% (0.98% - 1.01%) |
| (I10, I25, M51) | 0.99% (0.98% - 1.00%) |
| (I10, I63, I70) | 0.99% (0.97% - 1.00%) |
| (H81, I10, I63) | 0.99% (0.97% - 1.00%) |
| (I10, I50, I63) | 0.98% (0.97% - 0.99%) |
| (I10, I25, I70) | 0.97% (0.96% - 0.98%) |
| (I10, I25, L30) | 0.96% (0.95% - 0.97%) |
| (I10, I63, J40) | 0.95% (0.94% - 0.96%) |
| (I25, I50, I63) | 0.93% (0.92% - 0.94%) |
| (E78, G47, I10) | 0.93% (0.92% - 0.94%) |
| (E78, I10, J40) | 0.92% (0.91% - 0.93%) |
| (E78, I25, I63) | 0.92% (0.91% - 0.93%) |
| (I10, M51, M54) | 0.92% (0.91% - 0.93%) |
| (I10, I25, I67) | 0.91% (0.90% - 0.92%) |
| (I10, I67, K29) | 0.91% (0.90% - 0.92%) |
| (I10, I25, M54) | 0.90% (0.89% - 0.91%) |
| (J20, J40, K29) | 0.90% (0.89% - 0.91%) |
| (I25, J40, K29) | 0.89% (0.88% - 0.91%) |
| (I10, I63, M51) | 0.89% (0.88% - 0.90%) |
| (I10, I25, J20) | 0.89% (0.88% - 0.90%) |
| (I20, I25, I50) | 0.88% (0.87% - 0.89%) |

Supplementary Table S2. Prevalence of the 50 most common triads in female individuals.

| Condition triads | Prevalence (95% Confidence interval) |
| --- | --- |
| (I10, I25, K29) | 2.42% (2.40% - 2.45%) |
| (I10, I25, I63) | 1.96% (1.94% - 1.98%) |
| (I10, I63, K29) | 1.78% (1.76% - 1.80%) |
| (E78, I10, I25) | 1.73% (1.71% - 1.75%) |
| (I10, I25, I50) | 1.66% (1.64% - 1.68%) |
| (E78, I10, K29) | 1.64% (1.62% - 1.66%) |
| (E11, I10, I25) | 1.54% (1.52% - 1.56%) |
| (E78, I10, I63) | 1.50% (1.48% - 1.51%) |
| (I10, J40, K29) | 1.48% (1.47% - 1.50%) |
| (I10, K29, M50) | 1.42% (1.41% - 1.44%) |
| (I25, I63, K29) | 1.35% (1.33% - 1.37%) |
| (E11, E78, I10) | 1.33% (1.31% - 1.35%) |
| (E11, I10, K29) | 1.32% (1.31% - 1.34%) |
| (I25, I50, K29) | 1.30% (1.28% - 1.31%) |
| (I10, I63, M50) | 1.27% (1.25% - 1.28%) |
| (I10, K29, M51) | 1.26% (1.25% - 1.28%) |
| (G47, I10, K29) | 1.24% (1.22% - 1.25%) |
| (I10, I25, J40) | 1.22% (1.20% - 1.23%) |
| (E11, I10, I63) | 1.21% (1.19% - 1.23%) |
| (G47, I10, I25) | 1.20% (1.18% - 1.22%) |
| (I10, I50, K29) | 1.20% (1.18% - 1.21%) |
| (I10, K29, M54) | 1.18% (1.16% - 1.20%) |
| (E78, I25, K29) | 1.18% (1.16% - 1.19%) |
| (I10, I25, M50) | 1.14% (1.12% - 1.15%) |
| (H81, I10, I63) | 1.12% (1.10% - 1.13%) |
| (I10, J20, K29) | 1.11% (1.09% - 1.13%) |
| (I10, I63, I67) | 1.11% (1.09% - 1.12%) |
| (I10, I20, I25) | 1.07% (1.05% - 1.09%) |
| (G47, I10, I63) | 1.07% (1.05% - 1.08%) |
| (I10, I25, M51) | 1.07% (1.05% - 1.08%) |
| (H81, I10, K29) | 1.05% (1.03% - 1.06%) |
| (I10, I25, I49) | 1.03% (1.02% - 1.05%) |
| (K29, M51, M54) | 1.01% (1.00% - 1.03%) |
| (I63, K29, M50) | 1.01% (0.99% - 1.02%) |
| (E78, G47, I10) | 1.00% (0.98% - 1.01%) |
| (J20, J40, K29) | 1.00% (0.98% - 1.01%) |
| (I10, I67, K29) | 0.99% (0.98% - 1.01%) |
| (K29, M50, M51) | 0.99% (0.98% - 1.01%) |
| (I10, M51, M54) | 0.99% (0.98% - 1.01%) |
| (E78, I10, M50) | 0.99% (0.98% - 1.01%) |
| (E78, I25, I63) | 0.99% (0.97% - 1.00%) |
| (I10, K29, L30) | 0.98% (0.96% - 0.99%) |
| (I10, I63, M51) | 0.98% (0.96% - 0.99%) |
| (I10, I63, J40) | 0.97% (0.96% - 0.99%) |
| (I25, K29, M50) | 0.97% (0.96% - 0.99%) |
| (E78, I10, J40) | 0.97% (0.96% - 0.99%) |
| (E78, I63, K29) | 0.96% (0.95% - 0.98%) |
| (H81, I10, I25) | 0.96% (0.94% - 0.97%) |
| (I10, K29, M25) | 0.95% (0.93% - 0.96%) |
| (I25, J40, K29) | 0.95% (0.93% - 0.96%) |

Supplementary Table S3. Prevalence of the 50 most common triads in male individuals.

| Condition triads | Prevalence (95% Confidence interval) |
| --- | --- |
| (I10, I25, K29) | 2.24% (2.22% - 2.26%) |
| (I10, I25, I63) | 2.03% (2.01% - 2.05%) |
| (I10, I25, I50) | 1.92% (1.90% - 1.95%) |
| (E78, I10, I25) | 1.75% (1.73% - 1.77%) |
| (E11, I10, I25) | 1.74% (1.72% - 1.76%) |
| (I10, I63, K29) | 1.56% (1.54% - 1.58%) |
| (E11, E78, I10) | 1.46% (1.45% - 1.48%) |
| (E78, I10, K29) | 1.37% (1.36% - 1.39%) |
| (E78, I10, I63) | 1.36% (1.34% - 1.37%) |
| (I10, J40, K29) | 1.32% (1.30% - 1.34%) |
| (E11, I10, I63) | 1.32% (1.30% - 1.34%) |
| (I10, I20, I25) | 1.32% (1.30% - 1.34%) |
| (I25, I50, K29) | 1.30% (1.29% - 1.32%) |
| (I10, I25, J40) | 1.23% (1.22% - 1.25%) |
| (E11, I10, K29) | 1.22% (1.20% - 1.24%) |
| (I10, I50, K29) | 1.20% (1.18% - 1.21%) |
| (I10, I63, I67) | 1.17% (1.15% - 1.19%) |
| (I10, I25, N40) | 1.17% (1.15% - 1.19%) |
| (I10, I25, J44) | 1.17% (1.15% - 1.18%) |
| (I25, I63, K29) | 1.14% (1.12% - 1.16%) |
| (G47, I10, I25) | 1.13% (1.12% - 1.15%) |
| (I10, I25, I49) | 1.07% (1.06% - 1.09%) |
| (I10, I25, L30) | 1.06% (1.05% - 1.08%) |
| (I10, K29, N40) | 1.06% (1.04% - 1.07%) |
| (I10, I25, I70) | 1.05% (1.04% - 1.07%) |
| (I10, I63, I70) | 1.05% (1.04% - 1.07%) |
| (I10, I50, I63) | 1.03% (1.02% - 1.05%) |
| (I10, J44, K29) | 1.03% (1.01% - 1.04%) |
| (I10, K29, L30) | 1.02% (1.00% - 1.03%) |
| (I10, I63, N40) | 1.00% (0.99% - 1.02%) |
| (I10, K29, M54) | 1.00% (0.99% - 1.02%) |
| (I10, J20, K29) | 0.98% (0.97% - 1.00%) |
| (G47, I10, K29) | 0.97% (0.95% - 0.98%) |
| (I25, I50, I63) | 0.97% (0.95% - 0.98%) |
| (I20, I25, I50) | 0.97% (0.95% - 0.98%) |
| (I10, K29, M50) | 0.96% (0.95% - 0.98%) |
| (I10, I63, M50) | 0.96% (0.94% - 0.98%) |
| (I25, J44, K29) | 0.95% (0.94% - 0.97%) |
| (I10, K29, M51) | 0.94% (0.93% - 0.96%) |
| (I10, I63, J40) | 0.93% (0.92% - 0.95%) |
| (E78, I25, K29) | 0.93% (0.91% - 0.94%) |
| (G47, I10, I63) | 0.92% (0.90% - 0.93%) |
| (I25, I50, J44) | 0.92% (0.90% - 0.93%) |
| (E11, I10, I50) | 0.91% (0.89% - 0.92%) |
| (I10, I25, I67) | 0.91% (0.89% - 0.92%) |
| (I10, I25, M51) | 0.90% (0.89% - 0.92%) |
| (I10, I25, J15) | 0.90% (0.89% - 0.92%) |
| (I10, I25, M50) | 0.89% (0.87% - 0.90%) |
| (I10, I25, J20) | 0.88% (0.87% - 0.90%) |
| (I10, I25, M54) | 0.88% (0.86% - 0.89%) |

Supplementary Table S4. Prevalence of the 50 most common triads in individuals aged 1-9 years.

| Condition triads | Prevalence (95% Confidence interval) |
| --- | --- |
| (J20, J31, J32) | 3.92% (3.83% - 4.00%) |
| (J18, J20, J31) | 3.84% (3.76% - 3.93%) |
| (J20, J31, J35) | 3.78% (3.70% - 3.86%) |
| (J20, J30, J31) | 3.73% (3.65% - 3.81%) |
| (J31, J32, J35) | 3.57% (3.49% - 3.65%) |
| (J18, J20, J40) | 3.56% (3.49% - 3.64%) |
| (J20, J32, J35) | 3.54% (3.46% - 3.62%) |
| (J20, J31, J40) | 3.06% (2.99% - 3.13%) |
| (J30, J32, J35) | 3.05% (2.98% - 3.12%) |
| (J20, J30, J32) | 2.93% (2.86% - 3.00%) |
| (J20, J30, J35) | 2.87% (2.80% - 2.94%) |
| (J20, J31, L30) | 2.75% (2.68% - 2.82%) |
| (J30, J31, J32) | 2.71% (2.64% - 2.78%) |
| (J30, J31, J35) | 2.69% (2.62% - 2.76%) |
| (J18, J20, J21) | 2.68% (2.61% - 2.74%) |
| (J18, J20, L30) | 2.49% (2.42% - 2.55%) |
| (J18, J20, J30) | 2.43% (2.37% - 2.50%) |
| (J15, J18, J20) | 2.39% (2.32% - 2.45%) |
| (J18, J20, J32) | 2.12% (2.06% - 2.18%) |
| (J18, J20, J35) | 2.02% (1.96% - 2.08%) |
| (J20, J21, J31) | 1.95% (1.89% - 2.00%) |
| (J20, J30, J40) | 1.92% (1.86% - 1.97%) |
| (H52, J20, J31) | 1.88% (1.83% - 1.94%) |
| (I88, J18, J20) | 1.86% (1.80% - 1.91%) |
| (J20, J40, L30) | 1.85% (1.79% - 1.90%) |
| (J20, J30, L30) | 1.84% (1.79% - 1.90%) |
| (I88, J20, J31) | 1.74% (1.68% - 1.79%) |
| (J20, J21, J40) | 1.70% (1.64% - 1.75%) |
| (J20, J32, J40) | 1.68% (1.63% - 1.73%) |
| (J03, J18, J20) | 1.63% (1.58% - 1.68%) |
| (J20, J32, L30) | 1.63% (1.57% - 1.68%) |
| (J20, J35, L30) | 1.58% (1.52% - 1.63%) |
| (J18, J30, J31) | 1.57% (1.52% - 1.63%) |
| (J20, J35, J40) | 1.57% (1.52% - 1.62%) |
| (J18, J31, J32) | 1.55% (1.50% - 1.60%) |
| (J20, J21, J30) | 1.52% (1.46% - 1.57%) |
| (J18, J31, J35) | 1.51% (1.46% - 1.56%) |
| (J18, J20, K52) | 1.51% (1.45% - 1.56%) |
| (H52, J18, J20) | 1.49% (1.44% - 1.54%) |
| (J18, J20, K30) | 1.48% (1.43% - 1.53%) |
| (J18, J31, J40) | 1.48% (1.43% - 1.53%) |
| (J20, J31, K52) | 1.47% (1.42% - 1.52%) |
| (J18, J20, L50) | 1.44% (1.39% - 1.49%) |
| (J31, J32, L30) | 1.44% (1.39% - 1.49%) |
| (I88, J20, J40) | 1.43% (1.38% - 1.48%) |
| (J30, J31, L30) | 1.42% (1.37% - 1.47%) |
| (J06, J18, J20) | 1.42% (1.37% - 1.47%) |
| (J20, J31, K30) | 1.41% (1.36% - 1.46%) |
| (J20, J31, L50) | 1.41% (1.36% - 1.46%) |
| (J31, J35, L30) | 1.41% (1.36% - 1.46%) |

Supplementary Table S5. Prevalence of the 50 most common triads in individuals aged 10-19 years.

| Condition triads | Prevalence (95% Confidence interval) |
| --- | --- |
| (J30, J31, J32) | 0.75% (0.72% - 0.78%) |
| (J30, J32, J35) | 0.54% (0.51% - 0.57%) |
| (J31, J32, J35) | 0.53% (0.50% - 0.56%) |
| (H52, J31, J32) | 0.47% (0.45% - 0.50%) |
| (H52, J30, J32) | 0.44% (0.42% - 0.47%) |
| (J31, J32, J34) | 0.41% (0.38% - 0.43%) |
| (H52, J30, J31) | 0.41% (0.38% - 0.43%) |
| (J20, J31, J32) | 0.38% (0.36% - 0.41%) |
| (J30, J32, J34) | 0.37% (0.34% - 0.39%) |
| (J30, J31, J35) | 0.34% (0.32% - 0.37%) |
| (J20, J30, J31) | 0.33% (0.31% - 0.35%) |
| (H52, J20, J31) | 0.32% (0.30% - 0.34%) |
| (J20, J30, J32) | 0.32% (0.29% - 0.34%) |
| (H52, L30, L70) | 0.31% (0.29% - 0.33%) |
| (J31, J32, L30) | 0.29% (0.27% - 0.31%) |
| (J30, J31, J34) | 0.29% (0.27% - 0.31%) |
| (H52, J32, J35) | 0.29% (0.27% - 0.31%) |
| (H52, J20, L30) | 0.28% (0.26% - 0.30%) |
| (H52, J20, J40) | 0.27% (0.25% - 0.29%) |
| (J20, J31, J40) | 0.27% (0.25% - 0.29%) |
| (H52, J20, L70) | 0.27% (0.25% - 0.29%) |
| (J30, J32, L30) | 0.26% (0.24% - 0.28%) |
| (H52, J31, J35) | 0.26% (0.24% - 0.28%) |
| (J30, J31, L30) | 0.26% (0.24% - 0.28%) |
| (H52, J20, J30) | 0.26% (0.24% - 0.28%) |
| (J31, J32, L70) | 0.25% (0.23% - 0.27%) |
| (H52, J31, L30) | 0.25% (0.23% - 0.27%) |
| (H52, J20, J32) | 0.24% (0.22% - 0.26%) |
| (J20, J40, L30) | 0.23% (0.21% - 0.25%) |
| (H52, J30, J35) | 0.23% (0.21% - 0.25%) |
| (J30, J32, L70) | 0.23% (0.21% - 0.25%) |
| (J20, J40, K29) | 0.23% (0.21% - 0.25%) |
| (J31, J32, J40) | 0.23% (0.21% - 0.24%) |
| (J20, J32, J35) | 0.22% (0.20% - 0.24%) |
| (H52, J30, L30) | 0.22% (0.20% - 0.24%) |
| (J30, J31, L70) | 0.22% (0.20% - 0.24%) |
| (J20, J31, L30) | 0.21% (0.20% - 0.23%) |
| (H52, J31, L70) | 0.21% (0.19% - 0.23%) |
| (J20, J31, J35) | 0.20% (0.19% - 0.22%) |
| (J20, L30, L70) | 0.20% (0.18% - 0.22%) |
| (H52, J30, L70) | 0.20% (0.18% - 0.22%) |
| (J15, J18, J20) | 0.19% (0.18% - 0.21%) |
| (F32, F41, G47) | 0.19% (0.17% - 0.21%) |
| (H52, J20, M25) | 0.19% (0.17% - 0.20%) |
| (H52, J32, L30) | 0.19% (0.17% - 0.20%) |
| (H52, J20, K29) | 0.18% (0.17% - 0.20%) |
| (J30, J31, J40) | 0.18% (0.17% - 0.20%) |
| (J20, J30, L30) | 0.18% (0.17% - 0.20%) |
| (J20, J32, J40) | 0.18% (0.17% - 0.20%) |
| (H52, J31, J34) | 0.18% (0.16% - 0.20%) |

Supplementary Table S6. Prevalence of the 50 most common triads in individuals aged 20-29 years.

| Condition triads | Prevalence (95% Confidence interval) |
| --- | --- |
| (E28, N92, N97) | 0.47% (0.45% - 0.50%) |
| (E28, N92, N93) | 0.42% (0.40% - 0.44%) |
| (L30, N92, N93) | 0.42% (0.40% - 0.44%) |
| (E28, N91, N92) | 0.35% (0.33% - 0.37%) |
| (N92, N93, N97) | 0.34% (0.32% - 0.36%) |
| (N85, N92, N93) | 0.33% (0.32% - 0.35%) |
| (E16, E28, N92) | 0.30% (0.28% - 0.32%) |
| (N91, N92, N93) | 0.30% (0.28% - 0.32%) |
| (E28, L30, N92) | 0.29% (0.27% - 0.31%) |
| (J30, J31, J32) | 0.28% (0.26% - 0.30%) |
| (L30, L70, N92) | 0.28% (0.26% - 0.29%) |
| (E16, N92, N97) | 0.27% (0.26% - 0.29%) |
| (N72, N92, N93) | 0.27% (0.25% - 0.29%) |
| (J31, J32, J34) | 0.27% (0.25% - 0.29%) |
| (L30, N91, N92) | 0.27% (0.25% - 0.28%) |
| (L30, N85, N92) | 0.24% (0.23% - 0.26%) |
| (N62, N92, N93) | 0.24% (0.22% - 0.25%) |
| (N83, N92, N93) | 0.22% (0.21% - 0.24%) |
| (J30, J32, J34) | 0.22% (0.21% - 0.24%) |
| (N91, N92, N97) | 0.22% (0.21% - 0.24%) |
| (L30, N92, N97) | 0.21% (0.19% - 0.22%) |
| (L30, N62, N92) | 0.21% (0.19% - 0.22%) |
| (K29, N92, N93) | 0.21% (0.19% - 0.22%) |
| (L70, N92, N93) | 0.20% (0.19% - 0.22%) |
| (N85, N91, N92) | 0.20% (0.19% - 0.22%) |
| (J40, L30, N92) | 0.20% (0.19% - 0.21%) |
| (E16, E28, N97) | 0.20% (0.19% - 0.21%) |
| (K29, L30, N92) | 0.20% (0.18% - 0.21%) |
| (E28, N93, N97) | 0.20% (0.18% - 0.21%) |
| (L30, N72, N92) | 0.20% (0.18% - 0.21%) |
| (J20, J40, L30) | 0.19% (0.18% - 0.21%) |
| (L30, M54, N92) | 0.19% (0.18% - 0.21%) |
| (M54, N92, N93) | 0.19% (0.17% - 0.20%) |
| (K01, L30, N92) | 0.18% (0.17% - 0.19%) |
| (N85, N92, N97) | 0.18% (0.17% - 0.19%) |
| (J40, N92, N93) | 0.18% (0.17% - 0.19%) |
| (J31, J32, L30) | 0.18% (0.16% - 0.19%) |
| (K01, N92, N93) | 0.17% (0.16% - 0.19%) |
| (L23, L30, N92) | 0.17% (0.16% - 0.18%) |
| (L30, L50, N92) | 0.17% (0.16% - 0.18%) |
| (J30, J31, J34) | 0.17% (0.16% - 0.18%) |
| (K01, L30, L70) | 0.17% (0.16% - 0.18%) |
| (L30, N85, N93) | 0.17% (0.16% - 0.18%) |
| (J31, L30, N92) | 0.17% (0.15% - 0.18%) |
| (K29, L30, M54) | 0.17% (0.15% - 0.18%) |
| (N84, N92, N93) | 0.16% (0.15% - 0.17%) |
| (J40, K29, L30) | 0.16% (0.15% - 0.17%) |
| (J30, J31, L30) | 0.16% (0.15% - 0.17%) |
| (L23, L30, L70) | 0.16% (0.15% - 0.17%) |
| (J20, N92, N93) | 0.16% (0.15% - 0.17%) |

Supplementary Table S7. Prevalence of the 50 most common triads in individuals aged 30-39 years.

| Condition triads | Prevalence (95% Confidence interval) |
| --- | --- |
| (N85, N92, N93) | 0.41% (0.40% - 0.43%) |
| (L30, N92, N93) | 0.37% (0.35% - 0.39%) |
| (J20, J40, L30) | 0.35% (0.34% - 0.37%) |
| (J30, J31, J32) | 0.34% (0.32% - 0.36%) |
| (N72, N92, N93) | 0.31% (0.29% - 0.32%) |
| (J31, J40, L30) | 0.30% (0.29% - 0.32%) |
| (N62, N92, N93) | 0.30% (0.29% - 0.32%) |
| (J20, J40, K29) | 0.30% (0.29% - 0.32%) |
| (J31, J32, J34) | 0.29% (0.28% - 0.31%) |
| (J40, K29, L30) | 0.29% (0.27% - 0.30%) |
| (M54, N13, N20) | 0.28% (0.27% - 0.30%) |
| (J40, L30, M54) | 0.26% (0.25% - 0.27%) |
| (N83, N92, N93) | 0.26% (0.24% - 0.27%) |
| (N91, N92, N93) | 0.26% (0.24% - 0.27%) |
| (K29, L30, M54) | 0.25% (0.24% - 0.27%) |
| (J20, K29, L30) | 0.25% (0.24% - 0.27%) |
| (L30, N85, N92) | 0.25% (0.24% - 0.26%) |
| (M54, N92, N93) | 0.25% (0.24% - 0.26%) |
| (J31, K29, L30) | 0.25% (0.24% - 0.26%) |
| (J20, J31, J40) | 0.25% (0.23% - 0.26%) |
| (J30, J32, J34) | 0.24% (0.23% - 0.26%) |
| (L30, N62, N92) | 0.24% (0.23% - 0.26%) |
| (J31, J32, L30) | 0.24% (0.23% - 0.26%) |
| (L30, M51, M54) | 0.24% (0.23% - 0.25%) |
| (N84, N92, N93) | 0.24% (0.23% - 0.25%) |
| (J31, J40, K29) | 0.24% (0.23% - 0.25%) |
| (E28, N92, N97) | 0.24% (0.22% - 0.25%) |
| (G47, J40, L30) | 0.23% (0.22% - 0.25%) |
| (J40, L30, N92) | 0.23% (0.22% - 0.25%) |
| (K29, N92, N93) | 0.23% (0.22% - 0.24%) |
| (J40, N92, N93) | 0.23% (0.22% - 0.24%) |
| (J40, K29, M54) | 0.23% (0.21% - 0.24%) |
| (L30, M54, N92) | 0.23% (0.21% - 0.24%) |
| (N85, N91, N92) | 0.22% (0.21% - 0.24%) |
| (G47, K29, L30) | 0.22% (0.21% - 0.23%) |
| (E78, E79, I10) | 0.22% (0.21% - 0.23%) |
| (E11, E78, I10) | 0.22% (0.21% - 0.23%) |
| (E28, N92, N93) | 0.22% (0.21% - 0.23%) |
| (J30, J31, L30) | 0.22% (0.21% - 0.23%) |
| (J20, J40, M54) | 0.22% (0.20% - 0.23%) |
| (L30, N72, N92) | 0.22% (0.20% - 0.23%) |
| (J31, L30, M54) | 0.21% (0.20% - 0.23%) |
| (K29, L30, N92) | 0.21% (0.20% - 0.23%) |
| (J31, J32, J40) | 0.21% (0.20% - 0.23%) |
| (J20, J40, N92) | 0.21% (0.19% - 0.22%) |
| (B34, N72, N87) | 0.21% (0.19% - 0.22%) |
| (L30, M50, M54) | 0.21% (0.19% - 0.22%) |
| (J20, J31, L30) | 0.20% (0.19% - 0.22%) |
| (J31, K21, K29) | 0.20% (0.19% - 0.22%) |
| (K29, M51, M54) | 0.20% (0.19% - 0.22%) |

Supplementary Table S8. Prevalence of the 50 most common triads in individuals aged 40-49 years.

| Condition triads | Prevalence (95% Confidence interval) |
| --- | --- |
| (E11, E78, I10) | 0.61% (0.59% - 0.63%) |
| (E78, I10, K29) | 0.49% (0.47% - 0.51%) |
| (E78, I10, I25) | 0.47% (0.45% - 0.49%) |
| (K29, M51, M54) | 0.41% (0.39% - 0.43%) |
| (I10, I25, K29) | 0.40% (0.39% - 0.42%) |
| (M50, M51, M54) | 0.40% (0.38% - 0.41%) |
| (K29, M50, M54) | 0.40% (0.38% - 0.41%) |
| (I10, J40, K29) | 0.39% (0.38% - 0.41%) |
| (E78, I10, K76) | 0.39% (0.37% - 0.40%) |
| (J20, J40, K29) | 0.37% (0.36% - 0.39%) |
| (L30, N92, N93) | 0.37% (0.36% - 0.39%) |
| (E78, E79, I10) | 0.37% (0.35% - 0.38%) |
| (I10, K29, M50) | 0.36% (0.35% - 0.38%) |
| (J40, K29, M54) | 0.36% (0.34% - 0.38%) |
| (E78, G47, I10) | 0.36% (0.34% - 0.37%) |
| (I10, K29, L30) | 0.35% (0.34% - 0.37%) |
| (G47, I10, K29) | 0.35% (0.33% - 0.36%) |
| (N83, N92, N93) | 0.35% (0.33% - 0.36%) |
| (D25, N92, N93) | 0.35% (0.33% - 0.36%) |
| (E78, I10, L30) | 0.34% (0.33% - 0.36%) |
| (E11, I10, I25) | 0.34% (0.33% - 0.36%) |
| (I10, K29, M54) | 0.34% (0.33% - 0.36%) |
| (J40, K29, M50) | 0.34% (0.32% - 0.35%) |
| (J40, K29, L30) | 0.34% (0.32% - 0.35%) |
| (E78, I10, M50) | 0.33% (0.32% - 0.35%) |
| (K29, M50, M51) | 0.33% (0.32% - 0.35%) |
| (K29, N92, N93) | 0.33% (0.32% - 0.35%) |
| (E11, I10, K29) | 0.33% (0.31% - 0.34%) |
| (N72, N92, N93) | 0.33% (0.31% - 0.34%) |
| (N62, N92, N93) | 0.33% (0.31% - 0.34%) |
| (K29, L30, M54) | 0.32% (0.30% - 0.33%) |
| (E78, I10, J40) | 0.32% (0.30% - 0.33%) |
| (G47, K29, L30) | 0.32% (0.30% - 0.33%) |
| (J31, K21, K29) | 0.31% (0.30% - 0.33%) |
| (I10, J40, L30) | 0.31% (0.30% - 0.33%) |
| (I10, J20, K29) | 0.31% (0.29% - 0.32%) |
| (M25, M51, M54) | 0.31% (0.29% - 0.32%) |
| (N85, N92, N93) | 0.30% (0.29% - 0.32%) |
| (M54, N92, N93) | 0.30% (0.29% - 0.32%) |
| (J31, J40, K29) | 0.30% (0.29% - 0.32%) |
| (G47, I10, L30) | 0.30% (0.29% - 0.31%) |
| (I10, J20, J40) | 0.30% (0.28% - 0.31%) |
| (G47, K29, M50) | 0.30% (0.28% - 0.31%) |
| (E78, I10, N20) | 0.30% (0.28% - 0.31%) |
| (I10, M51, M54) | 0.30% (0.28% - 0.31%) |
| (I10, N13, N20) | 0.29% (0.28% - 0.31%) |
| (E78, I10, M54) | 0.29% (0.28% - 0.31%) |
| (J40, M50, M54) | 0.29% (0.28% - 0.31%) |
| (K29, L30, M50) | 0.29% (0.28% - 0.30%) |
| (L30, M51, M54) | 0.29% (0.28% - 0.30%) |

Supplementary Table S9. Prevalence of the 50 most common triads in individuals aged 50-59 years.

| Condition triads | Prevalence (95% Confidence interval) |
| --- | --- |
| (E11, E78, I10) | 1.27% (1.25% - 1.30%) |
| (E78, I10, I25) | 1.25% (1.22% - 1.27%) |
| (I10, I25, K29) | 1.24% (1.22% - 1.27%) |
| (E78, I10, K29) | 1.21% (1.18% - 1.23%) |
| (E11, I10, I25) | 1.03% (1.00% - 1.05%) |
| (E78, I10, I63) | 0.99% (0.97% - 1.01%) |
| (I10, J40, K29) | 0.99% (0.96% - 1.01%) |
| (I10, K29, M50) | 0.94% (0.92% - 0.96%) |
| (E11, I10, K29) | 0.90% (0.88% - 0.92%) |
| (I10, I25, I63) | 0.81% (0.79% - 0.83%) |
| (E78, I10, M50) | 0.80% (0.78% - 0.82%) |
| (I10, I63, K29) | 0.78% (0.76% - 0.80%) |
| (I10, K29, M54) | 0.78% (0.76% - 0.80%) |
| (G47, I10, K29) | 0.77% (0.75% - 0.79%) |
| (I10, I63, M50) | 0.75% (0.73% - 0.77%) |
| (I10, I25, I50) | 0.74% (0.72% - 0.76%) |
| (E11, I10, I63) | 0.73% (0.72% - 0.75%) |
| (E78, G47, I10) | 0.73% (0.71% - 0.75%) |
| (E78, I10, J40) | 0.73% (0.71% - 0.74%) |
| (E78, I25, K29) | 0.71% (0.69% - 0.73%) |
| (I10, J20, K29) | 0.70% (0.68% - 0.72%) |
| (I10, K29, M51) | 0.70% (0.68% - 0.72%) |
| (K29, M51, M54) | 0.70% (0.68% - 0.71%) |
| (I10, M51, M54) | 0.69% (0.68% - 0.71%) |
| (K29, M50, M51) | 0.69% (0.67% - 0.71%) |
| (I10, J40, M50) | 0.69% (0.67% - 0.71%) |
| (J40, K29, M50) | 0.68% (0.67% - 0.70%) |
| (E78, I10, K76) | 0.68% (0.66% - 0.70%) |
| (I10, K29, L30) | 0.66% (0.65% - 0.68%) |
| (I10, I25, J40) | 0.66% (0.65% - 0.68%) |
| (I10, I20, I25) | 0.66% (0.64% - 0.68%) |
| (K29, M50, M54) | 0.65% (0.63% - 0.67%) |
| (J40, K29, M54) | 0.65% (0.63% - 0.66%) |
| (I10, I25, M50) | 0.65% (0.63% - 0.66%) |
| (I10, M50, M51) | 0.64% (0.63% - 0.66%) |
| (G47, I10, I25) | 0.64% (0.63% - 0.66%) |
| (I10, I63, I67) | 0.64% (0.63% - 0.66%) |
| (I10, J40, M54) | 0.63% (0.61% - 0.65%) |
| (J20, J40, K29) | 0.63% (0.61% - 0.65%) |
| (M50, M51, M54) | 0.63% (0.61% - 0.64%) |
| (E78, I10, L30) | 0.63% (0.61% - 0.64%) |
| (E11, I10, J40) | 0.63% (0.61% - 0.64%) |
| (I10, K29, M25) | 0.62% (0.60% - 0.64%) |
| (E78, I10, M54) | 0.61% (0.59% - 0.63%) |
| (I10, M50, M54) | 0.61% (0.59% - 0.63%) |
| (E78, K29, M50) | 0.61% (0.59% - 0.62%) |
| (E78, I10, I70) | 0.60% (0.58% - 0.62%) |
| (I10, J20, J40) | 0.60% (0.58% - 0.61%) |
| (E78, I10, I67) | 0.59% (0.58% - 0.61%) |
| (E78, I10, M51) | 0.58% (0.56% - 0.60%) |

Supplementary Table S10. Prevalence of the 50 most common triads in individuals aged 60-69 years.

| Condition triads | Prevalence (95% Confidence interval) |
| --- | --- |
| (I10, I25, K29) | 3.12% (3.08% - 3.17%) |
| (I10, I25, I63) | 2.65% (2.61% - 2.69%) |
| (E78, I10, I25) | 2.49% (2.45% - 2.53%) |
| (E11, I10, I25) | 2.42% (2.38% - 2.46%) |
| (I10, I63, K29) | 2.29% (2.26% - 2.33%) |
| (E78, I10, I63) | 2.17% (2.14% - 2.21%) |
| (I10, I25, I50) | 2.17% (2.14% - 2.21%) |
| (E78, I10, K29) | 2.05% (2.02% - 2.09%) |
| (E11, E78, I10) | 1.97% (1.94% - 2.01%) |
| (E11, I10, I63) | 1.94% (1.90% - 1.97%) |
| (E11, I10, K29) | 1.84% (1.80% - 1.87%) |
| (I10, J40, K29) | 1.77% (1.74% - 1.80%) |
| (I10, I63, M50) | 1.73% (1.70% - 1.77%) |
| (I10, I20, I25) | 1.73% (1.70% - 1.76%) |
| (I10, K29, M50) | 1.64% (1.61% - 1.67%) |
| (I10, I63, I67) | 1.61% (1.58% - 1.64%) |
| (I10, I25, J40) | 1.57% (1.53% - 1.60%) |
| (I25, I63, K29) | 1.56% (1.53% - 1.59%) |
| (I25, I50, K29) | 1.52% (1.49% - 1.55%) |
| (G47, I10, I25) | 1.46% (1.43% - 1.49%) |
| (E78, I25, K29) | 1.45% (1.42% - 1.48%) |
| (I10, K29, M51) | 1.45% (1.42% - 1.48%) |
| (H81, I10, I63) | 1.44% (1.41% - 1.47%) |
| (I10, I25, M50) | 1.43% (1.40% - 1.46%) |
| (I10, I63, I70) | 1.42% (1.39% - 1.45%) |
| (I10, I50, K29) | 1.38% (1.35% - 1.41%) |
| (I10, K29, M54) | 1.35% (1.32% - 1.38%) |
| (I10, J20, K29) | 1.33% (1.30% - 1.36%) |
| (G47, I10, K29) | 1.33% (1.30% - 1.36%) |
| (G47, I10, I63) | 1.31% (1.28% - 1.34%) |
| (I10, I25, M51) | 1.31% (1.28% - 1.33%) |
| (I10, I25, I70) | 1.29% (1.27% - 1.32%) |
| (E78, I25, I63) | 1.29% (1.26% - 1.32%) |
| (I10, I25, I49) | 1.28% (1.25% - 1.31%) |
| (I10, I63, J40) | 1.28% (1.25% - 1.31%) |
| (I10, I63, M51) | 1.24% (1.21% - 1.27%) |
| (I10, M51, M54) | 1.23% (1.20% - 1.26%) |
| (I63, K29, M50) | 1.21% (1.18% - 1.23%) |
| (E78, I10, J40) | 1.20% (1.18% - 1.23%) |
| (E78, I10, I70) | 1.19% (1.16% - 1.22%) |
| (E78, I10, M50) | 1.19% (1.16% - 1.21%) |
| (I20, I25, I50) | 1.19% (1.16% - 1.21%) |
| (I10, I67, K29) | 1.18% (1.15% - 1.21%) |
| (I10, I25, J20) | 1.17% (1.14% - 1.19%) |
| (E78, G47, I10) | 1.16% (1.13% - 1.19%) |
| (E78, I63, K29) | 1.16% (1.13% - 1.19%) |
| (I10, I25, I67) | 1.15% (1.12% - 1.18%) |
| (I20, I25, K29) | 1.15% (1.12% - 1.17%) |
| (E11, I25, K29) | 1.14% (1.12% - 1.17%) |
| (I10, I25, M54) | 1.13% (1.10% - 1.16%) |

Supplementary Table S11. Prevalence of the 50 most common triads in individuals aged 70-79 years.

| Condition triads | Prevalence (95% Confidence interval) |
| --- | --- |
| (I10, I25, K29) | 6.15% (6.07% - 6.23%) |
| (I10, I25, I63) | 5.95% (5.88% - 6.03%) |
| (I10, I25, I50) | 5.46% (5.38% - 5.53%) |
| (I10, I63, K29) | 4.59% (4.52% - 4.66%) |
| (E11, I10, I25) | 4.13% (4.06% - 4.19%) |
| (I25, I50, K29) | 3.87% (3.80% - 3.93%) |
| (E78, I10, I25) | 3.83% (3.77% - 3.90%) |
| (I25, I63, K29) | 3.59% (3.53% - 3.65%) |
| (I10, I50, K29) | 3.55% (3.49% - 3.61%) |
| (E11, I10, I63) | 3.42% (3.36% - 3.48%) |
| (E78, I10, I63) | 3.38% (3.32% - 3.44%) |
| (I10, I20, I25) | 3.33% (3.27% - 3.39%) |
| (I10, I63, I67) | 3.19% (3.13% - 3.25%) |
| (I10, I50, I63) | 3.18% (3.13% - 3.24%) |
| (I10, I25, I49) | 3.04% (2.98% - 3.09%) |
| (I25, I50, I63) | 3.01% (2.95% - 3.06%) |
| (I10, I63, I70) | 3.00% (2.94% - 3.05%) |
| (I10, I25, J40) | 2.92% (2.87% - 2.98%) |
| (I10, I63, M50) | 2.86% (2.80% - 2.91%) |
| (E78, I10, K29) | 2.83% (2.78% - 2.89%) |
| (I10, I25, J44) | 2.81% (2.75% - 2.86%) |
| (I10, I25, I70) | 2.79% (2.73% - 2.84%) |
| (G47, I10, I25) | 2.77% (2.72% - 2.82%) |
| (E11, I10, K29) | 2.76% (2.71% - 2.82%) |
| (H81, I10, I63) | 2.72% (2.67% - 2.77%) |
| (I10, I25, M51) | 2.70% (2.65% - 2.76%) |
| (I10, J40, K29) | 2.68% (2.63% - 2.74%) |
| (I20, I25, I50) | 2.66% (2.61% - 2.72%) |
| (I10, I25, M50) | 2.56% (2.50% - 2.61%) |
| (I10, I25, I67) | 2.54% (2.49% - 2.59%) |
| (I10, K29, M51) | 2.52% (2.47% - 2.57%) |
| (I10, I63, M51) | 2.47% (2.42% - 2.53%) |
| (E11, E78, I10) | 2.44% (2.39% - 2.50%) |
| (I10, I25, J15) | 2.44% (2.39% - 2.49%) |
| (G47, I10, I63) | 2.43% (2.38% - 2.48%) |
| (I10, J44, K29) | 2.43% (2.38% - 2.48%) |
| (I10, K29, M50) | 2.40% (2.35% - 2.45%) |
| (E78, I25, I63) | 2.36% (2.31% - 2.41%) |
| (I10, I20, I50) | 2.35% (2.30% - 2.40%) |
| (I25, J44, K29) | 2.33% (2.28% - 2.38%) |
| (E11, I10, I50) | 2.33% (2.28% - 2.38%) |
| (I20, I25, K29) | 2.30% (2.25% - 2.35%) |
| (I25, I50, J44) | 2.29% (2.24% - 2.34%) |
| (I10, I63, J40) | 2.29% (2.24% - 2.34%) |
| (H81, I10, I25) | 2.27% (2.23% - 2.32%) |
| (I25, I49, I50) | 2.27% (2.22% - 2.32%) |
| (E78, I25, K29) | 2.25% (2.20% - 2.30%) |
| (I10, I25, J42) | 2.25% (2.20% - 2.30%) |
| (I10, I25, L30) | 2.22% (2.17% - 2.27%) |
| (I10, I25, J20) | 2.22% (2.17% - 2.27%) |

Supplementary Table S12. Prevalence of the 50 most common triads in individuals aged ≥ 80 years.

| Condition triads | Prevalence (95% Confidence interval) |
| --- | --- |
| (I10, I25, I50) | 9.47% (9.32% - 9.63%) |
| (I10, I25, K29) | 8.45% (8.30% - 8.60%) |
| (I10, I25, I63) | 8.03% (7.89% - 8.18%) |
| (I25, I50, K29) | 7.04% (6.90% - 7.18%) |
| (I10, I50, K29) | 6.43% (6.30% - 6.56%) |
| (I10, I63, K29) | 5.93% (5.80% - 6.05%) |
| (I10, I25, J44) | 5.92% (5.79% - 6.05%) |
| (I10, I50, I63) | 5.86% (5.74% - 5.99%) |
| (I25, I50, I63) | 5.66% (5.53% - 5.78%) |
| (I25, I50, J44) | 5.64% (5.52% - 5.77%) |
| (I10, I50, J44) | 5.23% (5.11% - 5.35%) |
| (I25, I63, K29) | 5.18% (5.07% - 5.30%) |
| (I10, I25, J15) | 4.96% (4.84% - 5.07%) |
| (I10, I25, I49) | 4.80% (4.69% - 4.92%) |
| (I25, J44, K29) | 4.76% (4.65% - 4.88%) |
| (I25, I50, J15) | 4.75% (4.64% - 4.87%) |
| (I10, I50, J15) | 4.71% (4.59% - 4.82%) |
| (I10, J44, K29) | 4.71% (4.59% - 4.82%) |
| (I25, I49, I50) | 4.51% (4.40% - 4.62%) |
| (I50, J44, K29) | 4.33% (4.22% - 4.44%) |
| (I10, I49, I50) | 4.26% (4.15% - 4.37%) |
| (G47, I10, I25) | 4.26% (4.15% - 4.37%) |
| (E11, I10, I25) | 4.19% (4.08% - 4.30%) |
| (I10, I63, J44) | 4.15% (4.04% - 4.25%) |
| (I10, I20, I25) | 4.14% (4.04% - 4.25%) |
| (I10, I63, J15) | 4.13% (4.02% - 4.24%) |
| (E87, I10, I25) | 4.09% (3.98% - 4.20%) |
| (E87, I10, I50) | 4.05% (3.95% - 4.16%) |
| (E87, I25, I50) | 4.04% (3.94% - 4.15%) |
| (I20, I25, I50) | 4.03% (3.93% - 4.14%) |
| (I50, I63, K29) | 4.01% (3.91% - 4.12%) |
| (I10, I25, K59) | 4.00% (3.89% - 4.10%) |
| (I10, J15, K29) | 3.92% (3.82% - 4.03%) |
| (I10, I25, J40) | 3.88% (3.78% - 3.98%) |
| (I10, I25, J42) | 3.82% (3.71% - 3.92%) |
| (E78, I10, I25) | 3.80% (3.70% - 3.90%) |
| (I25, I63, J44) | 3.69% (3.59% - 3.79%) |
| (I25, J15, K29) | 3.68% (3.58% - 3.78%) |
| (I10, I25, I70) | 3.63% (3.53% - 3.73%) |
| (I10, I63, I70) | 3.61% (3.51% - 3.71%) |
| (D64, I10, I25) | 3.59% (3.49% - 3.69%) |
| (I10, I63, I67) | 3.59% (3.49% - 3.69%) |
| (I10, K29, K59) | 3.57% (3.48% - 3.67%) |
| (I10, I20, I50) | 3.57% (3.47% - 3.67%) |
| (I25, I49, K29) | 3.54% (3.44% - 3.64%) |
| (I50, J15, K29) | 3.51% (3.41% - 3.61%) |
| (G47, I10, I63) | 3.49% (3.39% - 3.59%) |
| (I10, I25, L30) | 3.44% (3.34% - 3.54%) |
| (D64, I10, I50) | 3.41% (3.32% - 3.51%) |
| (I10, J15, J44) | 3.41% (3.31% - 3.51%) |

Supplementary Table S13. Multimorbidity coefficient (MMC), number of connections, and average partial correlation for the 100 highest-MMC conditions in the overall study population.

| Condition | MMC | Number of connections | Average partial correlation per connection |
| --- | --- | --- | --- |
| K29 | 2.47 | 340 | 0.005645 |
| E88 | 2.37 | 450 | 0.004571 |
| I50 | 2.30 | 284 | 0.006067 |
| D64 | 2.26 | 323 | 0.005797 |
| L30 | 2.21 | 293 | 0.006728 |
| E87 | 2.19 | 309 | 0.006120 |
| I70 | 2.18 | 298 | 0.005816 |
| J15 | 2.17 | 296 | 0.006280 |
| K76 | 2.09 | 313 | 0.005314 |
| I63 | 2.08 | 299 | 0.005001 |
| M81 | 2.07 | 315 | 0.005373 |
| G47 | 2.04 | 310 | 0.005761 |
| D61 | 1.94 | 255 | 0.005797 |
| I10 | 1.88 | 349 | 0.002669 |
| E78 | 1.86 | 325 | 0.004110 |
| J44 | 1.84 | 322 | 0.003550 |
| J31 | 1.82 | 263 | 0.005825 |
| I25 | 1.81 | 298 | 0.004166 |
| N18 | 1.80 | 240 | 0.005632 |
| I69 | 1.75 | 262 | 0.005012 |
| H16 | 1.75 | 257 | 0.005921 |
| F41 | 1.75 | 239 | 0.006054 |
| N40 | 1.75 | 303 | 0.004052 |
| M25 | 1.73 | 259 | 0.005719 |
| N28 | 1.72 | 266 | 0.005357 |
| N25 | 1.70 | 243 | 0.004435 |
| E04 | 1.70 | 303 | 0.003959 |
| J96 | 1.69 | 279 | 0.004934 |
| H25 | 1.69 | 268 | 0.003683 |
| N39 | 1.68 | 273 | 0.004926 |
| J20 | 1.67 | 328 | 0.002976 |
| J18 | 1.66 | 325 | 0.002905 |
| N92 | 1.65 | 242 | 0.004695 |
| D37 | 1.64 | 151 | 0.009489 |
| M50 | 1.64 | 274 | 0.004279 |
| I67 | 1.64 | 274 | 0.004283 |
| D70 | 1.61 | 256 | 0.005372 |
| K59 | 1.60 | 194 | 0.006378 |
| C79 | 1.60 | 273 | 0.004993 |
| M54 | 1.60 | 282 | 0.004383 |
| K92 | 1.60 | 256 | 0.005106 |
| K21 | 1.60 | 281 | 0.004437 |
| C78 | 1.59 | 171 | 0.006727 |
| K63 | 1.59 | 240 | 0.005044 |
| J32 | 1.59 | 197 | 0.006988 |
| I11 | 1.59 | 257 | 0.003849 |
| J94 | 1.57 | 284 | 0.004236 |
| E79 | 1.56 | 237 | 0.004755 |
| K52 | 1.55 | 252 | 0.005300 |
| I49 | 1.53 | 221 | 0.005756 |
| M51 | 1.52 | 255 | 0.004550 |
| M13 | 1.52 | 283 | 0.003330 |
| E03 | 1.50 | 262 | 0.004265 |
| N62 | 1.50 | 235 | 0.005495 |
| D68 | 1.49 | 246 | 0.004750 |
| B34 | 1.46 | 193 | 0.006550 |
| J40 | 1.45 | 252 | 0.004068 |
| N19 | 1.44 | 204 | 0.005648 |
| J42 | 1.44 | 272 | 0.003766 |
| N03 | 1.44 | 207 | 0.005282 |
| E11 | 1.44 | 282 | 0.003091 |
| N93 | 1.41 | 244 | 0.004009 |
| J06 | 1.40 | 218 | 0.004929 |
| A49 | 1.40 | 271 | 0.003686 |
| E14 | 1.40 | 292 | 0.003630 |
| N73 | 1.38 | 175 | 0.006222 |
| I48 | 1.37 | 221 | 0.003827 |
| C77 | 1.37 | 243 | 0.004686 |
| K74 | 1.36 | 185 | 0.005148 |
| N95 | 1.36 | 166 | 0.005930 |
| G62 | 1.36 | 287 | 0.003107 |
| K66 | 1.36 | 214 | 0.004351 |
| J30 | 1.35 | 197 | 0.005291 |
| M79 | 1.35 | 241 | 0.004869 |
| A16 | 1.35 | 267 | 0.003414 |
| G31 | 1.35 | 170 | 0.005757 |
| A16 | 1.34 | 218 | 0.005139 |
| I80 | 1.34 | 147 | 0.007673 |
| N83 | 1.34 | 261 | 0.003709 |
| M35 | 1.33 | 156 | 0.007487 |
| D48 | 1.32 | 206 | 0.004483 |
| I65 | 1.30 | 236 | 0.004050 |
| E83 | 1.29 | 232 | 0.003417 |
| I20 | 1.29 | 249 | 0.003773 |
| L23 | 1.29 | 150 | 0.005265 |
| C73 | 1.28 | 259 | 0.003026 |
| H52 | 1.26 | 238 | 0.004145 |
| I84 | 1.26 | 241 | 0.003804 |
| K12 | 1.26 | 210 | 0.004311 |
| N20 | 1.25 | 219 | 0.004623 |
| K05 | 1.25 | 228 | 0.004385 |
| D69 | 1.25 | 197 | 0.004655 |
| D25 | 1.24 | 203 | 0.004789 |
| K62 | 1.23 | 206 | 0.004505 |
| K31 | 1.23 | 192 | 0.003779 |
| I15 | 1.23 | 231 | 0.003662 |
| H81 | 1.23 | 188 | 0.004717 |
| K65 | 1.23 | 179 | 0.005203 |
| N72 | 1.23 | 176 | 0.005552 |
| G93 | 1.23 | 142 | 0.007089 |

Supplementary Table S14. Multimorbidity coefficient (MMC), number of connections, and average partial correlation for the 100 highest-MMC conditions female individuals.

| Condition | MMC | Number of connections | Average partial correlation per connection |
| --- | --- | --- | --- |
| K29 | 2.53 | 183 | 0.013286 |
| E88 | 2.31 | 171 | 0.013001 |
| I50 | 2.31 | 104 | 0.021466 |
| E87 | 2.22 | 193 | 0.010975 |
| M81 | 2.22 | 191 | 0.011119 |
| L30 | 2.21 | 197 | 0.010744 |
| I70 | 2.18 | 166 | 0.012475 |
| D64 | 2.18 | 174 | 0.011833 |
| I63 | 2.09 | 126 | 0.015711 |
| G47 | 2.09 | 218 | 0.008951 |
| J15 | 2.06 | 164 | 0.011926 |
| K76 | 2.05 | 179 | 0.010755 |
| D61 | 1.97 | 93 | 0.020155 |
| I10 | 1.94 | 145 | 0.012728 |
| E78 | 1.89 | 174 | 0.010279 |
| H16 | 1.88 | 179 | 0.009904 |
| J31 | 1.84 | 157 | 0.010893 |
| N18 | 1.82 | 94 | 0.018606 |
| F41 | 1.82 | 148 | 0.011413 |
| I25 | 1.79 | 106 | 0.015974 |
| M25 | 1.77 | 158 | 0.010463 |
| E04 | 1.74 | 160 | 0.010009 |
| I69 | 1.71 | 121 | 0.013089 |
| N25 | 1.70 | 77 | 0.020710 |
| H25 | 1.70 | 142 | 0.011346 |
| M50 | 1.69 | 121 | 0.012985 |
| C78 | 1.67 | 63 | 0.025062 |
| N92 | 1.67 | 124 | 0.012651 |
| N39 | 1.67 | 162 | 0.009535 |
| J20 | 1.67 | 131 | 0.012110 |
| M54 | 1.67 | 162 | 0.009662 |
| D70 | 1.66 | 170 | 0.008880 |
| N28 | 1.66 | 173 | 0.008843 |
| I67 | 1.64 | 129 | 0.012069 |
| D37 | 1.62 | 91 | 0.016514 |
| K21 | 1.61 | 153 | 0.009654 |
| M51 | 1.60 | 126 | 0.011754 |
| C79 | 1.60 | 89 | 0.016834 |
| K59 | 1.59 | 178 | 0.008046 |
| K52 | 1.59 | 176 | 0.008194 |
| J94 | 1.58 | 113 | 0.013035 |
| I11 | 1.58 | 107 | 0.013394 |
| J44 | 1.57 | 94 | 0.015635 |
| N62 | 1.57 | 153 | 0.009642 |
| J32 | 1.56 | 111 | 0.012621 |
| K63 | 1.55 | 103 | 0.013612 |
| J96 | 1.54 | 89 | 0.016484 |
| K92 | 1.54 | 149 | 0.009415 |
| M13 | 1.53 | 106 | 0.013849 |
| E03 | 1.52 | 135 | 0.010050 |
| D68 | 1.51 | 126 | 0.011257 |
| I49 | 1.51 | 137 | 0.009954 |
| J18 | 1.51 | 88 | 0.016406 |
| M35 | 1.51 | 128 | 0.010835 |
| N73 | 1.50 | 90 | 0.015555 |
| J40 | 1.50 | 116 | 0.012125 |
| B34 | 1.50 | 120 | 0.011286 |
| I48 | 1.48 | 76 | 0.018297 |
| N19 | 1.48 | 111 | 0.012268 |
| E79 | 1.47 | 143 | 0.009352 |
| N95 | 1.46 | 154 | 0.008722 |
| N03 | 1.46 | 97 | 0.013244 |
| E11 | 1.45 | 107 | 0.012468 |
| M79 | 1.44 | 175 | 0.007340 |
| K66 | 1.43 | 93 | 0.013919 |
| J42 | 1.43 | 127 | 0.010204 |
| J06 | 1.41 | 127 | 0.010349 |
| N93 | 1.41 | 112 | 0.011495 |
| N83 | 1.40 | 86 | 0.014623 |
| E14 | 1.39 | 109 | 0.011669 |
| I80 | 1.39 | 135 | 0.009310 |
| A49 | 1.38 | 167 | 0.007519 |
| D48 | 1.35 | 92 | 0.013011 |
| I20 | 1.35 | 112 | 0.010838 |
| E83 | 1.34 | 118 | 0.010246 |
| G62 | 1.34 | 161 | 0.007284 |
| K74 | 1.33 | 56 | 0.021926 |
| D25 | 1.33 | 98 | 0.012508 |
| J30 | 1.33 | 107 | 0.011260 |
| C73 | 1.33 | 34 | 0.035416 |
| A16 | 1.33 | 43 | 0.028176 |
| L23 | 1.32 | 120 | 0.010040 |
| C77 | 1.31 | 72 | 0.016880 |
| K31 | 1.31 | 103 | 0.011290 |
| K12 | 1.31 | 111 | 0.010807 |
| I84 | 1.30 | 137 | 0.008473 |
| G31 | 1.30 | 119 | 0.010017 |
| H52 | 1.28 | 110 | 0.010844 |
| M17 | 1.28 | 106 | 0.010965 |
| I65 | 1.28 | 89 | 0.013331 |
| I15 | 1.27 | 55 | 0.020815 |
| J37 | 1.27 | 104 | 0.010920 |
| N85 | 1.26 | 83 | 0.013995 |
| N63 | 1.26 | 91 | 0.012502 |
| K05 | 1.26 | 123 | 0.009154 |
| H81 | 1.26 | 102 | 0.010792 |
| M48 | 1.25 | 119 | 0.009569 |
| G93 | 1.25 | 94 | 0.011413 |
| C95 | 1.24 | 36 | 0.030265 |
| N20 | 1.24 | 105 | 0.010295 |

Supplementary Table S15. Multimorbidity coefficient (MMC), number of connections, and average partial correlation for the 100 highest-MMC conditions in male individuals.

| Condition | MMC | Number of connections | Average partial correlation per connection |
| --- | --- | --- | --- |
| K29 | 2.37 | 172 | 0.013320 |
| E88 | 2.37 | 167 | 0.013695 |
| I50 | 2.35 | 107 | 0.021145 |
| D64 | 2.26 | 173 | 0.012445 |
| J15 | 2.26 | 180 | 0.011926 |
| I70 | 2.23 | 165 | 0.012713 |
| E87 | 2.18 | 183 | 0.011339 |
| L30 | 2.14 | 184 | 0.011038 |
| K76 | 2.11 | 175 | 0.011312 |
| I63 | 2.08 | 124 | 0.015964 |
| N40 | 2.05 | 159 | 0.012068 |
| J44 | 1.97 | 114 | 0.016527 |
| G47 | 1.93 | 208 | 0.008555 |
| I25 | 1.87 | 112 | 0.015906 |
| E78 | 1.83 | 165 | 0.010458 |
| J31 | 1.82 | 148 | 0.011563 |
| I69 | 1.82 | 114 | 0.014794 |
| J96 | 1.82 | 95 | 0.018192 |
| I10 | 1.81 | 133 | 0.013063 |
| N18 | 1.81 | 88 | 0.019476 |
| N25 | 1.80 | 86 | 0.019739 |
| J18 | 1.80 | 109 | 0.015761 |
| D37 | 1.80 | 82 | 0.020254 |
| N28 | 1.79 | 161 | 0.010283 |
| D61 | 1.79 | 94 | 0.017908 |
| H25 | 1.74 | 149 | 0.011009 |
| M81 | 1.73 | 160 | 0.010028 |
| F41 | 1.71 | 145 | 0.010703 |
| M25 | 1.68 | 149 | 0.010462 |
| J20 | 1.68 | 122 | 0.013272 |
| K63 | 1.68 | 104 | 0.014853 |
| I11 | 1.67 | 105 | 0.014611 |
| I67 | 1.66 | 130 | 0.01207 |
| E79 | 1.66 | 162 | 0.009329 |
| K92 | 1.66 | 145 | 0.010475 |
| J32 | 1.65 | 112 | 0.013359 |
| C79 | 1.63 | 76 | 0.019986 |
| K59 | 1.63 | 167 | 0.008862 |
| C78 | 1.62 | 57 | 0.026822 |
| J94 | 1.60 | 106 | 0.014361 |
| K21 | 1.60 | 154 | 0.009557 |
| I49 | 1.58 | 139 | 0.010394 |
| N39 | 1.58 | 119 | 0.012410 |
| H16 | 1.55 | 149 | 0.009433 |
| K52 | 1.55 | 171 | 0.008302 |
| M13 | 1.54 | 95 | 0.015317 |
| M50 | 1.52 | 115 | 0.012314 |
| D70 | 1.51 | 153 | 0.008880 |
| M54 | 1.51 | 132 | 0.010734 |
| J42 | 1.51 | 148 | 0.009392 |
| C77 | 1.51 | 71 | 0.019765 |
| N19 | 1.48 | 107 | 0.012817 |
| E04 | 1.48 | 139 | 0.009581 |
| N03 | 1.48 | 107 | 0.012497 |
| E11 | 1.47 | 105 | 0.012926 |
| E14 | 1.46 | 99 | 0.013490 |
| G31 | 1.46 | 129 | 0.010361 |
| J06 | 1.43 | 117 | 0.011105 |
| M51 | 1.42 | 119 | 0.011038 |
| J40 | 1.42 | 103 | 0.013091 |
| G62 | 1.41 | 165 | 0.007727 |
| I65 | 1.41 | 89 | 0.014512 |
| K74 | 1.41 | 49 | 0.026628 |
| N41 | 1.40 | 132 | 0.009856 |
| A16 | 1.39 | 48 | 0.027002 |
| A49 | 1.39 | 168 | 0.007503 |
| J30 | 1.38 | 106 | 0.012029 |
| I80 | 1.36 | 125 | 0.009805 |
| E03 | 1.36 | 159 | 0.007585 |
| I48 | 1.35 | 83 | 0.014905 |
| D69 | 1.35 | 140 | 0.008576 |
| K65 | 1.33 | 90 | 0.013811 |
| H35 | 1.33 | 88 | 0.013508 |
| B18 | 1.32 | 75 | 0.015490 |
| K62 | 1.32 | 110 | 0.010619 |
| I20 | 1.31 | 106 | 0.011116 |
| J43 | 1.31 | 119 | 0.009911 |
| D68 | 1.30 | 122 | 0.009822 |
| K12 | 1.30 | 115 | 0.010448 |
| G93 | 1.30 | 102 | 0.011471 |
| K05 | 1.30 | 122 | 0.009437 |
| L23 | 1.27 | 115 | 0.010237 |
| E46 | 1.27 | 152 | 0.007652 |
| M79 | 1.27 | 157 | 0.007149 |
| L08 | 1.27 | 123 | 0.009416 |
| H52 | 1.27 | 107 | 0.010934 |
| I15 | 1.27 | 52 | 0.021937 |
| C34 | 1.27 | 47 | 0.025190 |
| J35 | 1.27 | 70 | 0.016953 |
| I34 | 1.26 | 61 | 0.018481 |
| M48 | 1.26 | 115 | 0.009737 |
| D41 | 1.25 | 52 | 0.021274 |
| I27 | 1.25 | 64 | 0.017887 |
| N20 | 1.24 | 92 | 0.012207 |
| I84 | 1.24 | 133 | 0.008277 |
| E83 | 1.24 | 108 | 0.010080 |
| A41 | 1.23 | 122 | 0.009271 |
| K30 | 1.23 | 121 | 0.009149 |
| C73 | 1.22 | 38 | 0.028801 |
| H02 | 1.22 | 122 | 0.008328 |

Supplementary Table S16. Multimorbidity coefficient (MMC), number of connections, and average partial correlation for the 100 highest-MMC conditions in individuals aged 1-9 years.

| Condition | MMC | Number of connections | Average partial correlation per connection |
| --- | --- | --- | --- |
| J31 | 1.76 | 67 | 0.022395 |
| L30 | 1.64 | 76 | 0.018165 |
| J18 | 1.53 | 49 | 0.028118 |
| J35 | 1.44 | 27 | 0.042668 |
| J32 | 1.44 | 43 | 0.026717 |
| J30 | 1.43 | 48 | 0.023842 |
| K52 | 1.42 | 66 | 0.017094 |
| J06 | 1.39 | 62 | 0.018806 |
| J20 | 1.34 | 46 | 0.024547 |
| A49 | 1.34 | 41 | 0.026589 |
| J03 | 1.32 | 47 | 0.023519 |
| J15 | 1.30 | 39 | 0.027888 |
| I88 | 1.28 | 66 | 0.015163 |
| E87 | 1.22 | 49 | 0.019832 |
| A41 | 1.14 | 44 | 0.019495 |
| J21 | 1.11 | 33 | 0.025819 |
| H52 | 1.10 | 50 | 0.017518 |
| K02 | 1.10 | 36 | 0.022881 |
| K59 | 1.07 | 49 | 0.01568 |
| H65 | 1.05 | 25 | 0.029571 |
| H61 | 1.01 | 52 | 0.013742 |
| K30 | 1.01 | 47 | 0.015871 |
| M25 | 1.01 | 49 | 0.013822 |
| J45 | 1.01 | 30 | 0.024206 |
| L50 | 0.99 | 48 | 0.015292 |
| K00 | 0.99 | 41 | 0.016738 |
| E55 | 0.98 | 45 | 0.017590 |
| J40 | 0.96 | 37 | 0.019224 |
| C91 | 0.95 | 9 | 0.094623 |
| D64 | 0.95 | 38 | 0.018189 |
| D70 | 0.95 | 44 | 0.015344 |
| B34 | 0.94 | 34 | 0.022789 |
| K29 | 0.92 | 39 | 0.014217 |
| M79 | 0.90 | 50 | 0.011645 |
| L28 | 0.89 | 42 | 0.013633 |
| K04 | 0.87 | 22 | 0.027279 |
| C95 | 0.87 | 12 | 0.067872 |
| H02 | 0.84 | 25 | 0.021958 |
| E34 | 0.82 | 18 | 0.028268 |
| E86 | 0.82 | 21 | 0.028696 |
| K12 | 0.81 | 35 | 0.013770 |
| L23 | 0.80 | 38 | 0.013945 |
| J10 | 0.80 | 28 | 0.022039 |
| H66 | 0.78 | 24 | 0.024204 |
| H16 | 0.78 | 28 | 0.016514 |
| A09 | 0.78 | 24 | 0.023514 |
| J34 | 0.77 | 25 | 0.019552 |
| N28 | 0.76 | 16 | 0.038646 |
| B08 | 0.76 | 30 | 0.017451 |
| J13 | 0.75 | 19 | 0.028995 |
| K35 | 0.75 | 16 | 0.033880 |
| K58 | 0.75 | 39 | 0.011946 |
| L08 | 0.73 | 17 | 0.031723 |
| N47 | 0.73 | 30 | 0.015433 |
| E06 | 0.72 | 18 | 0.030877 |
| H10 | 0.71 | 36 | 0.013138 |
| N48 | 0.71 | 25 | 0.015542 |
| N04 | 0.70 | 12 | 0.049583 |
| B35 | 0.70 | 30 | 0.013863 |
| J12 | 0.69 | 18 | 0.030098 |
| L20 | 0.69 | 26 | 0.016594 |
| I51 | 0.69 | 23 | 0.017900 |
| E58 | 0.68 | 26 | 0.019280 |
| E23 | 0.68 | 14 | 0.035250 |
| K92 | 0.66 | 28 | 0.015985 |
| B49 | 0.66 | 31 | 0.013508 |
| K08 | 0.66 | 20 | 0.021411 |
| J14 | 0.66 | 23 | 0.021545 |
| J98 | 0.63 | 19 | 0.020907 |
| H53 | 0.63 | 13 | 0.024205 |
| I49 | 0.63 | 20 | 0.019011 |
| D69 | 0.63 | 28 | 0.013767 |
| K13 | 0.63 | 26 | 0.012026 |
| J04 | 0.62 | 22 | 0.017044 |
| E83 | 0.62 | 21 | 0.016095 |
| K07 | 0.62 | 18 | 0.022129 |
| E56 | 0.62 | 27 | 0.015147 |
| J38 | 0.62 | 8 | 0.047939 |
| N39 | 0.62 | 25 | 0.014053 |
| L21 | 0.61 | 32 | 0.011340 |
| A08 | 0.61 | 25 | 0.018153 |
| L25 | 0.61 | 27 | 0.011486 |
| L29 | 0.60 | 27 | 0.011966 |
| K56 | 0.59 | 24 | 0.017286 |
| E04 | 0.59 | 25 | 0.017240 |
| H60 | 0.59 | 17 | 0.018769 |
| N05 | 0.59 | 19 | 0.024982 |
| G80 | 0.59 | 18 | 0.025987 |
| L04 | 0.58 | 23 | 0.018096 |
| L81 | 0.58 | 18 | 0.016223 |
| H91 | 0.58 | 19 | 0.022005 |
| N00 | 0.58 | 19 | 0.025337 |
| B00 | 0.58 | 24 | 0.014828 |
| M89 | 0.58 | 22 | 0.015817 |
| J37 | 0.58 | 15 | 0.022351 |
| L90 | 0.58 | 29 | 0.011298 |
| D22 | 0.58 | 24 | 0.010636 |
| K60 | 0.58 | 21 | 0.013994 |
| M65 | 0.57 | 19 | 0.017492 |
| M60 | 0.57 | 22 | 0.014488 |

Supplementary Table S17. Multimorbidity coefficient (MMC), number of connections, and average partial correlation for the 100 highest-MMC conditions in individuals aged 10-19 years.

| Condition | MMC | Number of connections | Average partial correlation per connection |
| --- | --- | --- | --- |
| C95 | 1.64 | 68 | 0.021805 |
| I61 | 1.61 | 81 | 0.017451 |
| M25 | 1.57 | 73 | 0.016738 |
| E79 | 1.52 | 69 | 0.016207 |
| D64 | 1.51 | 52 | 0.022645 |
| K29 | 1.49 | 71 | 0.015634 |
| E87 | 1.49 | 74 | 0.015035 |
| F32 | 1.47 | 45 | 0.025031 |
| J32 | 1.46 | 29 | 0.036149 |
| L30 | 1.46 | 67 | 0.016255 |
| J15 | 1.45 | 49 | 0.024974 |
| E11 | 1.45 | 48 | 0.022985 |
| F41 | 1.40 | 52 | 0.020491 |
| G47 | 1.39 | 56 | 0.017224 |
| M81 | 1.38 | 56 | 0.01878 |
| J31 | 1.38 | 50 | 0.020583 |
| M54 | 1.37 | 64 | 0.014938 |
| K52 | 1.37 | 66 | 0.014230 |
| E88 | 1.36 | 63 | 0.018197 |
| J06 | 1.35 | 68 | 0.015252 |
| A49 | 1.35 | 48 | 0.020533 |
| J44 | 1.30 | 80 | 0.013251 |
| M32 | 1.29 | 29 | 0.038729 |
| E78 | 1.28 | 49 | 0.019010 |
| J03 | 1.27 | 42 | 0.023810 |
| D61 | 1.27 | 53 | 0.020893 |
| H16 | 1.26 | 59 | 0.014240 |
| J30 | 1.24 | 36 | 0.024512 |
| N92 | 1.24 | 34 | 0.027737 |
| D70 | 1.23 | 63 | 0.014406 |
| M50 | 1.21 | 66 | 0.012666 |
| K59 | 1.20 | 45 | 0.015704 |
| L93 | 1.19 | 34 | 0.029428 |
| J45 | 1.18 | 43 | 0.018170 |
| N03 | 1.17 | 29 | 0.032734 |
| L08 | 1.16 | 45 | 0.018179 |
| I10 | 1.15 | 52 | 0.015198 |
| N19 | 1.15 | 52 | 0.016428 |
| E14 | 1.14 | 46 | 0.020153 |
| I49 | 1.14 | 40 | 0.018096 |
| K76 | 1.13 | 45 | 0.018105 |
| M79 | 1.13 | 51 | 0.013287 |
| F20 | 1.12 | 29 | 0.029776 |
| F45 | 1.11 | 60 | 0.012539 |
| K35 | 1.11 | 31 | 0.026349 |
| K21 | 1.11 | 56 | 0.013013 |
| D50 | 1.10 | 35 | 0.021530 |
| E03 | 1.10 | 37 | 0.021568 |
| I88 | 1.09 | 53 | 0.013565 |
| K66 | 1.09 | 39 | 0.021738 |
| N83 | 1.09 | 33 | 0.024706 |
| I50 | 1.09 | 49 | 0.014366 |
| D68 | 1.09 | 59 | 0.014282 |
| J94 | 1.08 | 46 | 0.017675 |
| E16 | 1.08 | 35 | 0.020056 |
| K71 | 1.08 | 53 | 0.015728 |
| N28 | 1.07 | 48 | 0.017082 |
| I51 | 1.07 | 37 | 0.018574 |
| A18 | 1.07 | 55 | 0.014636 |
| E83 | 1.06 | 47 | 0.016050 |
| N04 | 1.06 | 22 | 0.039502 |
| F31 | 1.05 | 23 | 0.032081 |
| N39 | 1.05 | 54 | 0.014036 |
| A16 | 1.05 | 15 | 0.059380 |
| N02 | 1.04 | 30 | 0.030384 |
| J47 | 1.03 | 51 | 0.015505 |
| M47 | 1.03 | 49 | 0.014098 |
| J18 | 1.02 | 28 | 0.028878 |
| E04 | 1.02 | 25 | 0.026223 |
| D69 | 1.02 | 48 | 0.013654 |
| K12 | 1.01 | 54 | 0.011307 |
| K30 | 1.01 | 50 | 0.013574 |
| N05 | 1.01 | 28 | 0.029594 |
| M35 | 1.00 | 52 | 0.014629 |
| H65 | 1.00 | 33 | 0.018919 |
| K92 | 1.00 | 44 | 0.015436 |
| I25 | 0.99 | 59 | 0.011669 |
| H53 | 0.99 | 44 | 0.012592 |
| C91 | 0.99 | 24 | 0.035276 |
| M65 | 0.99 | 43 | 0.014362 |
| N73 | 0.98 | 39 | 0.017921 |
| I40 | 0.98 | 43 | 0.013871 |
| M13 | 0.98 | 47 | 0.013004 |
| J35 | 0.97 | 31 | 0.023882 |
| K50 | 0.97 | 41 | 0.018882 |
| I24 | 0.96 | 56 | 0.013712 |
| A09 | 0.95 | 45 | 0.015184 |
| E28 | 0.95 | 16 | 0.043883 |
| E23 | 0.94 | 24 | 0.028186 |
| E10 | 0.94 | 15 | 0.050411 |
| D48 | 0.94 | 33 | 0.020679 |
| J37 | 0.94 | 27 | 0.022729 |
| K05 | 0.94 | 24 | 0.018881 |
| L23 | 0.93 | 42 | 0.013590 |
| H93 | 0.93 | 35 | 0.015987 |
| I63 | 0.93 | 55 | 0.012088 |
| B99 | 0.92 | 45 | 0.012894 |
| I67 | 0.92 | 49 | 0.011400 |
| J20 | 0.92 | 37 | 0.019409 |
| H81 | 0.92 | 48 | 0.010841 |

Supplementary Table S18. Multimorbidity coefficient (MMC), number of connections, and average partial correlation for the 100 highest-MMC conditions in individuals aged 20-29 years.

| Condition | MMC | Number of connections | Average partial correlation per connection |
| --- | --- | --- | --- |
| K29 | 1.84 | 100 | 0.015268 |
| L30 | 1.83 | 118 | 0.013365 |
| E87 | 1.64 | 110 | 0.012425 |
| N92 | 1.63 | 66 | 0.020544 |
| G47 | 1.62 | 93 | 0.013920 |
| J15 | 1.59 | 73 | 0.018411 |
| N25 | 1.56 | 58 | 0.022704 |
| J31 | 1.56 | 75 | 0.016132 |
| I10 | 1.55 | 75 | 0.016777 |
| I50 | 1.52 | 64 | 0.020489 |
| M25 | 1.48 | 75 | 0.014857 |
| M54 | 1.47 | 84 | 0.012813 |
| N18 | 1.47 | 39 | 0.032699 |
| A16 | 1.45 | 20 | 0.064098 |
| D64 | 1.43 | 65 | 0.018733 |
| F41 | 1.42 | 52 | 0.021277 |
| H16 | 1.42 | 83 | 0.013108 |
| M81 | 1.42 | 69 | 0.016321 |
| B34 | 1.42 | 62 | 0.017318 |
| E78 | 1.41 | 66 | 0.016521 |
| K76 | 1.41 | 64 | 0.016676 |
| K59 | 1.41 | 90 | 0.011466 |
| E88 | 1.41 | 81 | 0.014032 |
| E04 | 1.38 | 72 | 0.014841 |
| E79 | 1.37 | 61 | 0.016236 |
| N83 | 1.37 | 38 | 0.027468 |
| K52 | 1.36 | 69 | 0.013383 |
| D68 | 1.35 | 60 | 0.018429 |
| N85 | 1.35 | 53 | 0.021439 |
| I15 | 1.35 | 41 | 0.02821 |
| M32 | 1.33 | 35 | 0.033852 |
| N03 | 1.33 | 39 | 0.026562 |
| N93 | 1.33 | 57 | 0.017939 |
| J32 | 1.32 | 35 | 0.027863 |
| N73 | 1.31 | 48 | 0.022624 |
| M50 | 1.30 | 74 | 0.013167 |
| K66 | 1.30 | 45 | 0.023800 |
| M35 | 1.30 | 61 | 0.016383 |
| C73 | 1.30 | 18 | 0.060463 |
| K21 | 1.30 | 56 | 0.016304 |
| D70 | 1.3 | 65 | 0.014500 |
| L93 | 1.28 | 52 | 0.018674 |
| N62 | 1.28 | 69 | 0.013848 |
| E16 | 1.27 | 42 | 0.022334 |
| D61 | 1.26 | 48 | 0.020717 |
| E11 | 1.24 | 48 | 0.019732 |
| F32 | 1.24 | 37 | 0.023584 |
| E83 | 1.23 | 59 | 0.016110 |
| E03 | 1.23 | 43 | 0.021838 |
| M13 | 1.22 | 70 | 0.013387 |
| J40 | 1.22 | 76 | 0.011854 |
| I11 | 1.19 | 50 | 0.019328 |
| C77 | 1.17 | 34 | 0.027986 |
| N19 | 1.15 | 45 | 0.020542 |
| N63 | 1.15 | 41 | 0.02165 |
| E28 | 1.15 | 28 | 0.031491 |
| J37 | 1.14 | 32 | 0.025095 |
| J20 | 1.13 | 71 | 0.011565 |
| L23 | 1.12 | 76 | 0.011268 |
| N41 | 1.12 | 55 | 0.015913 |
| N28 | 1.12 | 61 | 0.012548 |
| N97 | 1.11 | 29 | 0.033827 |
| K92 | 1.11 | 49 | 0.015217 |
| J94 | 1.11 | 52 | 0.016005 |
| N80 | 1.09 | 31 | 0.026473 |
| N72 | 1.09 | 57 | 0.014713 |
| K65 | 1.09 | 46 | 0.018359 |
| K12 | 1.08 | 65 | 0.011205 |
| J30 | 1.08 | 39 | 0.019614 |
| K30 | 1.07 | 61 | 0.01178 |
| J06 | 1.07 | 58 | 0.012665 |
| K01 | 1.06 | 56 | 0.013929 |
| N94 | 1.06 | 71 | 0.010653 |
| M79 | 1.05 | 68 | 0.009926 |
| H47 | 1.05 | 52 | 0.013982 |
| K71 | 1.04 | 56 | 0.012978 |
| K05 | 1.04 | 53 | 0.012862 |
| F31 | 1.03 | 29 | 0.027328 |
| N39 | 1.03 | 59 | 0.011841 |
| E27 | 1.03 | 56 | 0.012188 |
| E14 | 1.02 | 39 | 0.019991 |
| I84 | 1.01 | 32 | 0.020037 |
| N87 | 1.01 | 23 | 0.034351 |
| E07 | 1.01 | 27 | 0.025553 |
| H93 | 1.00 | 46 | 0.014923 |
| J45 | 1.00 | 36 | 0.018060 |
| K62 | 1.00 | 41 | 0.016069 |
| D35 | 1.00 | 28 | 0.026231 |
| G93 | 1.00 | 62 | 0.010997 |
| J34 | 1.00 | 20 | 0.033317 |
| I69 | 1.00 | 47 | 0.016086 |
| N88 | 0.99 | 34 | 0.021638 |
| N84 | 0.99 | 44 | 0.015365 |
| A49 | 0.99 | 61 | 0.010117 |
| I49 | 0.98 | 47 | 0.012796 |
| N91 | 0.98 | 33 | 0.020680 |
| F48 | 0.98 | 52 | 0.012201 |
| N20 | 0.98 | 27 | 0.027580 |
| F45 | 0.97 | 52 | 0.012375 |
| E89 | 0.97 | 27 | 0.028274 |

Supplementary Table S19. Multimorbidity coefficient (MMC), number of connections, and average partial correlation for the 100 highest-MMC conditions in individuals aged 30-39 years.

| Condition | MMC | Number of connections | Average partial correlation per connection |
| --- | --- | --- | --- |
| I50 | 2.30 | 81 | 0.025594 |
| K29 | 2.13 | 120 | 0.015280 |
| L30 | 2.04 | 140 | 0.012570 |
| E87 | 1.95 | 129 | 0.013037 |
| G47 | 1.88 | 130 | 0.011792 |
| D64 | 1.84 | 93 | 0.016793 |
| E88 | 1.83 | 103 | 0.015400 |
| E78 | 1.81 | 93 | 0.016110 |
| J15 | 1.74 | 82 | 0.017661 |
| J31 | 1.74 | 101 | 0.014209 |
| D61 | 1.74 | 65 | 0.023348 |
| I10 | 1.72 | 93 | 0.016234 |
| K76 | 1.71 | 111 | 0.013287 |
| N18 | 1.68 | 44 | 0.033840 |
| M25 | 1.65 | 103 | 0.013016 |
| N25 | 1.63 | 46 | 0.031227 |
| H16 | 1.58 | 106 | 0.011902 |
| N92 | 1.57 | 77 | 0.017250 |
| F41 | 1.55 | 69 | 0.018195 |
| M81 | 1.54 | 99 | 0.013067 |
| D68 | 1.53 | 66 | 0.019971 |
| E04 | 1.53 | 81 | 0.014761 |
| K52 | 1.52 | 93 | 0.012378 |
| J32 | 1.52 | 70 | 0.016997 |
| M50 | 1.52 | 93 | 0.013147 |
| I15 | 1.51 | 45 | 0.028106 |
| K59 | 1.49 | 105 | 0.010940 |
| M54 | 1.48 | 99 | 0.011911 |
| N73 | 1.47 | 62 | 0.020491 |
| D70 | 1.47 | 88 | 0.013122 |
| M35 | 1.43 | 69 | 0.017544 |
| I11 | 1.43 | 41 | 0.030498 |
| E79 | 1.42 | 68 | 0.016148 |
| N62 | 1.42 | 82 | 0.013747 |
| C73 | 1.41 | 20 | 0.058541 |
| B34 | 1.39 | 69 | 0.016266 |
| J40 | 1.39 | 89 | 0.012360 |
| C77 | 1.39 | 40 | 0.029090 |
| K21 | 1.38 | 74 | 0.014776 |
| N83 | 1.38 | 47 | 0.024016 |
| N03 | 1.37 | 43 | 0.026169 |
| A16 | 1.37 | 21 | 0.056003 |
| E83 | 1.36 | 72 | 0.015552 |
| I25 | 1.34 | 60 | 0.017682 |
| N19 | 1.33 | 57 | 0.019291 |
| E11 | 1.33 | 70 | 0.015230 |
| M13 | 1.32 | 76 | 0.014009 |
| J94 | 1.32 | 80 | 0.013567 |
| I70 | 1.32 | 78 | 0.013217 |
| N85 | 1.31 | 55 | 0.019910 |
| I69 | 1.31 | 42 | 0.025112 |
| E03 | 1.31 | 58 | 0.017011 |
| J96 | 1.30 | 69 | 0.016179 |
| N93 | 1.29 | 62 | 0.016547 |
| C78 | 1.29 | 51 | 0.021865 |
| N80 | 1.28 | 34 | 0.030929 |
| K66 | 1.28 | 52 | 0.020403 |
| K65 | 1.28 | 63 | 0.016574 |
| N63 | 1.28 | 45 | 0.022124 |
| N41 | 1.27 | 75 | 0.013901 |
| M32 | 1.26 | 47 | 0.023354 |
| E21 | 1.25 | 45 | 0.023438 |
| C79 | 1.25 | 51 | 0.020949 |
| K63 | 1.24 | 47 | 0.020699 |
| N72 | 1.24 | 64 | 0.015595 |
| J20 | 1.24 | 95 | 0.010172 |
| I49 | 1.24 | 66 | 0.012217 |
| K92 | 1.24 | 66 | 0.013838 |
| I61 | 1.23 | 41 | 0.025298 |
| A49 | 1.23 | 95 | 0.009497 |
| G81 | 1.22 | 48 | 0.022305 |
| L23 | 1.21 | 84 | 0.011374 |
| J37 | 1.21 | 32 | 0.027139 |
| N97 | 1.20 | 37 | 0.027865 |
| E16 | 1.20 | 48 | 0.018665 |
| I63 | 1.20 | 54 | 0.016981 |
| M79 | 1.20 | 94 | 0.008938 |
| N20 | 1.19 | 39 | 0.022835 |
| I38 | 1.19 | 29 | 0.034416 |
| K12 | 1.19 | 78 | 0.011233 |
| F32 | 1.18 | 47 | 0.018578 |
| G93 | 1.17 | 74 | 0.011772 |
| N28 | 1.17 | 61 | 0.013703 |
| N88 | 1.17 | 51 | 0.018025 |
| N39 | 1.17 | 72 | 0.012031 |
| J30 | 1.17 | 51 | 0.016522 |
| J06 | 1.16 | 76 | 0.011661 |
| I84 | 1.16 | 54 | 0.014771 |
| I67 | 1.16 | 70 | 0.012231 |
| K05 | 1.15 | 69 | 0.011828 |
| E14 | 1.15 | 56 | 0.016663 |
| H35 | 1.15 | 38 | 0.023916 |
| H93 | 1.14 | 56 | 0.013927 |
| E28 | 1.14 | 37 | 0.022765 |
| N87 | 1.12 | 22 | 0.041357 |
| K30 | 1.12 | 80 | 0.010398 |
| K62 | 1.12 | 57 | 0.014011 |
| J45 | 1.11 | 50 | 0.015759 |
| D48 | 1.11 | 45 | 0.018235 |
| D35 | 1.10 | 38 | 0.022366 |

Supplementary Table S20. Multimorbidity coefficient (MMC), number of connections, and average partial correlation for the 100 highest-MMC conditions in individuals aged 40-49 years.

| Condition | MMC | Number of connections | Average partial correlation per connection |
| --- | --- | --- | --- |
| K29 | 2.40 | 142 | 0.015395 |
| I50 | 2.23 | 77 | 0.026568 |
| E87 | 2.17 | 134 | 0.014168 |
| D64 | 2.14 | 110 | 0.017316 |
| K76 | 2.08 | 124 | 0.014316 |
| L30 | 2.08 | 140 | 0.013060 |
| E88 | 2.07 | 121 | 0.015372 |
| D61 | 2.01 | 79 | 0.023021 |
| J15 | 2.01 | 123 | 0.014256 |
| G47 | 1.97 | 134 | 0.011983 |
| E78 | 1.88 | 118 | 0.013365 |
| M81 | 1.79 | 120 | 0.012833 |
| M25 | 1.75 | 114 | 0.012842 |
| J31 | 1.75 | 105 | 0.013762 |
| I10 | 1.72 | 93 | 0.016066 |
| N18 | 1.71 | 52 | 0.029102 |
| M50 | 1.69 | 98 | 0.014497 |
| N73 | 1.67 | 76 | 0.019445 |
| I70 | 1.67 | 100 | 0.014181 |
| H16 | 1.67 | 115 | 0.012006 |
| F41 | 1.65 | 86 | 0.015631 |
| N92 | 1.64 | 91 | 0.015700 |
| N25 | 1.63 | 44 | 0.032898 |
| E04 | 1.62 | 90 | 0.014874 |
| D70 | 1.61 | 92 | 0.014118 |
| C78 | 1.60 | 51 | 0.027824 |
| J96 | 1.58 | 75 | 0.018895 |
| I11 | 1.55 | 54 | 0.024885 |
| E83 | 1.54 | 85 | 0.015064 |
| I63 | 1.53 | 63 | 0.020208 |
| E79 | 1.53 | 83 | 0.014655 |
| I25 | 1.52 | 65 | 0.019547 |
| C77 | 1.51 | 50 | 0.025683 |
| K52 | 1.50 | 99 | 0.011856 |
| K66 | 1.50 | 69 | 0.018898 |
| M54 | 1.49 | 105 | 0.011600 |
| K21 | 1.49 | 84 | 0.014477 |
| J32 | 1.49 | 59 | 0.019029 |
| K63 | 1.48 | 62 | 0.019717 |
| N28 | 1.48 | 101 | 0.011462 |
| N83 | 1.48 | 52 | 0.023351 |
| K59 | 1.46 | 109 | 0.010392 |
| D37 | 1.46 | 60 | 0.019988 |
| C73 | 1.46 | 24 | 0.051448 |
| N62 | 1.46 | 91 | 0.013172 |
| J94 | 1.46 | 81 | 0.015552 |
| N93 | 1.44 | 66 | 0.018185 |
| B34 | 1.44 | 67 | 0.017100 |
| N39 | 1.42 | 88 | 0.012284 |
| D68 | 1.42 | 90 | 0.013917 |
| C79 | 1.41 | 59 | 0.020562 |
| D25 | 1.41 | 62 | 0.019018 |
| J40 | 1.40 | 90 | 0.013081 |
| M13 | 1.40 | 87 | 0.013936 |
| E03 | 1.40 | 78 | 0.014015 |
| K92 | 1.39 | 78 | 0.013963 |
| N19 | 1.37 | 57 | 0.019362 |
| A16 | 1.36 | 37 | 0.032979 |
| N95 | 1.36 | 105 | 0.010236 |
| I15 | 1.36 | 38 | 0.030734 |
| B18 | 1.35 | 45 | 0.023928 |
| I67 | 1.35 | 78 | 0.013714 |
| N03 | 1.35 | 42 | 0.024748 |
| M35 | 1.34 | 76 | 0.014651 |
| G93 | 1.34 | 74 | 0.014797 |
| N63 | 1.33 | 61 | 0.018042 |
| E11 | 1.33 | 72 | 0.015588 |
| K65 | 1.33 | 66 | 0.016763 |
| L23 | 1.32 | 93 | 0.011769 |
| I38 | 1.30 | 28 | 0.038155 |
| M79 | 1.30 | 106 | 0.008734 |
| I69 | 1.30 | 47 | 0.022531 |
| I49 | 1.29 | 80 | 0.012262 |
| M32 | 1.29 | 44 | 0.025095 |
| A49 | 1.28 | 107 | 0.009118 |
| N88 | 1.27 | 61 | 0.016949 |
| D69 | 1.27 | 87 | 0.010848 |
| K12 | 1.27 | 90 | 0.011644 |
| E21 | 1.25 | 50 | 0.020957 |
| I84 | 1.25 | 76 | 0.012167 |
| K74 | 1.25 | 29 | 0.037520 |
| J37 | 1.25 | 51 | 0.019799 |
| K62 | 1.24 | 58 | 0.016556 |
| N41 | 1.24 | 79 | 0.012628 |
| J20 | 1.24 | 93 | 0.010678 |
| D48 | 1.23 | 51 | 0.018910 |
| J44 | 1.23 | 54 | 0.017557 |
| N72 | 1.23 | 55 | 0.017912 |
| N20 | 1.22 | 51 | 0.018729 |
| H35 | 1.22 | 42 | 0.023703 |
| K05 | 1.22 | 77 | 0.011962 |
| M51 | 1.21 | 63 | 0.014719 |
| C50 | 1.21 | 28 | 0.037431 |
| G81 | 1.21 | 47 | 0.021753 |
| E14 | 1.21 | 59 | 0.017098 |
| N80 | 1.21 | 37 | 0.026519 |
| H52 | 1.21 | 89 | 0.010030 |
| J30 | 1.20 | 61 | 0.014464 |
| I80 | 1.19 | 72 | 0.012652 |
| I27 | 1.19 | 63 | 0.015990 |

Supplementary Table S21. Multimorbidity coefficient (MMC), number of connections, and average partial correlation for the 100 highest-MMC conditions in individuals aged 50-59 years.

| Condition | MMC | Number of connections | Average partial correlation per connection |
| --- | --- | --- | --- |
| K29 | 2.44 | 147 | 0.015471 |
| D64 | 2.32 | 152 | 0.01393 |
| E88 | 2.27 | 139 | 0.015234 |
| I50 | 2.24 | 81 | 0.025605 |
| D61 | 2.22 | 87 | 0.023813 |
| E87 | 2.21 | 170 | 0.011900 |
| K76 | 2.21 | 147 | 0.013445 |
| J15 | 2.16 | 143 | 0.013595 |
| L30 | 2.01 | 150 | 0.012052 |
| G47 | 2.01 | 167 | 0.010407 |
| M81 | 1.99 | 138 | 0.012782 |
| I70 | 1.97 | 130 | 0.013415 |
| E78 | 1.97 | 154 | 0.011423 |
| I63 | 1.86 | 80 | 0.020415 |
| N18 | 1.75 | 57 | 0.027658 |
| D70 | 1.74 | 134 | 0.011182 |
| J31 | 1.73 | 125 | 0.012075 |
| F41 | 1.71 | 96 | 0.014838 |
| N95 | 1.71 | 142 | 0.010996 |
| E04 | 1.70 | 113 | 0.012960 |
| D37 | 1.69 | 72 | 0.020689 |
| M25 | 1.69 | 122 | 0.012319 |
| I25 | 1.69 | 76 | 0.019222 |
| H16 | 1.67 | 128 | 0.011463 |
| C78 | 1.65 | 51 | 0.029540 |
| C79 | 1.65 | 67 | 0.022010 |
| M50 | 1.64 | 101 | 0.014583 |
| K63 | 1.64 | 78 | 0.018140 |
| I10 | 1.63 | 97 | 0.014756 |
| J96 | 1.60 | 82 | 0.018296 |
| N25 | 1.60 | 48 | 0.030364 |
| N39 | 1.59 | 129 | 0.010663 |
| N28 | 1.59 | 120 | 0.011115 |
| J44 | 1.55 | 70 | 0.019566 |
| I11 | 1.55 | 68 | 0.019795 |
| K92 | 1.55 | 105 | 0.012490 |
| J32 | 1.54 | 78 | 0.015906 |
| K21 | 1.53 | 96 | 0.013404 |
| I67 | 1.52 | 93 | 0.014198 |
| D68 | 1.50 | 107 | 0.012222 |
| C77 | 1.50 | 67 | 0.020203 |
| J94 | 1.50 | 81 | 0.015986 |
| E79 | 1.49 | 101 | 0.012335 |
| K66 | 1.48 | 81 | 0.016117 |
| K59 | 1.48 | 131 | 0.009129 |
| K74 | 1.48 | 35 | 0.038601 |
| E03 | 1.47 | 104 | 0.011539 |
| K52 | 1.47 | 120 | 0.010159 |
| J40 | 1.47 | 94 | 0.013948 |
| B34 | 1.46 | 79 | 0.015379 |
| N19 | 1.45 | 83 | 0.014921 |
| M54 | 1.44 | 103 | 0.011892 |
| I69 | 1.43 | 72 | 0.016992 |
| M13 | 1.43 | 90 | 0.014402 |
| N40 | 1.43 | 101 | 0.011674 |
| M51 | 1.42 | 87 | 0.013608 |
| N73 | 1.41 | 61 | 0.020519 |
| I49 | 1.41 | 100 | 0.011273 |
| D48 | 1.41 | 65 | 0.017815 |
| A16 | 1.41 | 35 | 0.035838 |
| M35 | 1.39 | 103 | 0.011687 |
| D25 | 1.39 | 75 | 0.016271 |
| E11 | 1.39 | 82 | 0.014801 |
| E83 | 1.36 | 90 | 0.012604 |
| N83 | 1.36 | 54 | 0.021406 |
| I15 | 1.36 | 46 | 0.024937 |
| J06 | 1.35 | 91 | 0.012997 |
| I80 | 1.35 | 92 | 0.011933 |
| G62 | 1.35 | 120 | 0.008639 |
| D69 | 1.34 | 103 | 0.010657 |
| M79 | 1.34 | 121 | 0.008861 |
| K12 | 1.34 | 97 | 0.012268 |
| E14 | 1.33 | 84 | 0.013892 |
| H35 | 1.33 | 51 | 0.021896 |
| K62 | 1.33 | 72 | 0.015020 |
| A49 | 1.32 | 128 | 0.008552 |
| L23 | 1.32 | 97 | 0.011666 |
| I27 | 1.32 | 63 | 0.017986 |
| G93 | 1.3 | 77 | 0.014318 |
| K31 | 1.3 | 71 | 0.014686 |
| J37 | 1.29 | 66 | 0.016245 |
| N03 | 1.29 | 65 | 0.015998 |
| N62 | 1.29 | 110 | 0.009839 |
| N93 | 1.28 | 55 | 0.019194 |
| I38 | 1.27 | 32 | 0.032936 |
| K05 | 1.26 | 88 | 0.011816 |
| N20 | 1.26 | 66 | 0.015869 |
| C73 | 1.26 | 24 | 0.044063 |
| N63 | 1.25 | 70 | 0.014572 |
| H26 | 1.25 | 69 | 0.013523 |
| K65 | 1.25 | 68 | 0.016200 |
| E21 | 1.24 | 50 | 0.021244 |
| B18 | 1.24 | 52 | 0.018813 |
| I48 | 1.23 | 52 | 0.019294 |
| I84 | 1.22 | 99 | 0.009844 |
| H43 | 1.22 | 92 | 0.010610 |
| J20 | 1.21 | 98 | 0.010312 |
| C95 | 1.21 | 32 | 0.032129 |
| N72 | 1.20 | 53 | 0.019104 |
| J42 | 1.20 | 89 | 0.010643 |

Supplementary Table S22. Multimorbidity coefficient (MMC), number of connections, and average partial correlation for the 100 highest-MMC conditions in individuals aged 60-69 years.

| Condition | MMC | Number of connections | Average partial correlation per connection |
| --- | --- | --- | --- |
| K29 | 2.56 | 148 | 0.015776 |
| E88 | 2.46 | 137 | 0.016318 |
| D64 | 2.41 | 148 | 0.014528 |
| I50 | 2.38 | 89 | 0.024688 |
| E87 | 2.32 | 165 | 0.012718 |
| I70 | 2.27 | 137 | 0.014727 |
| K76 | 2.26 | 138 | 0.014465 |
| J15 | 2.25 | 151 | 0.013391 |
| I63 | 2.16 | 99 | 0.019383 |
| G47 | 2.11 | 164 | 0.010730 |
| M81 | 2.09 | 142 | 0.013037 |
| D61 | 2.08 | 84 | 0.022653 |
| L30 | 1.98 | 128 | 0.013284 |
| D37 | 1.92 | 65 | 0.026059 |
| F41 | 1.88 | 111 | 0.014367 |
| E78 | 1.86 | 130 | 0.012376 |
| K63 | 1.84 | 88 | 0.018211 |
| I25 | 1.84 | 85 | 0.018697 |
| N18 | 1.84 | 56 | 0.029173 |
| N40 | 1.83 | 127 | 0.012498 |
| K92 | 1.82 | 109 | 0.014154 |
| C78 | 1.82 | 60 | 0.027858 |
| N39 | 1.81 | 127 | 0.012212 |
| J44 | 1.81 | 70 | 0.022877 |
| J96 | 1.79 | 76 | 0.021433 |
| N28 | 1.79 | 128 | 0.011684 |
| C79 | 1.79 | 74 | 0.021401 |
| N25 | 1.76 | 67 | 0.023656 |
| E04 | 1.74 | 102 | 0.014292 |
| D70 | 1.73 | 127 | 0.011265 |
| I67 | 1.72 | 112 | 0.013601 |
| H16 | 1.72 | 122 | 0.012106 |
| J31 | 1.70 | 118 | 0.012317 |
| M25 | 1.66 | 105 | 0.013359 |
| J94 | 1.66 | 96 | 0.015420 |
| H25 | 1.65 | 72 | 0.018212 |
| K21 | 1.65 | 110 | 0.012311 |
| I69 | 1.65 | 98 | 0.014633 |
| I10 | 1.64 | 91 | 0.015354 |
| M51 | 1.64 | 91 | 0.015144 |
| M50 | 1.63 | 93 | 0.015291 |
| I49 | 1.61 | 101 | 0.013265 |
| D48 | 1.61 | 80 | 0.015935 |
| I11 | 1.59 | 75 | 0.018087 |
| J32 | 1.59 | 88 | 0.014510 |
| K74 | 1.58 | 38 | 0.036470 |
| E79 | 1.57 | 95 | 0.013571 |
| K59 | 1.57 | 121 | 0.010236 |
| K66 | 1.56 | 82 | 0.016286 |
| G62 | 1.54 | 115 | 0.010317 |
| M13 | 1.54 | 83 | 0.016573 |
| J40 | 1.53 | 90 | 0.015232 |
| J42 | 1.53 | 101 | 0.012039 |
| M54 | 1.52 | 92 | 0.014008 |
| N19 | 1.52 | 73 | 0.017278 |
| K52 | 1.51 | 107 | 0.011045 |
| E03 | 1.51 | 107 | 0.011093 |
| C77 | 1.51 | 67 | 0.019679 |
| K31 | 1.51 | 85 | 0.01465 |
| N95 | 1.51 | 102 | 0.012647 |
| K65 | 1.51 | 82 | 0.016152 |
| B34 | 1.50 | 74 | 0.016210 |
| D69 | 1.48 | 100 | 0.011381 |
| A16 | 1.47 | 37 | 0.034229 |
| G31 | 1.47 | 104 | 0.011748 |
| I80 | 1.45 | 97 | 0.012087 |
| J06 | 1.45 | 96 | 0.012862 |
| D68 | 1.44 | 98 | 0.012587 |
| M79 | 1.44 | 114 | 0.009960 |
| E14 | 1.44 | 84 | 0.014590 |
| C85 | 1.43 | 52 | 0.023017 |
| N03 | 1.43 | 70 | 0.016226 |
| K62 | 1.41 | 64 | 0.017568 |
| K12 | 1.41 | 93 | 0.013081 |
| E11 | 1.41 | 72 | 0.016612 |
| I65 | 1.40 | 71 | 0.016593 |
| D41 | 1.40 | 36 | 0.031783 |
| M35 | 1.39 | 87 | 0.013013 |
| M48 | 1.39 | 81 | 0.013633 |
| N20 | 1.37 | 70 | 0.015605 |
| G93 | 1.36 | 67 | 0.016702 |
| E83 | 1.36 | 101 | 0.011126 |
| A49 | 1.36 | 118 | 0.009164 |
| B18 | 1.35 | 67 | 0.015530 |
| C34 | 1.34 | 38 | 0.030771 |
| I48 | 1.34 | 57 | 0.020087 |
| N73 | 1.33 | 56 | 0.019510 |
| I34 | 1.33 | 52 | 0.021102 |
| H35 | 1.33 | 57 | 0.018979 |
| K05 | 1.32 | 87 | 0.012356 |
| L08 | 1.32 | 95 | 0.011311 |
| I27 | 1.32 | 51 | 0.022458 |
| L23 | 1.32 | 84 | 0.013333 |
| J20 | 1.31 | 94 | 0.011457 |
| M17 | 1.31 | 66 | 0.016018 |
| J43 | 1.31 | 81 | 0.012821 |
| N83 | 1.31 | 51 | 0.020484 |
| K80 | 1.30 | 69 | 0.014466 |
| I84 | 1.30 | 97 | 0.010093 |
| J37 | 1.29 | 73 | 0.013811 |

Supplementary Table S23. Multimorbidity coefficient (MMC), number of connections, and average partial correlation for the 100 highest-MMC conditions in individuals aged 70-79 years.

| Condition | MMC | Number of connections | Average partial correlation per connection |
| --- | --- | --- | --- |
| E88 | 2.60 | 131 | 0.017120 |
| I70 | 2.59 | 129 | 0.017281 |
| K29 | 2.52 | 133 | 0.016639 |
| I50 | 2.45 | 79 | 0.027466 |
| D64 | 2.42 | 118 | 0.017732 |
| E87 | 2.37 | 135 | 0.014975 |
| J15 | 2.31 | 119 | 0.016594 |
| K76 | 2.24 | 129 | 0.014711 |
| M81 | 2.23 | 130 | 0.014570 |
| I63 | 2.20 | 84 | 0.022202 |
| L30 | 2.20 | 123 | 0.014820 |
| H25 | 2.15 | 81 | 0.021440 |
| G47 | 2.14 | 133 | 0.012869 |
| N40 | 2.12 | 113 | 0.015558 |
| K92 | 2.09 | 96 | 0.017806 |
| N28 | 2.07 | 125 | 0.013449 |
| D61 | 2.04 | 81 | 0.021635 |
| I69 | 2.03 | 116 | 0.014831 |
| N39 | 2.02 | 116 | 0.014877 |
| F41 | 2.00 | 102 | 0.015797 |
| J44 | 1.99 | 67 | 0.025093 |
| N18 | 1.97 | 64 | 0.026392 |
| D37 | 1.97 | 65 | 0.025831 |
| N25 | 1.97 | 66 | 0.025369 |
| K63 | 1.96 | 77 | 0.020679 |
| I67 | 1.94 | 100 | 0.016401 |
| J96 | 1.91 | 71 | 0.023725 |
| E04 | 1.91 | 107 | 0.014528 |
| C78 | 1.89 | 54 | 0.030737 |
| C79 | 1.87 | 58 | 0.026630 |
| J94 | 1.87 | 85 | 0.019009 |
| K21 | 1.87 | 117 | 0.012476 |
| E78 | 1.86 | 108 | 0.013639 |
| H16 | 1.84 | 101 | 0.014347 |
| I49 | 1.83 | 82 | 0.017122 |
| M51 | 1.80 | 78 | 0.018477 |
| I11 | 1.80 | 90 | 0.016473 |
| I25 | 1.80 | 60 | 0.024835 |
| K59 | 1.79 | 106 | 0.012839 |
| G62 | 1.78 | 119 | 0.011825 |
| E79 | 1.78 | 91 | 0.015226 |
| J31 | 1.77 | 111 | 0.012346 |
| I65 | 1.74 | 71 | 0.020030 |
| D70 | 1.74 | 109 | 0.012148 |
| E46 | 1.73 | 111 | 0.012183 |
| D48 | 1.73 | 96 | 0.014406 |
| N19 | 1.72 | 78 | 0.017445 |
| G31 | 1.71 | 80 | 0.016619 |
| M25 | 1.71 | 88 | 0.015745 |
| N03 | 1.70 | 69 | 0.018613 |
| K52 | 1.70 | 108 | 0.011825 |
| J42 | 1.68 | 84 | 0.014732 |
| E14 | 1.67 | 75 | 0.017117 |
| D41 | 1.66 | 39 | 0.032695 |
| M50 | 1.66 | 82 | 0.016804 |
| K74 | 1.65 | 41 | 0.033282 |
| M13 | 1.65 | 76 | 0.018839 |
| D69 | 1.65 | 96 | 0.012686 |
| J32 | 1.64 | 81 | 0.015097 |
| I20 | 1.64 | 79 | 0.015852 |
| I80 | 1.63 | 98 | 0.012810 |
| M79 | 1.63 | 105 | 0.011490 |
| K31 | 1.62 | 79 | 0.015803 |
| K66 | 1.61 | 76 | 0.017573 |
| I48 | 1.60 | 63 | 0.021122 |
| M54 | 1.60 | 82 | 0.015793 |
| I10 | 1.59 | 70 | 0.017923 |
| K65 | 1.59 | 71 | 0.019046 |
| A16 | 1.59 | 37 | 0.034026 |
| J40 | 1.58 | 77 | 0.017380 |
| K62 | 1.57 | 64 | 0.017518 |
| K04 | 1.57 | 85 | 0.013258 |
| I34 | 1.55 | 47 | 0.02582 |
| M17 | 1.55 | 67 | 0.017772 |
| D68 | 1.55 | 96 | 0.012668 |
| M48 | 1.54 | 76 | 0.015753 |
| E11 | 1.54 | 65 | 0.018552 |
| E03 | 1.52 | 82 | 0.012965 |
| J18 | 1.51 | 76 | 0.015515 |
| C77 | 1.50 | 60 | 0.020045 |
| I77 | 1.50 | 87 | 0.013623 |
| H02 | 1.49 | 60 | 0.016452 |
| G93 | 1.49 | 64 | 0.017708 |
| M35 | 1.49 | 91 | 0.012691 |
| B18 | 1.49 | 62 | 0.017678 |
| C34 | 1.49 | 40 | 0.031659 |
| E72 | 1.48 | 77 | 0.015671 |
| J43 | 1.47 | 73 | 0.015185 |
| J06 | 1.47 | 89 | 0.012754 |
| K05 | 1.46 | 76 | 0.014044 |
| K12 | 1.46 | 77 | 0.014965 |
| N20 | 1.46 | 74 | 0.015076 |
| L08 | 1.46 | 76 | 0.014547 |
| K83 | 1.46 | 58 | 0.020084 |
| N95 | 1.45 | 78 | 0.014583 |
| K80 | 1.45 | 60 | 0.017250 |
| H35 | 1.45 | 54 | 0.019522 |
| B49 | 1.44 | 94 | 0.011288 |
| I84 | 1.44 | 94 | 0.011027 |
| K56 | 1.44 | 86 | 0.013647 |

Supplementary Table S24. Multimorbidity coefficient (MMC), number of connections, and average partial correlation for the 100 highest-MMC conditions in individuals aged ≥ 80 years.

| Condition | MMC | Number of connections | Average partial correlation per connection |
| --- | --- | --- | --- |
| I70 | 2.85 | 102 | 0.021617 |
| E88 | 2.80 | 99 | 0.021649 |
| M81 | 2.56 | 100 | 0.018817 |
| D64 | 2.53 | 83 | 0.022356 |
| I69 | 2.44 | 94 | 0.020126 |
| I50 | 2.42 | 55 | 0.035175 |
| N28 | 2.41 | 98 | 0.017854 |
| E87 | 2.41 | 90 | 0.020244 |
| H25 | 2.40 | 66 | 0.025086 |
| K29 | 2.38 | 79 | 0.022850 |
| L30 | 2.37 | 87 | 0.019765 |
| K76 | 2.37 | 86 | 0.019762 |
| N40 | 2.37 | 95 | 0.019856 |
| J15 | 2.36 | 83 | 0.020510 |
| N39 | 2.36 | 98 | 0.018384 |
| N18 | 2.34 | 58 | 0.030004 |
| K92 | 2.33 | 82 | 0.020545 |
| N25 | 2.33 | 83 | 0.022476 |
| E56 | 2.32 | 88 | 0.020215 |
| G47 | 2.32 | 89 | 0.018619 |
| I63 | 2.31 | 61 | 0.028550 |
| F41 | 2.28 | 79 | 0.019595 |
| J96 | 2.24 | 62 | 0.028783 |
| G62 | 2.20 | 94 | 0.016271 |
| C78 | 2.19 | 53 | 0.032272 |
| K21 | 2.16 | 95 | 0.015571 |
| H16 | 2.16 | 84 | 0.018332 |
| K59 | 2.15 | 84 | 0.017343 |
| I11 | 2.15 | 79 | 0.019726 |
| G31 | 2.13 | 80 | 0.019632 |
| J94 | 2.12 | 59 | 0.027450 |
| I65 | 2.12 | 70 | 0.022081 |
| J44 | 2.12 | 61 | 0.027281 |
| D37 | 2.09 | 50 | 0.031277 |
| N03 | 2.09 | 79 | 0.019328 |
| I67 | 2.09 | 74 | 0.019736 |
| J37 | 2.09 | 83 | 0.018264 |
| C79 | 2.08 | 57 | 0.027256 |
| I49 | 2.06 | 67 | 0.021116 |
| M25 | 2.05 | 70 | 0.019840 |
| M51 | 2.05 | 67 | 0.021374 |
| I80 | 2.03 | 72 | 0.019576 |
| K52 | 2.02 | 72 | 0.017192 |
| N19 | 2.01 | 63 | 0.021419 |
| I20 | 2.00 | 72 | 0.019527 |
| E14 | 2.00 | 68 | 0.021217 |
| K63 | 1.99 | 65 | 0.021995 |
| E78 | 1.99 | 82 | 0.016797 |
| J30 | 1.99 | 77 | 0.016417 |
| J42 | 1.99 | 73 | 0.017383 |
| E04 | 1.98 | 81 | 0.016171 |
| J31 | 1.98 | 88 | 0.016363 |
| E46 | 1.95 | 82 | 0.016283 |
| D48 | 1.94 | 59 | 0.021382 |
| K04 | 1.93 | 64 | 0.018639 |
| I25 | 1.92 | 47 | 0.030665 |
| E79 | 1.92 | 59 | 0.021282 |
| D61 | 1.92 | 65 | 0.021447 |
| M79 | 1.91 | 76 | 0.014836 |
| J32 | 1.90 | 76 | 0.016842 |
| M17 | 1.89 | 51 | 0.022229 |
| I48 | 1.89 | 50 | 0.026520 |
| E11 | 1.88 | 60 | 0.020902 |
| M13 | 1.88 | 61 | 0.023299 |
| K65 | 1.88 | 60 | 0.023364 |
| L82 | 1.87 | 76 | 0.016325 |
| J18 | 1.87 | 61 | 0.021161 |
| M54 | 1.86 | 66 | 0.019966 |
| I34 | 1.85 | 27 | 0.046128 |
| H02 | 1.85 | 58 | 0.019980 |
| H35 | 1.84 | 58 | 0.020518 |
| J40 | 1.83 | 64 | 0.020314 |
| D68 | 1.83 | 82 | 0.015352 |
| K08 | 1.82 | 58 | 0.019660 |
| B49 | 1.82 | 76 | 0.016434 |
| K80 | 1.82 | 62 | 0.019836 |
| J84 | 1.82 | 73 | 0.015402 |
| K73 | 1.81 | 61 | 0.019027 |
| L29 | 1.81 | 72 | 0.015359 |
| D69 | 1.81 | 59 | 0.019193 |
| M48 | 1.80 | 58 | 0.019017 |
| K66 | 1.80 | 58 | 0.020979 |
| D41 | 1.80 | 25 | 0.047184 |
| E03 | 1.79 | 77 | 0.014649 |
| M50 | 1.79 | 67 | 0.018904 |
| D50 | 1.79 | 74 | 0.016081 |
| G93 | 1.79 | 58 | 0.020276 |
| B35 | 1.78 | 82 | 0.014080 |
| D70 | 1.78 | 58 | 0.018339 |
| I84 | 1.78 | 76 | 0.015232 |
| K74 | 1.77 | 48 | 0.025506 |
| L08 | 1.76 | 62 | 0.019002 |
| I10 | 1.75 | 48 | 0.023998 |
| K30 | 1.75 | 68 | 0.017039 |
| K56 | 1.74 | 62 | 0.019626 |
| K05 | 1.74 | 52 | 0.019784 |
| K12 | 1.73 | 65 | 0.017007 |
| C77 | 1.72 | 57 | 0.021971 |
| L23 | 1.72 | 71 | 0.017405 |
| F01 | 1.72 | 58 | 0.021586 |

Supplementary Figure S1. Number of times a health condition occurs in the 50 most common triad in female individuals.

The graph shows the number of times a health condition occurs in the 50 most common triads by selected age groups. The x-axis shows the frequency of occurrence. For clarity, diseases are represented by their ICD-10 codes; a complete list of codes and their corresponding full disease names is provided in Supplementary Table S25. An interactive version of this figure, with features for exploring full disease details, is available online at: https://pumc-multimorbidity.shinyapps.io/triad-frequency-visualization/

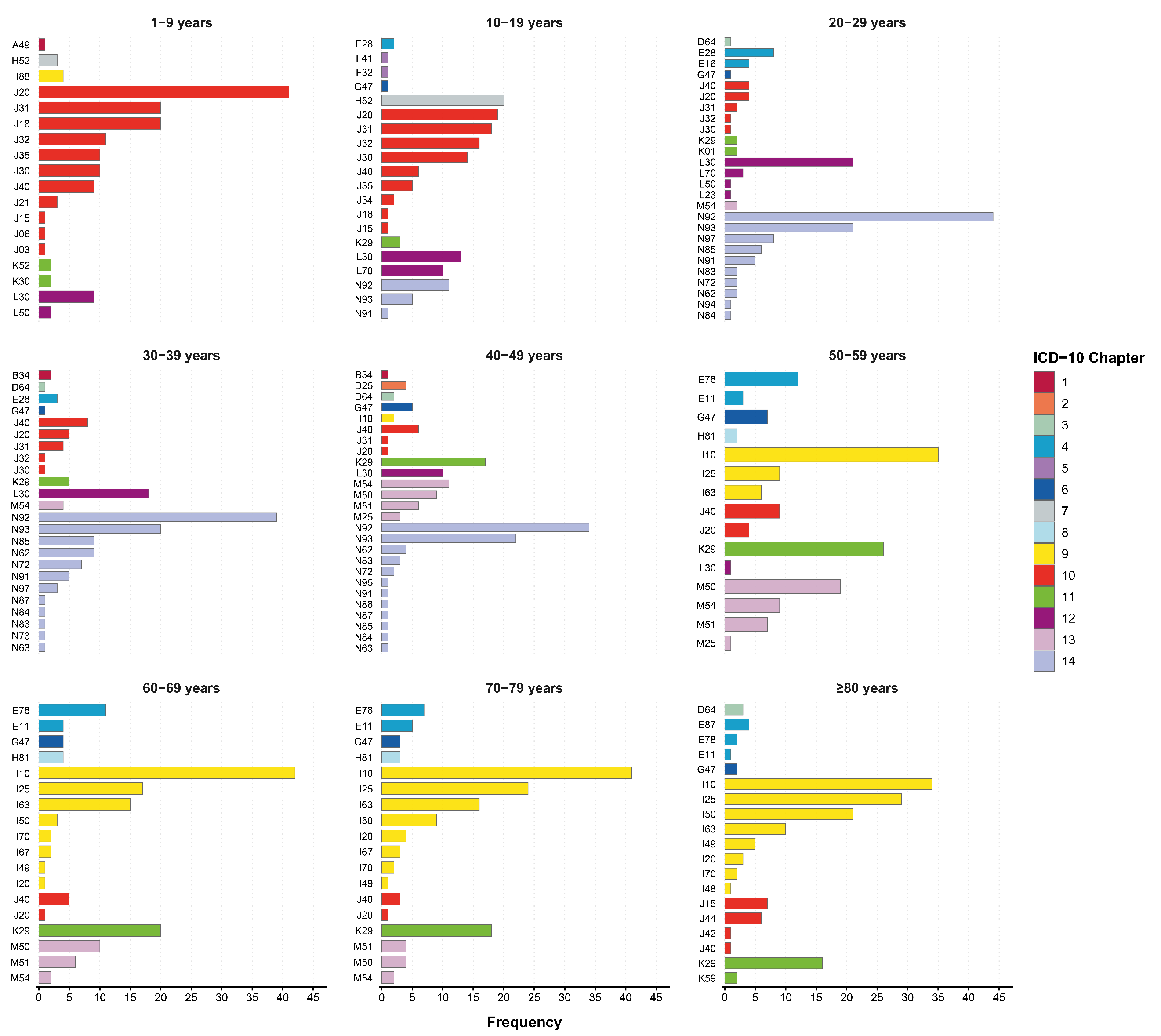

Supplementary Figure S2. Number of times a health condition occurs in the 50 most common triad in male individuals.

The graph shows the number of times a health condition occurs in the 50 most common triads by selected age groups. The x-axis shows the frequency of occurrence. For clarity, diseases are represented by their ICD-10 codes; a complete list of codes and their corresponding full disease names is provided in Supplementary Table S25. An interactive version of this figure, with features for exploring full disease details, is available online at: https://pumc-multimorbidity.shinyapps.io/triad-frequency-visualization/

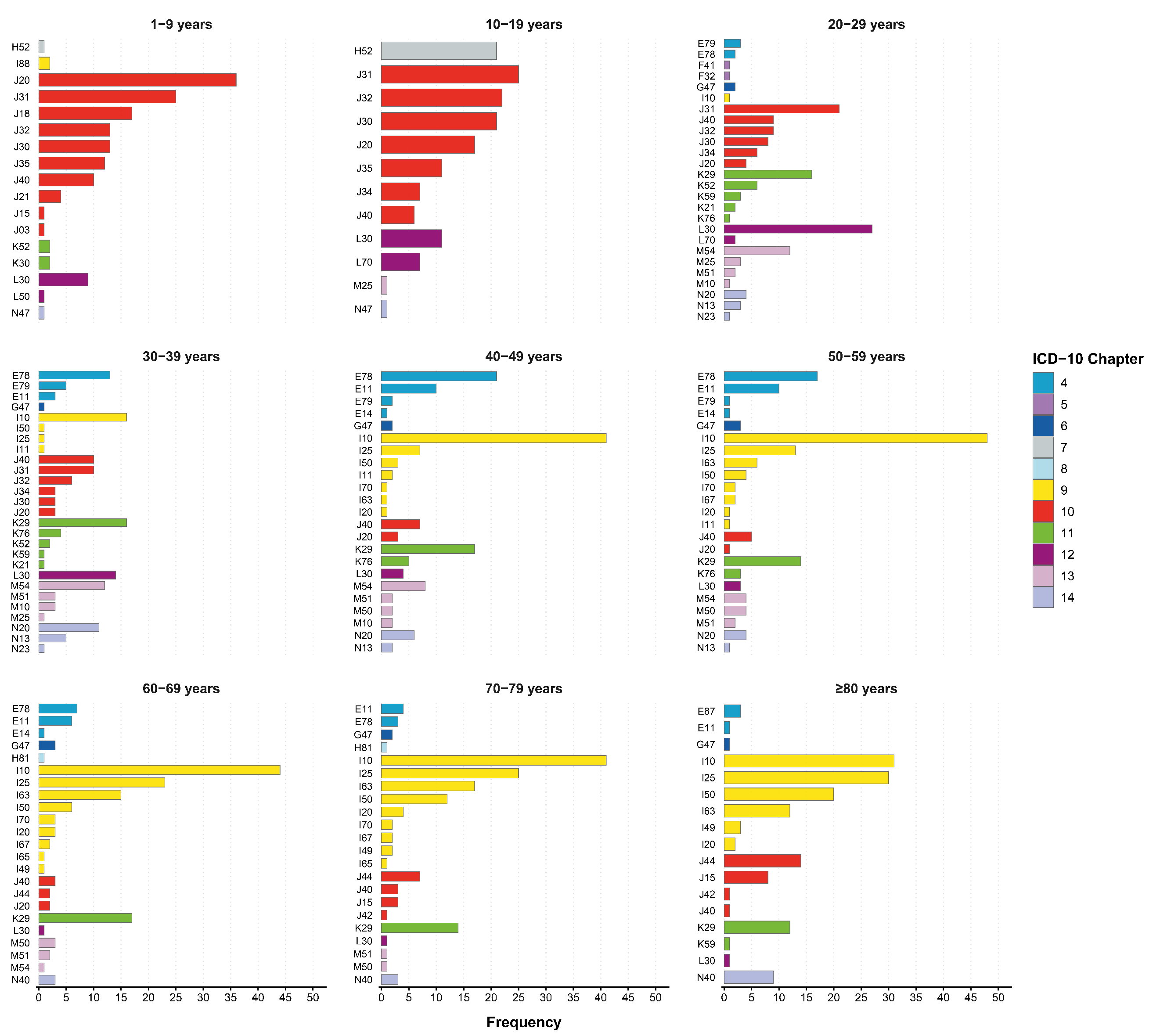

Supplementary Figure S3. multimorbidity network in overall population.

Nodes represent individual ICD‑10 conditions; node size is proportional to each condition’s period prevalence (2016–2023), and node color denotes the ICD‑10 chapter. Edges depict significant pairwise partial correlation coefficients (φ) between conditions after controlling for all other diseases; edge width is scaled to the magnitude of φ.

An interactive version of this network is available online, allowing for dynamic exploration by adjusting thresholds for both the partial correlation coefficient (φ) and the FDR-adjusted P-value. The tool can be accessed at: https://pumc-multimorbidity.shinyapps.io/multimorbidity-network/.

The static figures below are presented as illustrative examples of the network under different filtering criteria:

(S3A) Associations meeting FDR-adjusted *P* < 0.05 and φ ≥ 0.01;

(S3B) Associations meeting FDR-adjusted *P* < 0.01 and φ ≥ 0.01;

(S3C) Associations meeting FDR-adjusted *P* < 0.05 and φ ≥ 0.1.

S3A

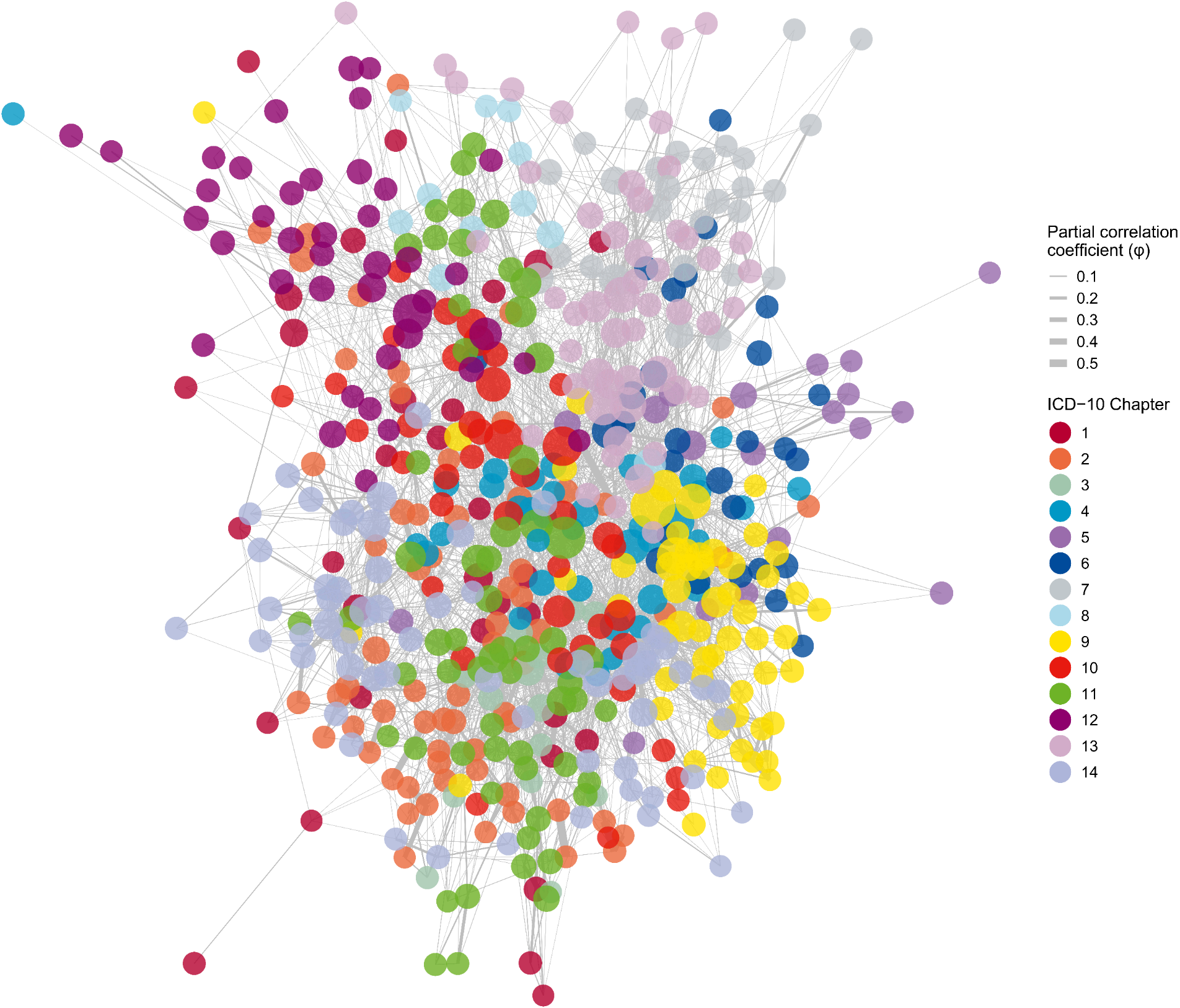

S3B

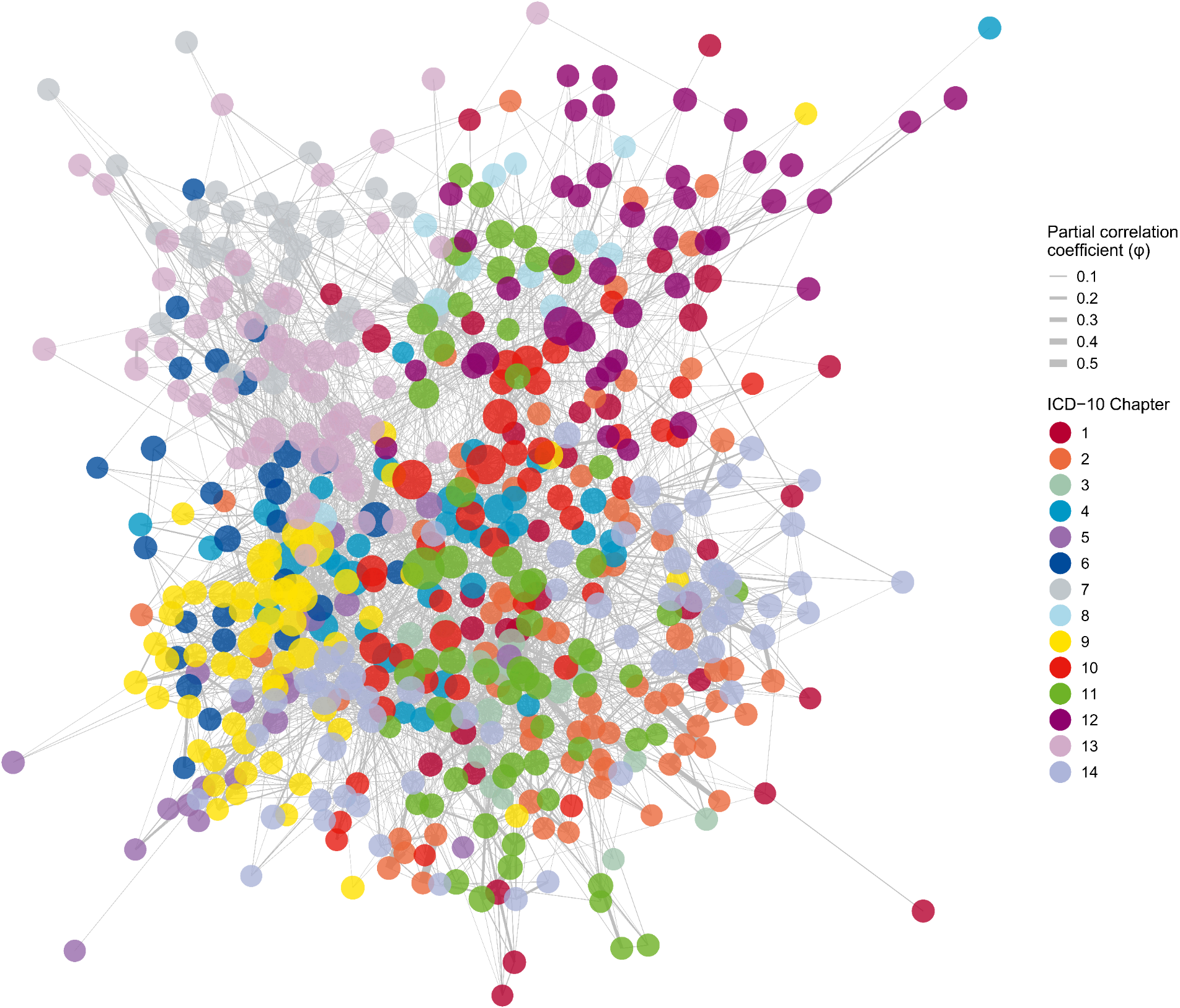

S3C

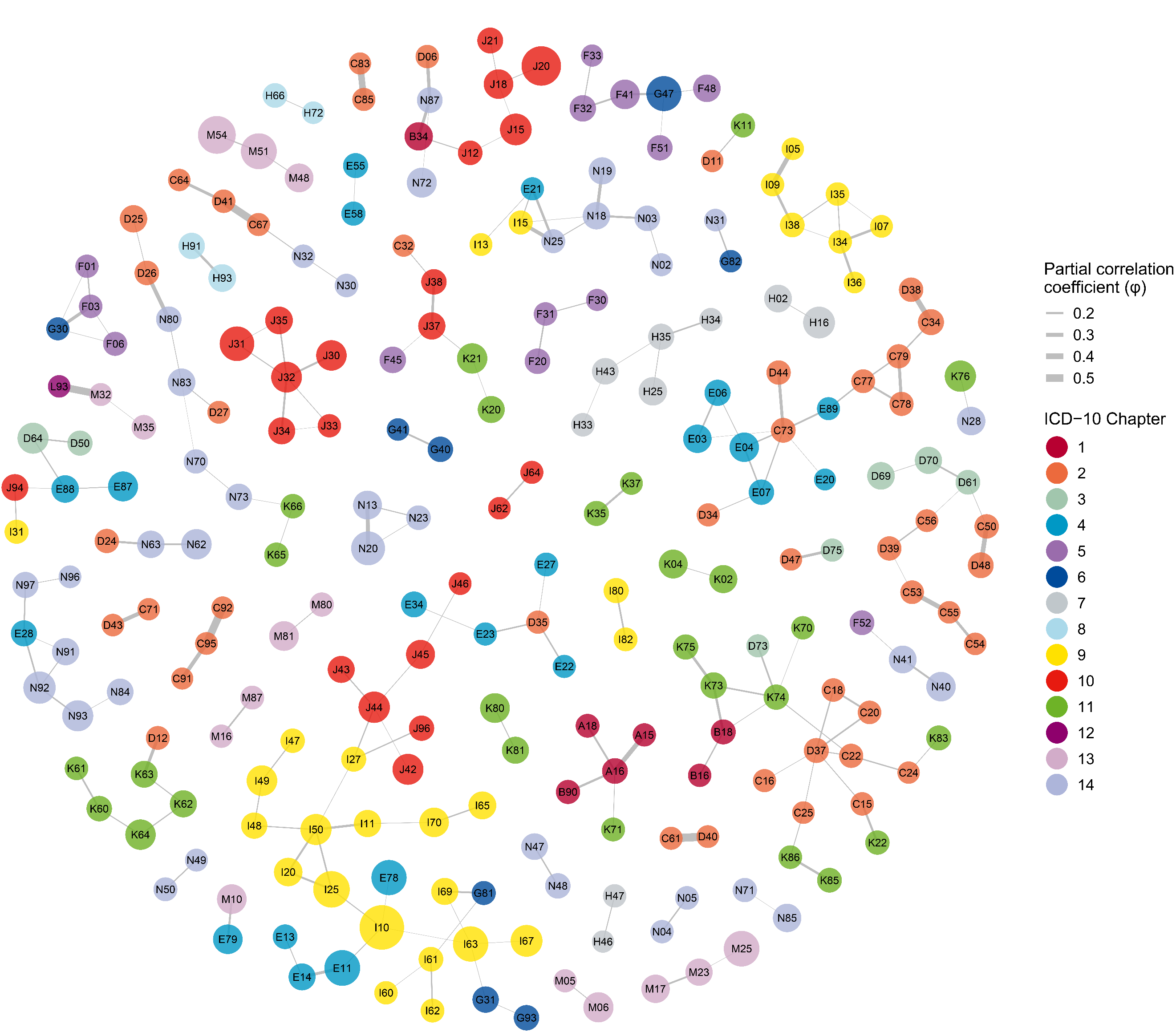

Supplementary Figure S4. Multimorbidity Network in female individuals.

Nodes represent individual ICD‑10 conditions; node size is proportional to each condition’s period prevalence (2016–2023), and node color denotes the ICD‑10 chapter. Edges depict significant pairwise partial correlation coefficients (φ) between conditions after controlling for all other diseases; edge width is scaled to the magnitude of φ.

An interactive version of this network is available online, allowing for dynamic exploration by adjusting thresholds for both the partial correlation coefficient (φ) and the FDR-adjusted *P*-value. The tool can be accessed at: https://pumc-multimorbidity.shinyapps.io/multimorbidity-network/.

The static figures below are presented as illustrative examples of the network under different filtering criteria:

(S4A) Associations meeting FDR-adjusted *P* < 0.05 and φ ≥ 0.01;

(S4B) Associations meeting FDR-adjusted *P* < 0.01 and φ ≥ 0.01;

(S4C) Associations meeting FDR-adjusted *P* < 0.05 and φ ≥ 0.05;

(S4D) Associations meeting FDR-adjusted *P* < 0.05 and φ ≥ 0.1.

S4A

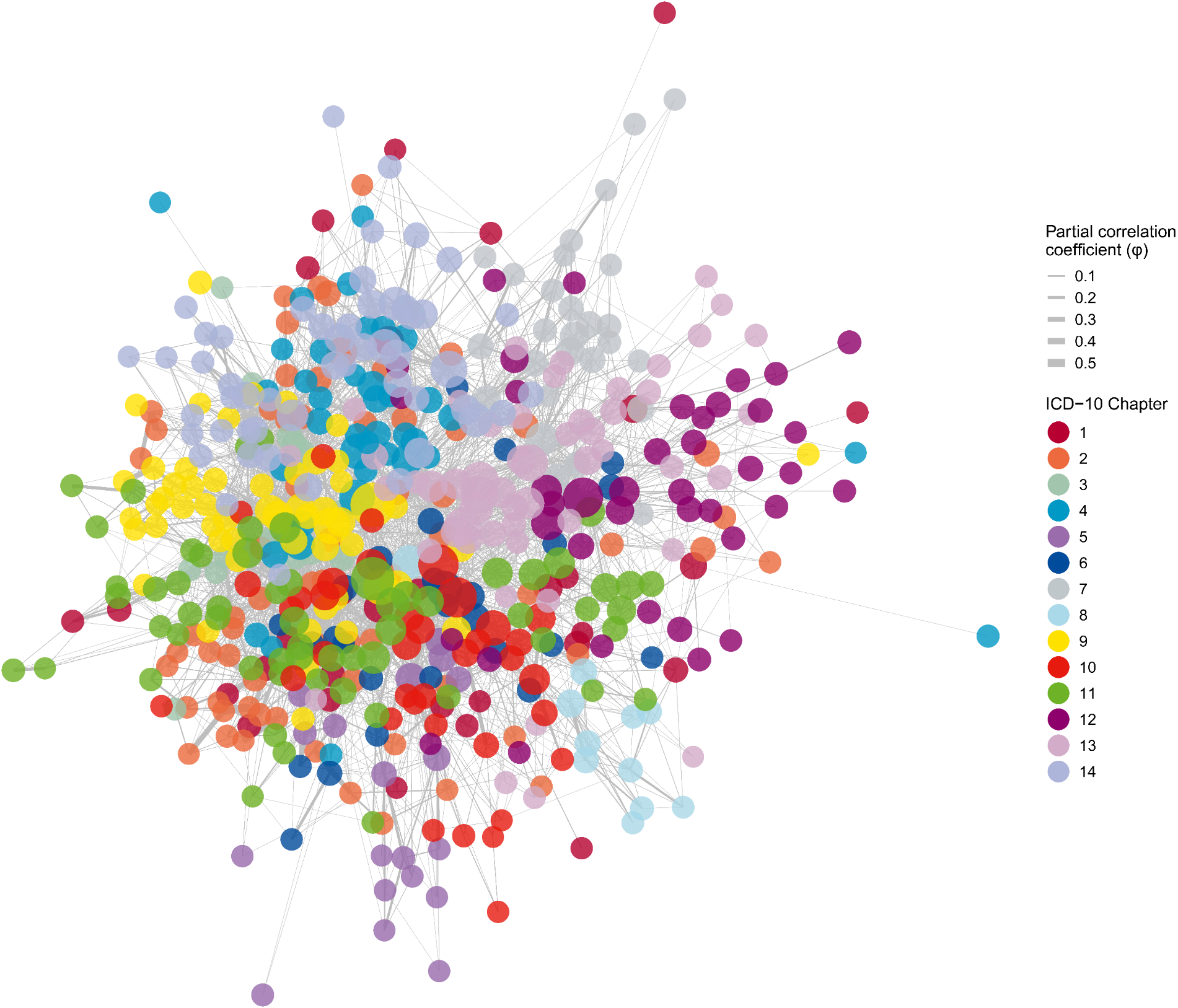

S4B

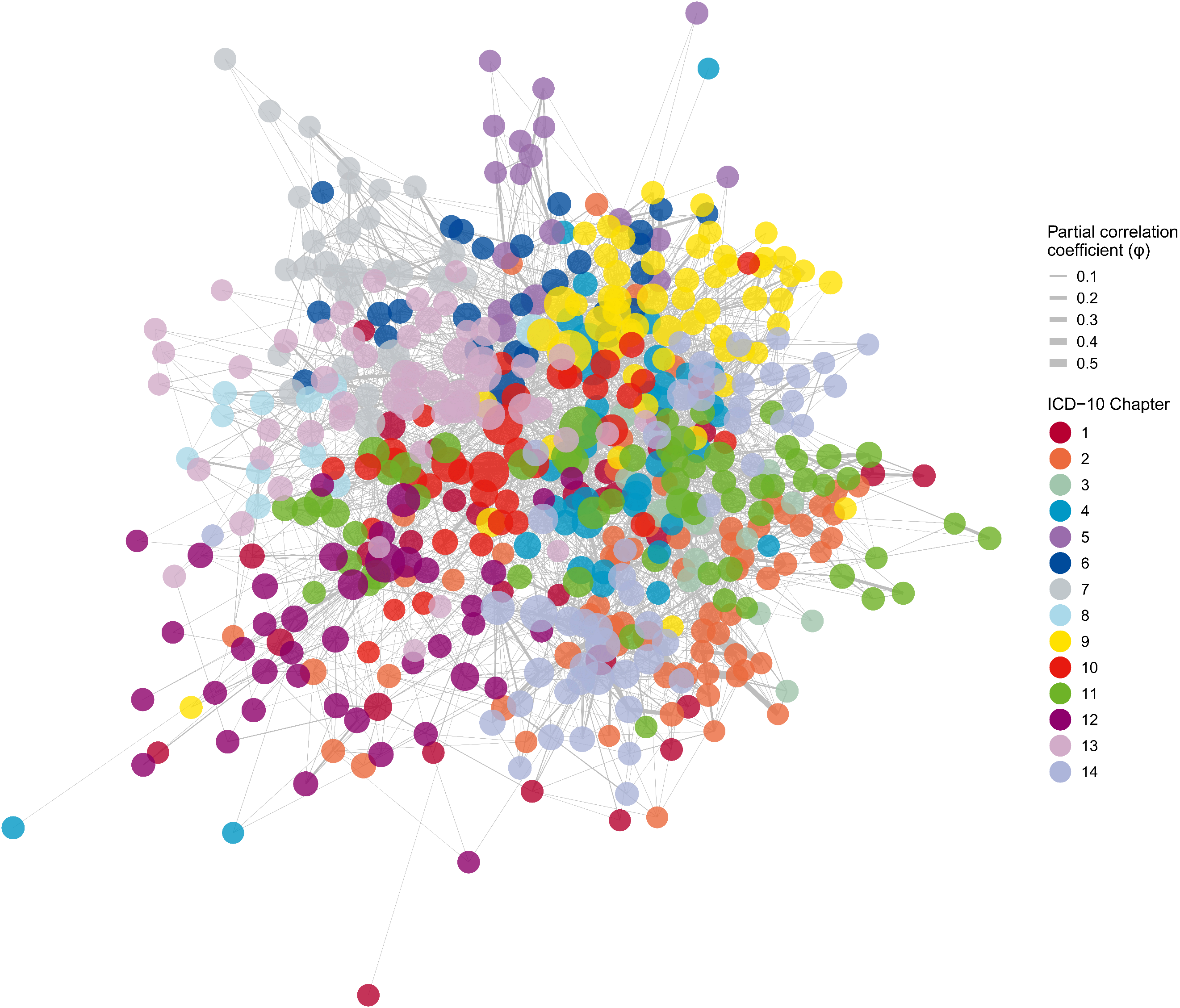

S4C

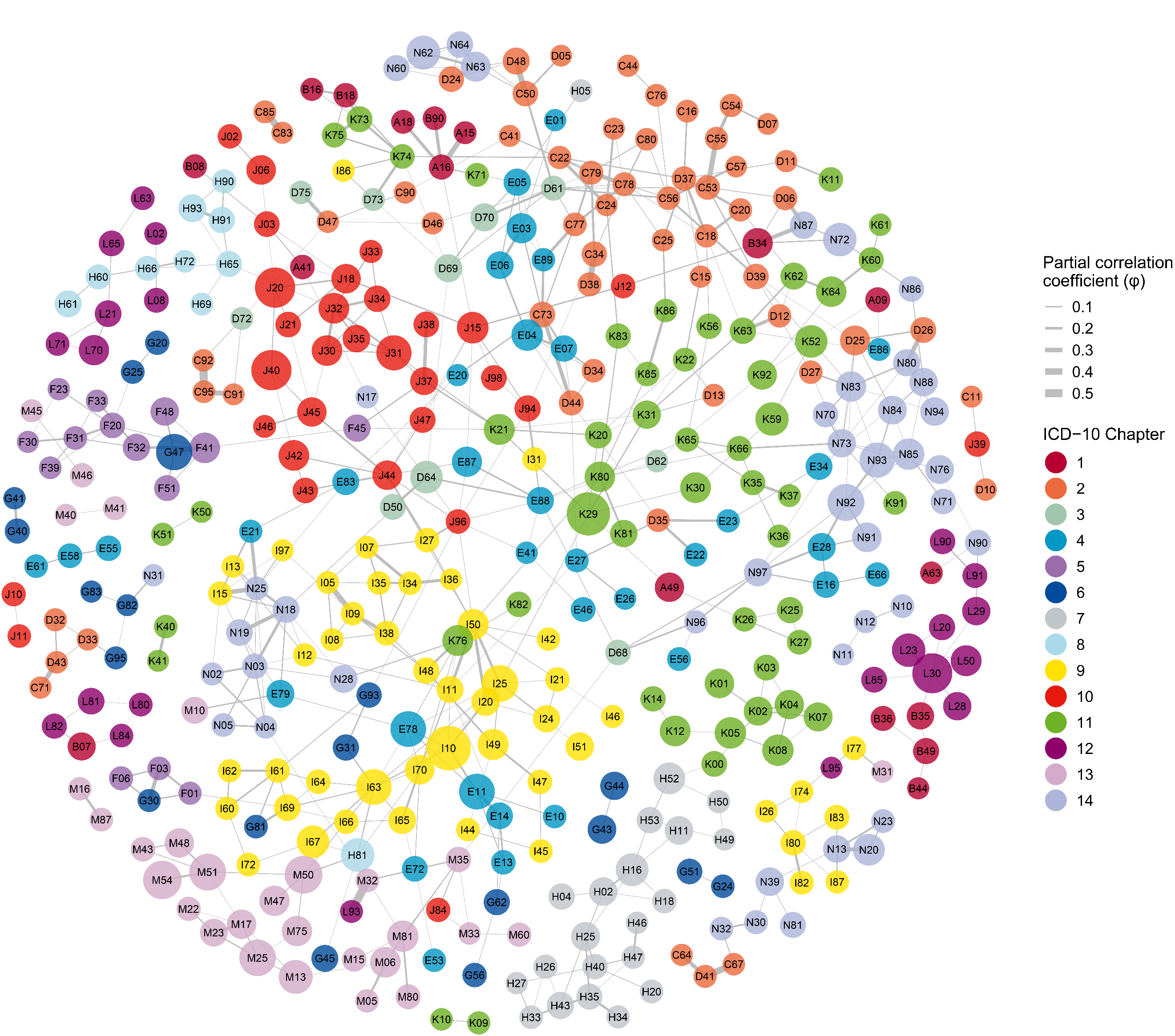

S4D

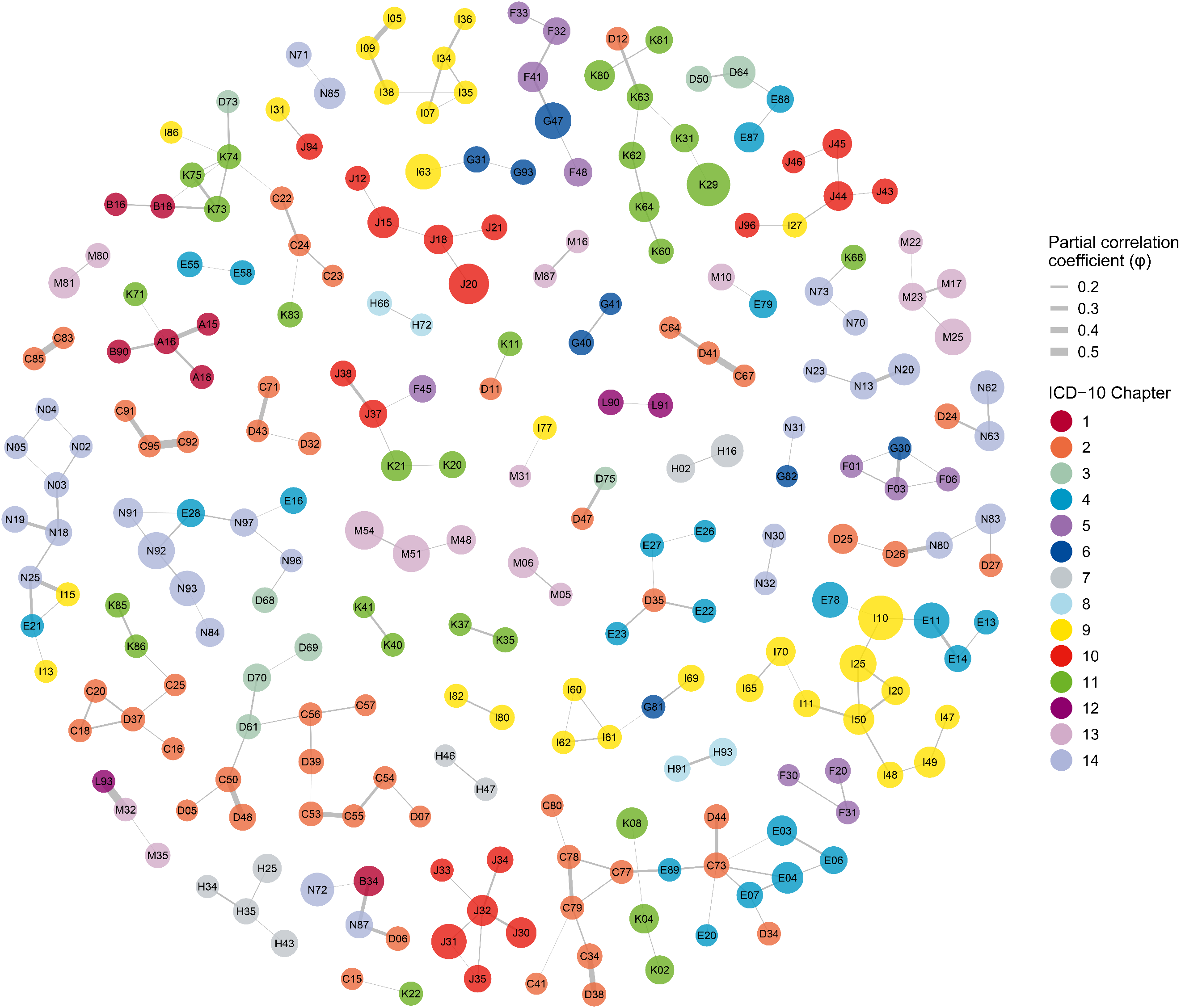

Supplementary Figure S5. Multimorbidity Network in male individuals.

Nodes represent individual ICD‑10 conditions; node size is proportional to each condition’s period prevalence (2016–2023), and node color denotes the ICD‑10 chapter. Edges depict significant pairwise partial correlation coefficients (φ) between conditions after controlling for all other diseases; edge width is scaled to the magnitude of φ.

An interactive version of this network is available online, allowing for dynamic exploration by adjusting thresholds for both the partial correlation coefficient (φ) and the FDR-adjusted *P*-value. The tool can be accessed at: https://pumc-multimorbidity.shinyapps.io/multimorbidity-network/.

The static figures below are presented as illustrative examples of the network under different filtering criteria:

(S5A) Associations meeting FDR-adjusted *P* < 0.05 and φ ≥ 0.01;

(S5B) Associations meeting FDR-adjusted *P* < 0.01 and φ ≥ 0.01;

(S5C) Associations meeting FDR-adjusted *P* < 0.05 and φ ≥ 0.05;

(S5D) Associations meeting FDR-adjusted *P* < 0.05 and φ ≥ 0.1.

S5A

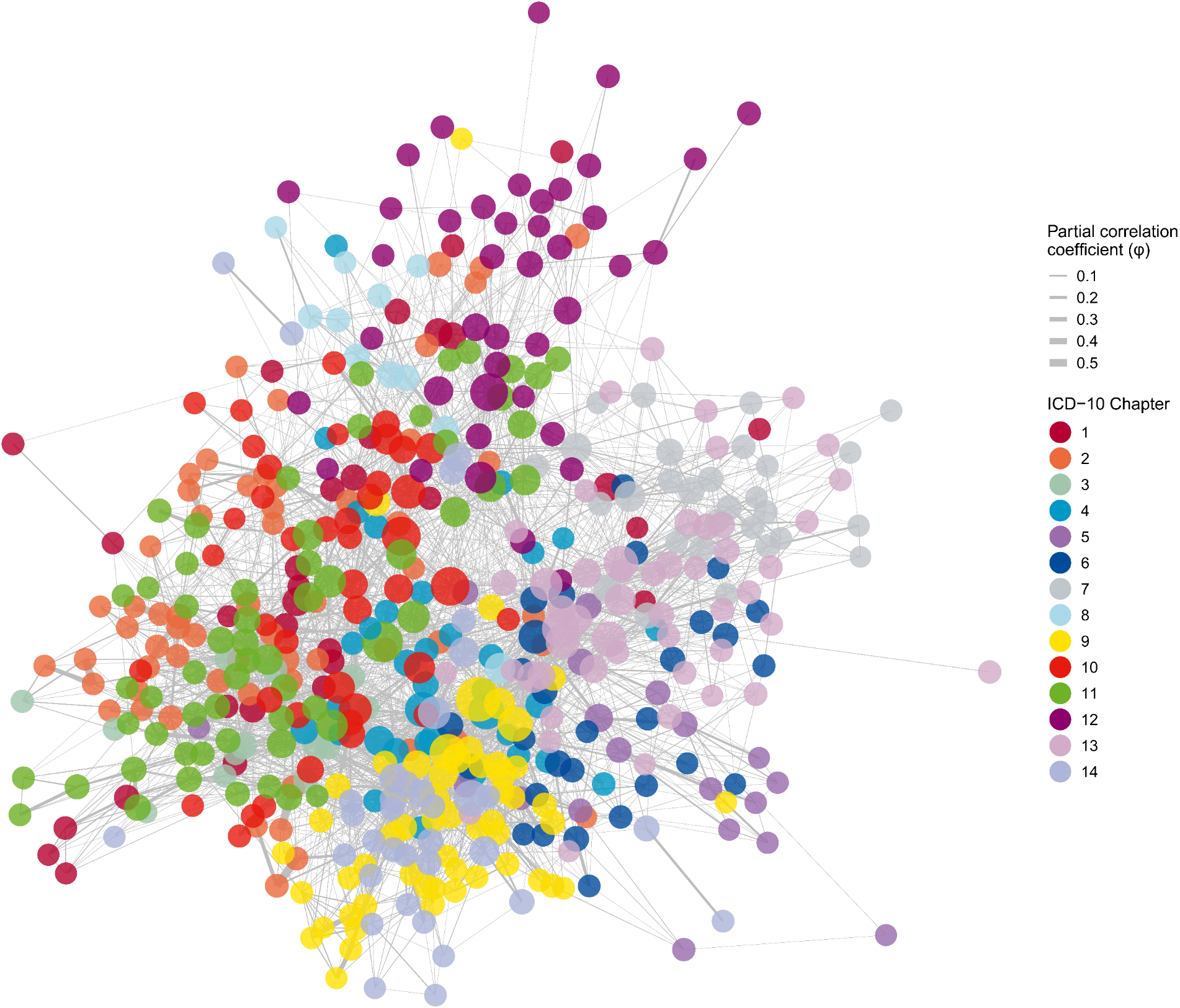

S5B

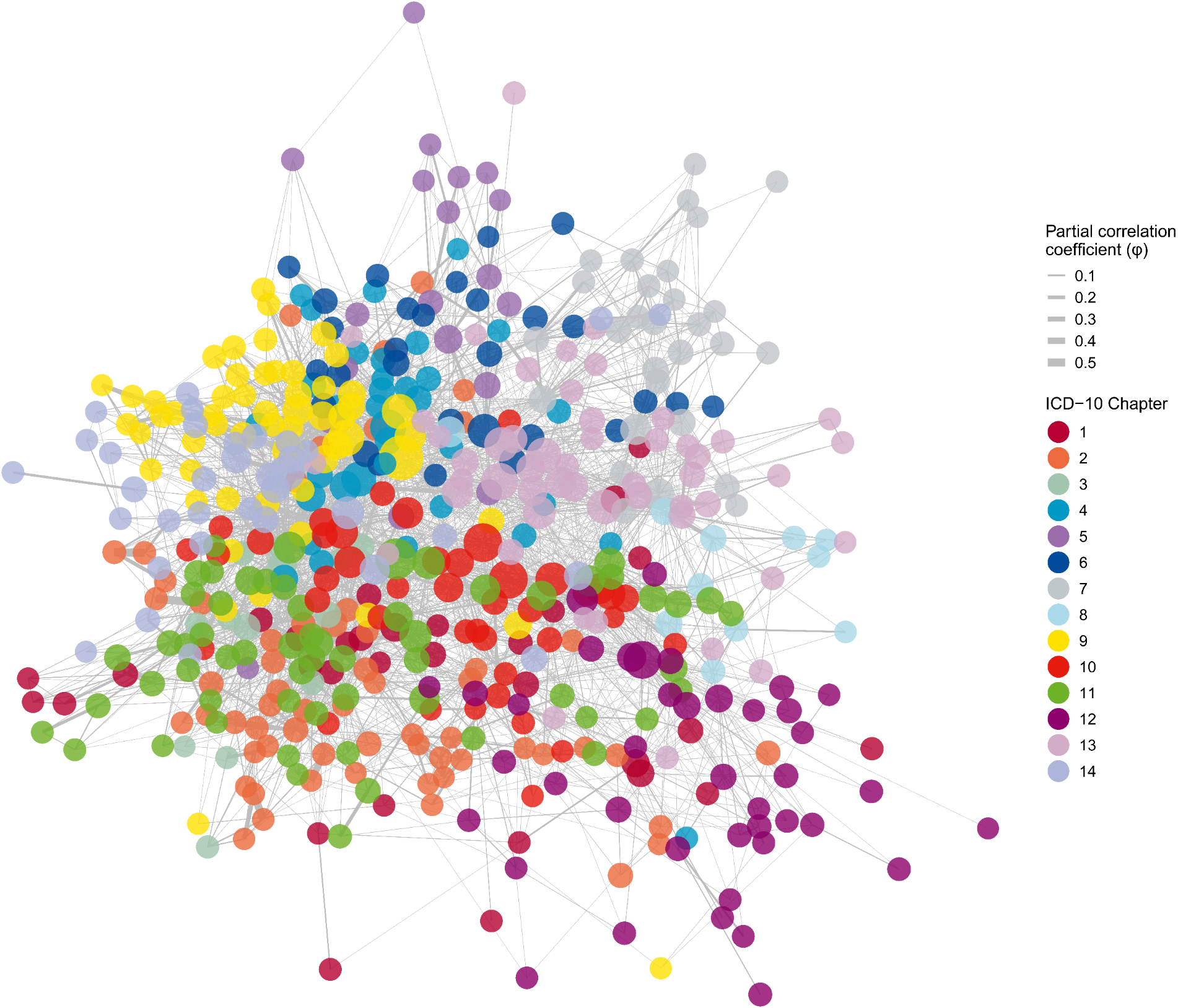

S5C

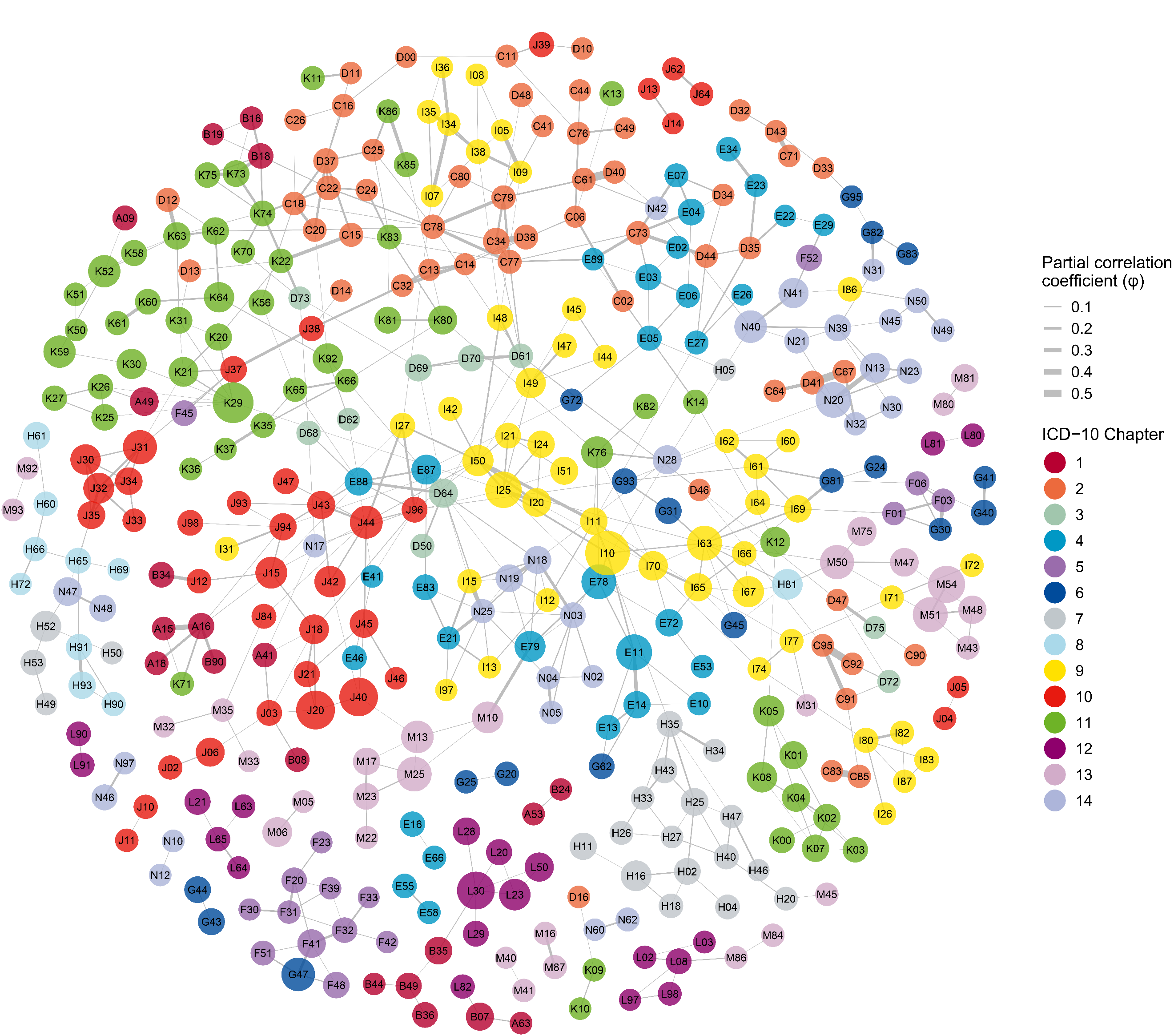

S5D

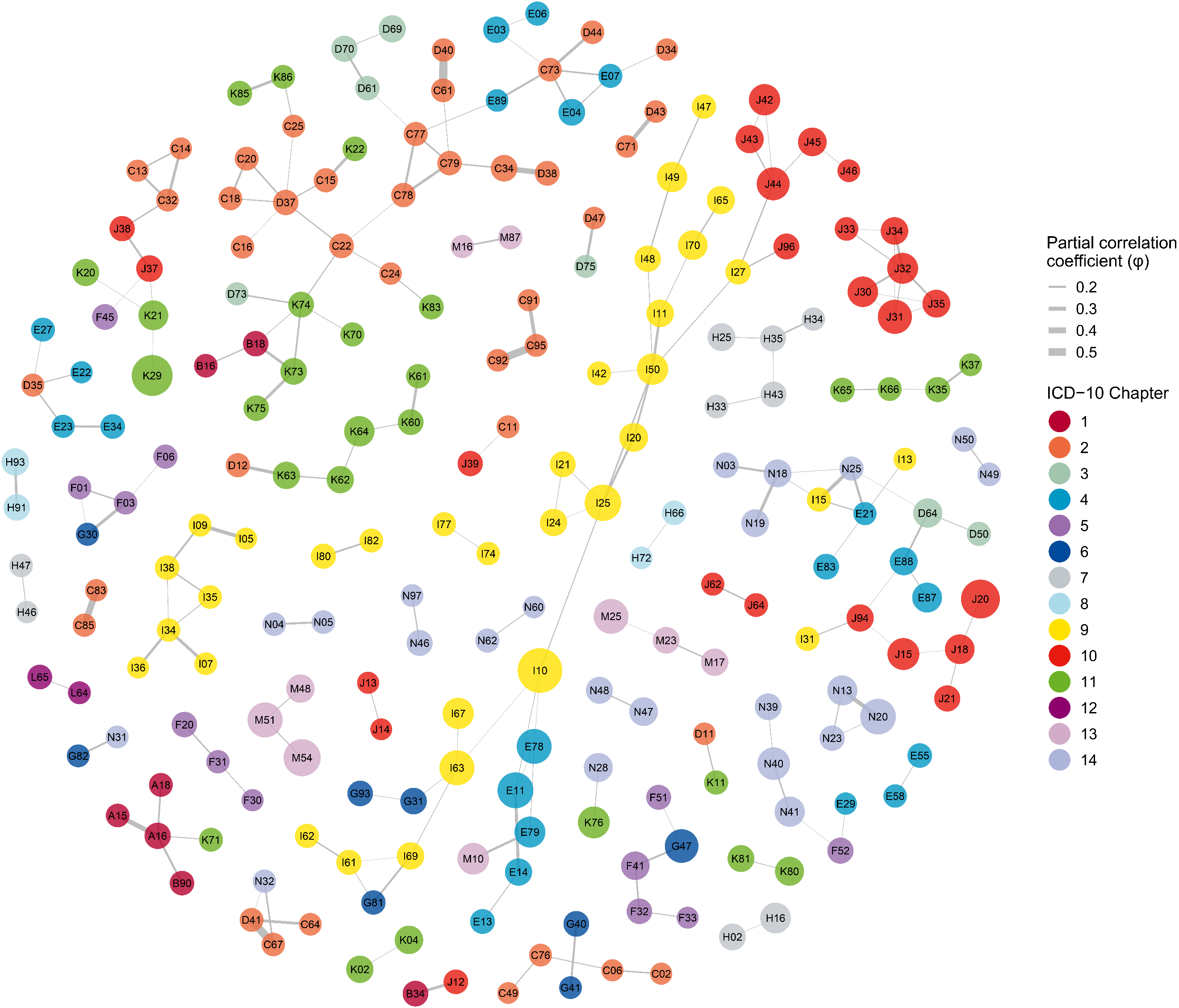

Supplementary Figure S6. Multimorbidity Networks across different age groups.

Nodes represent individual ICD‑10 conditions; node size is proportional to each condition’s period prevalence (2016–2023) within the specific age group, and node color denotes the ICD‑10 chapter. Edges depict pairwise partial correlation coefficients (φ) between conditions after controlling for all other diseases; edge width is scaled to the magnitude of φ. As networks including all statistically significant positive correlations (φ > 0) can be very dense and difficult to interpret, these figures present the networks filtered for associations meeting FDR-adjusted *P* < 0.05 and partial correlation φ ≥ 0.05. Sub-figures represent different age groups: (A) 0–9 years, (B) 10–19 years, (C) 20–29 years, (D) 30–39 years, (E) 40–49 years, (F) 50–59 years, (G) 60–69 years, (H) 70–79 years, and (I) ≥ 80 years.

S6A

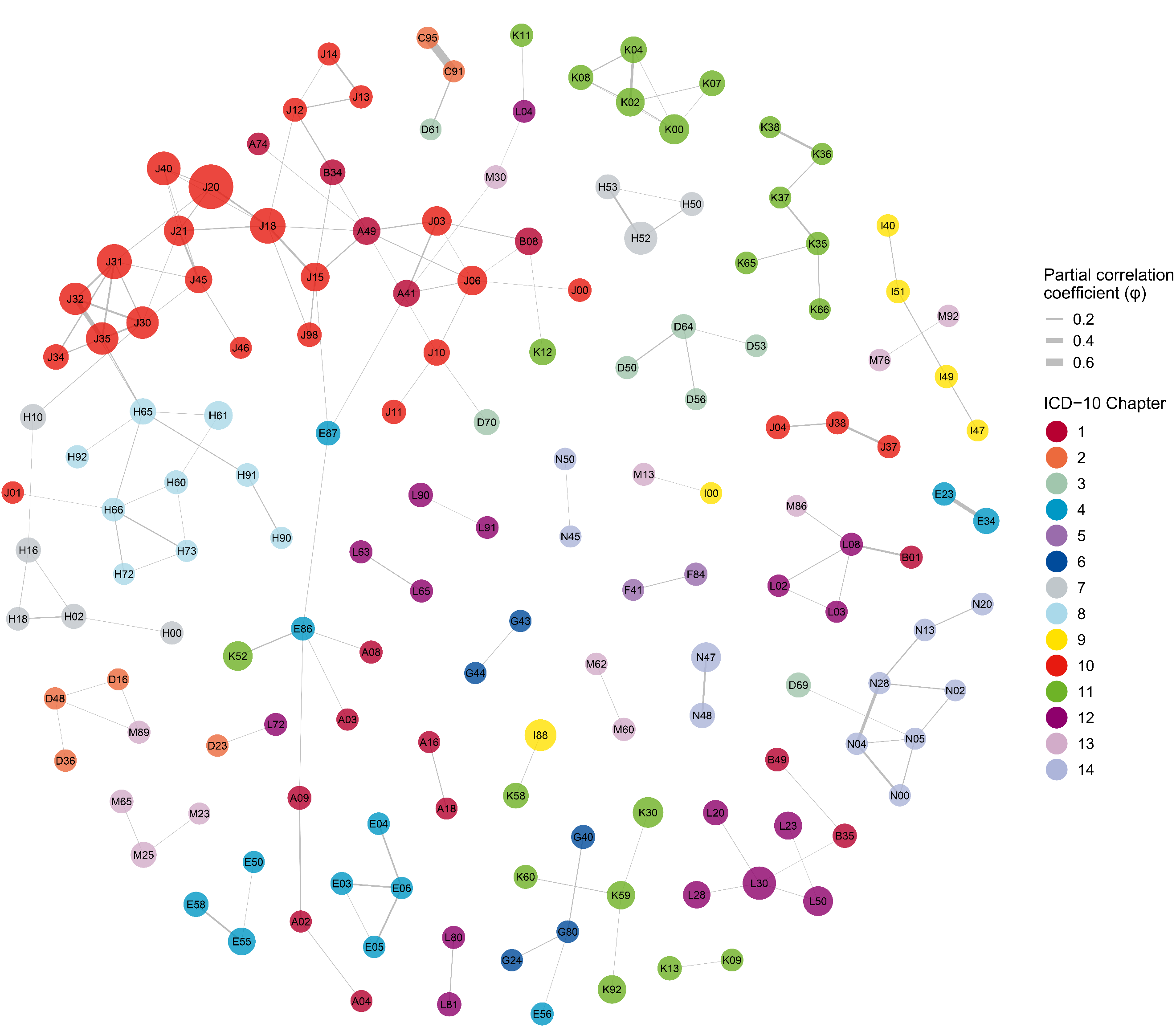

S6B

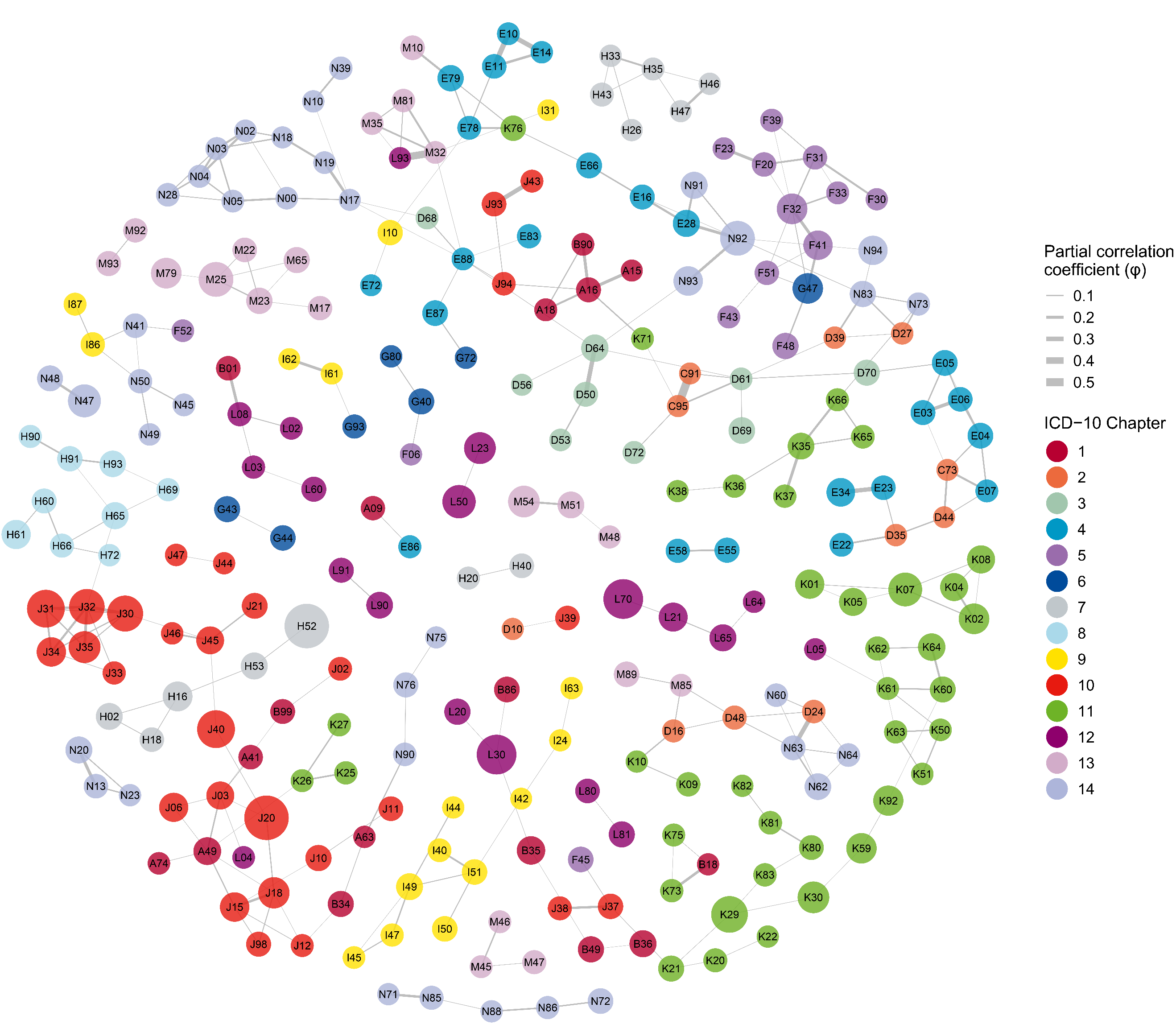

S6C

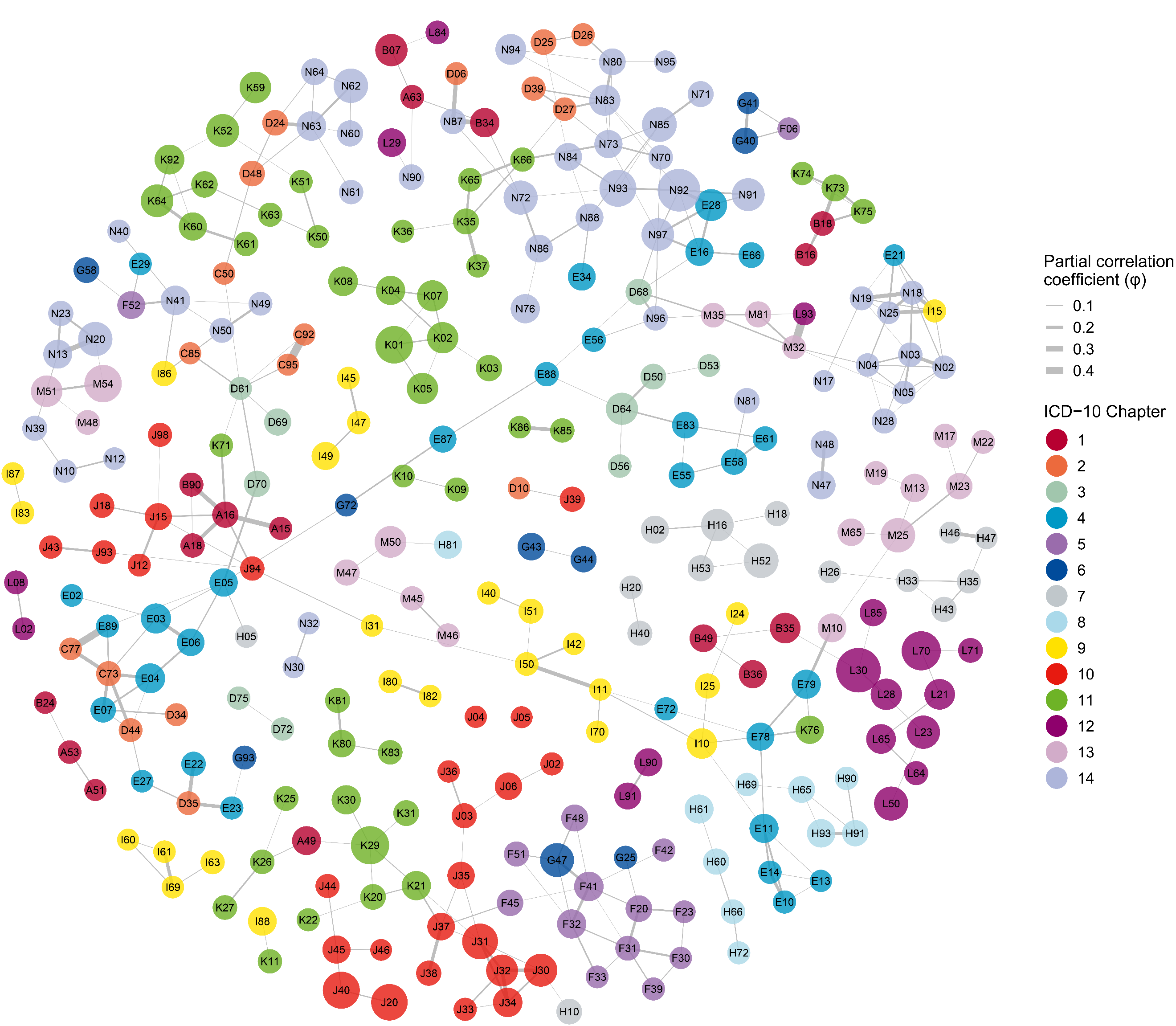

S6D

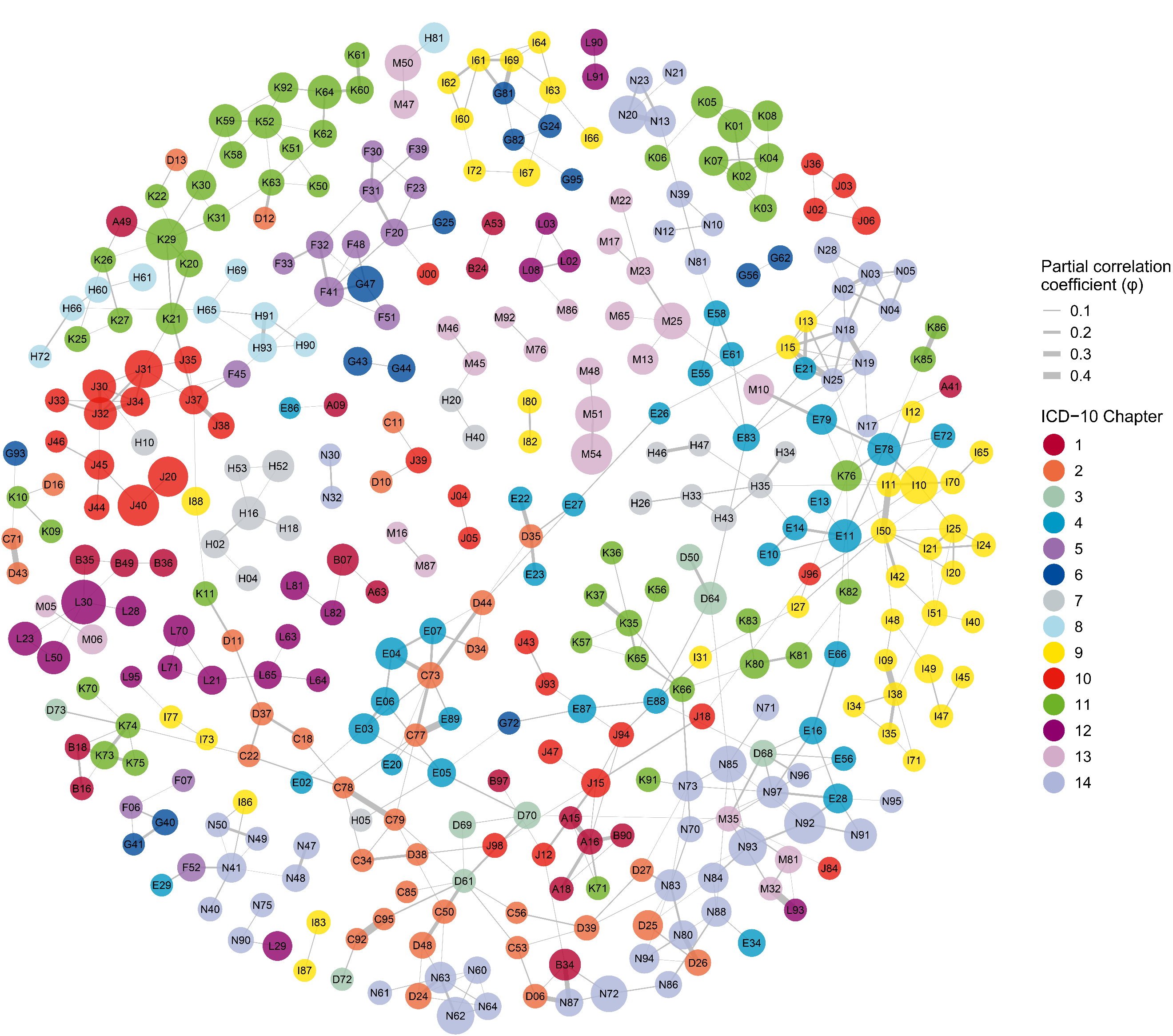

S6E

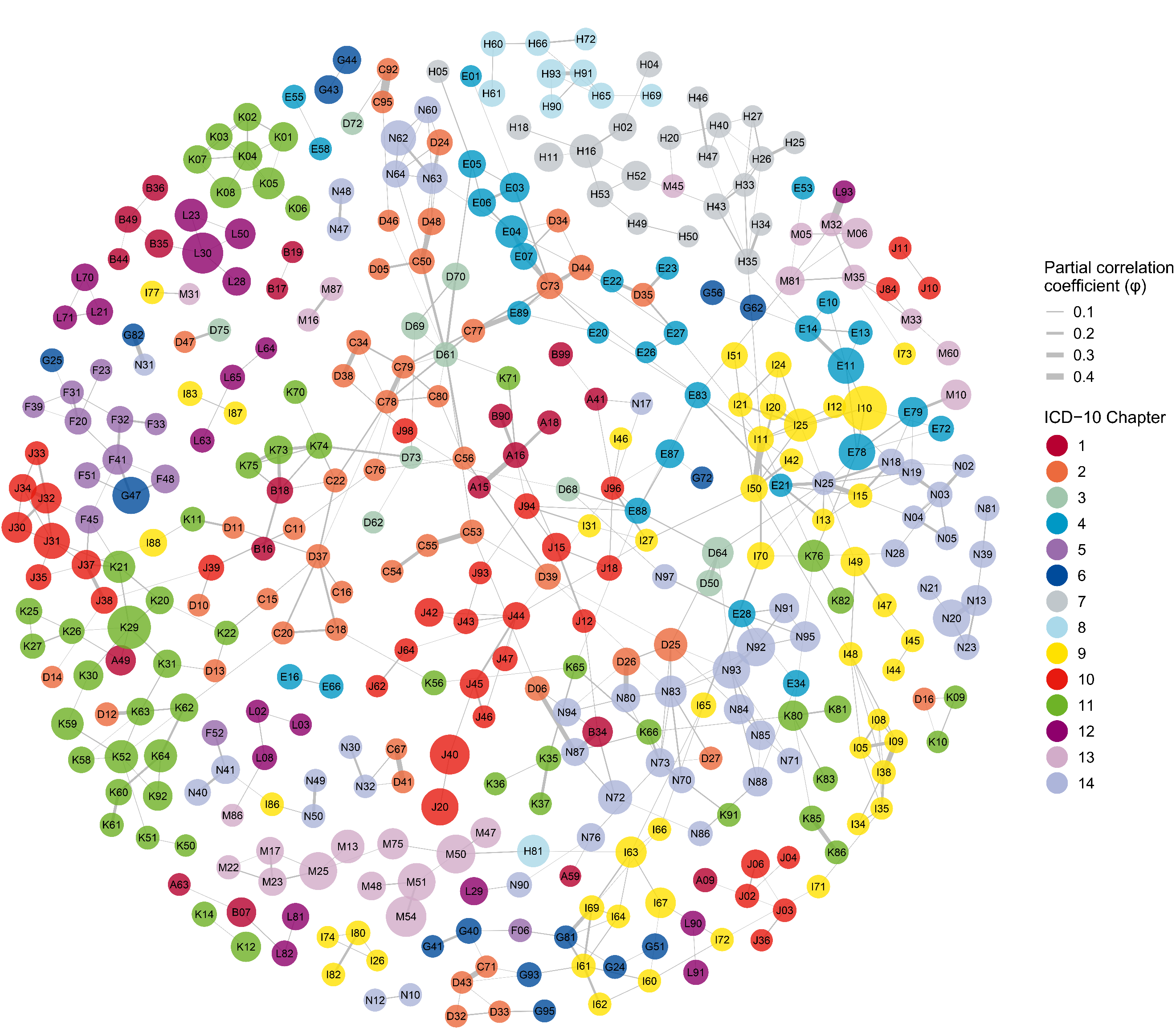

S6F

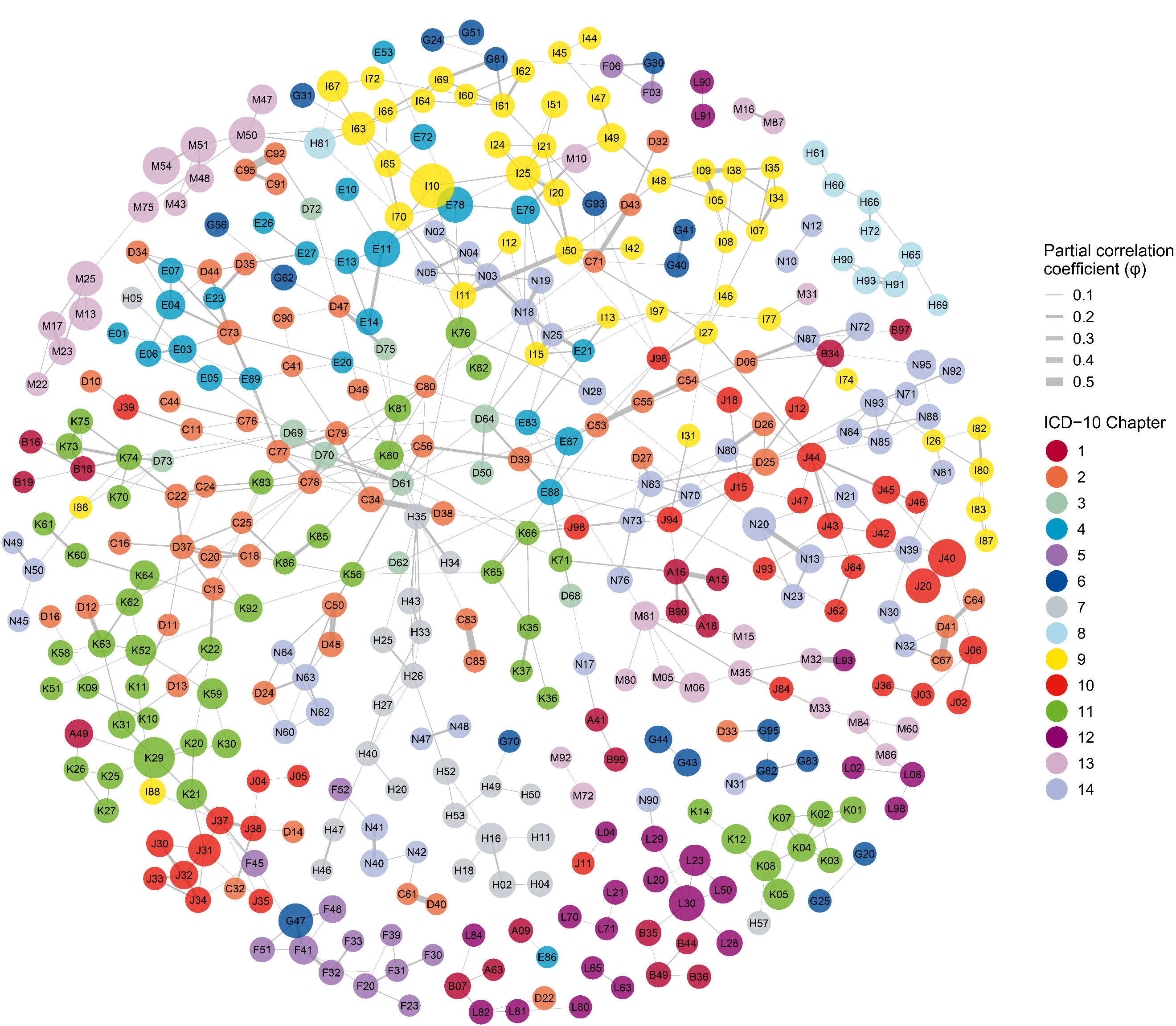

S6G

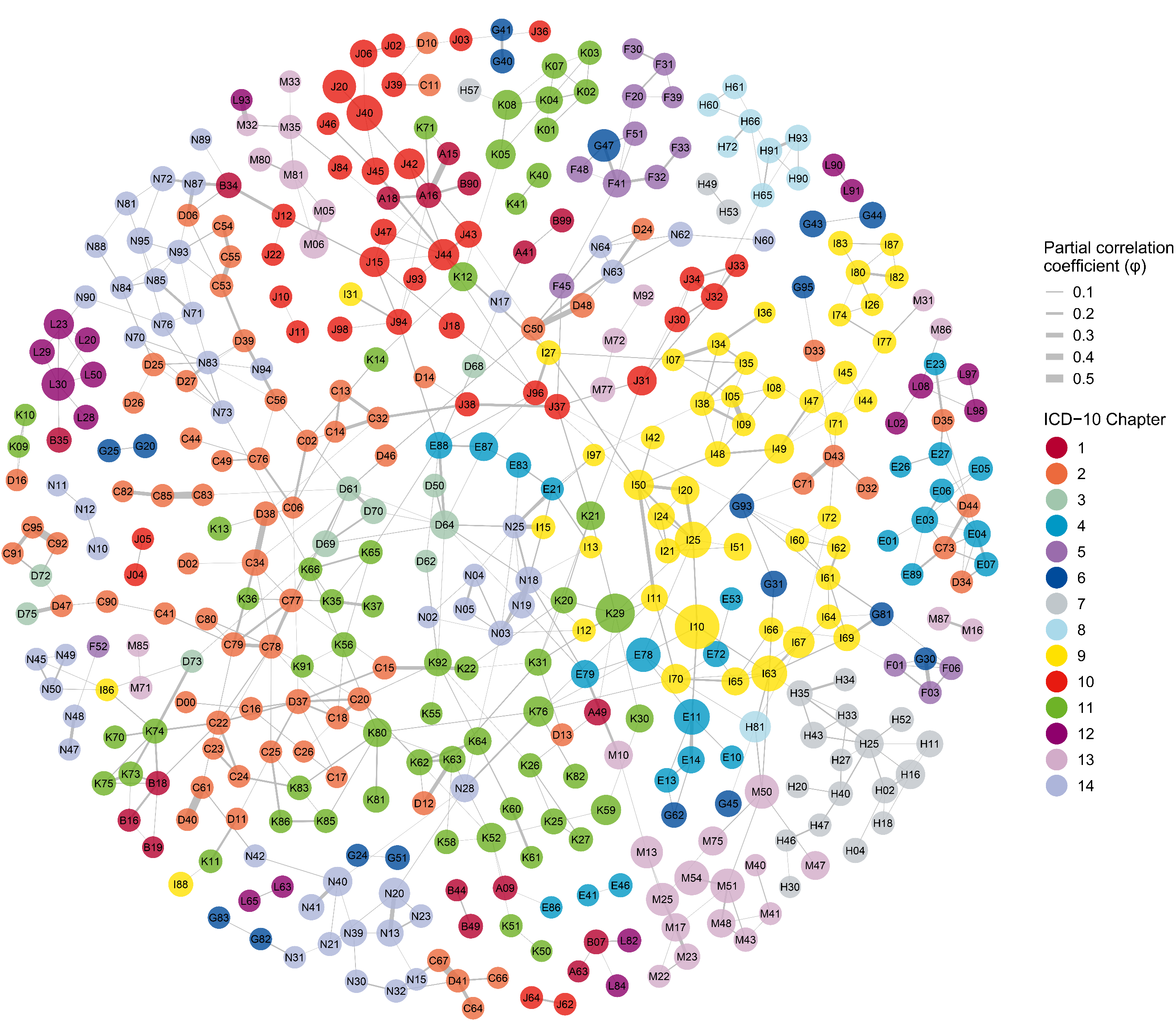

S6H

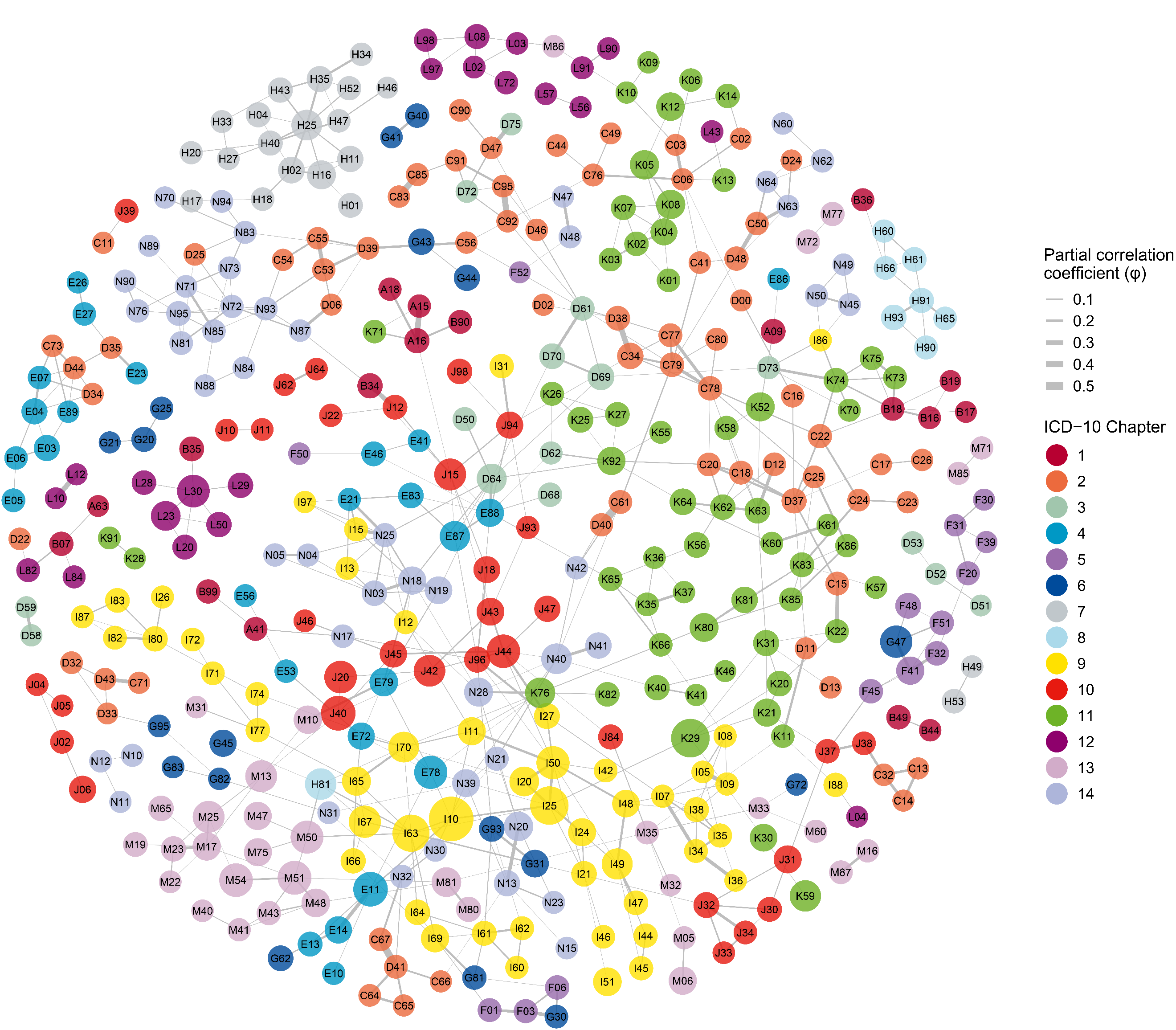

S6I

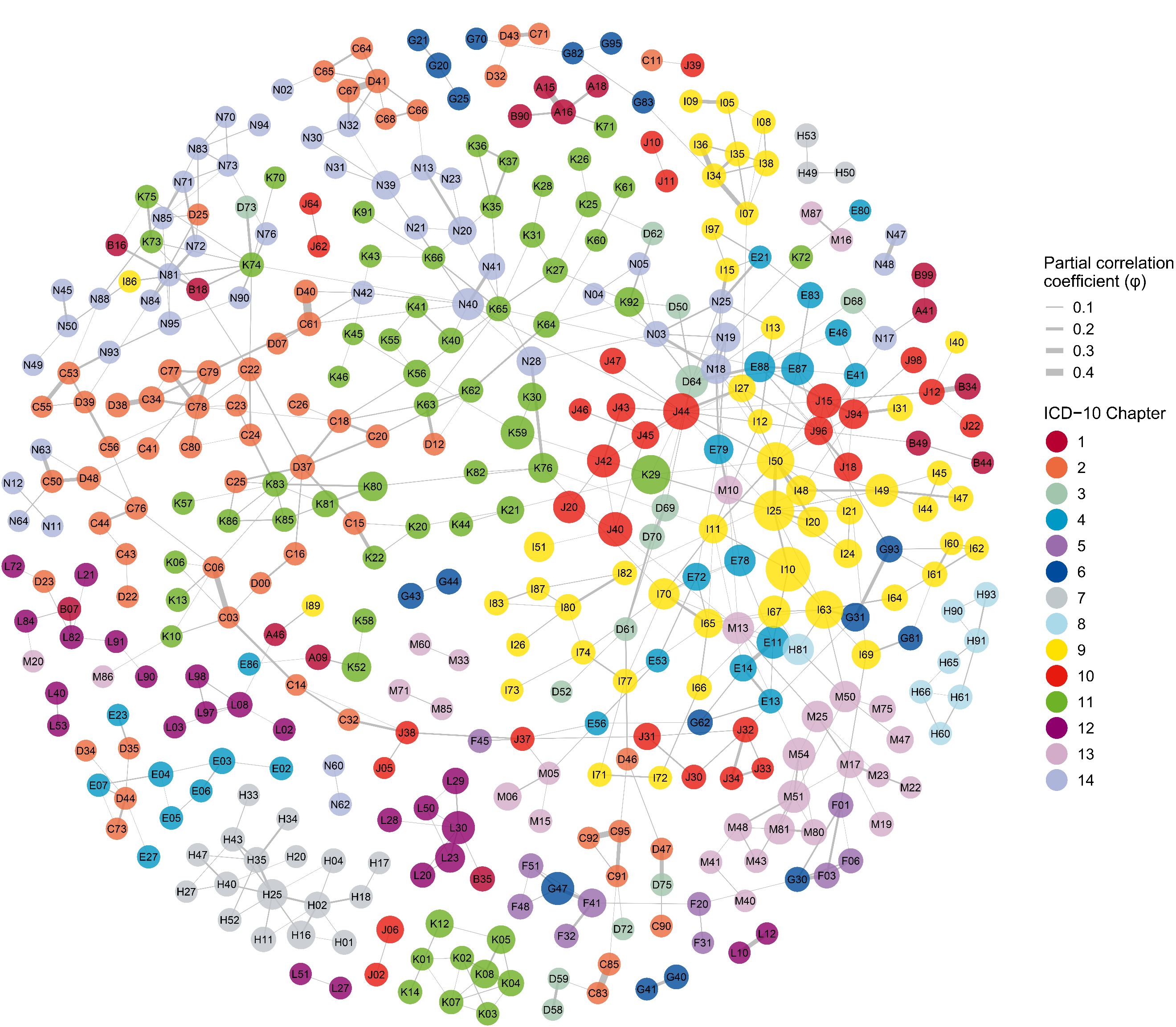

Supplementary Figure S7. Multimorbidity subnetworks for the top 20 hub diseases in overall population identified by Multimorbidity Coefficient (MMC).

(A-T) Each panel displays one of the top 20 hub diseases (center node) and its neighboring conditions using a star layout. Nodes represent individual ICD‑10 conditions (labeled with ICD-3 codes); node size is proportional to each condition’s period prevalence (2016–2023), and node color denotes the ICD‑10 chapter. Edges depict significant adjusted associations between pairs of conditions after controlling for all other diseases in the network (quantified by the partial correlation coefficient, φ). Only associations meeting FDR‑adjusted *P* < 0.05 and φ > 0.01 are shown. Edge width is scaled to the magnitude of φ. Hubs are presented in descending order of MMC: (A) [K29], (B) [E88], (C) [I50], (D) [D64], (E) [L30], (F) [E87], (G) [I70], (H) [J15], (I) [K76], (J) [I63], (K) [M81], (L) [G47], (M) [D61], (N) [I10], (O) [E78], (P) [J44], (Q) [J31], (R) [I25], (S) [N18], (T) [I69]. An interactive version of these subnetworks for the overall, female, and male populations is available at: https://pumc-multimorbidity.shinyapps.io/hub-disease-network/.

S7A

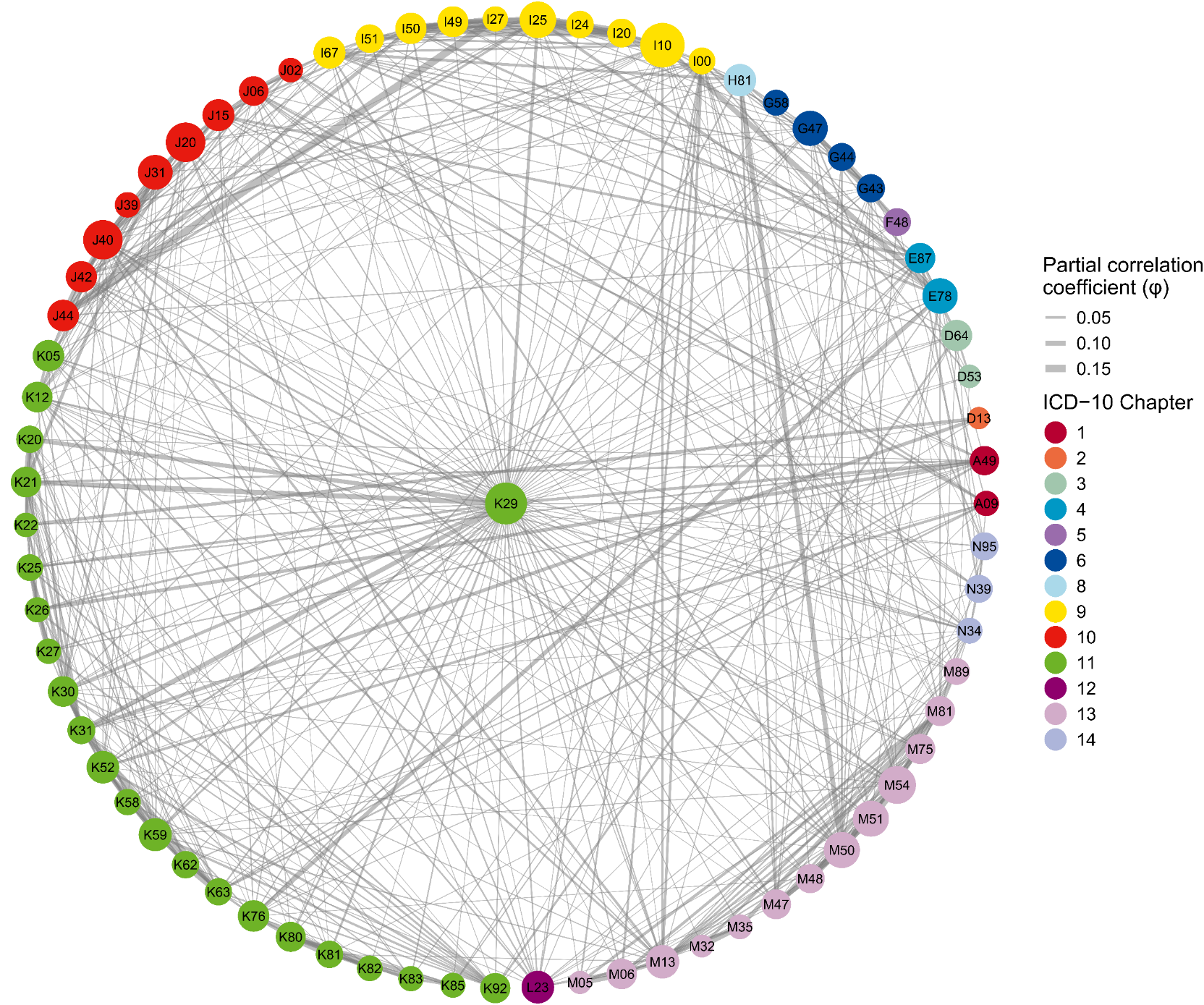

S7B

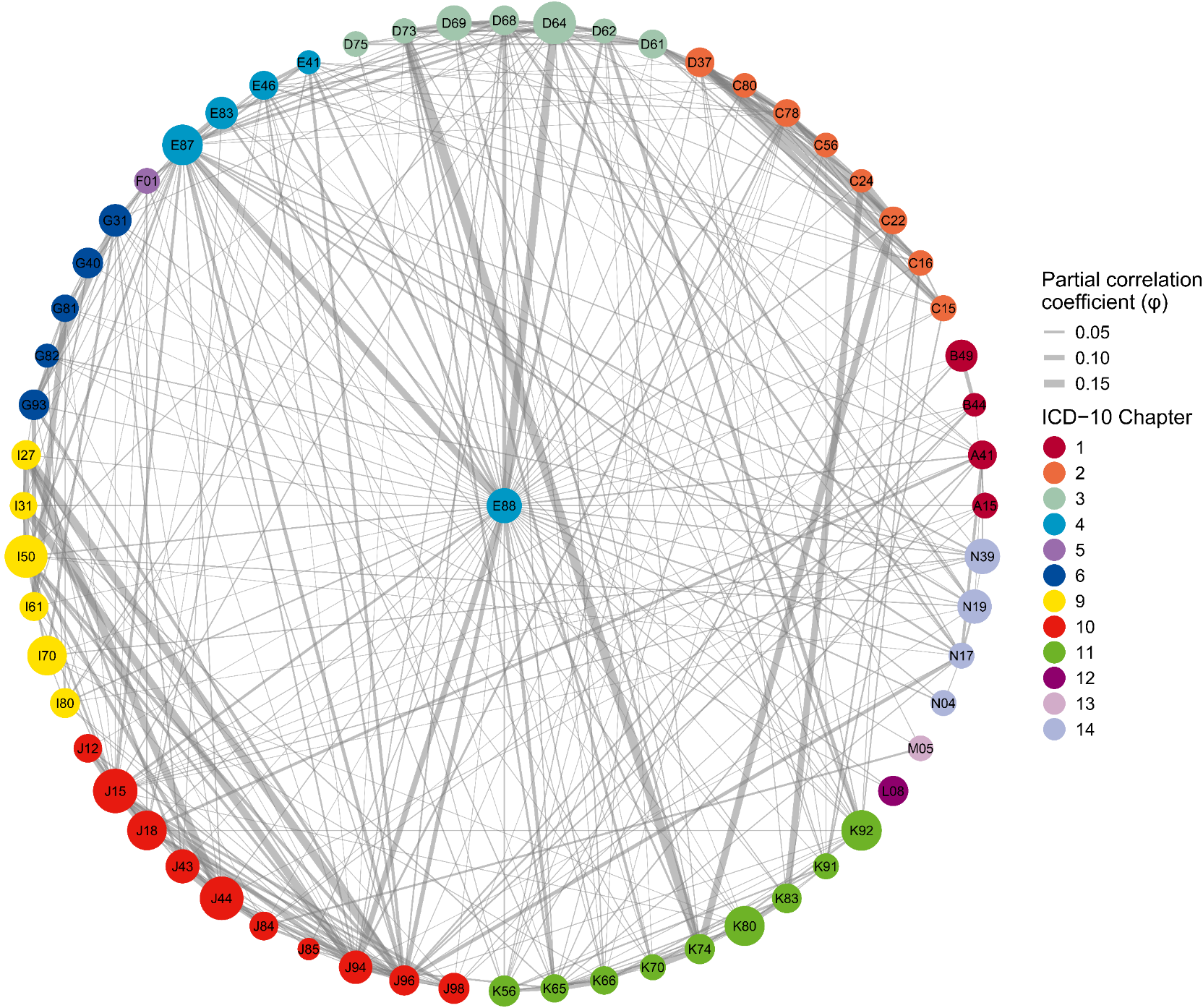

S7C

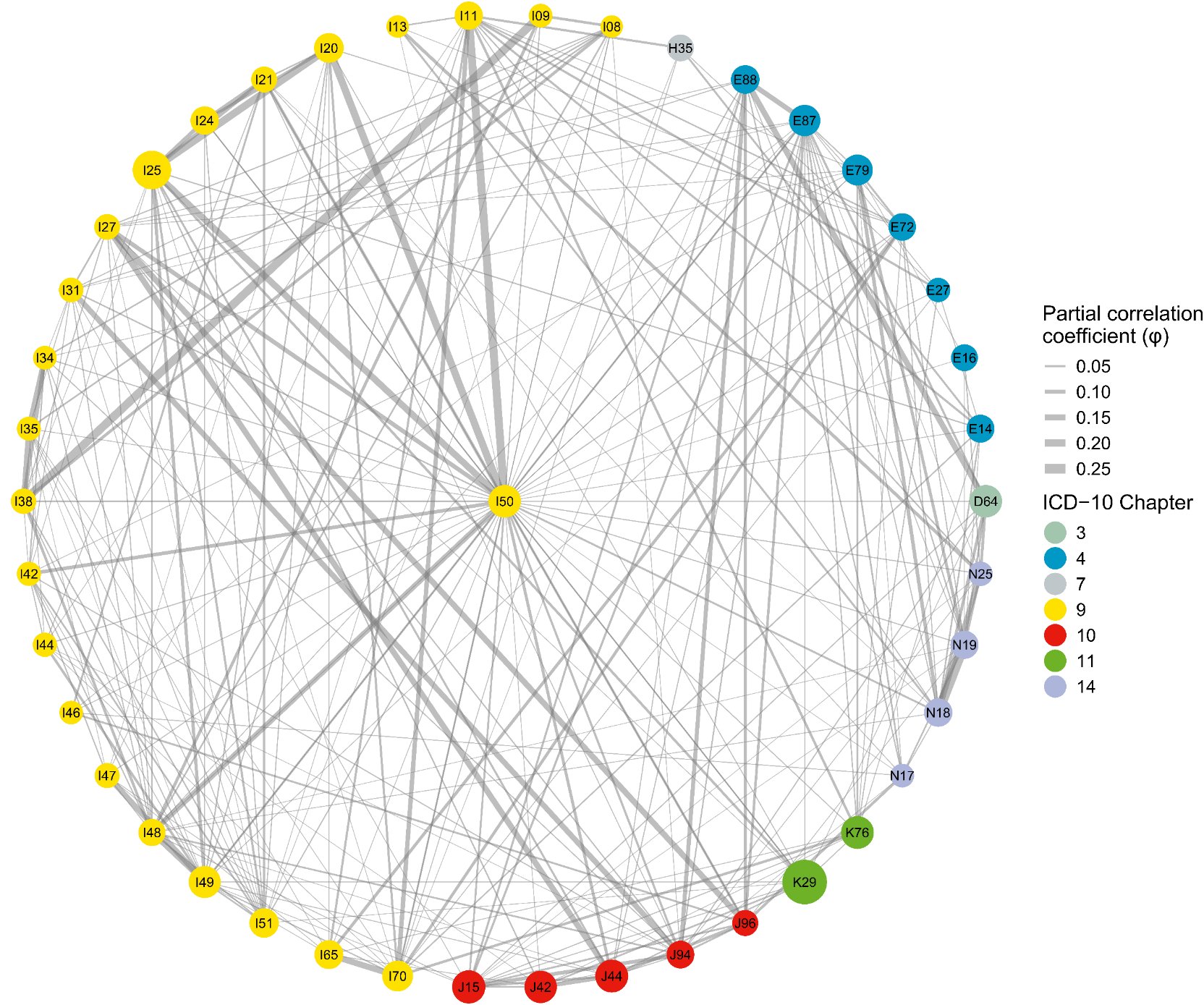

S7D

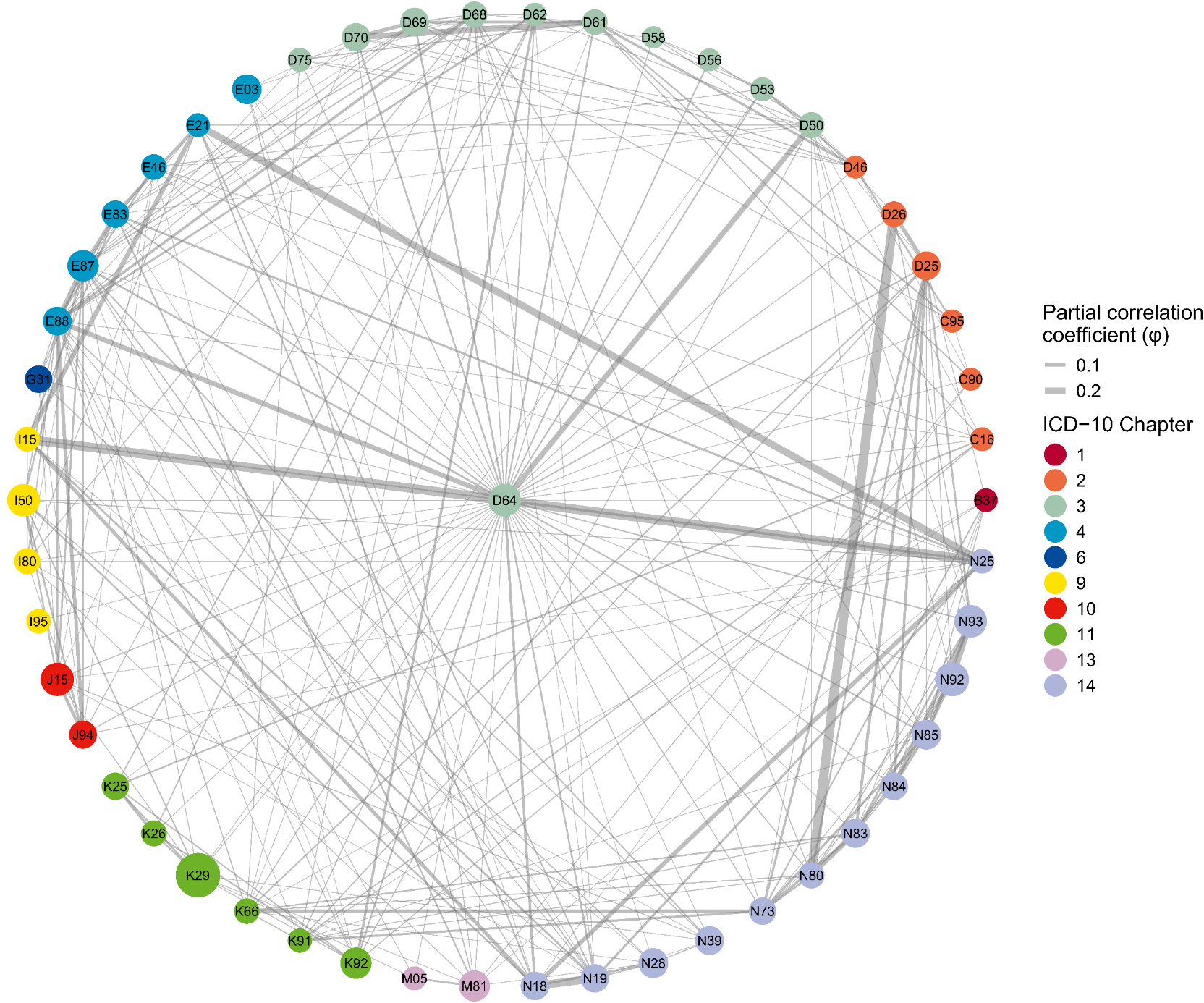

S7E

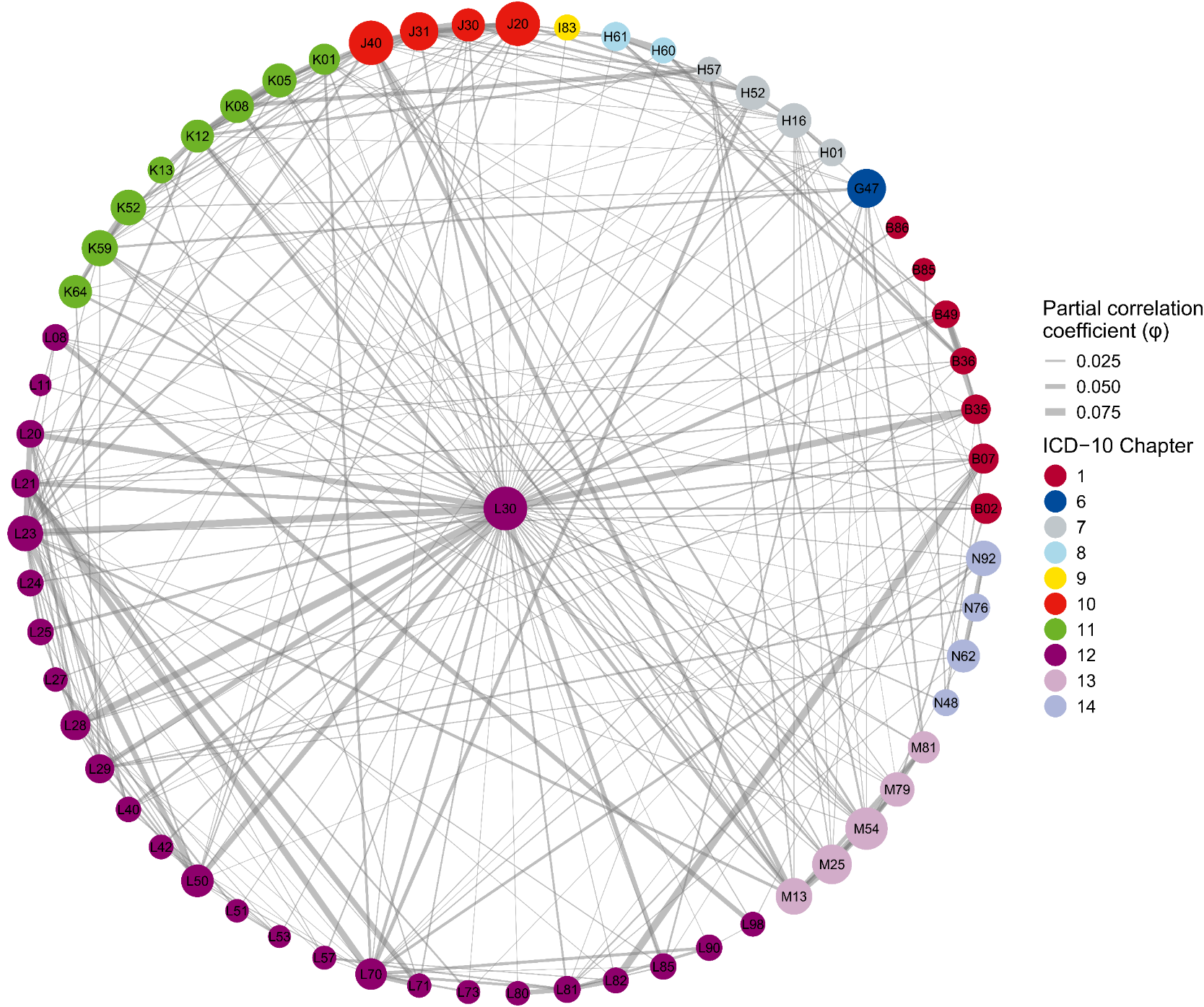

S7F

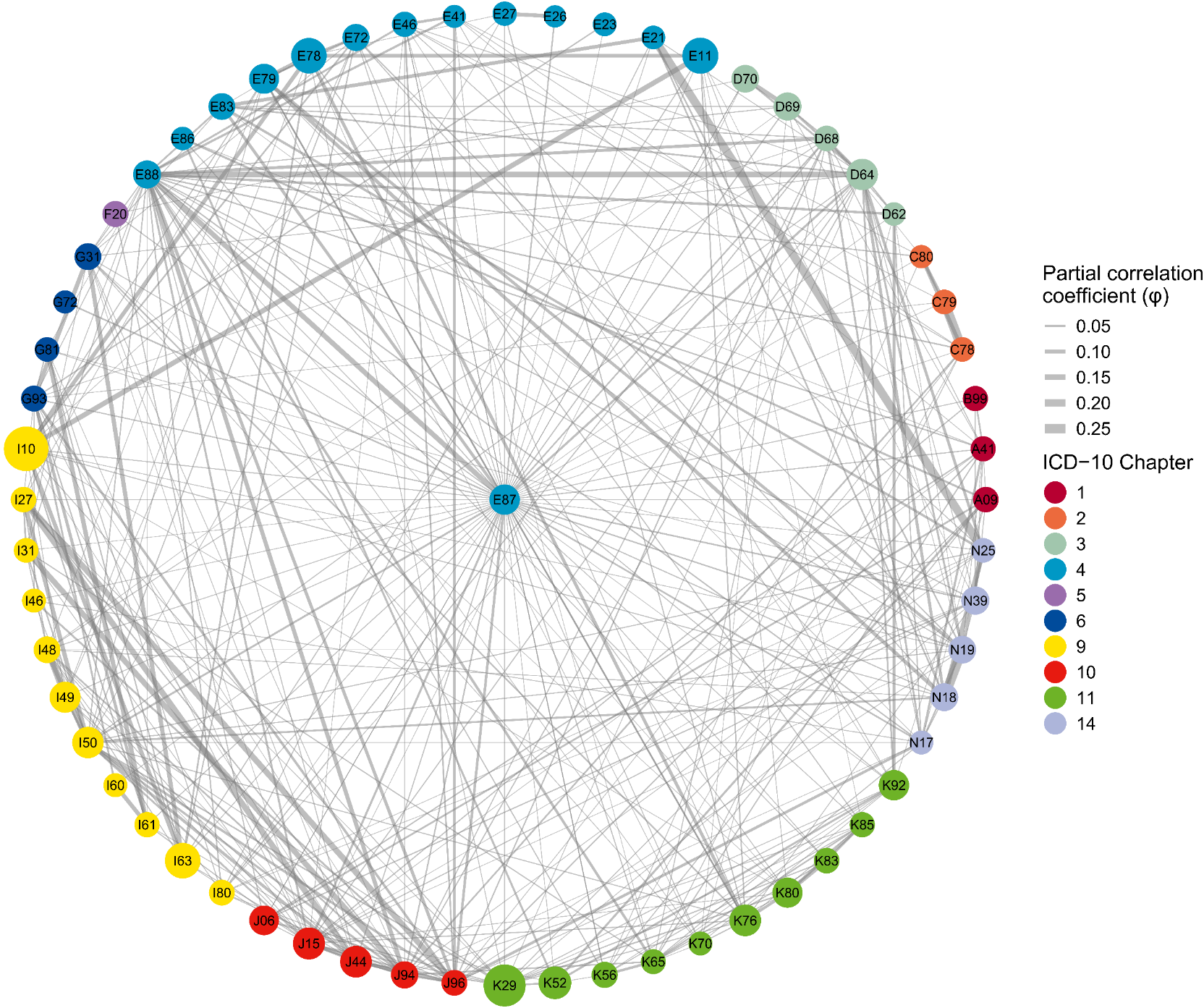

S7G

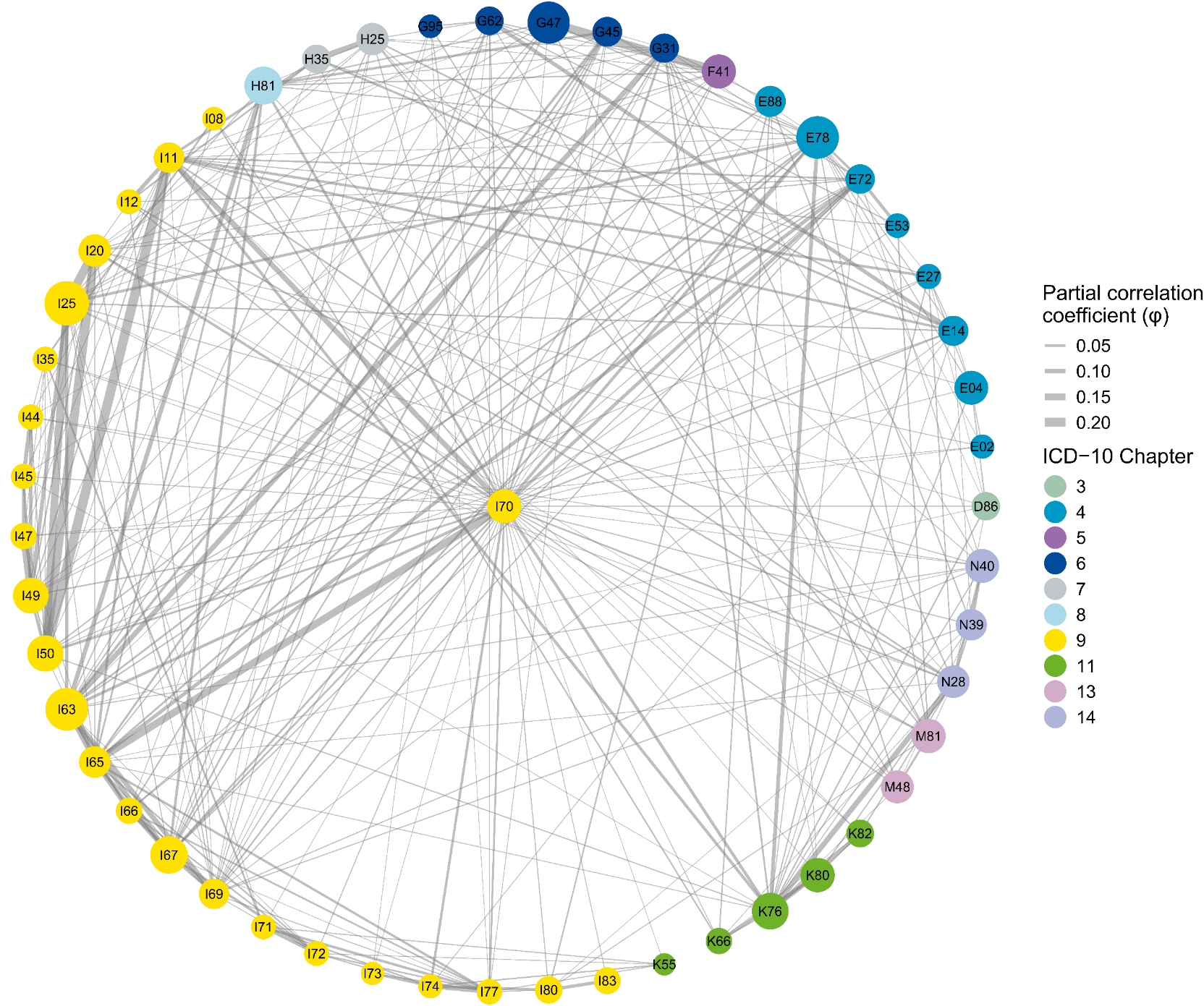

S7H

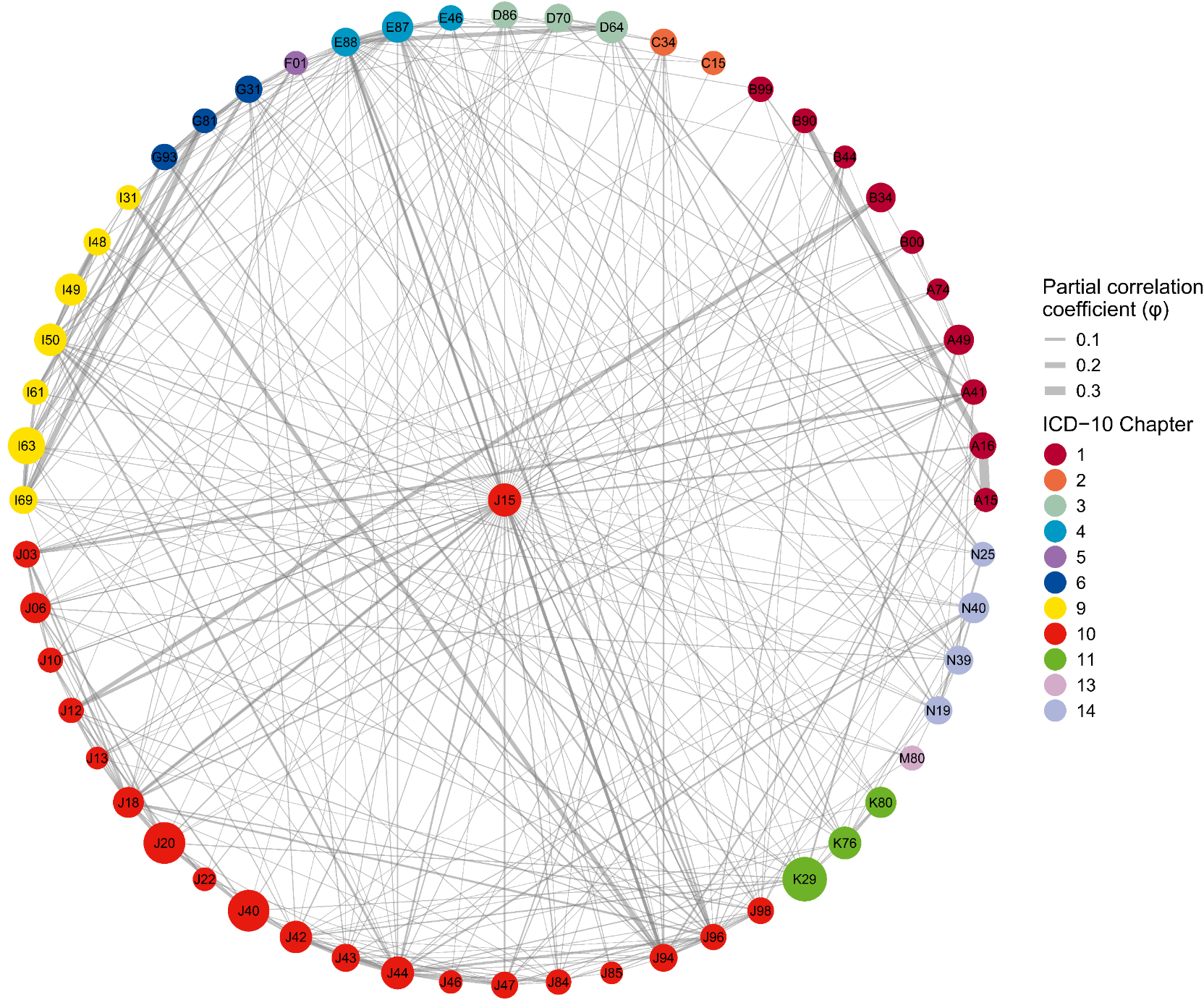

S7I

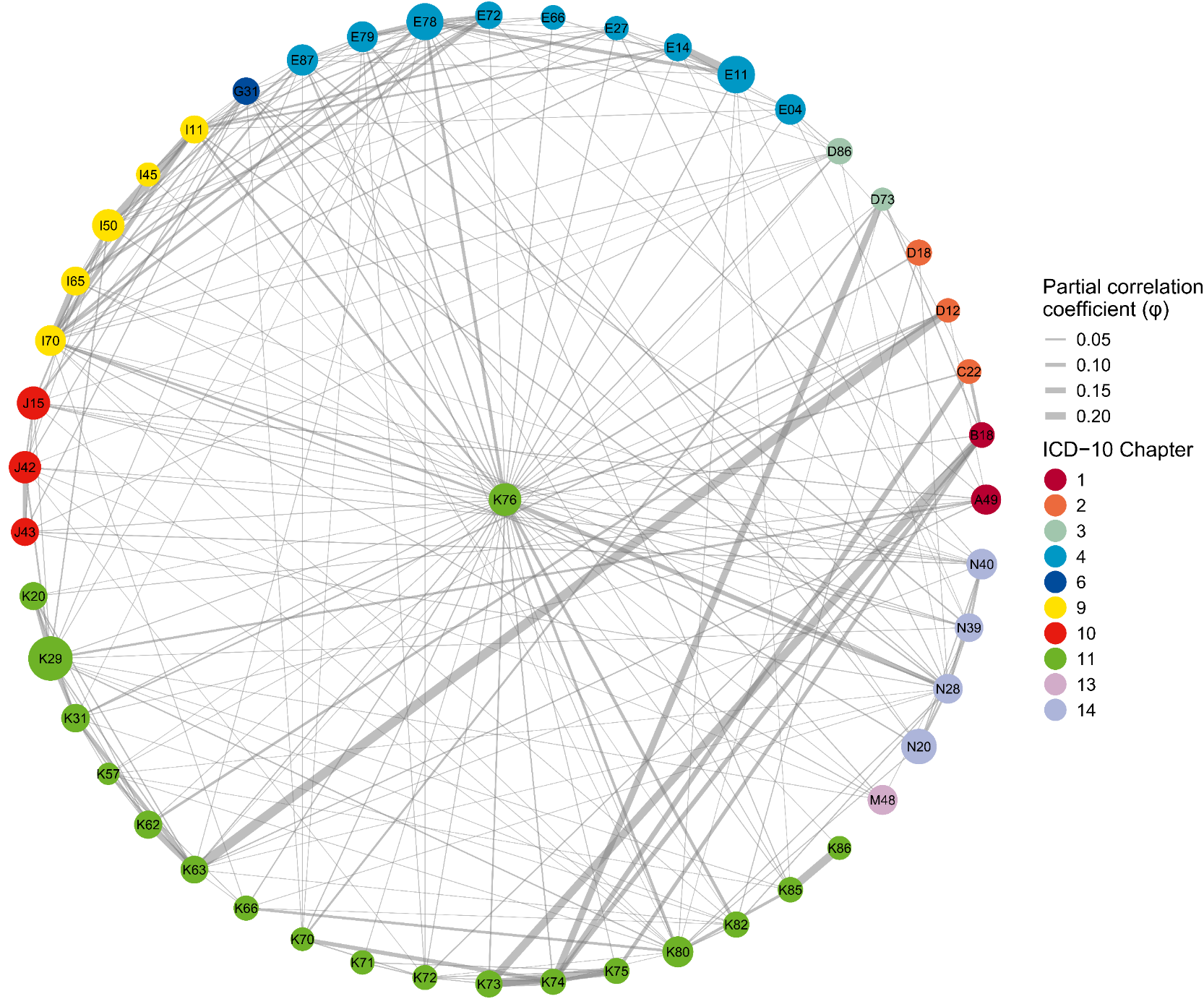

S7J

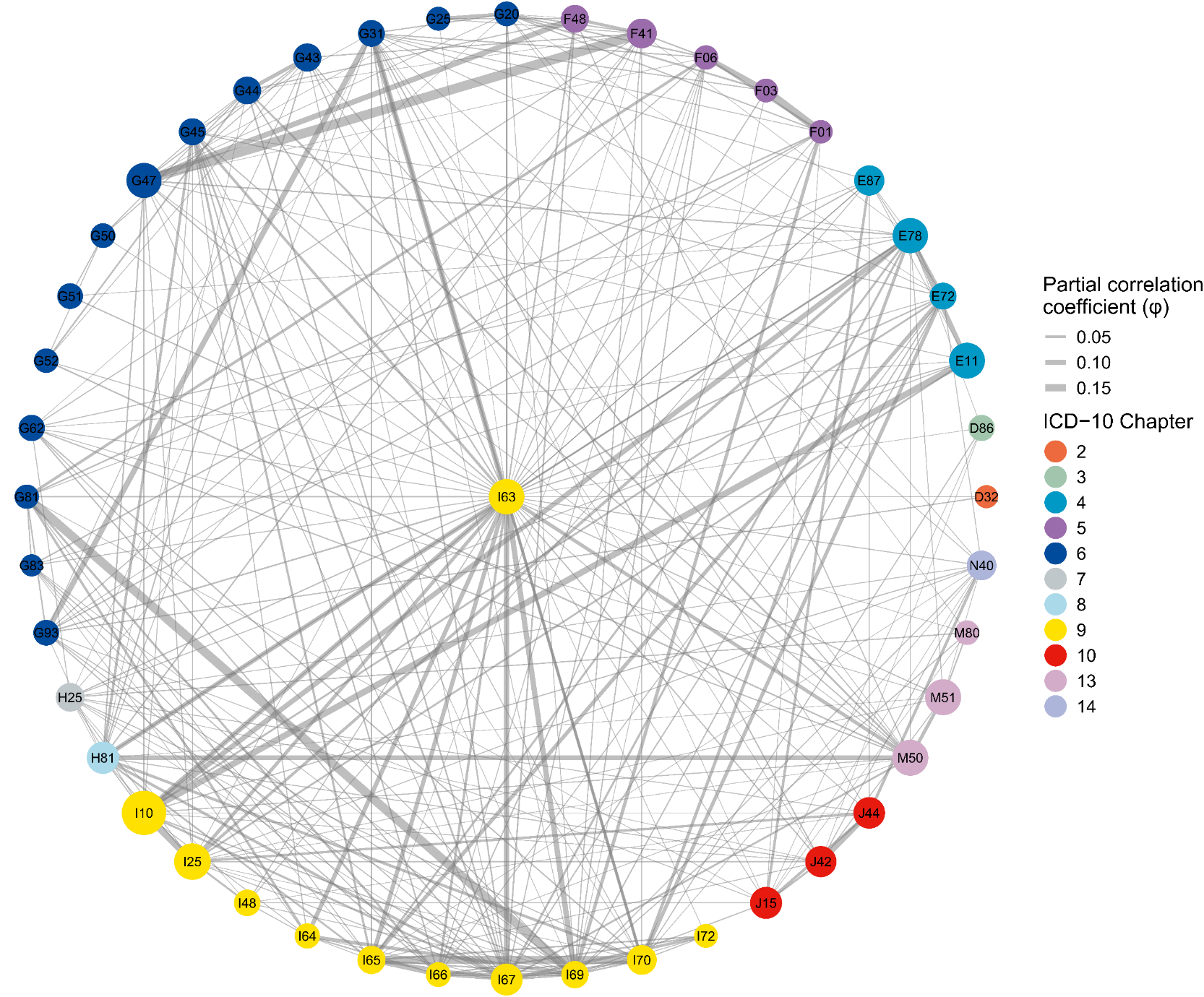

S7K

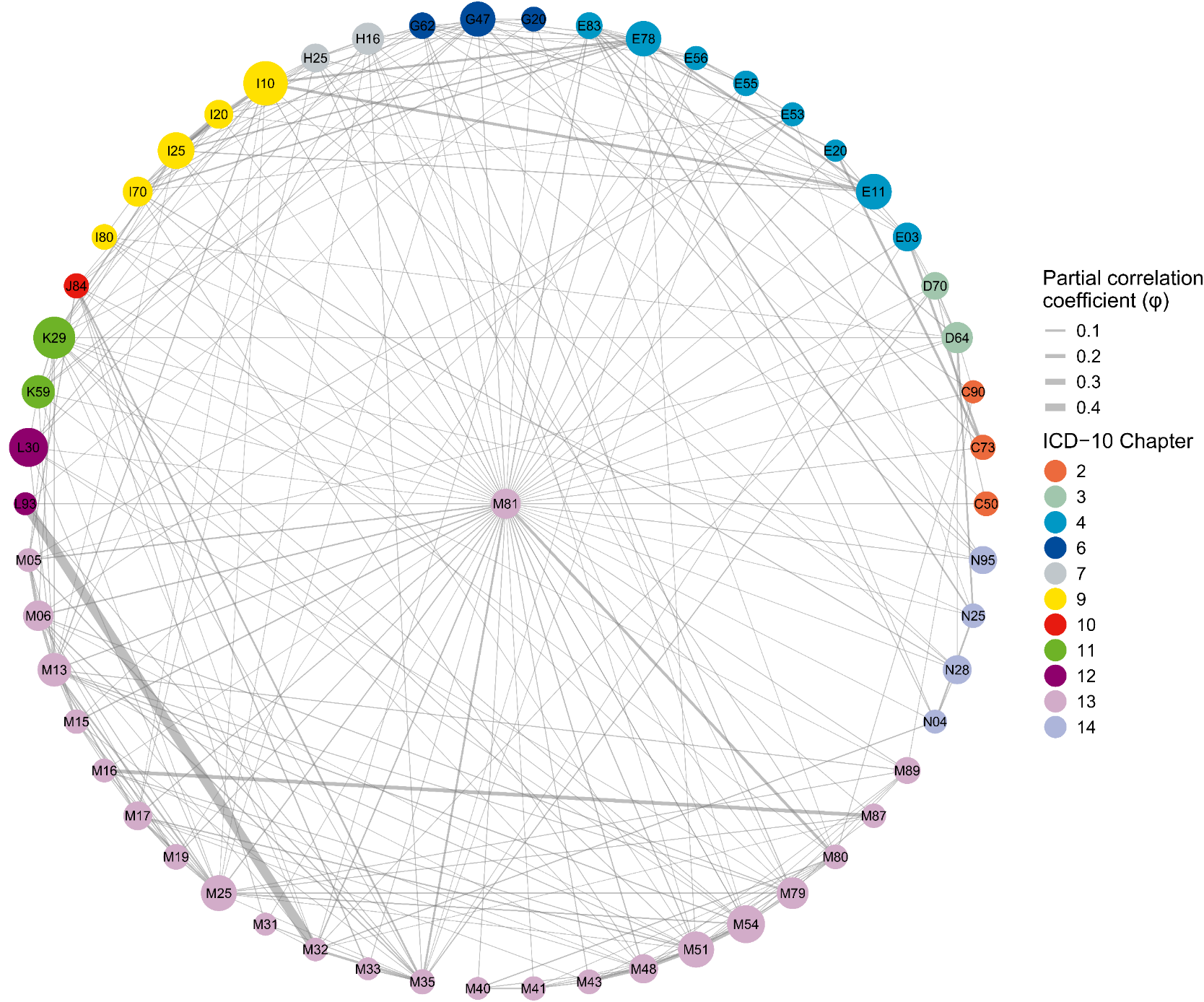

S7L

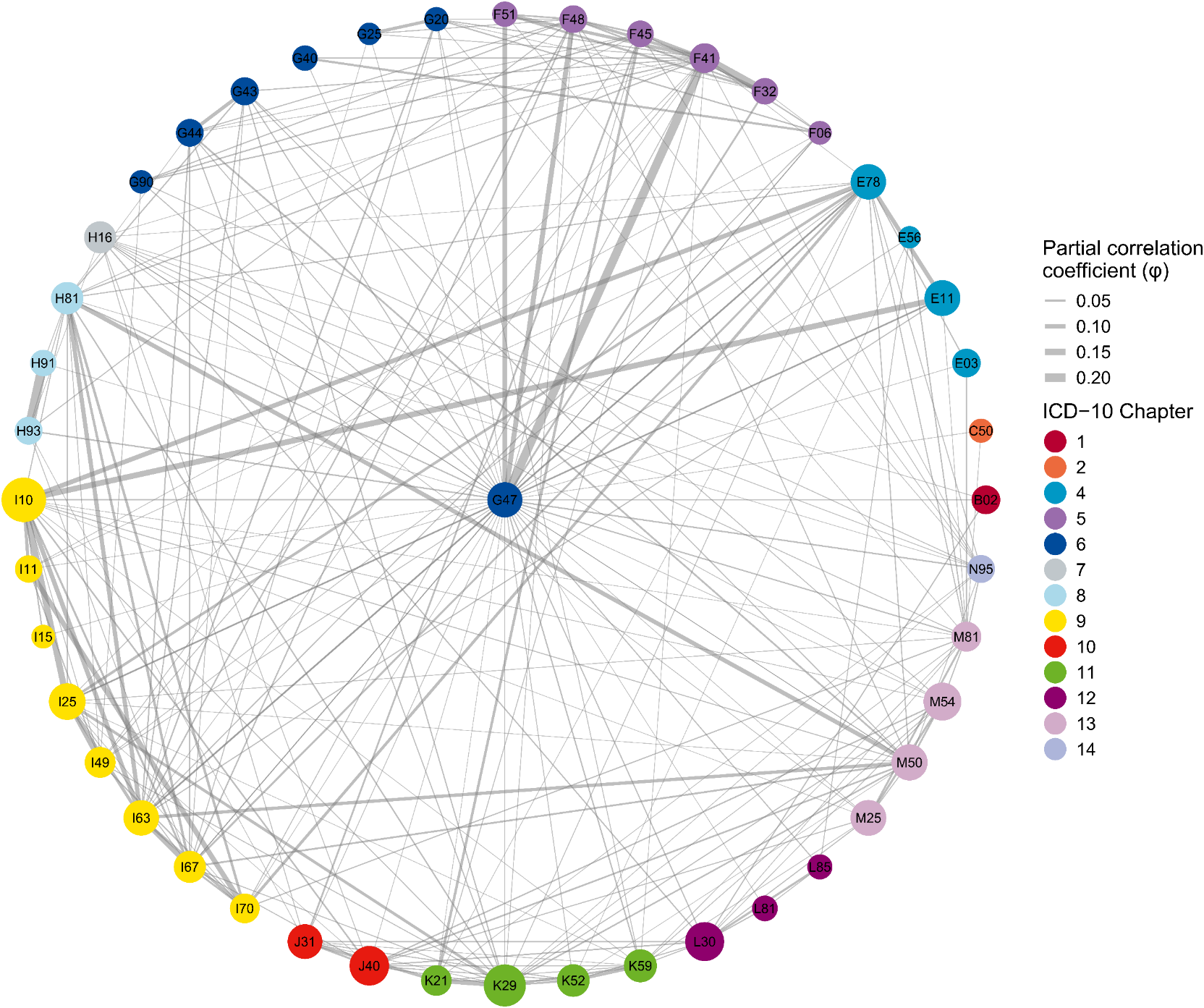

S7M

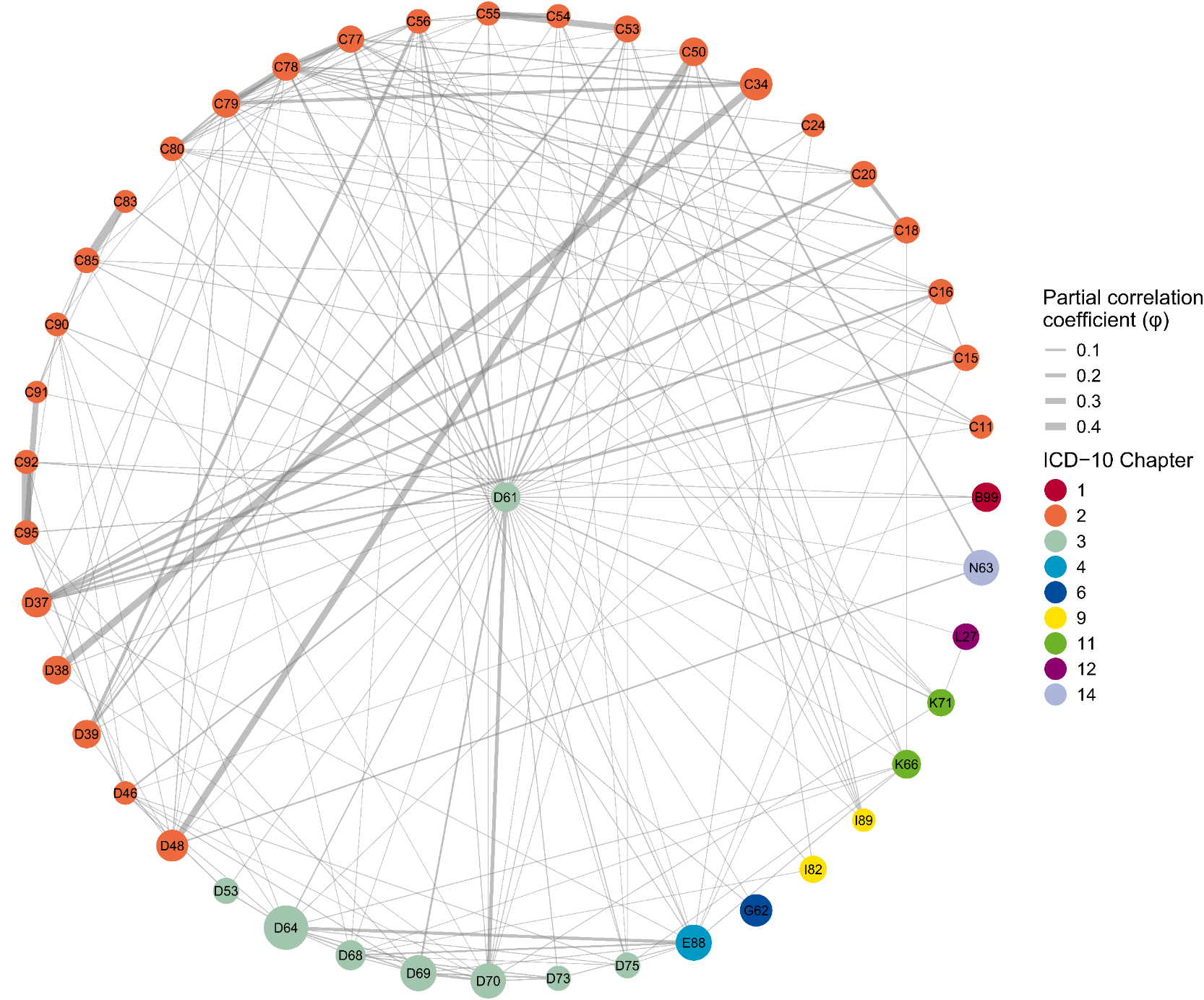

S7N

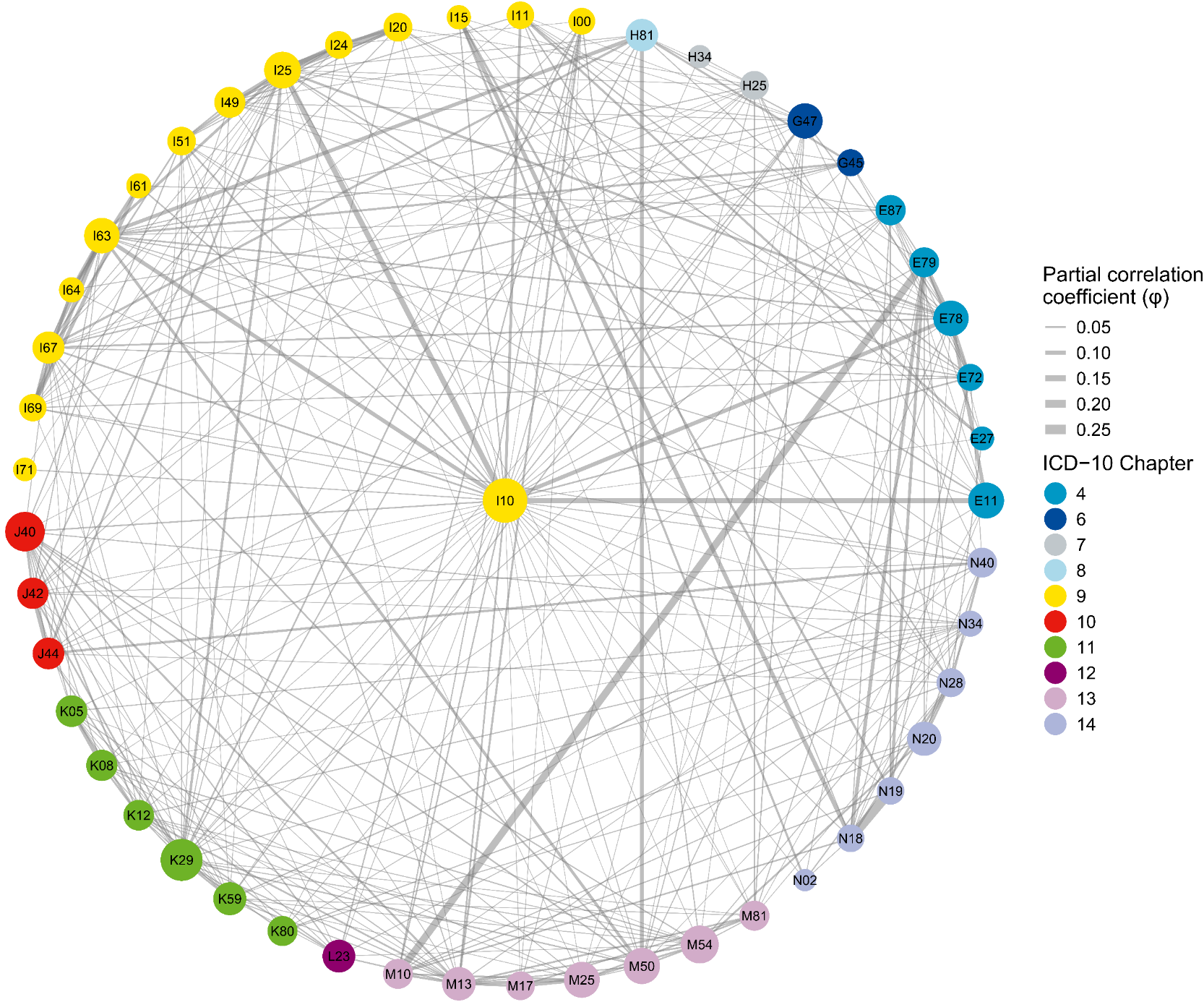

S7O

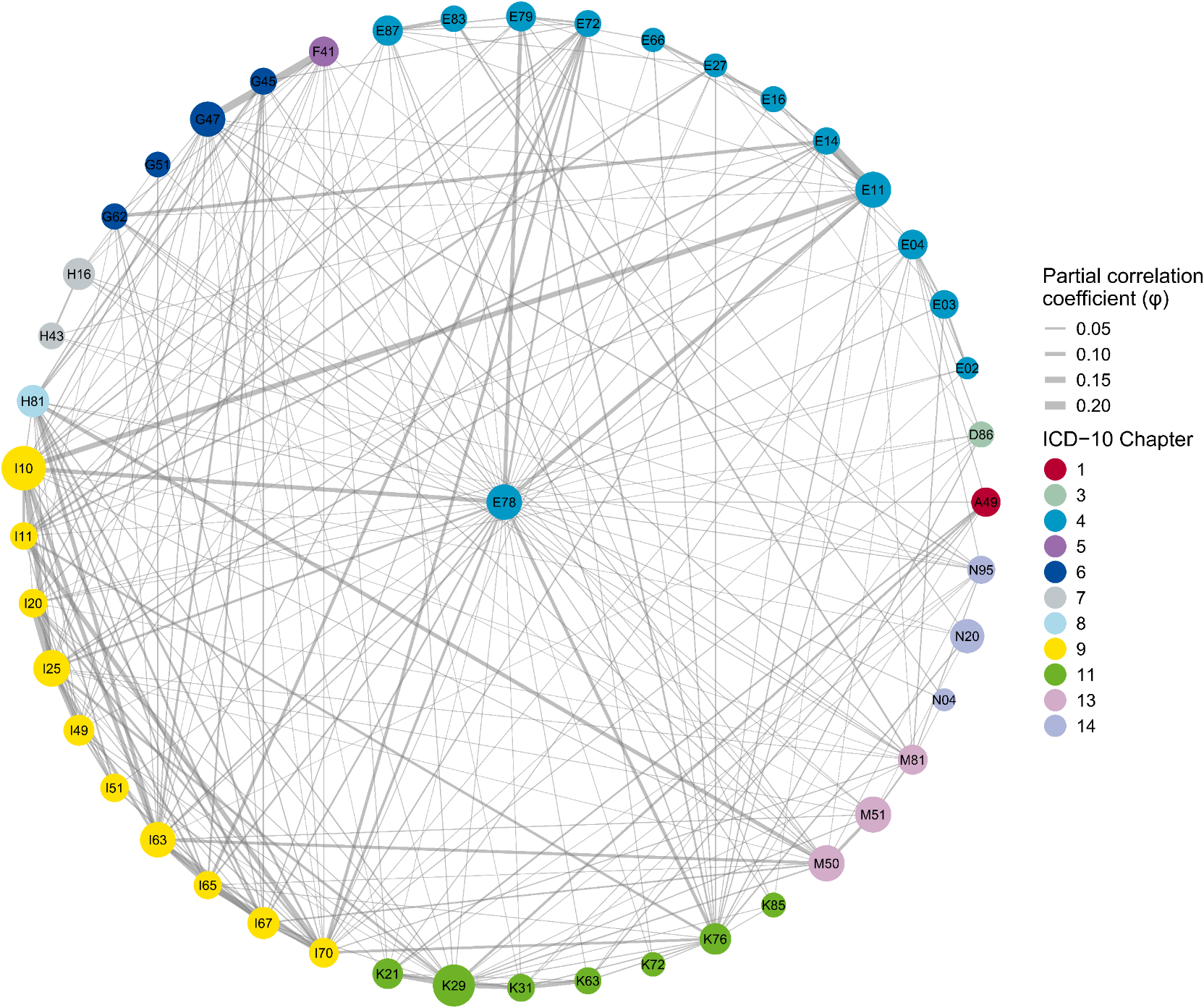

S7P

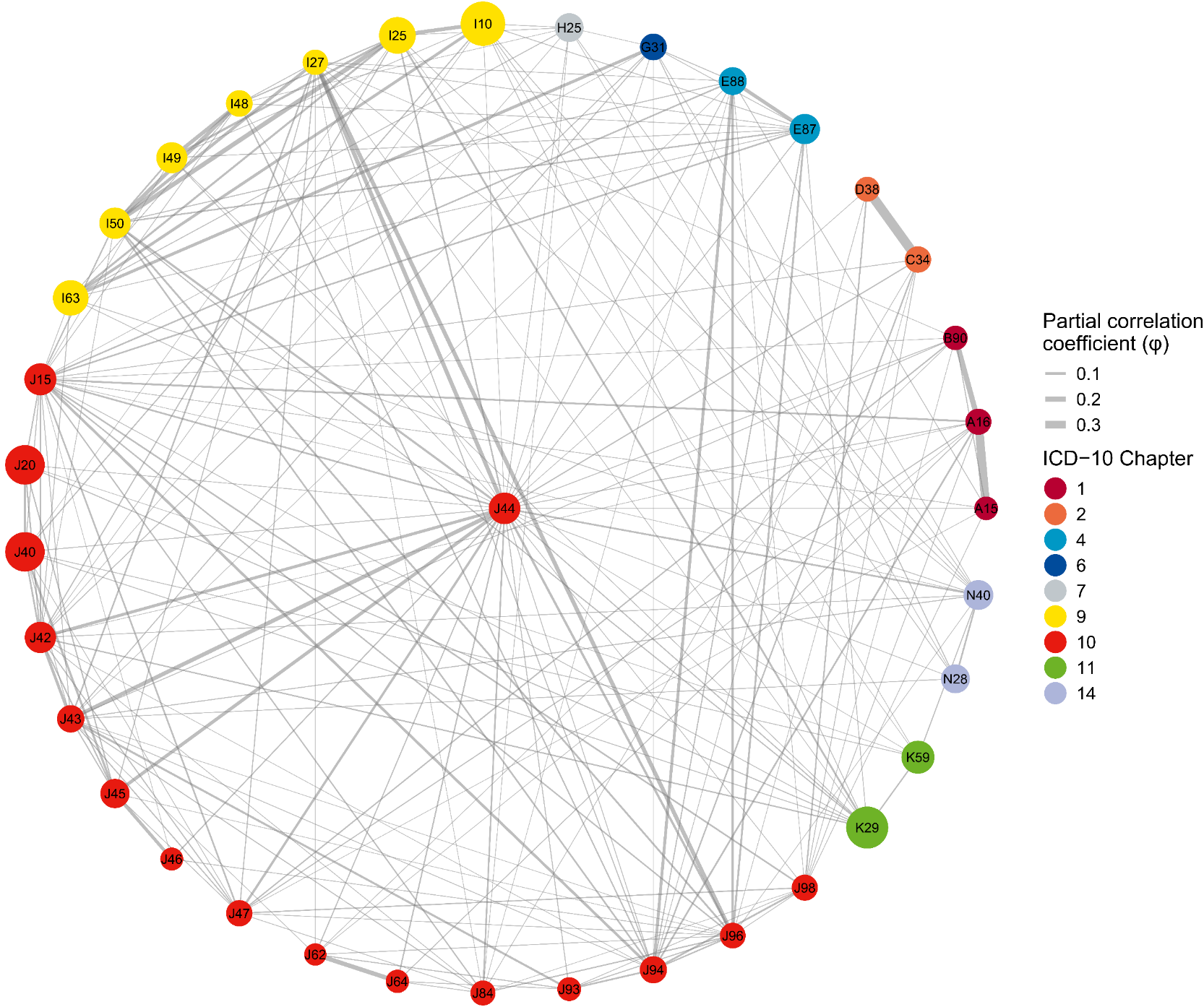

S7Q

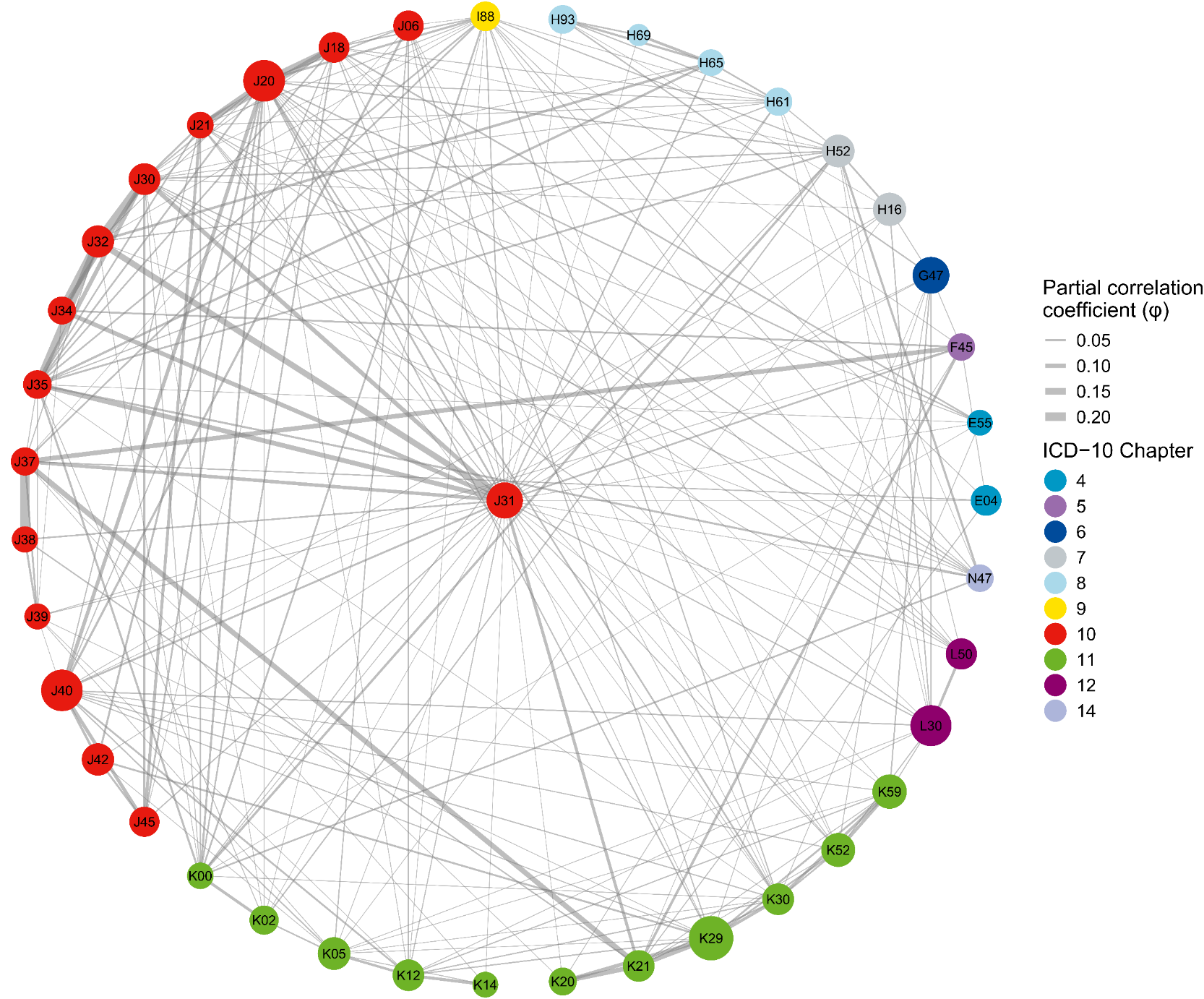

S7R

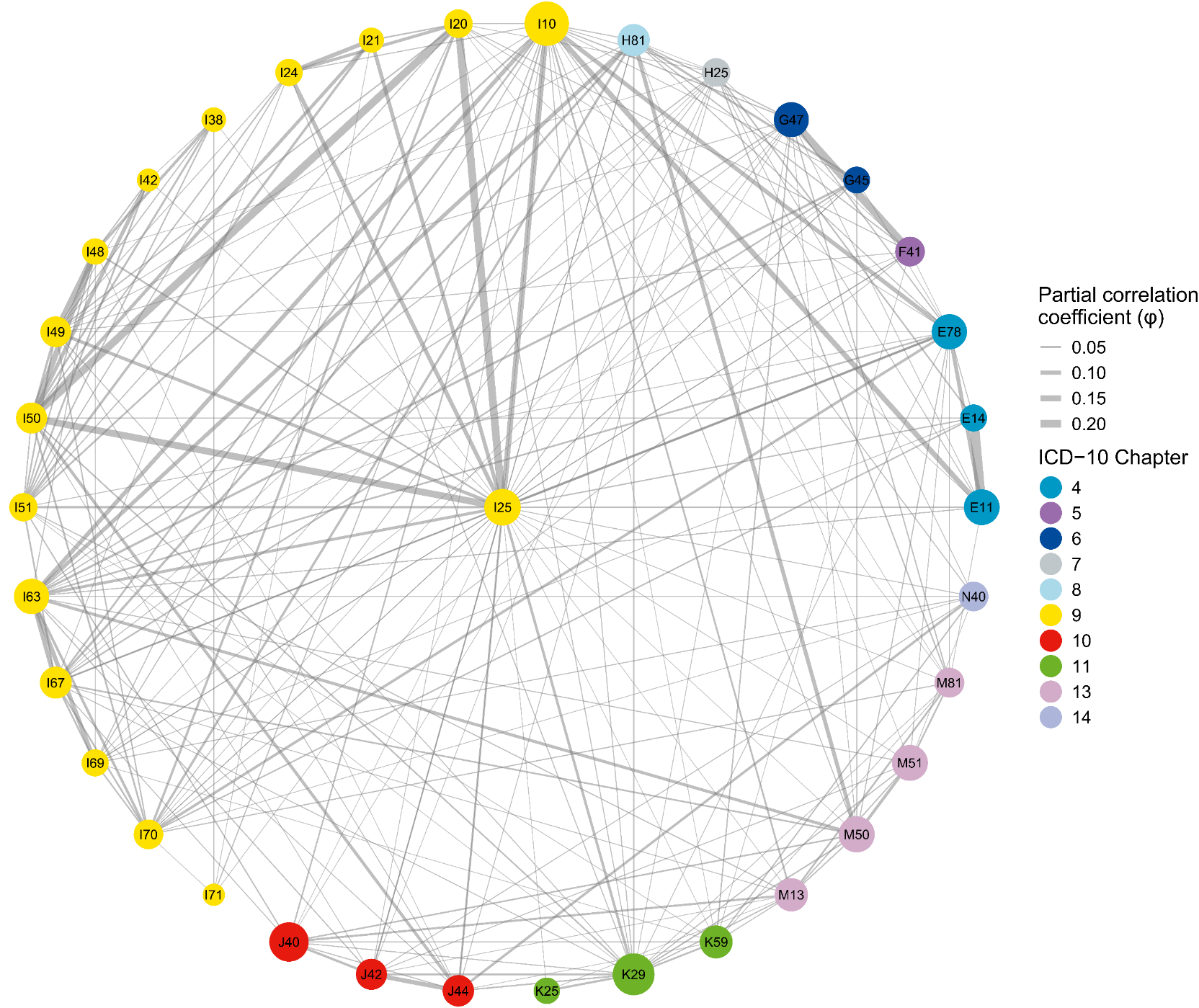

S7S

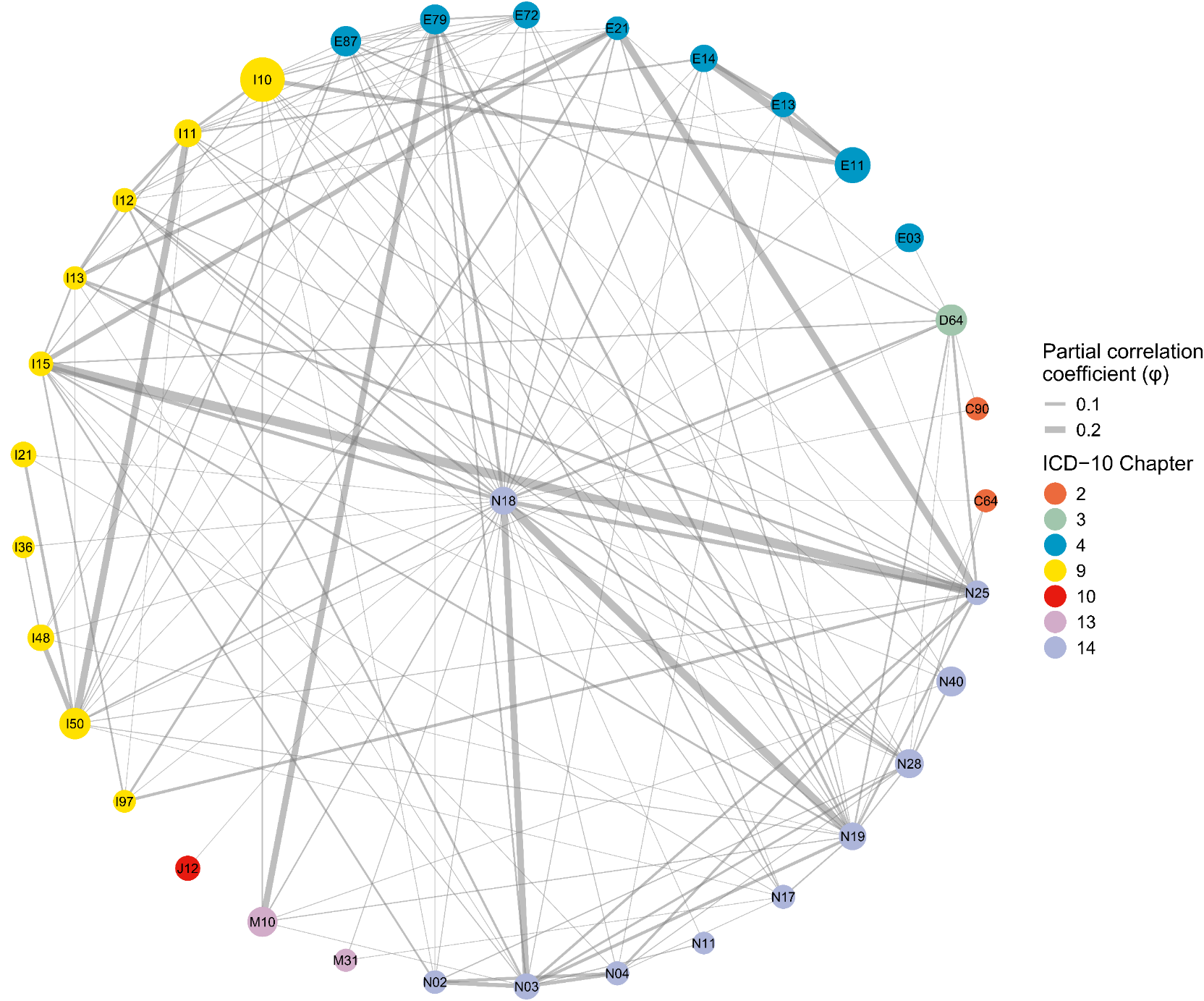

S7T

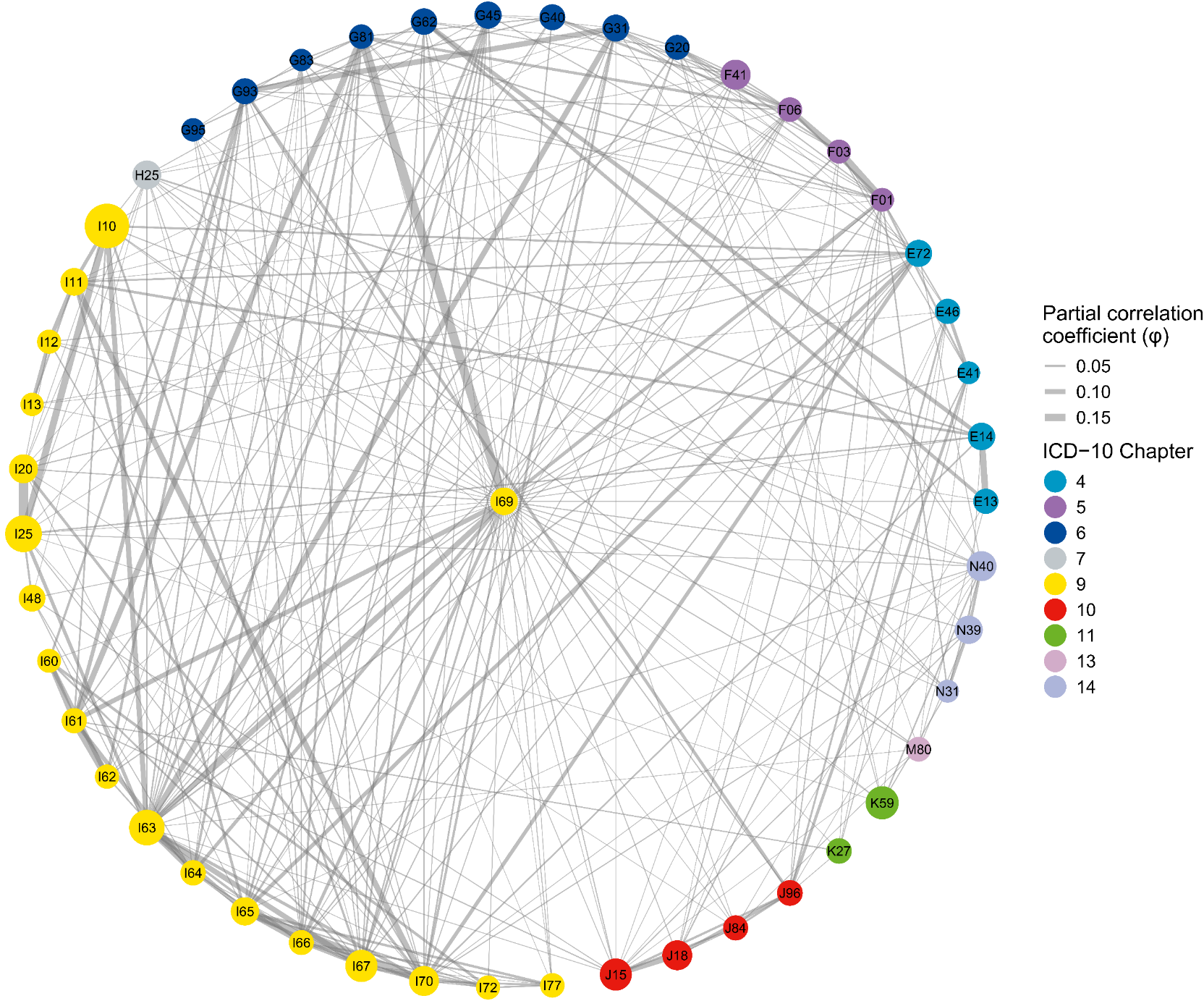

Supplementary Figure S8. Multimorbidity subnetworks for the top 20 hub diseases in female population identified by Multimorbidity Coefficient (MMC).

(A-T) Each panel displays one of the top 20 hub diseases (center node) and its neighboring conditions using a star layout. Nodes represent individual ICD‑10 conditions (labeled with ICD-3 codes); node size is proportional to each condition’s period prevalence (2016–2023), and node color denotes the ICD‑10 chapter. Edges depict significant adjusted associations between pairs of conditions after controlling for all other diseases in the network (quantified by the partial correlation coefficient, φ). Only associations meeting FDR‑adjusted *P* < 0.05 and φ > 0.01 are shown. Edge width is scaled to the magnitude of φ. Hubs are presented in descending order of MMC: (A) [K29], (B) [E88], (C) [I50], (D) [E87], (E) [M81], (F) [L30], (G) [I70], (H) [D64], (I) [I63], (J) [G47], (K) [J15], (L) [K76], (M) [D61], (N) [I10], (O) [E78], (P) [H16], (Q) [J31], (R) [N18], (S) [F41], (T) [I25]. These subnetworks can be explored interactively at: https://pumc-multimorbidity.shinyapps.io/hub-disease-network/

S8A

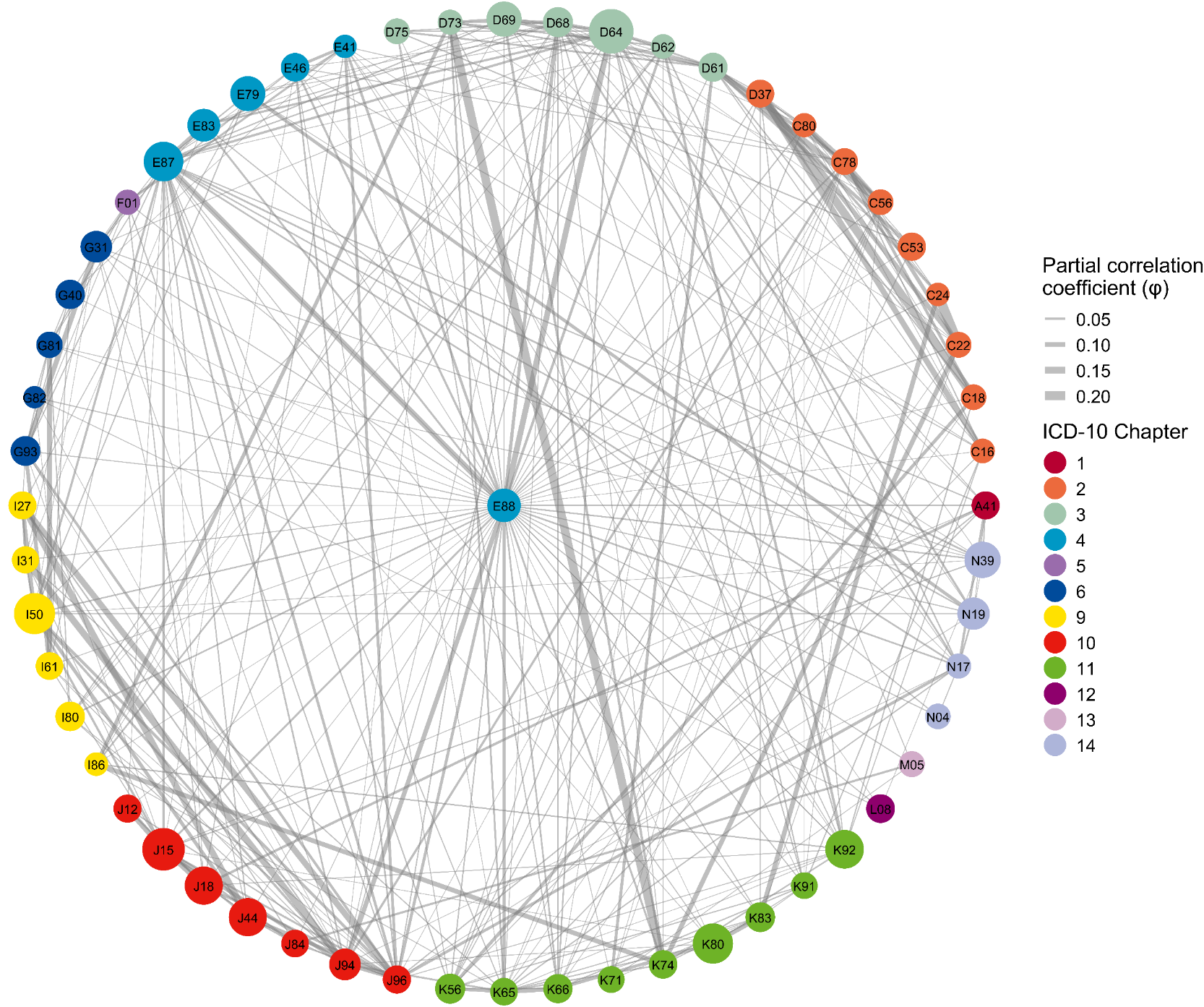

S8B

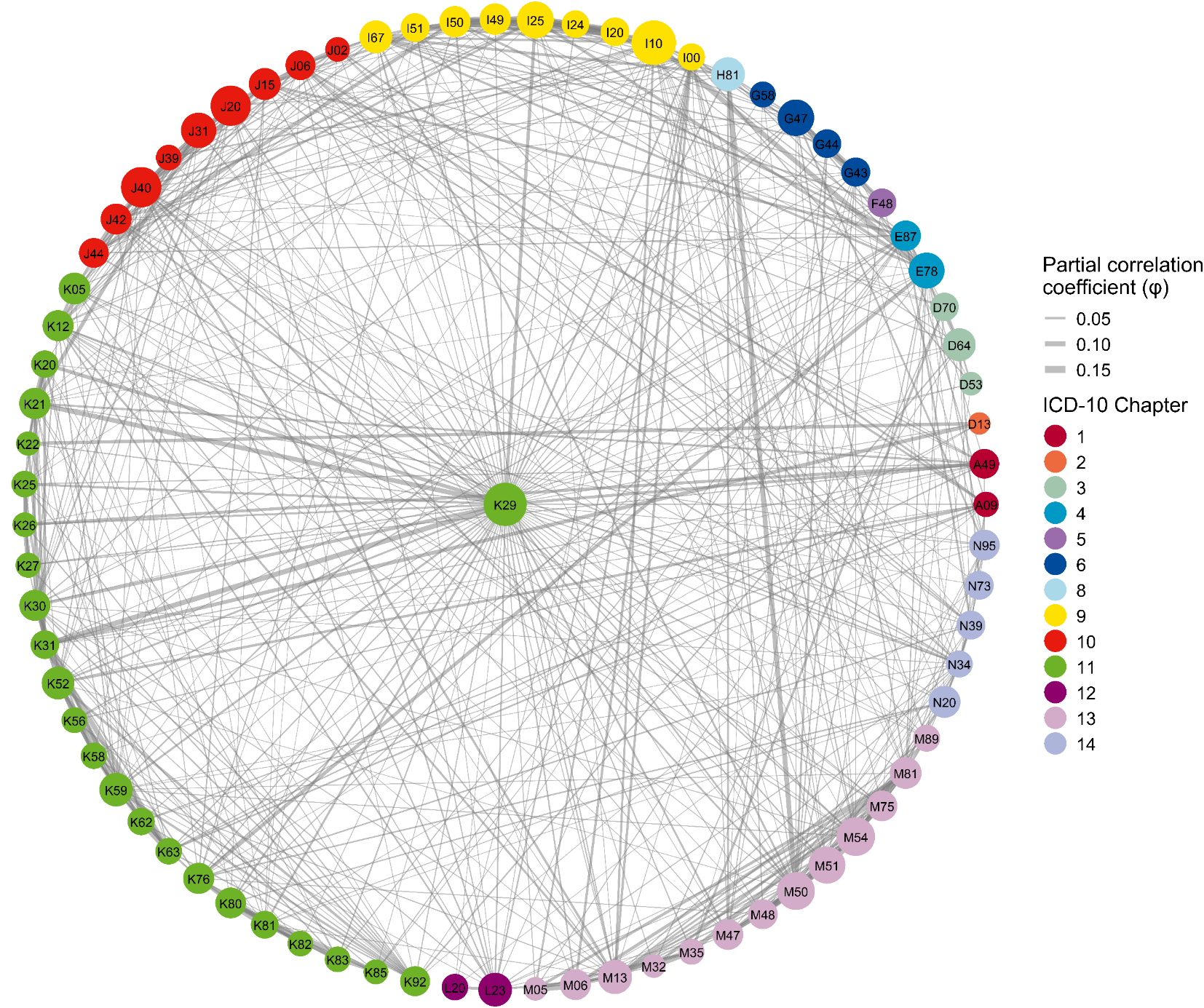

S8C

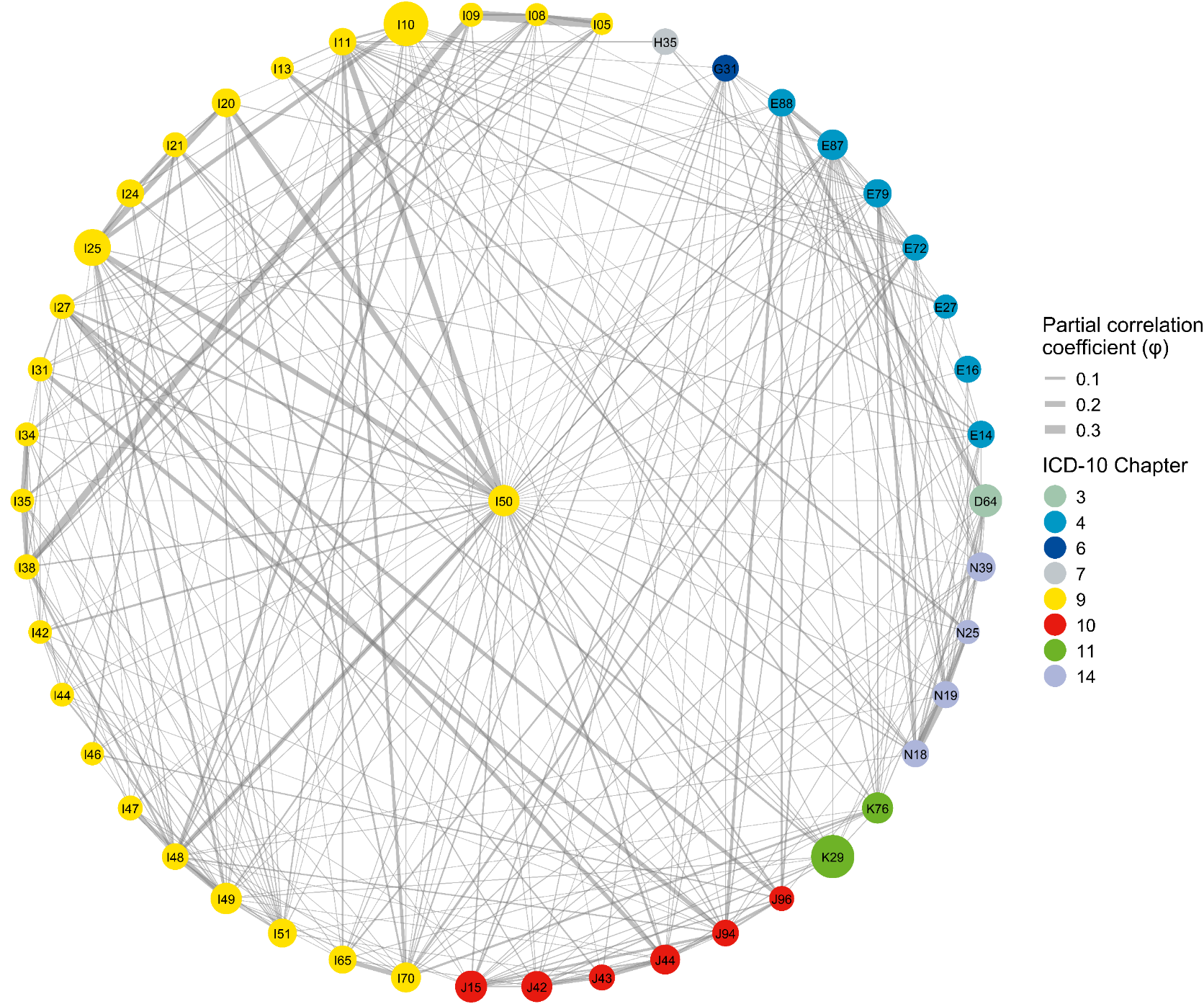

S8D

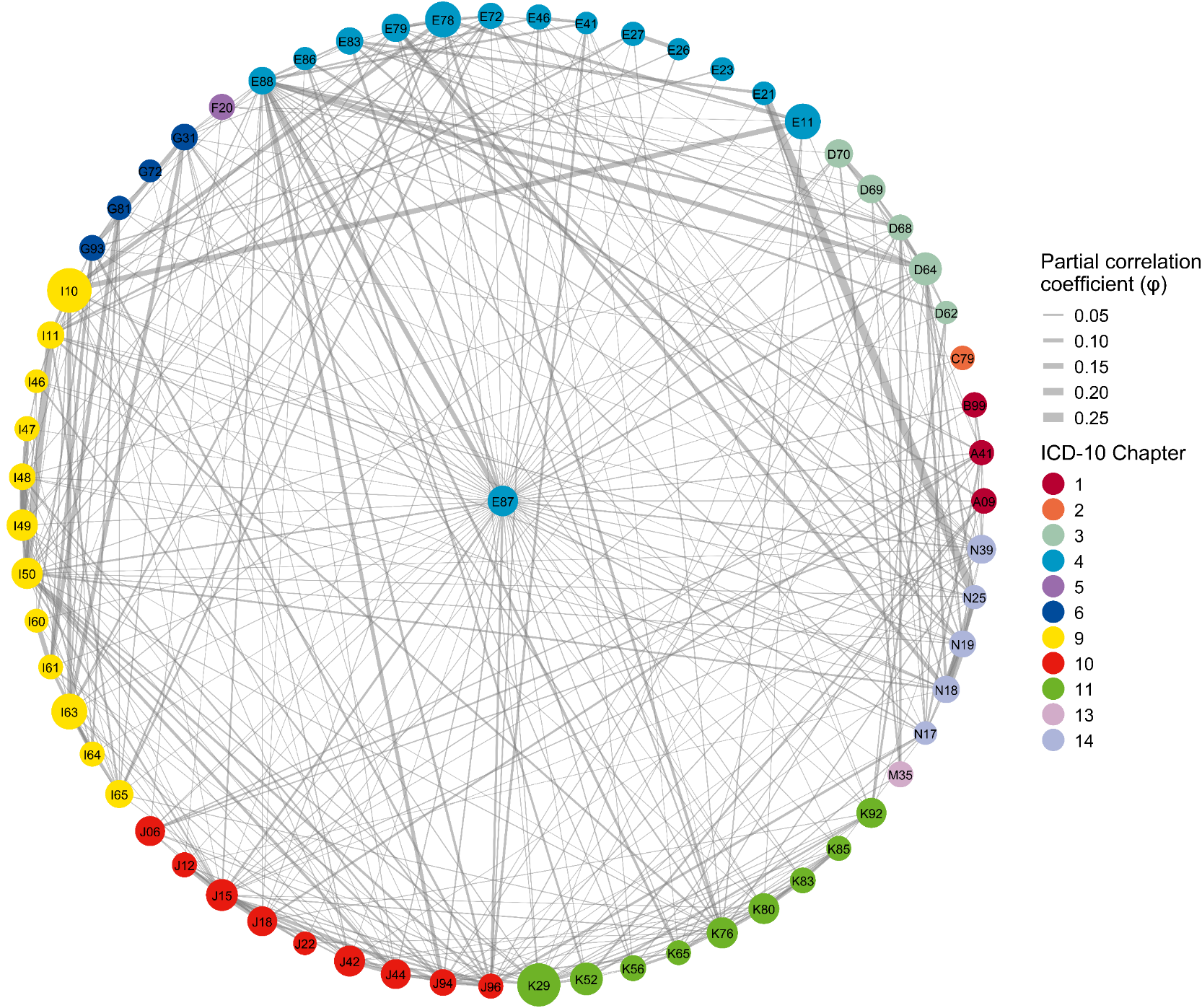

S8E

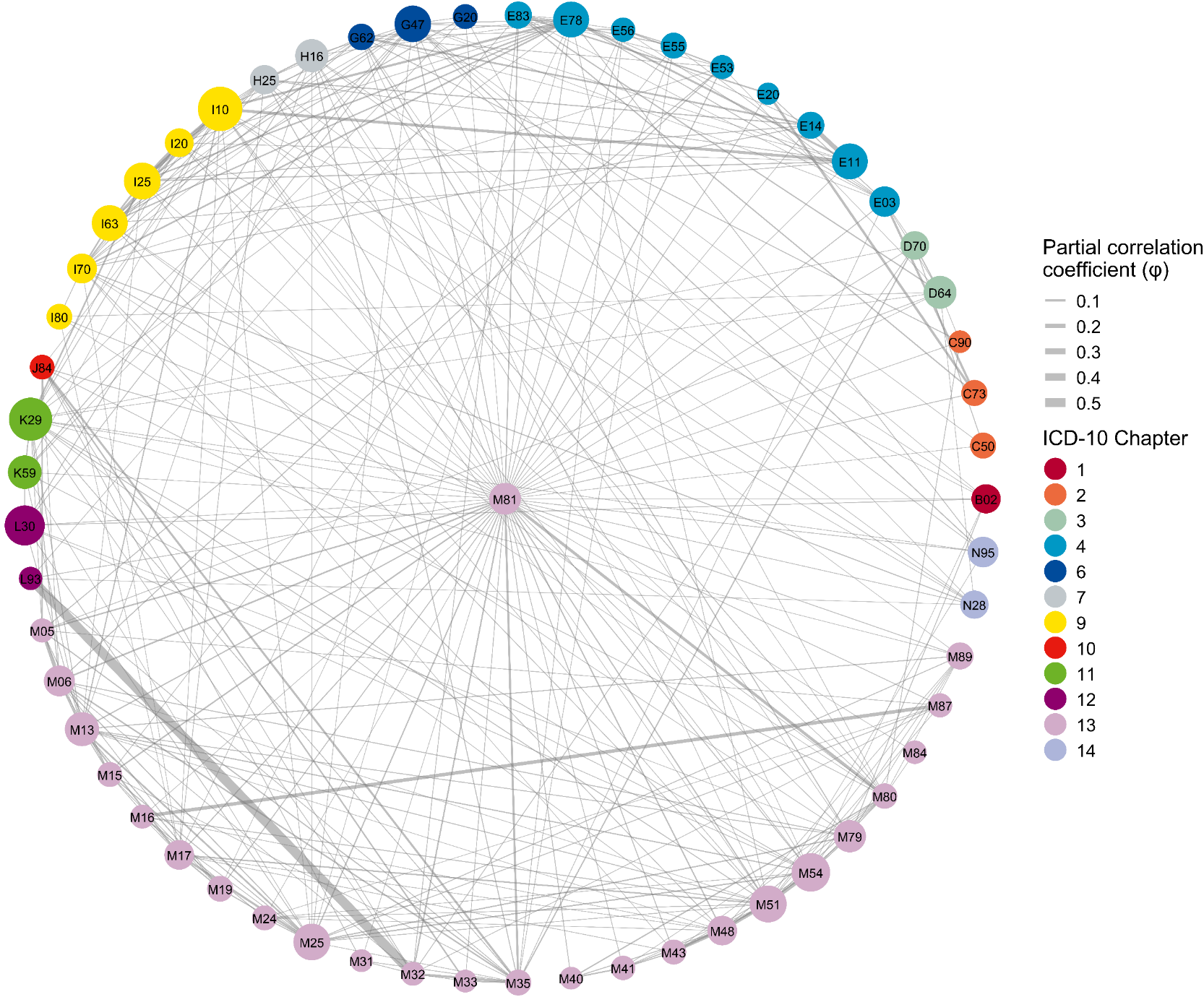

S8F

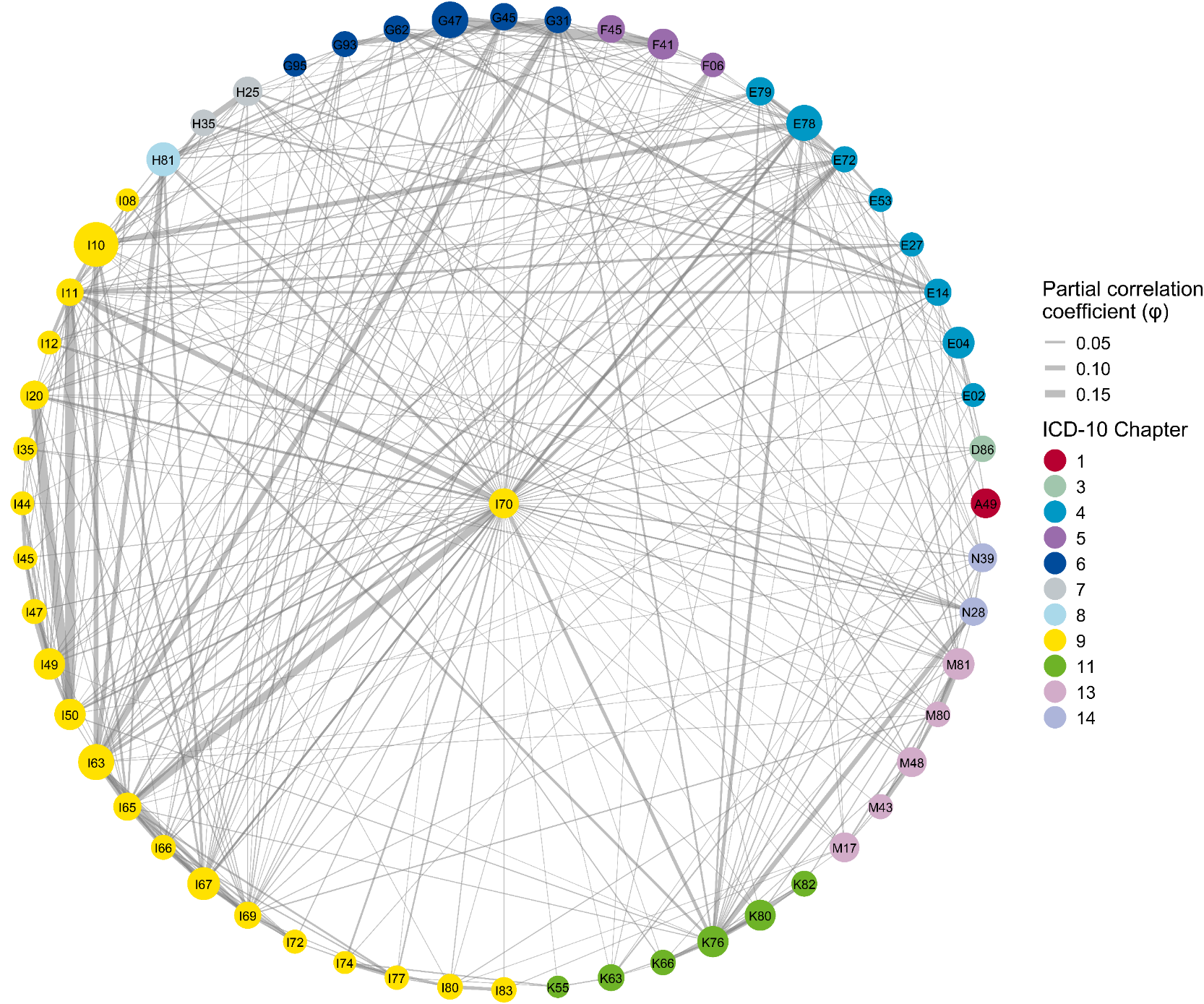

S8G

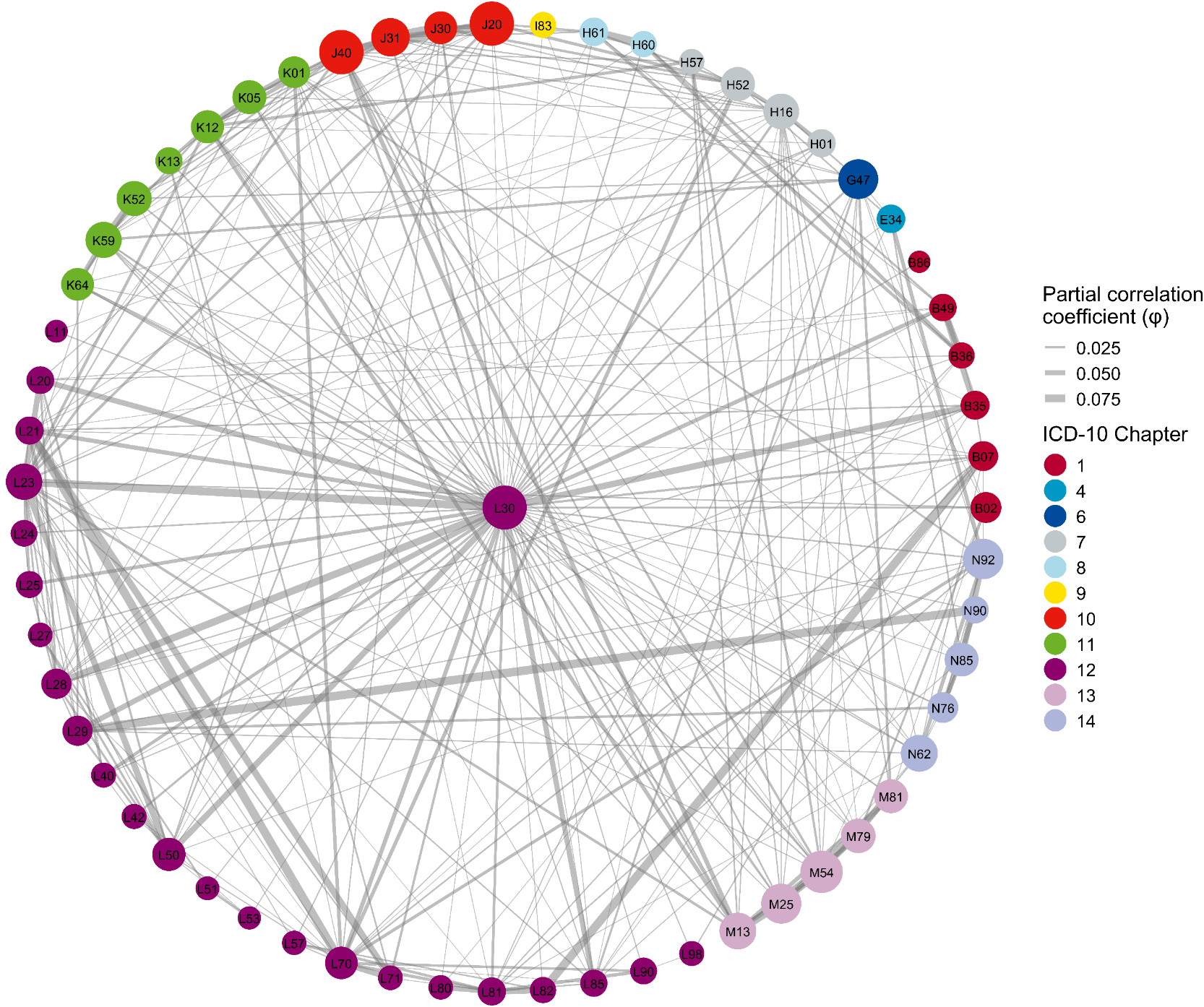

S8H

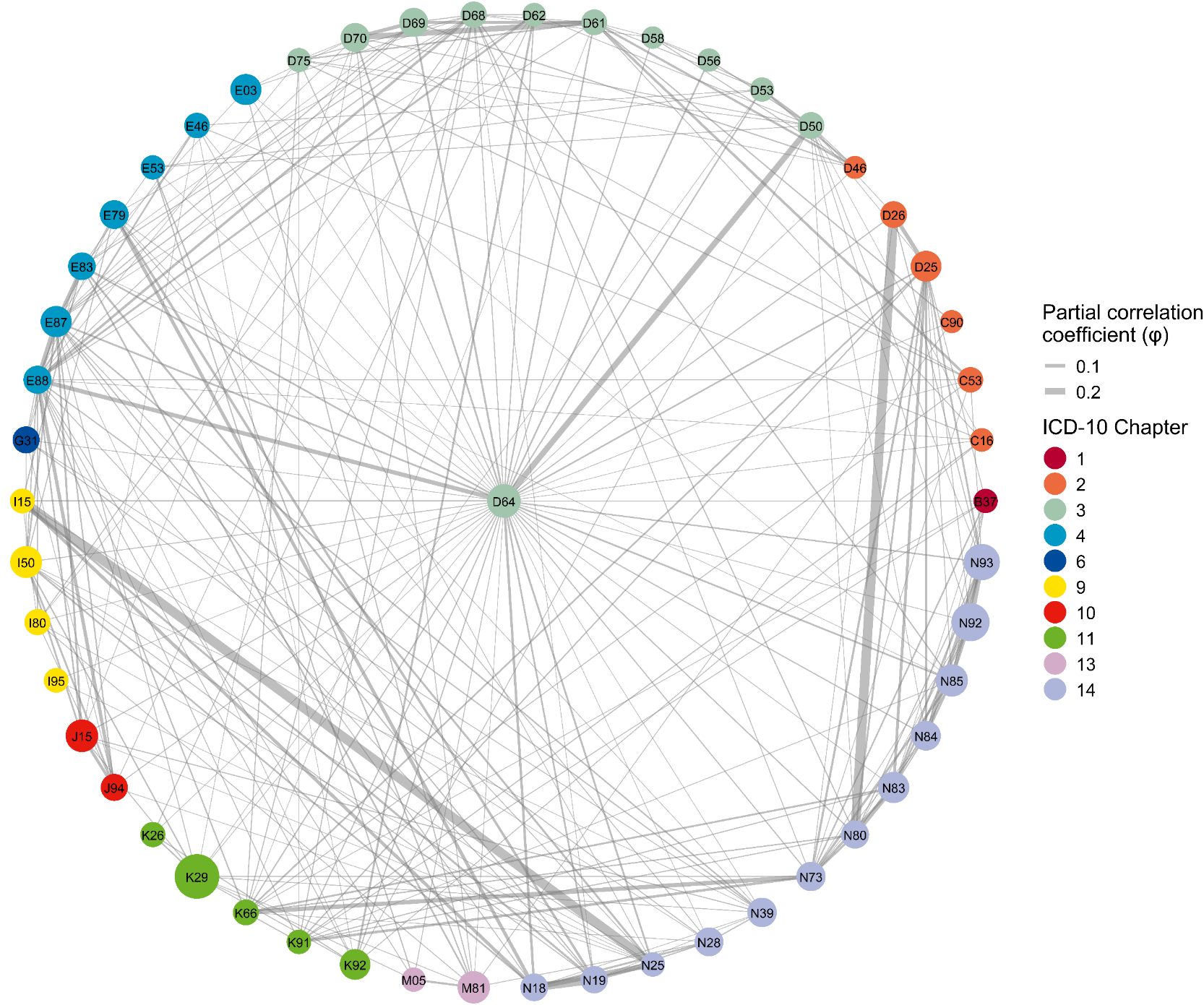

S8I

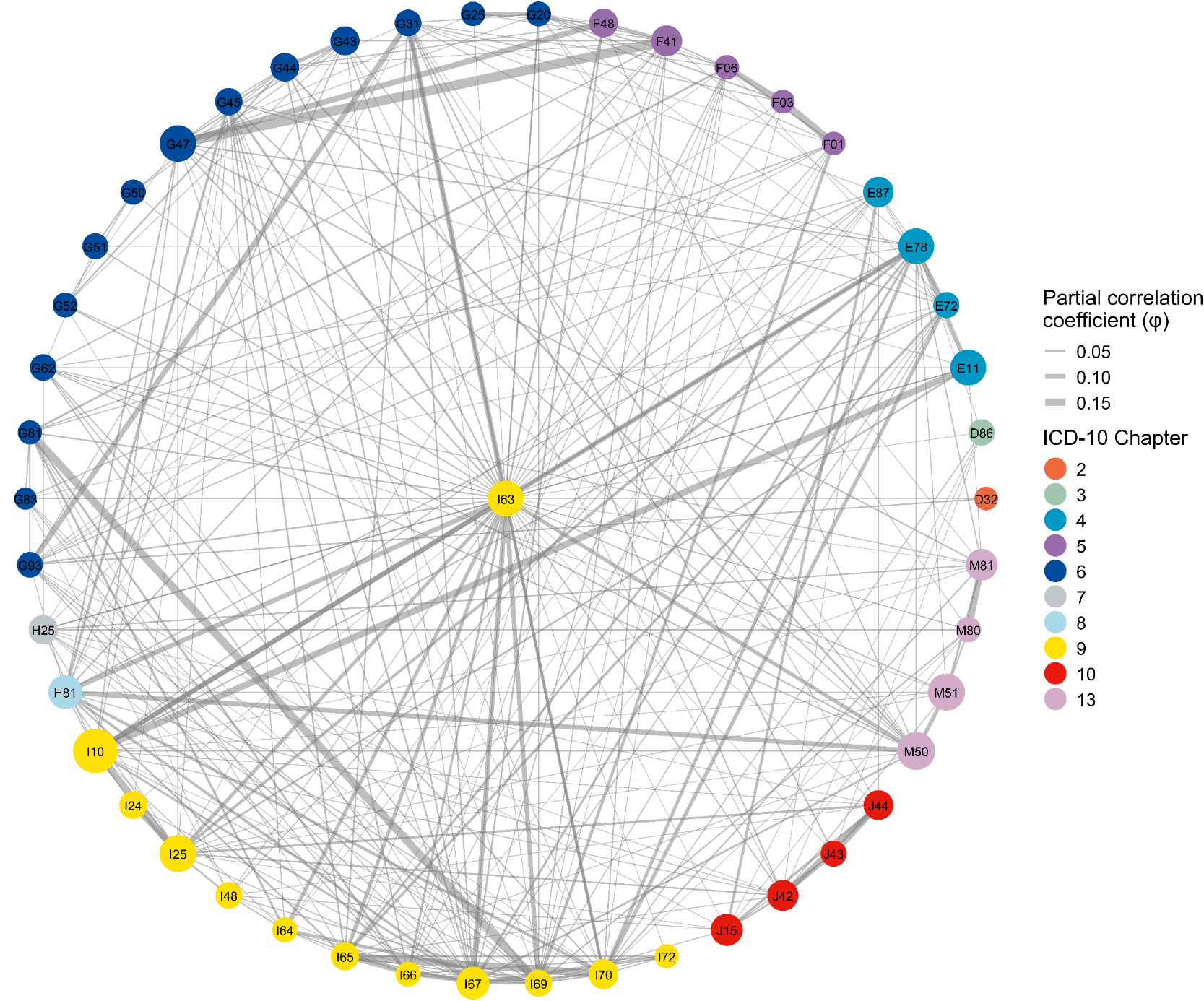

S8J

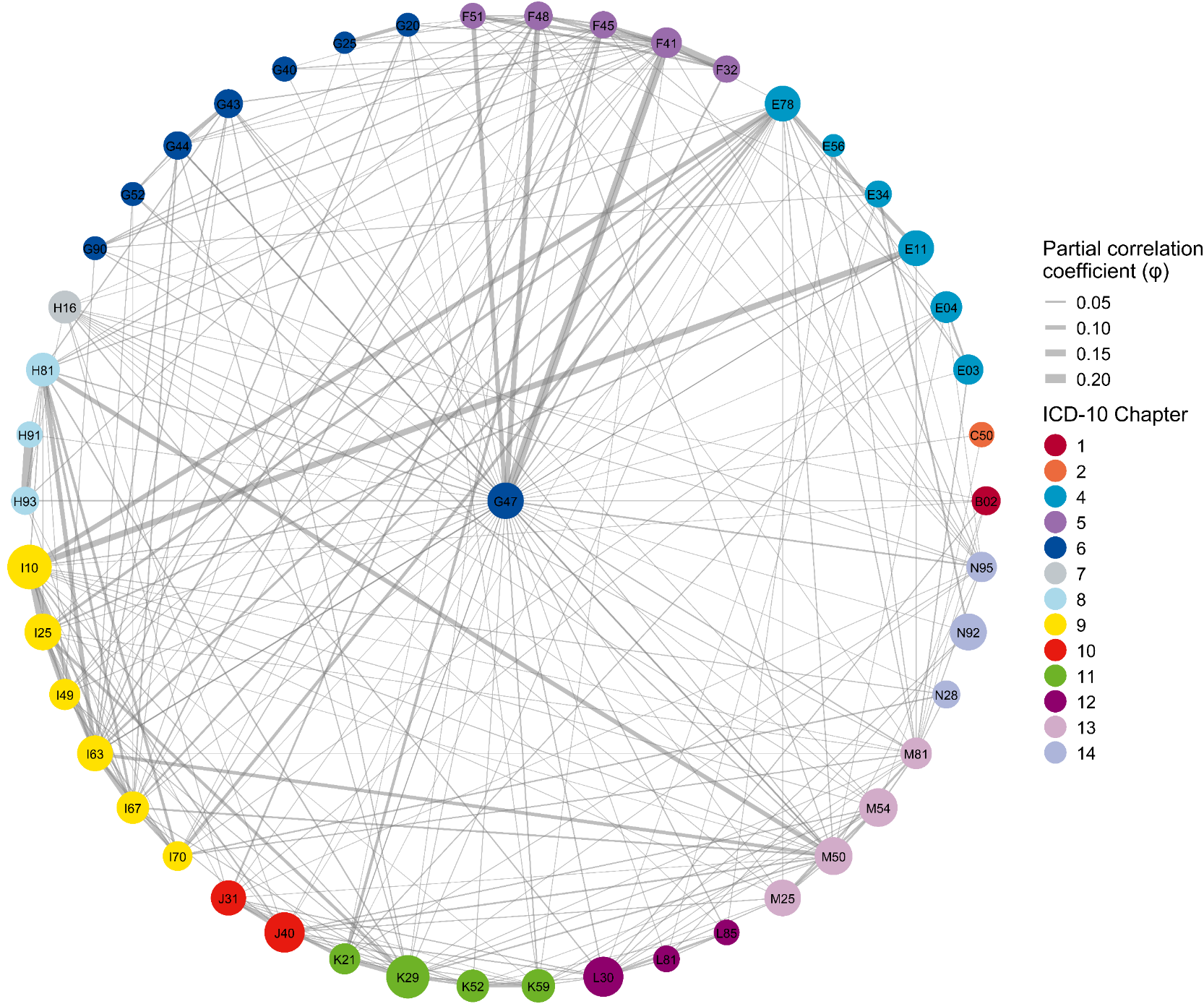

S8K

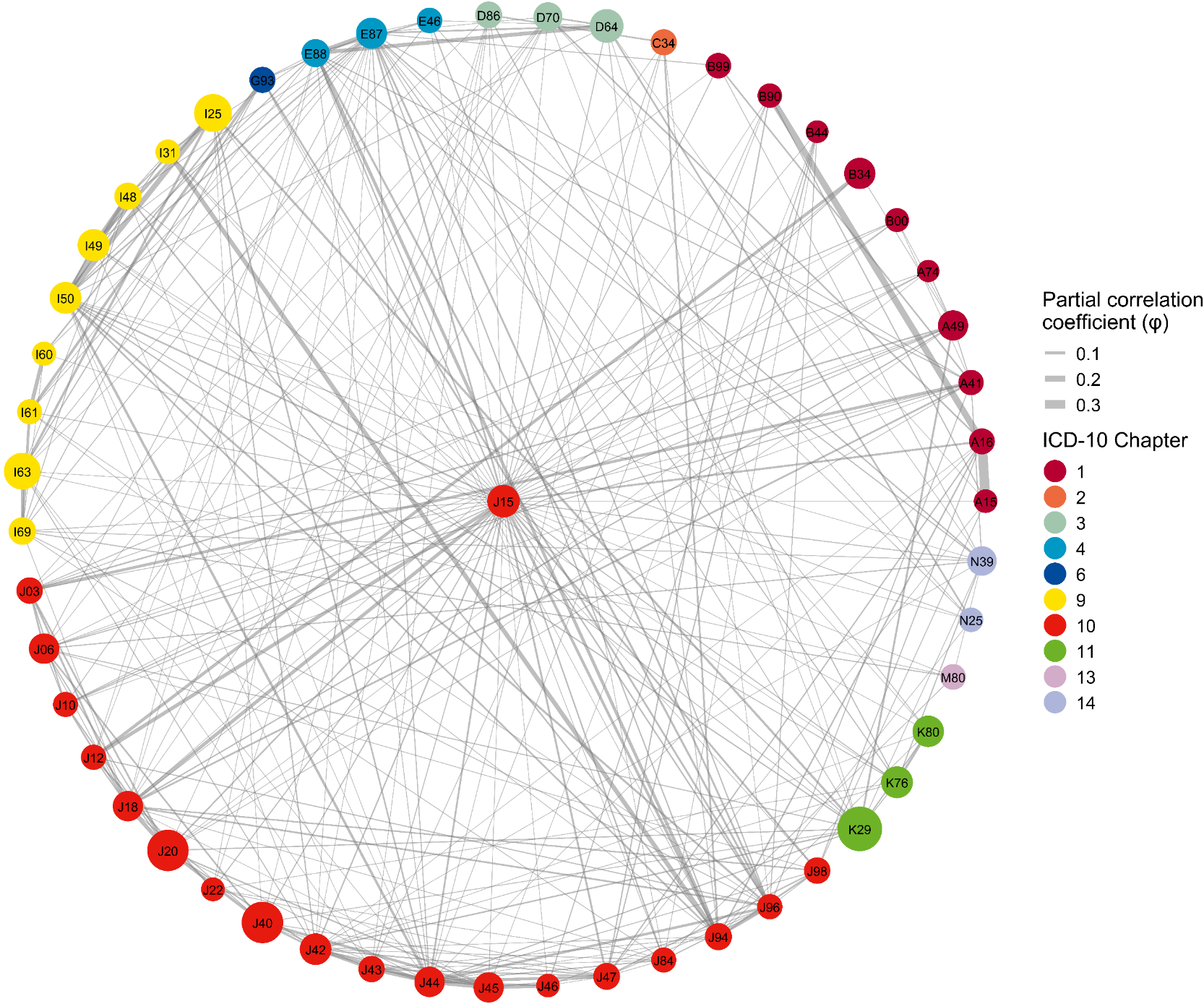

S8L

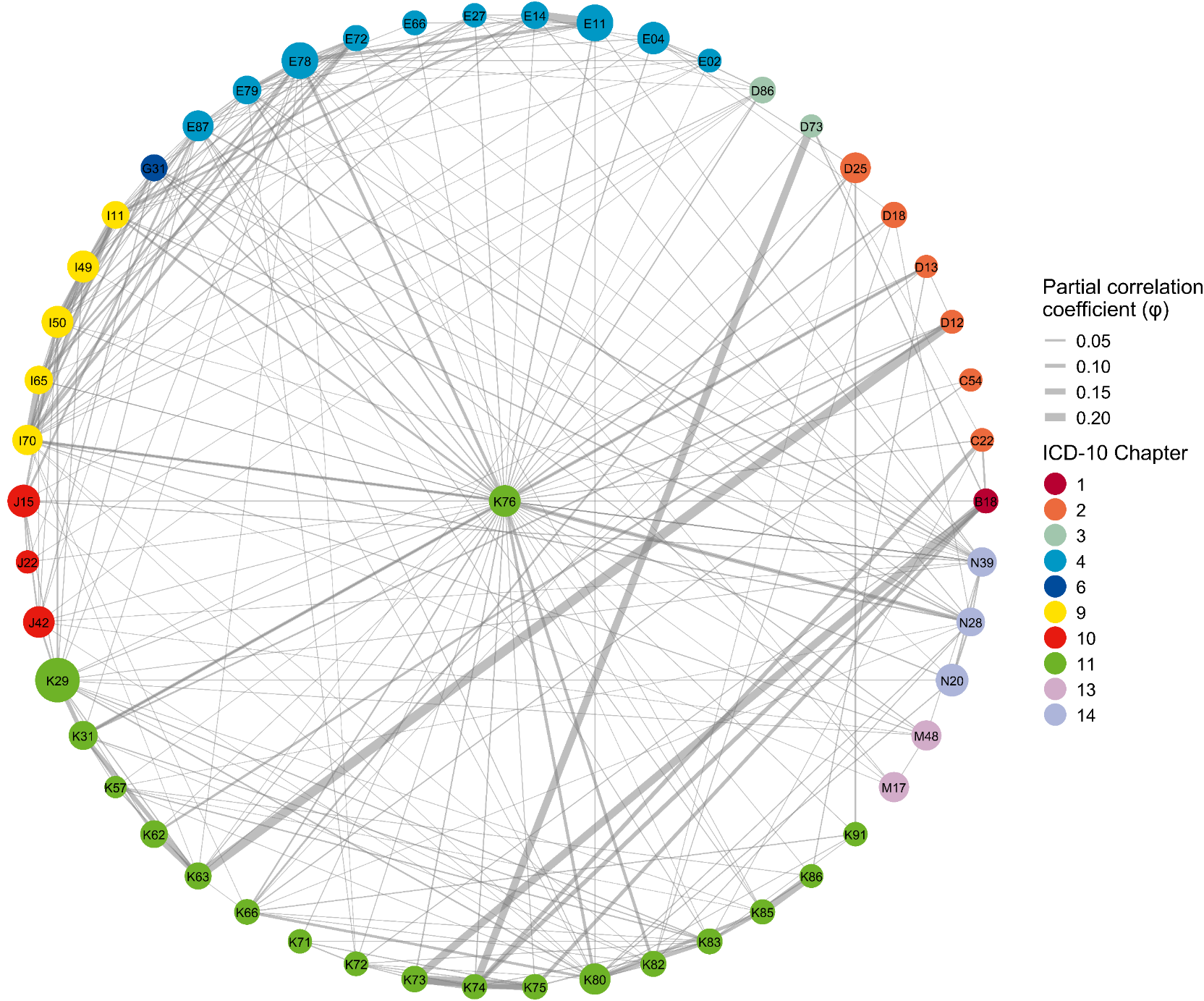

S8M

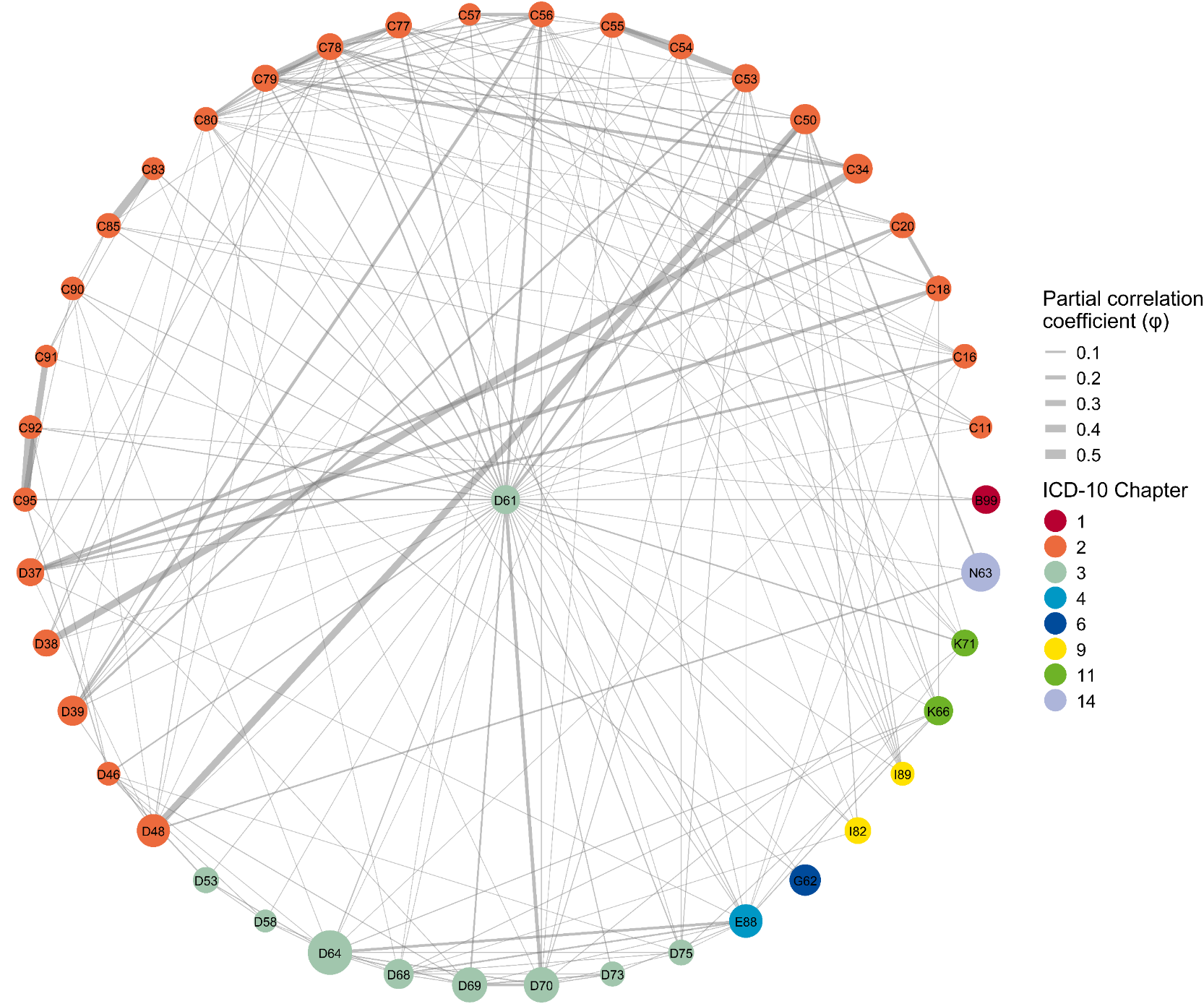

S8N

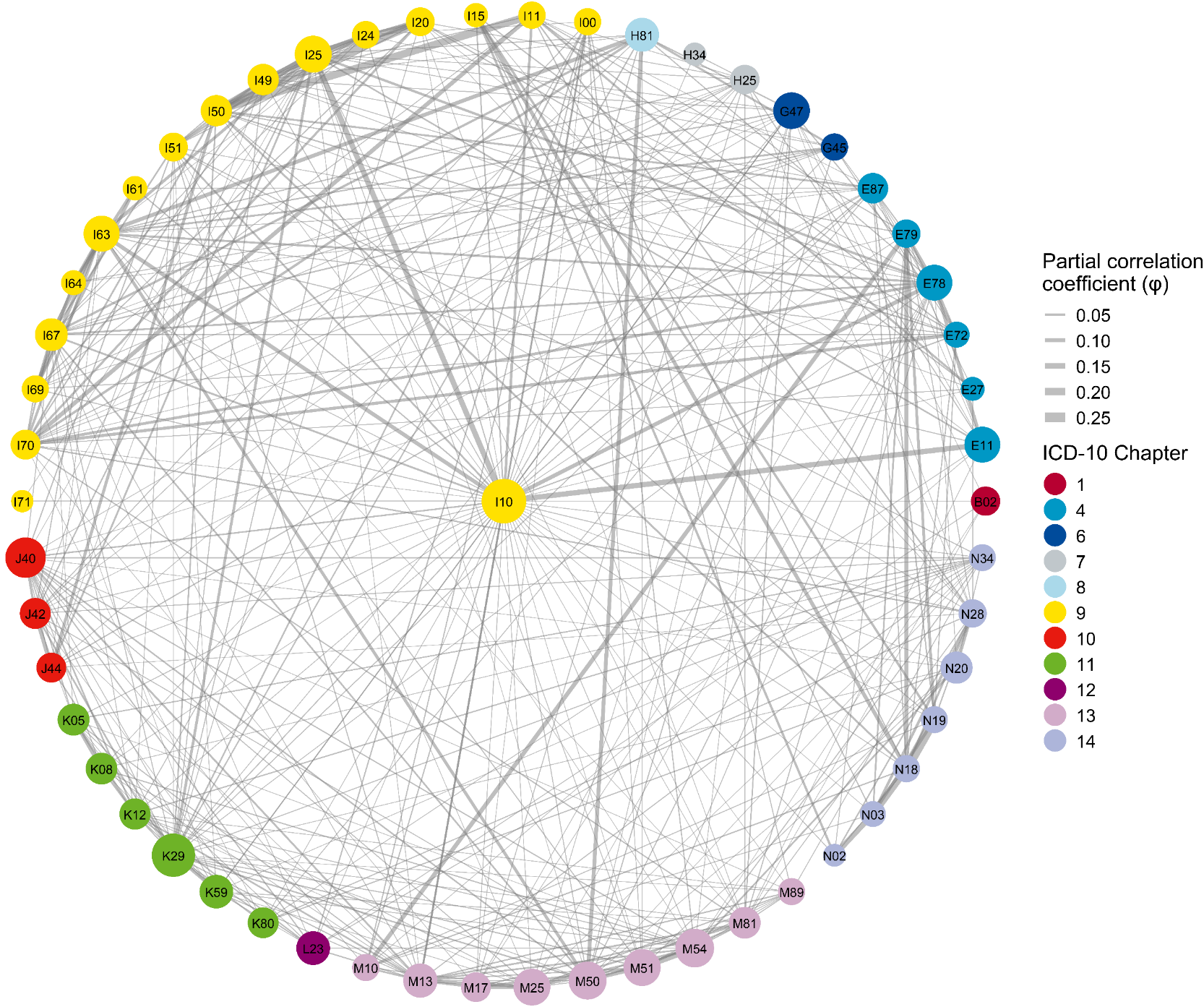

S8O

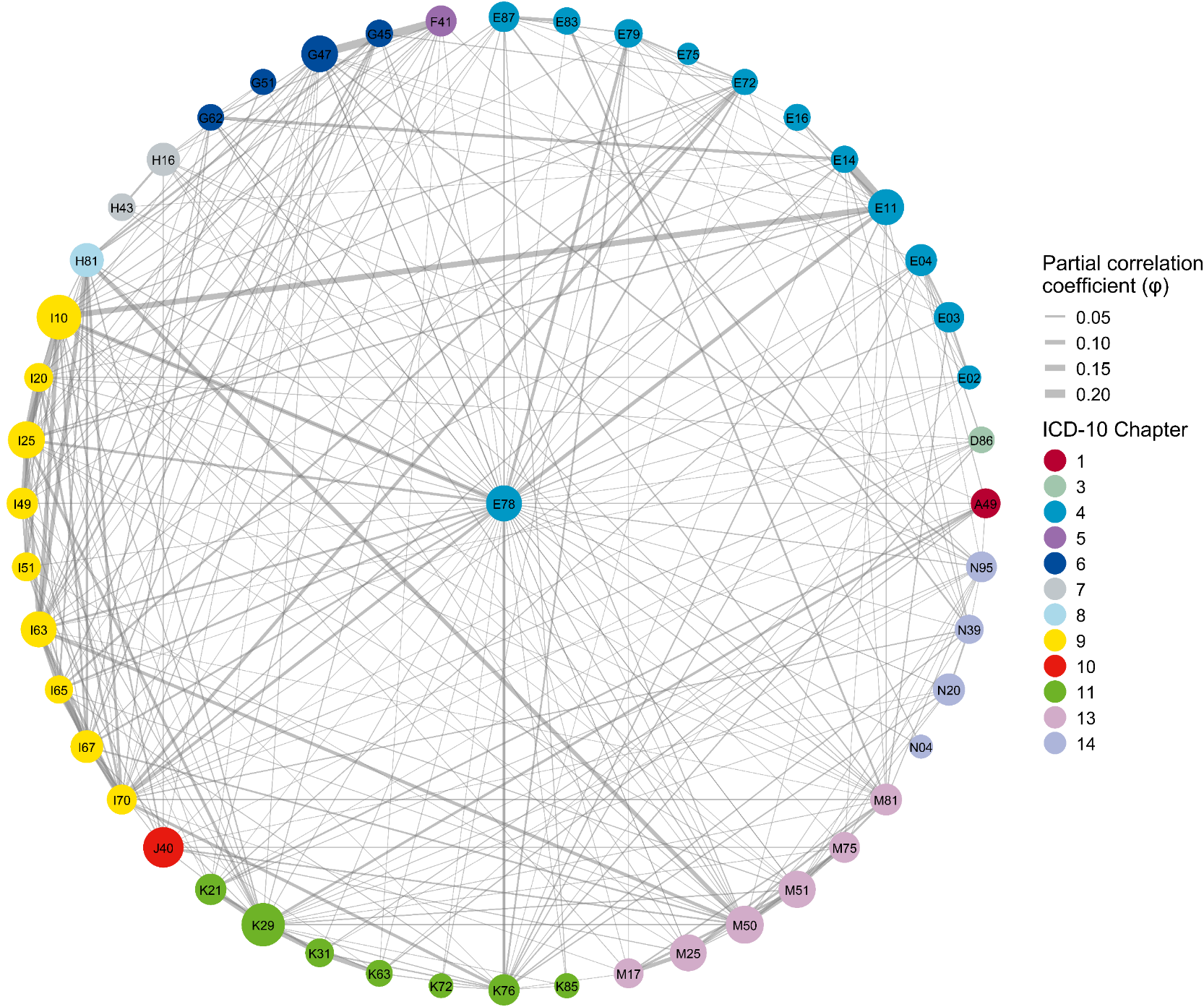

S8P

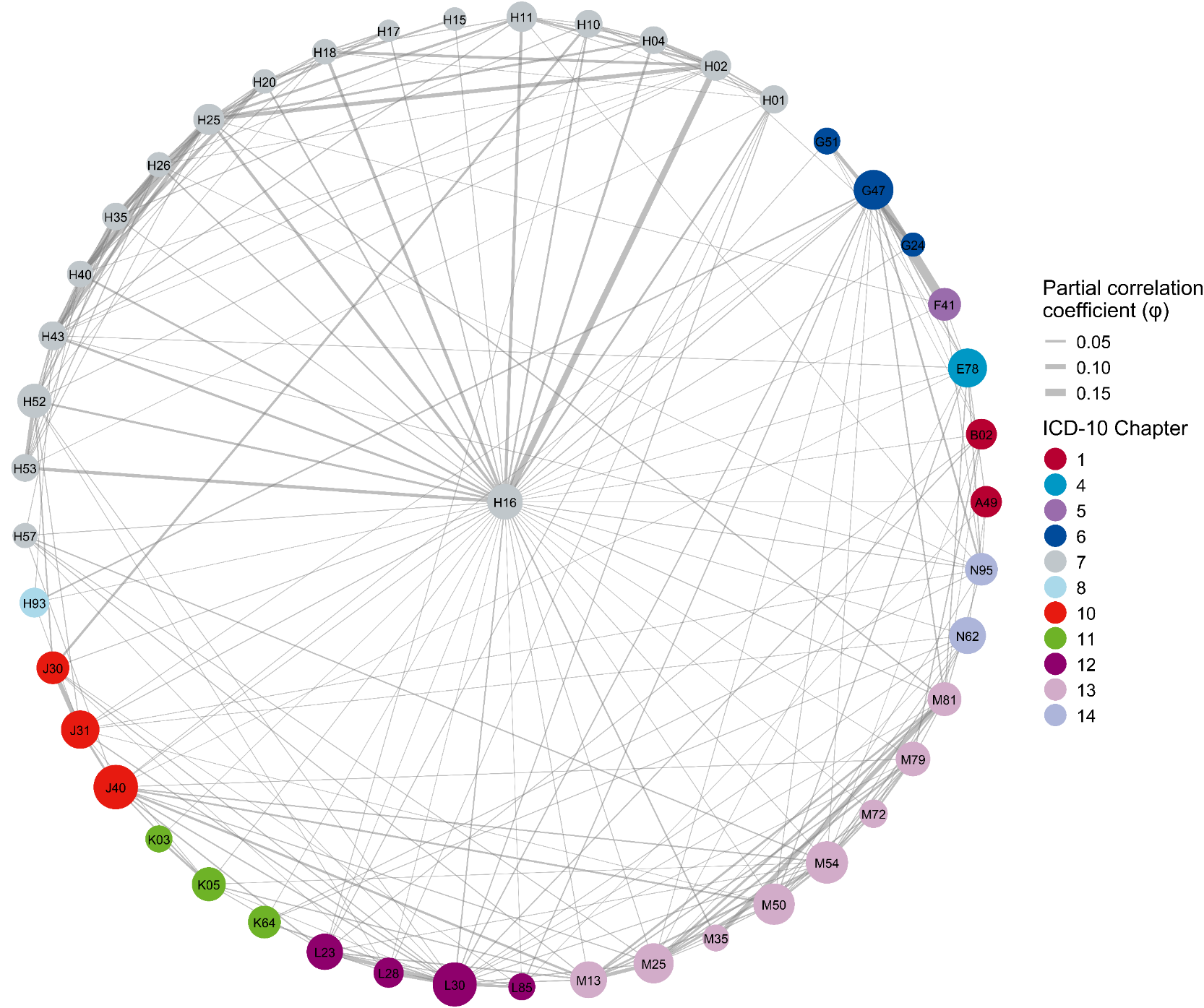

S8Q

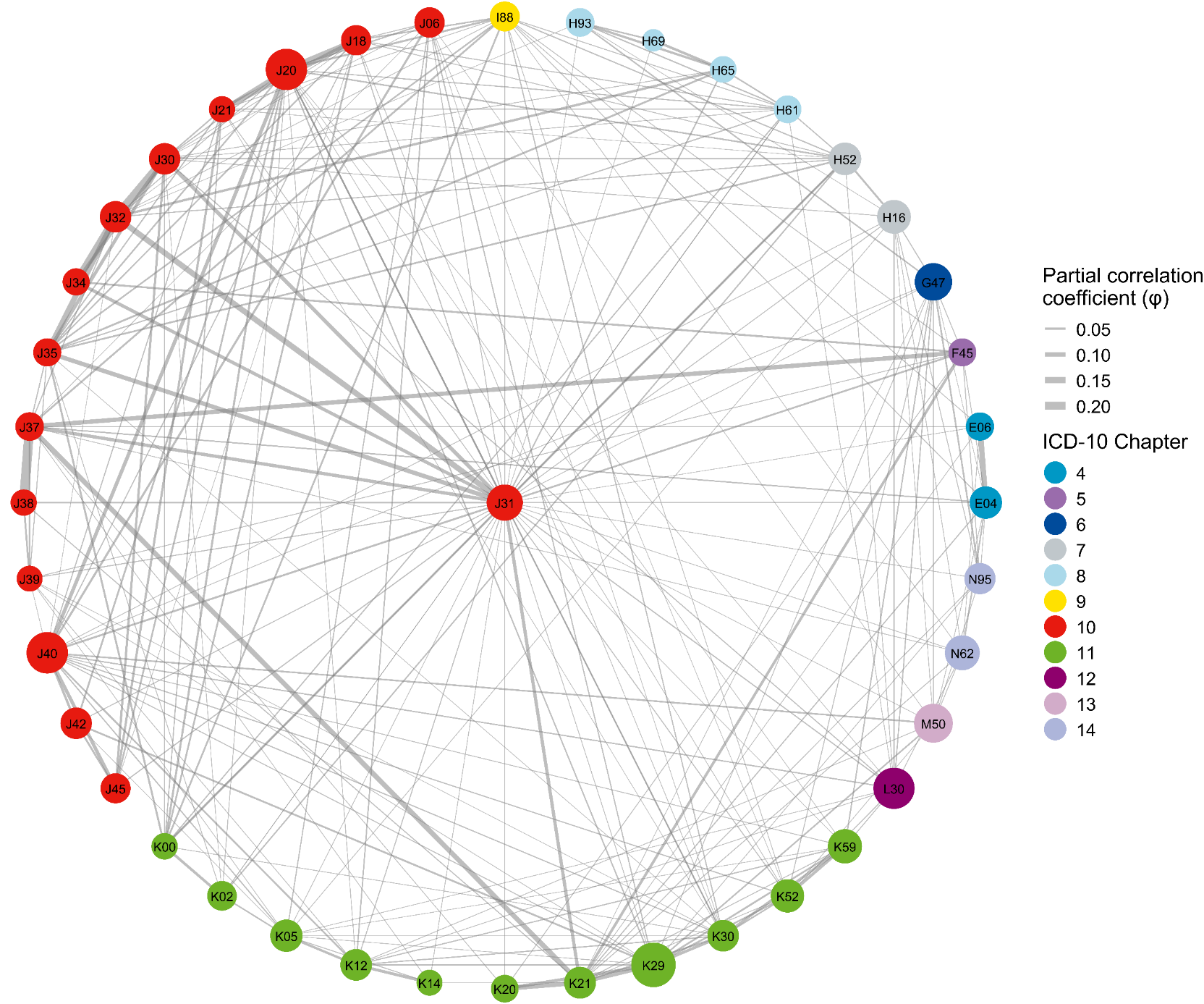

S8R

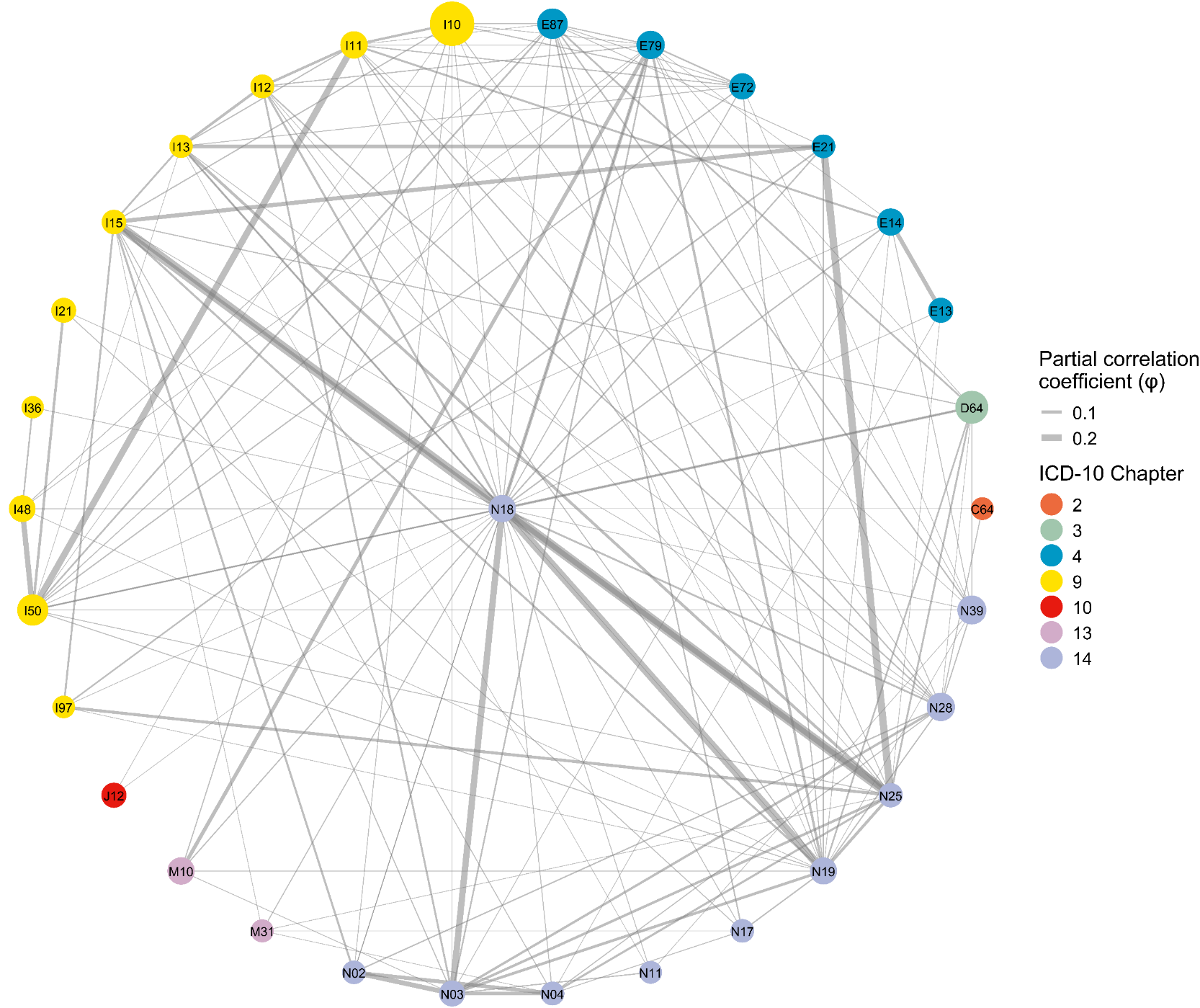

S8S

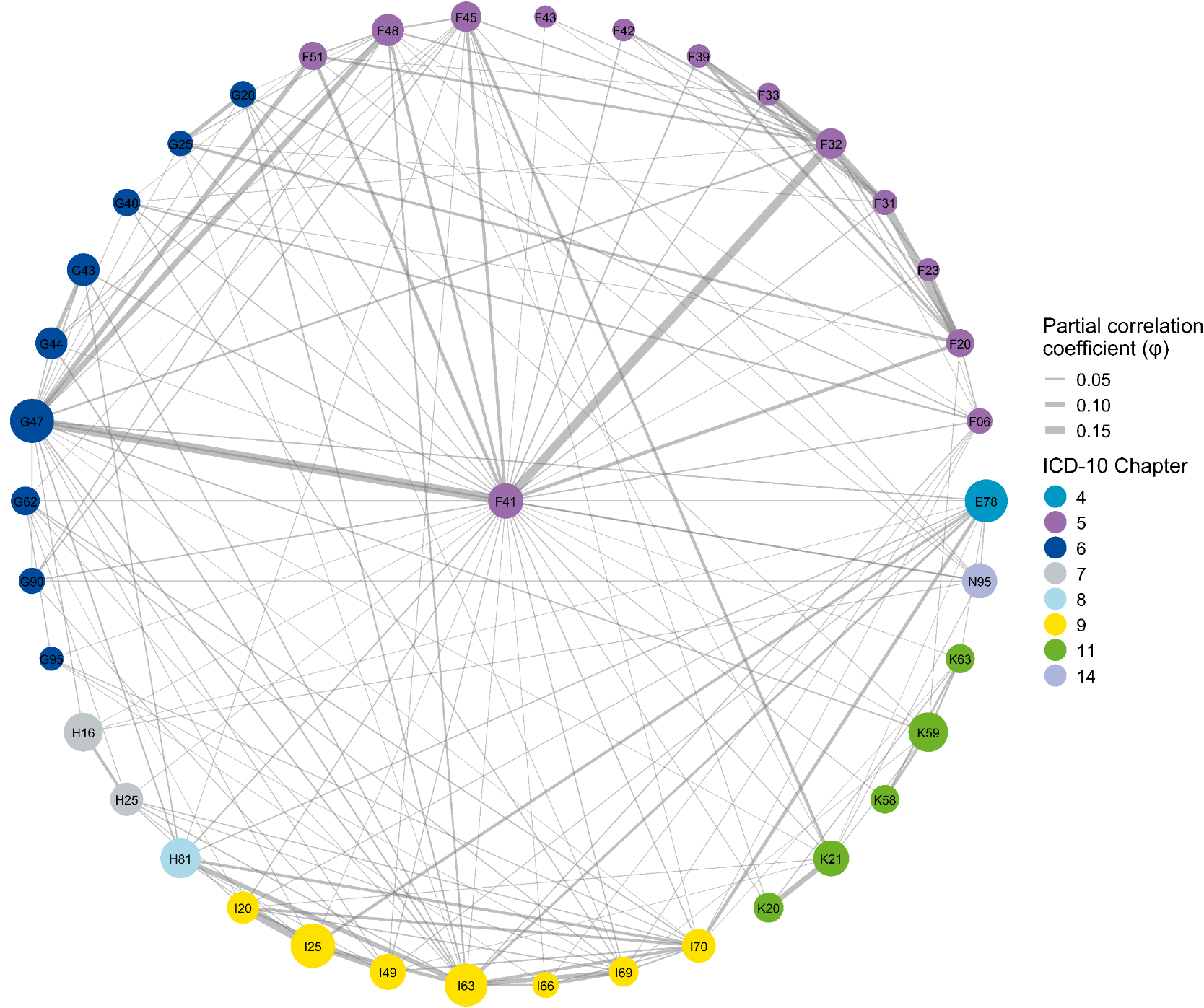

S8T

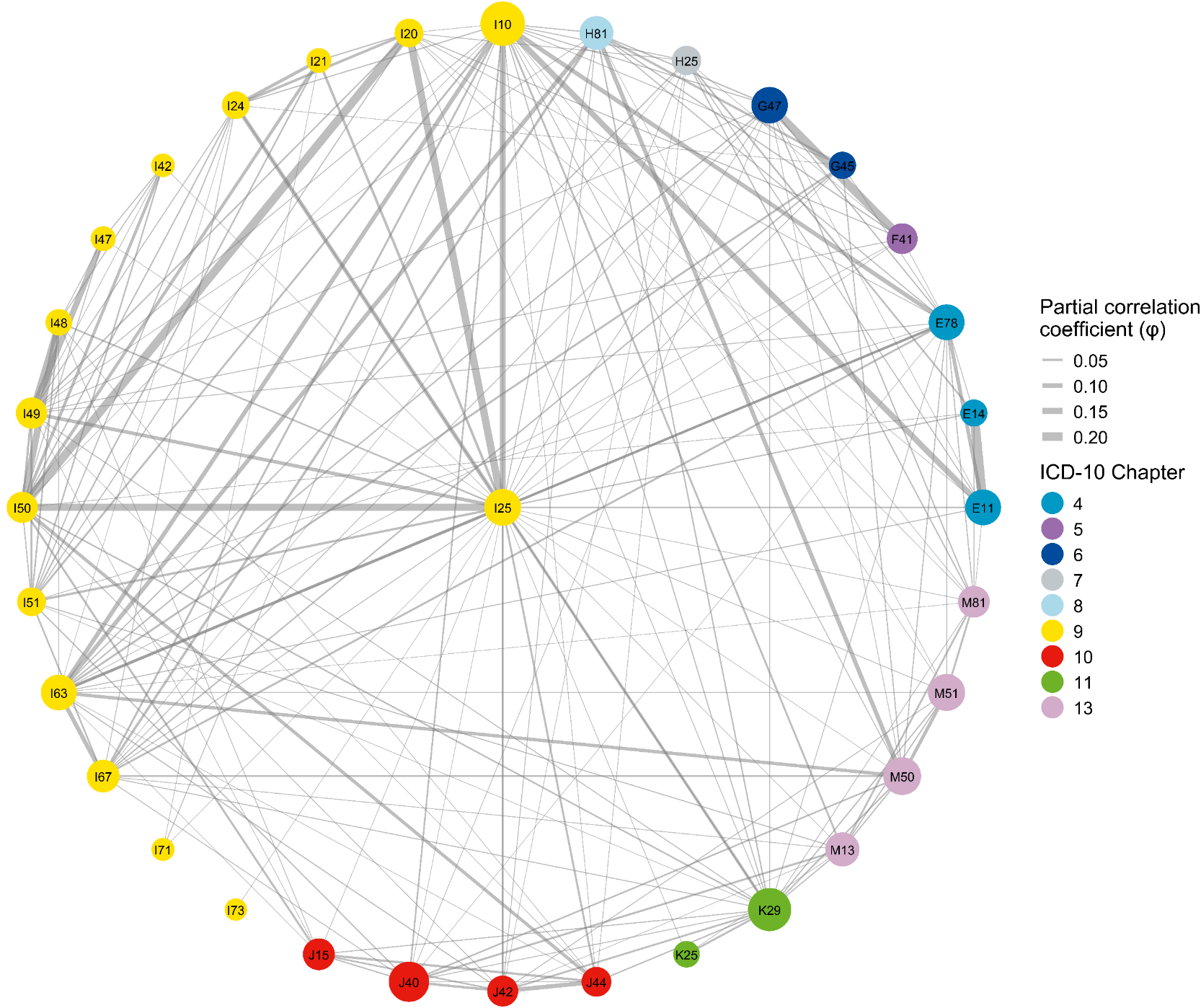

Supplementary Figure S9. Multimorbidity subnetworks for the top 20 hub diseases in male population identified by Multimorbidity Coefficient (MMC).

(A-T) Each panel displays one of the top 20 hub diseases (center node) and its neighboring conditions using a star layout. Nodes represent individual ICD‑10 conditions (labeled with ICD-3 codes); node size is proportional to each condition’s period prevalence (2016–2023), and node color denotes the ICD‑10 chapter. Edges depict significant adjusted associations between pairs of conditions after controlling for all other diseases in the network (quantified by the partial correlation coefficient, φ). Only associations meeting FDR‑adjusted *P* < 0.05 and φ > 0.01 are shown. Edge width is scaled to the magnitude of φ. Hubs are presented in descending order of MMC: (A) [K29], (B) [E88], (C) [I50], (D) [D64], (E) [J15], (F) [I70], (G) [E87], (H) [L30], (I) [K76], (J) [I63], (K) [N40], (L) [J44], (M) [G47], (N) [I25], (O) [E78], (P) [J31], (Q) [I69], (R) [J96], (S) [I10], (T) [N18]. These subnetworks can be explored interactively at: https://pumc-multimorbidity.shinyapps.io/hub-disease-network/

S8A

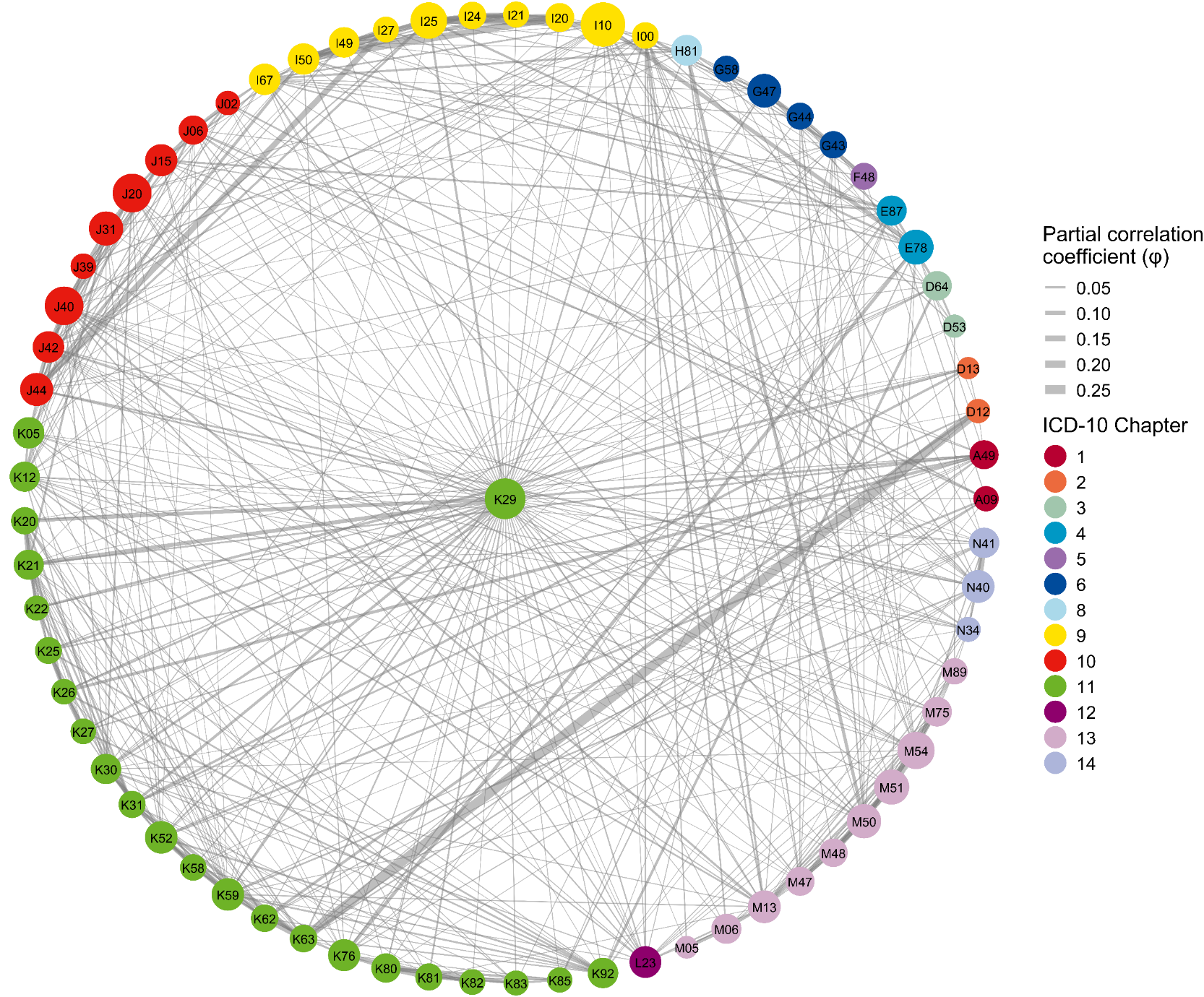

S8B

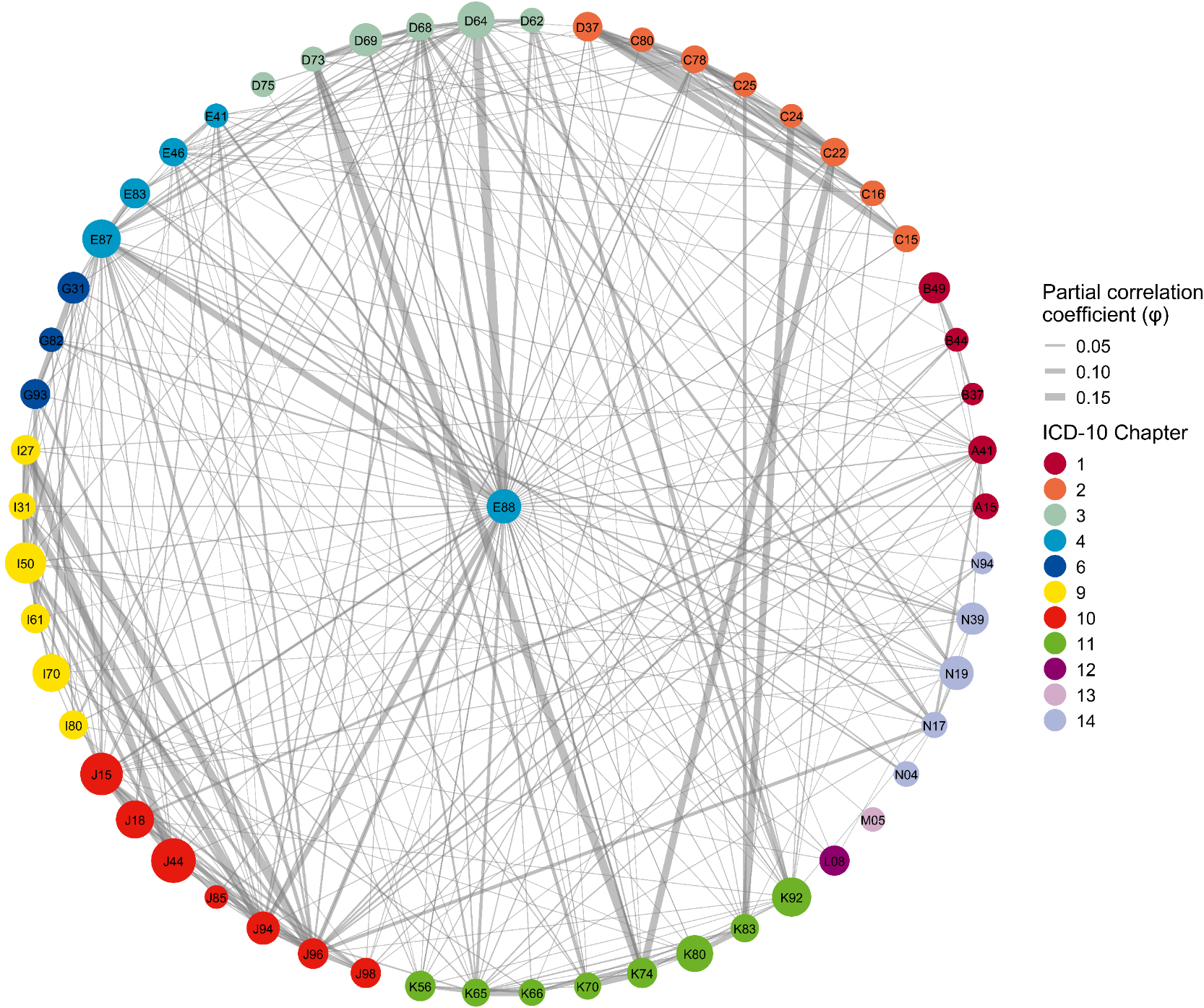

S8C

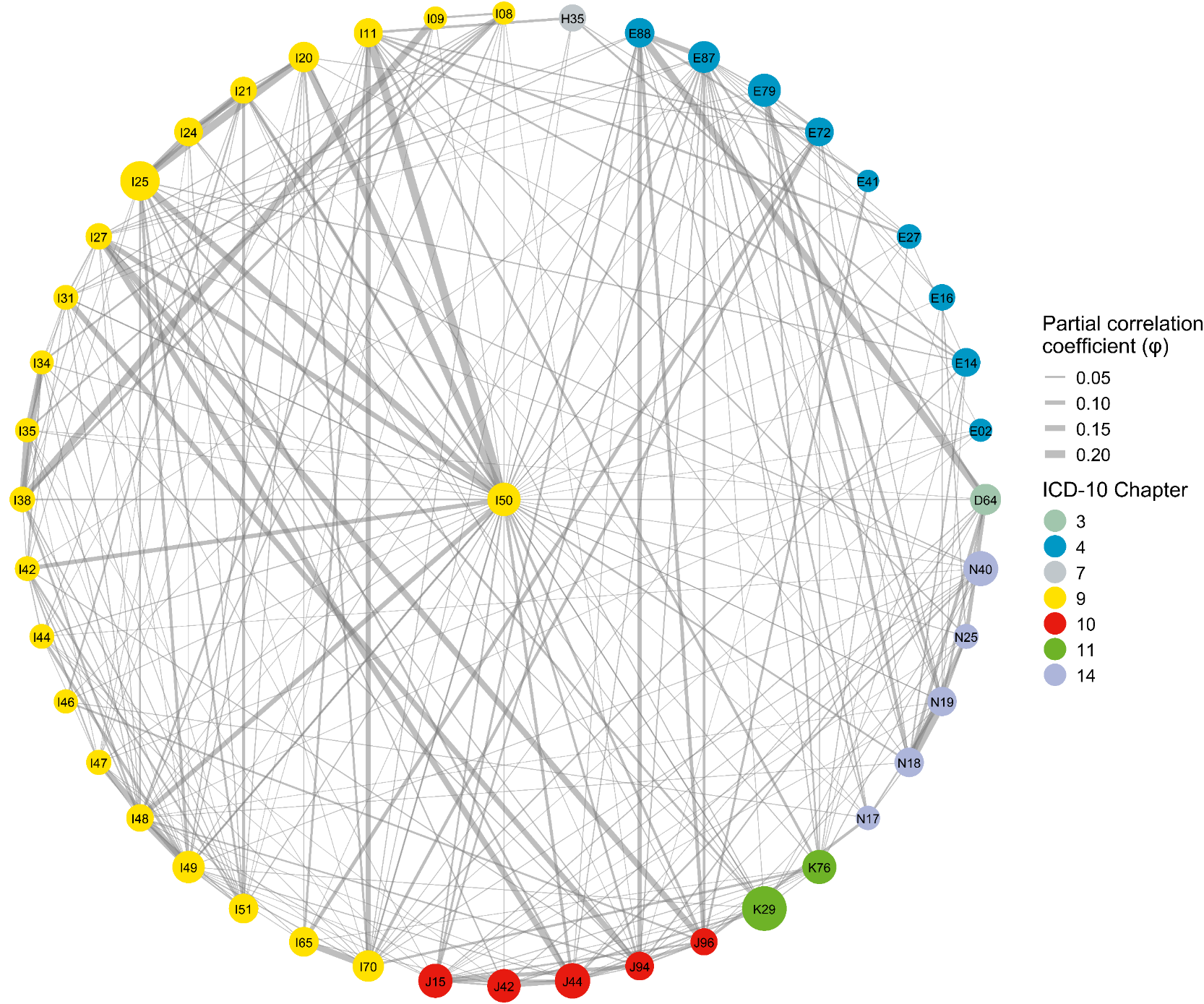

S8D

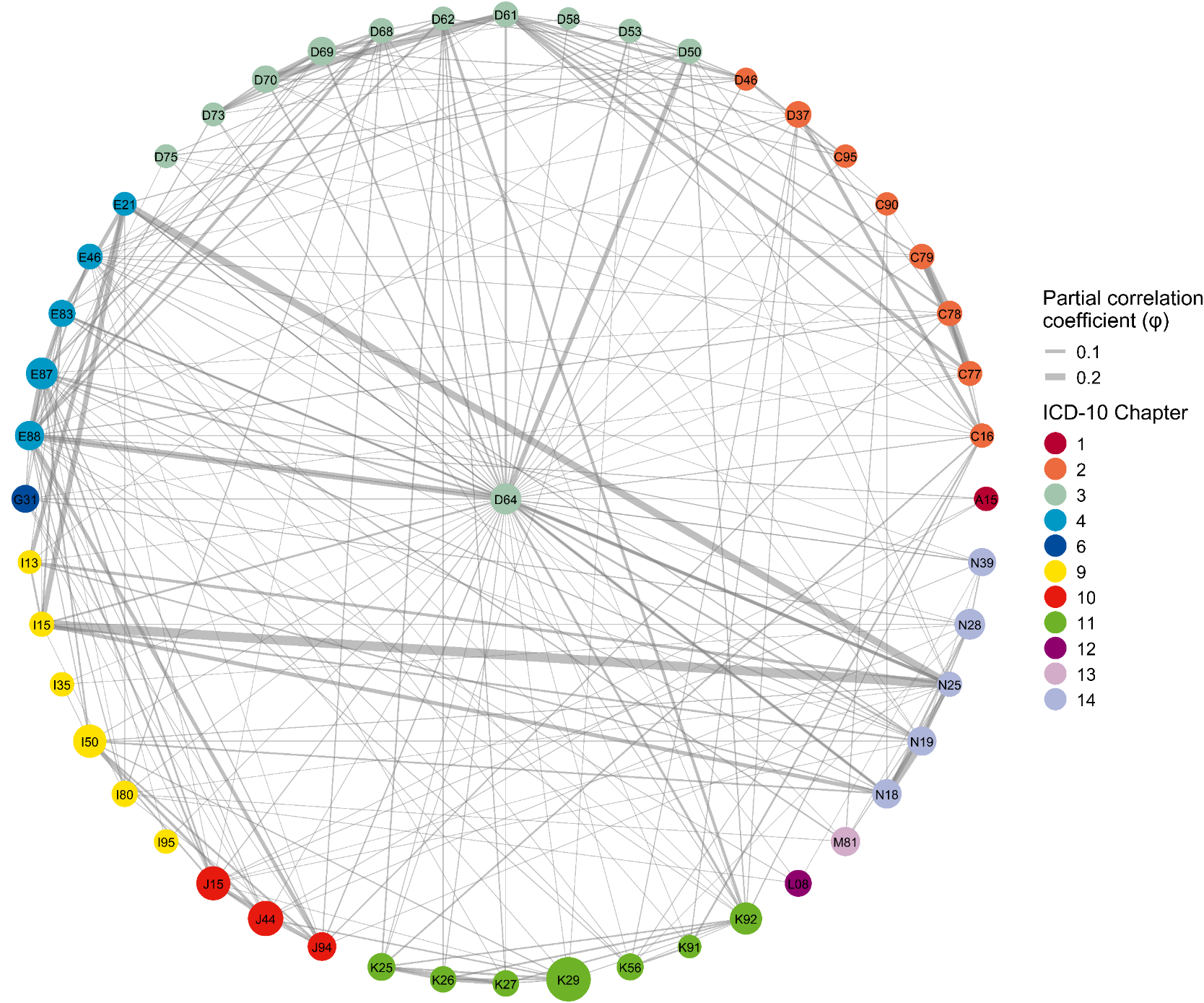

S8E

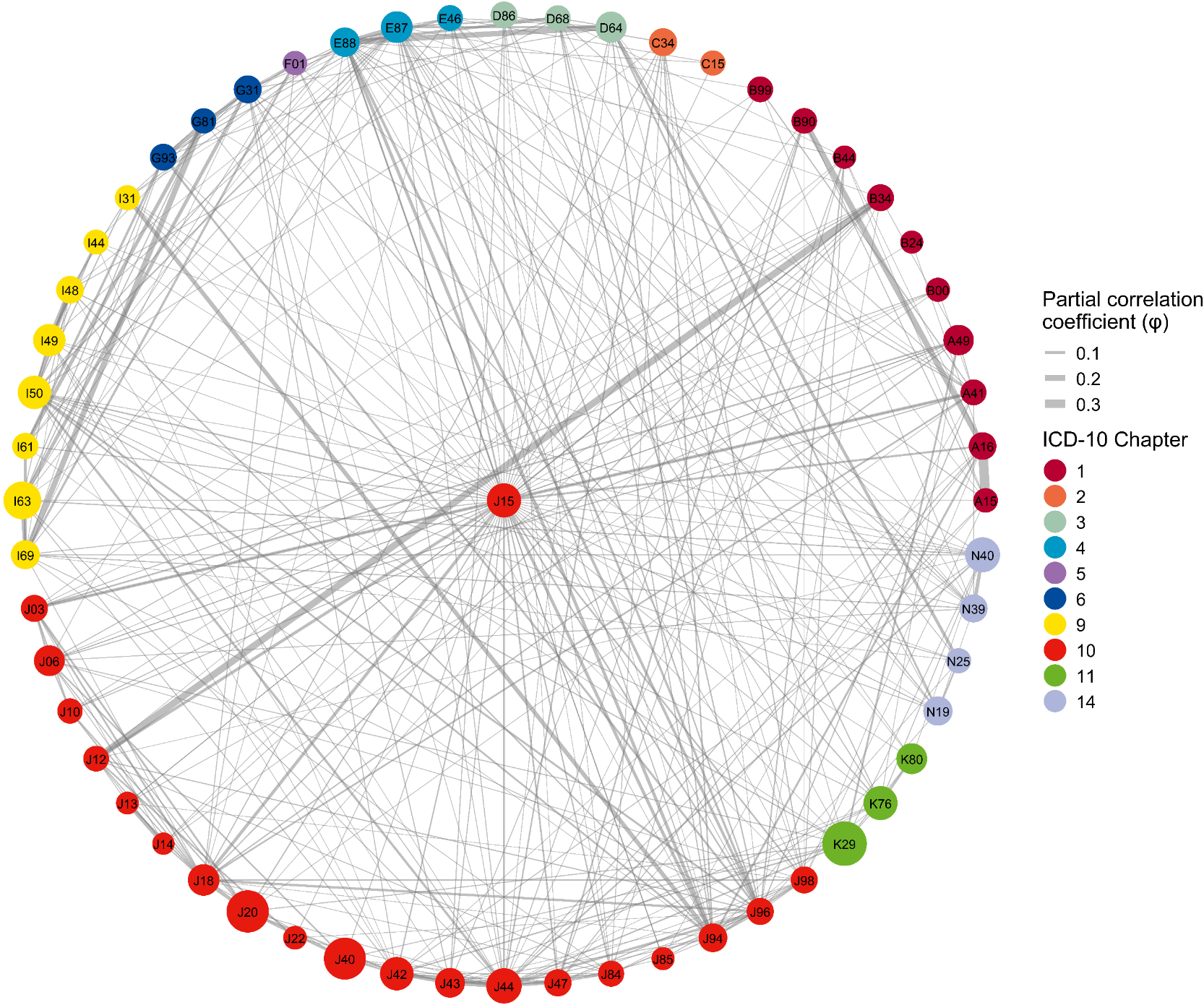

S8F

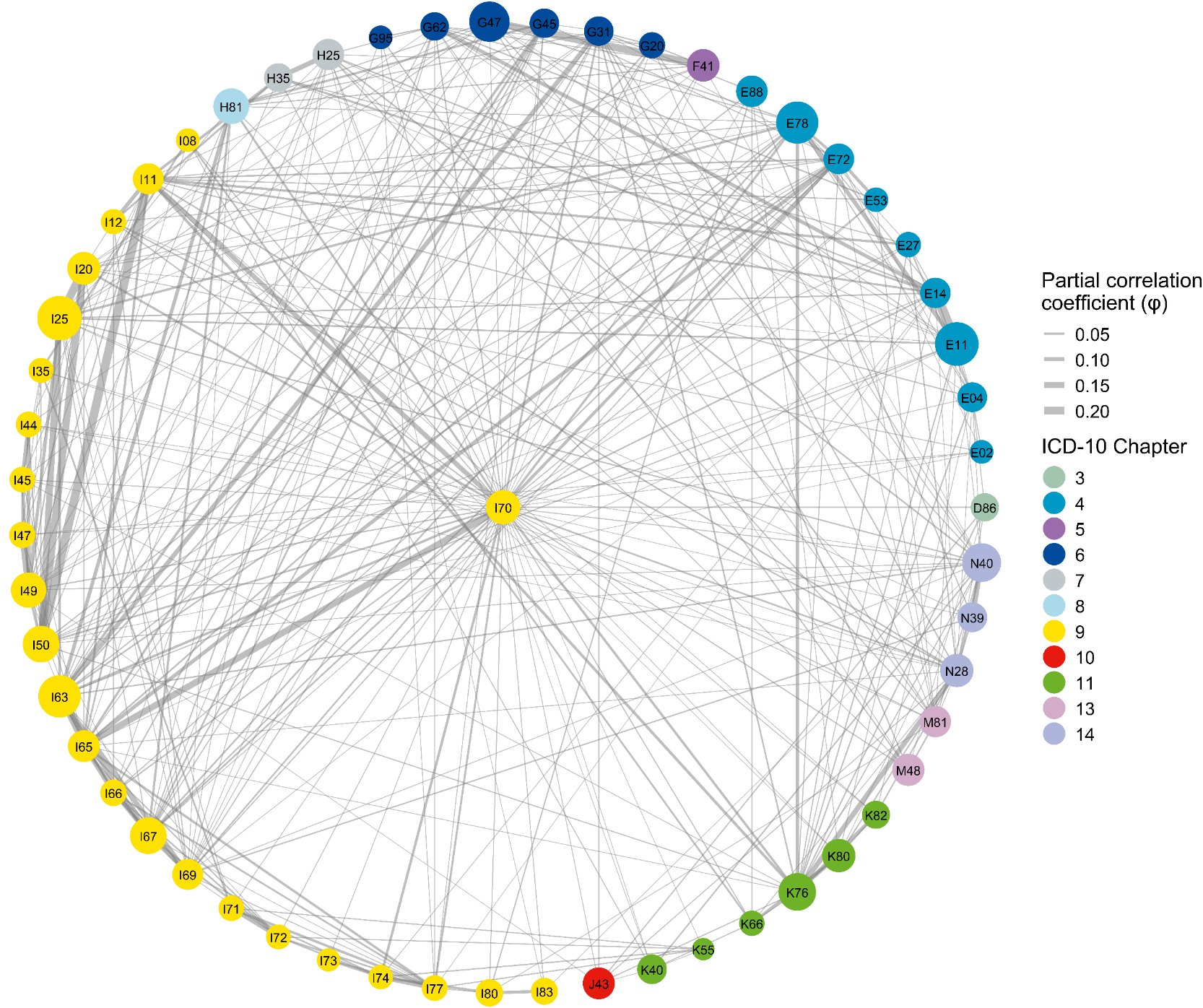

S8G

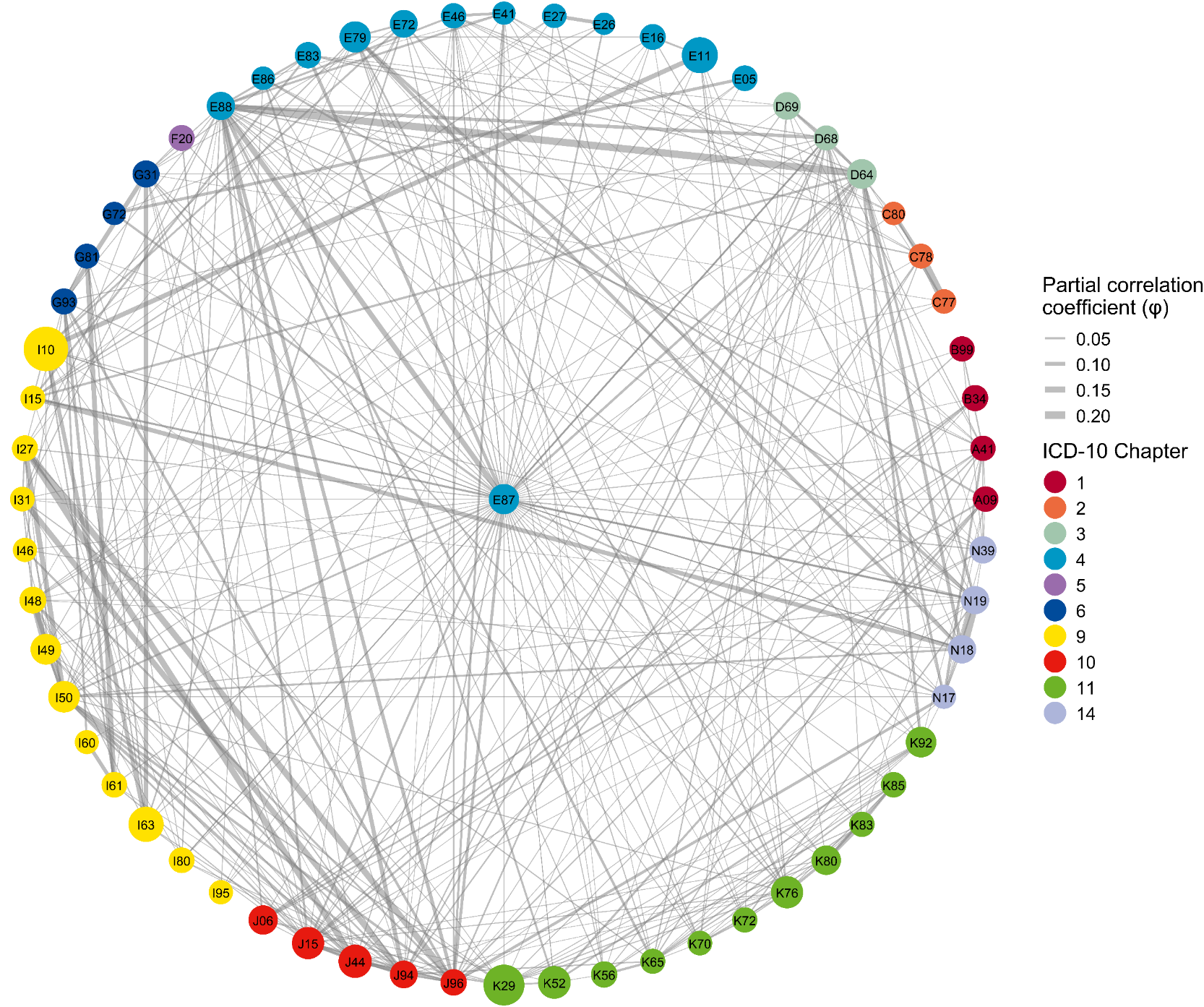

S8H

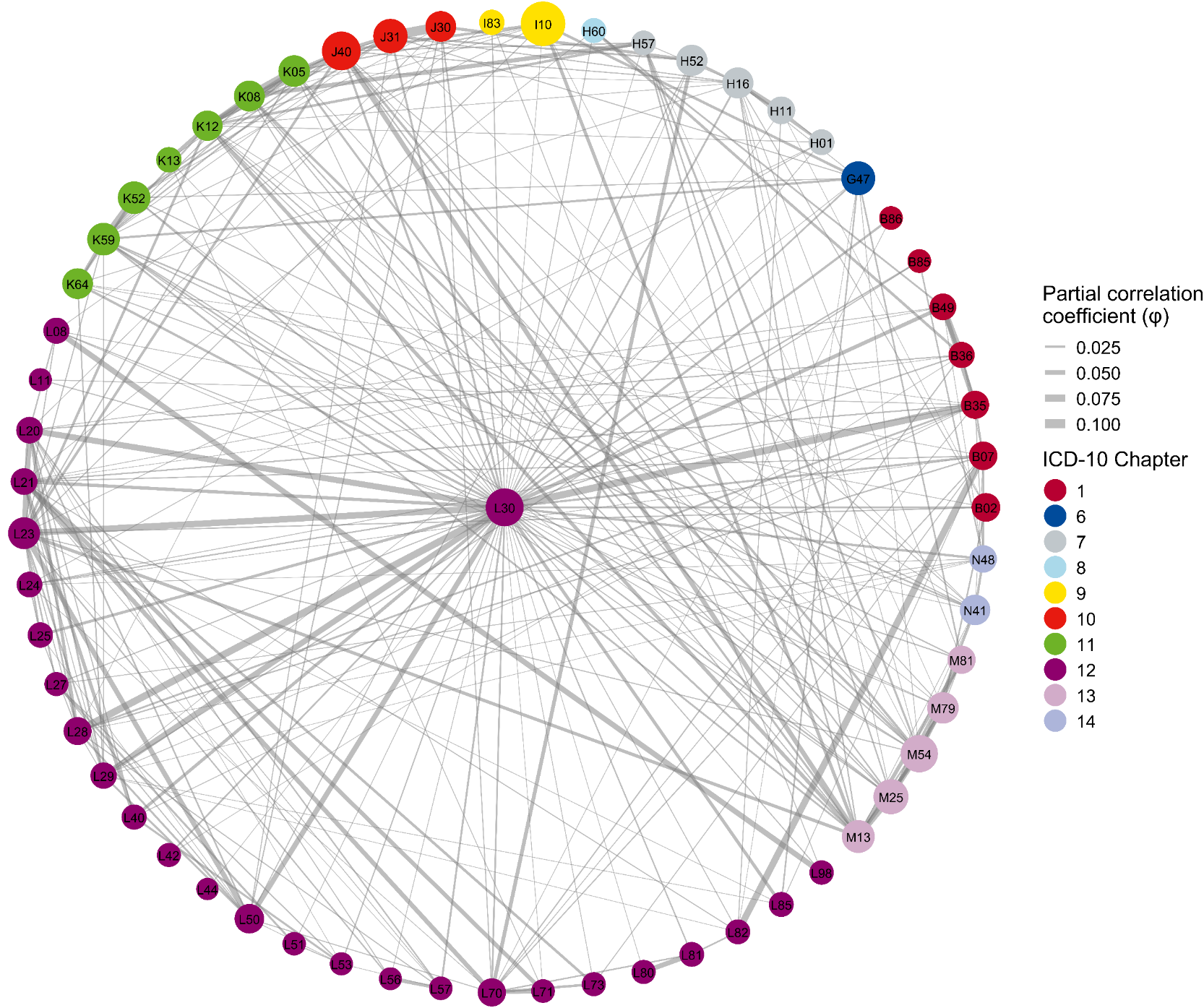

S8I

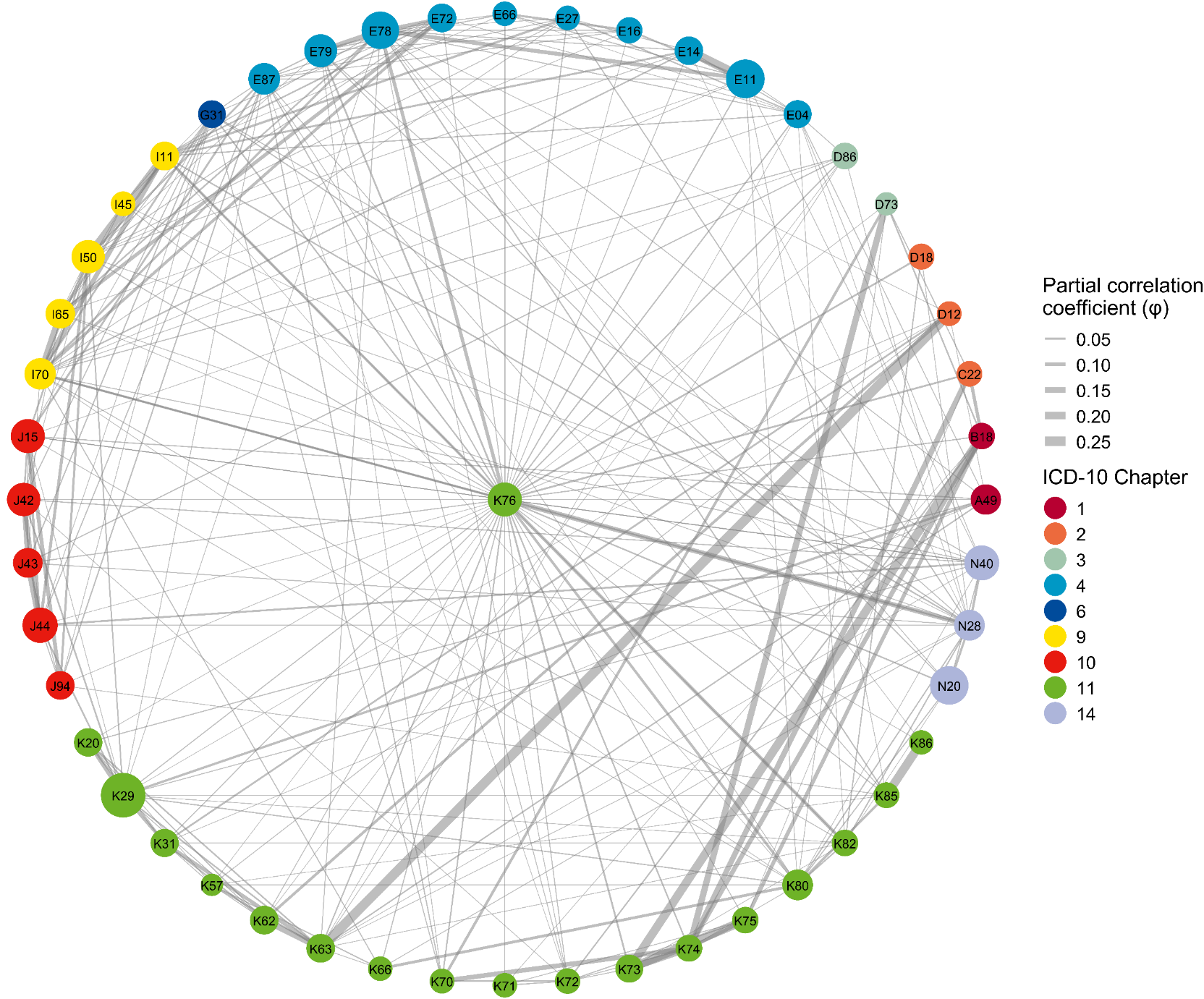

S8J

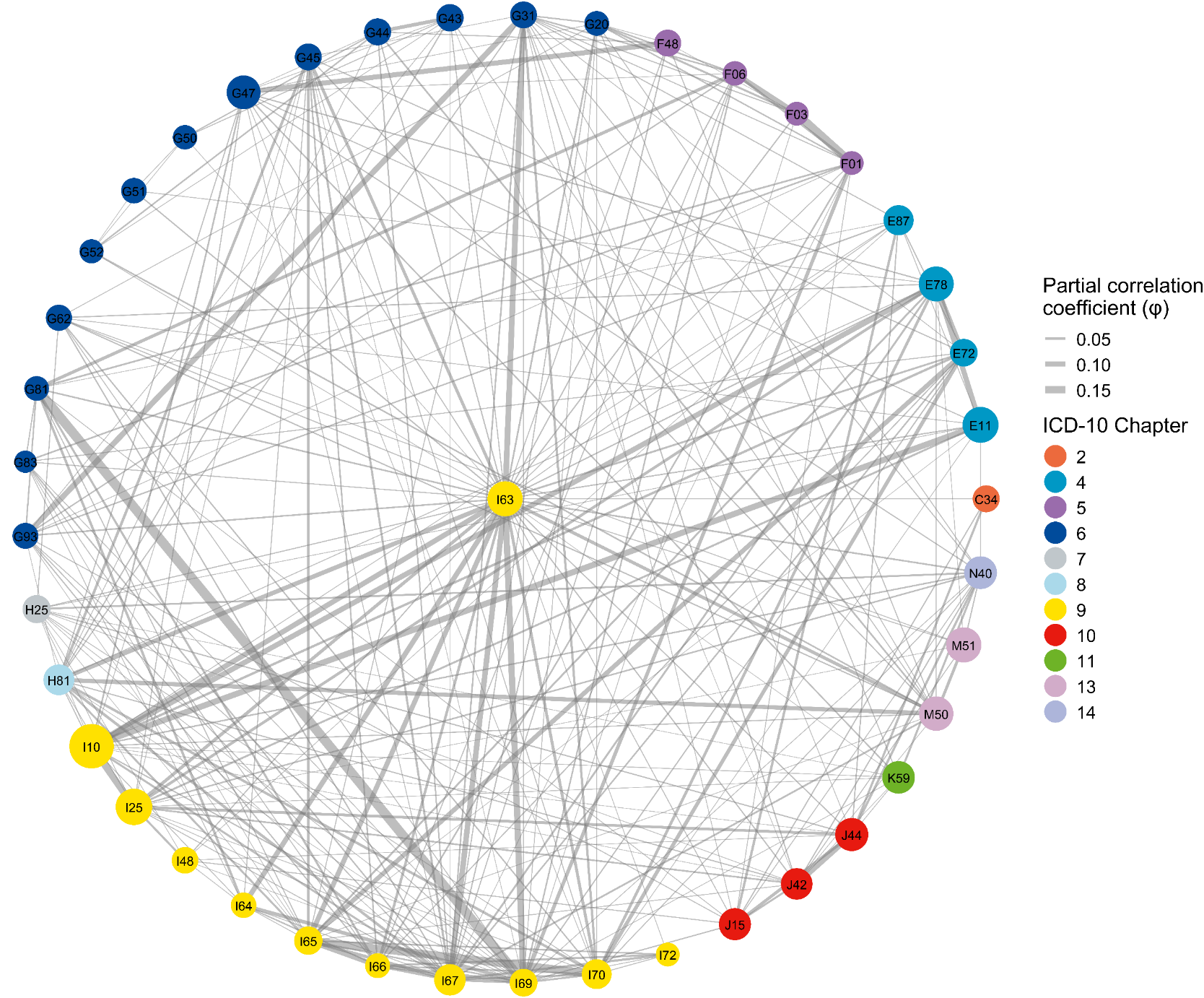

S8K

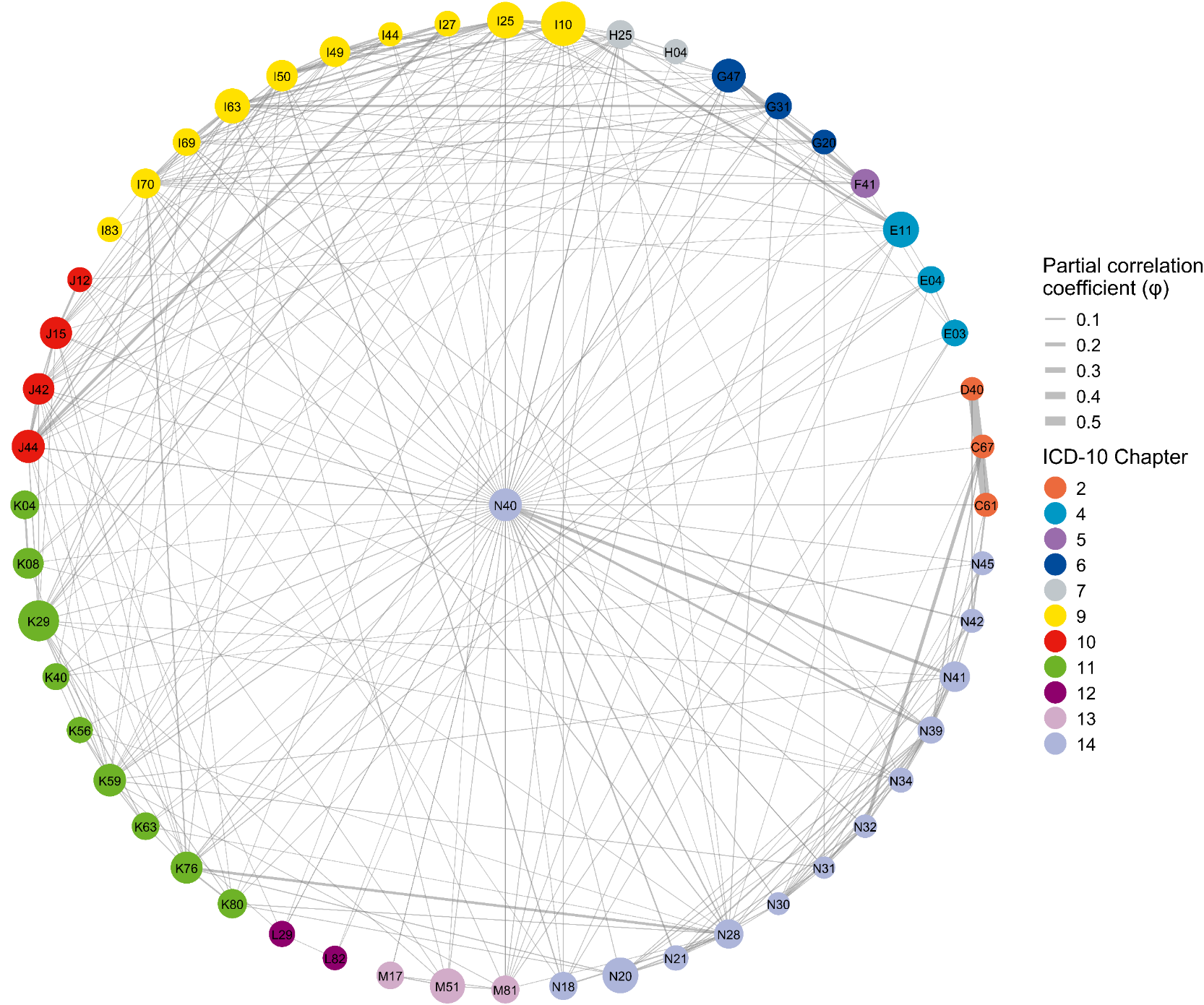

S8L

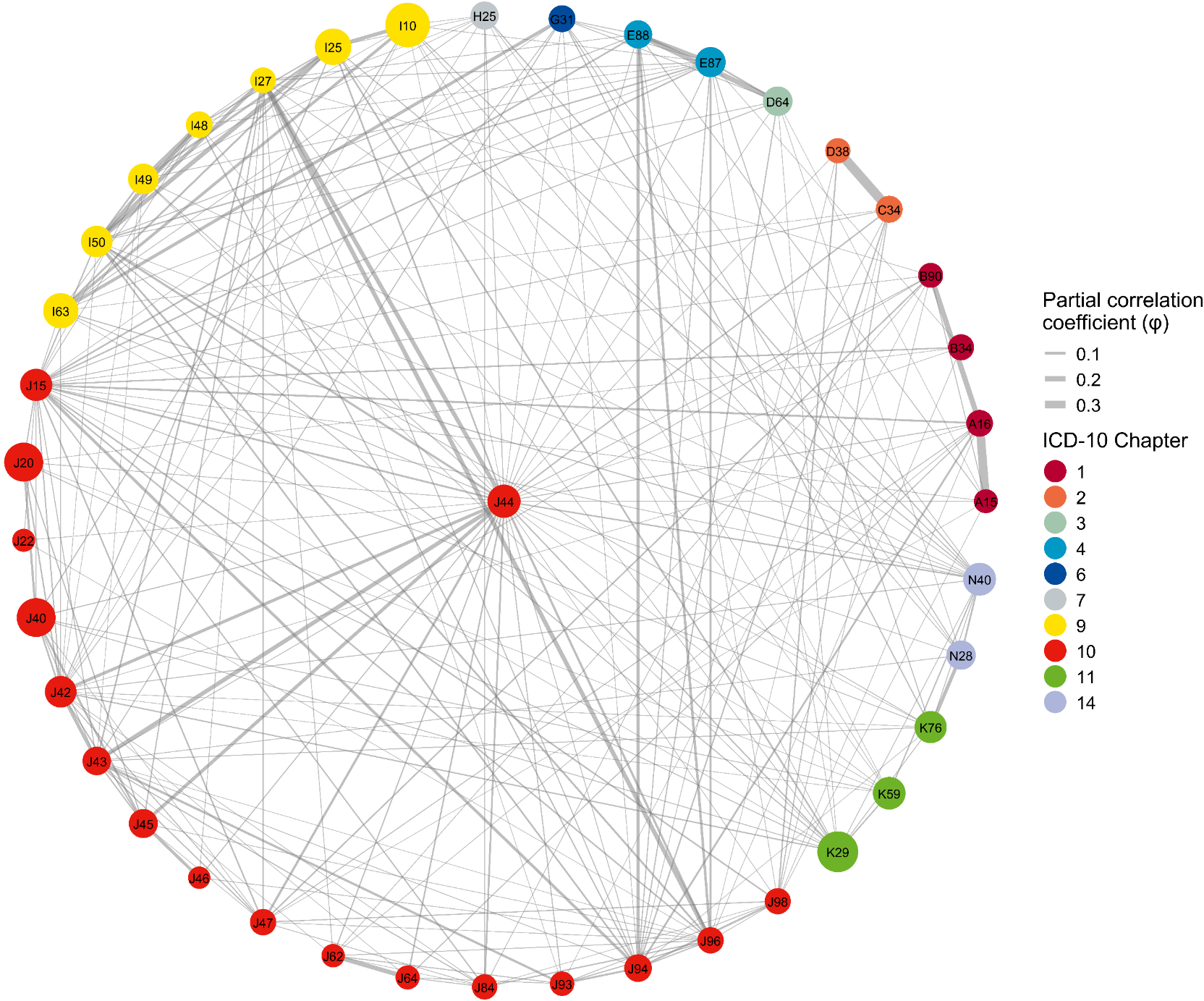

S8M

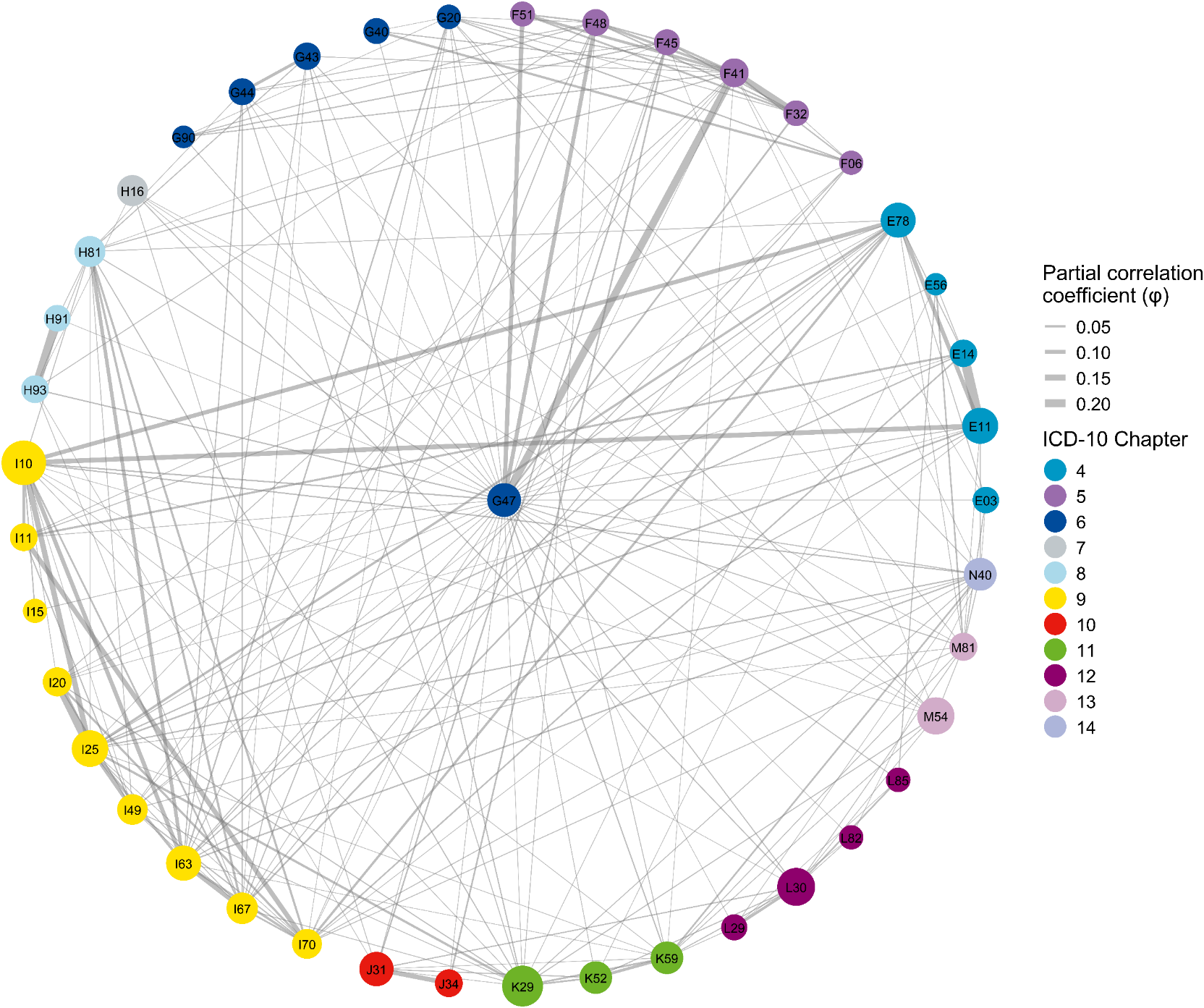

S8N

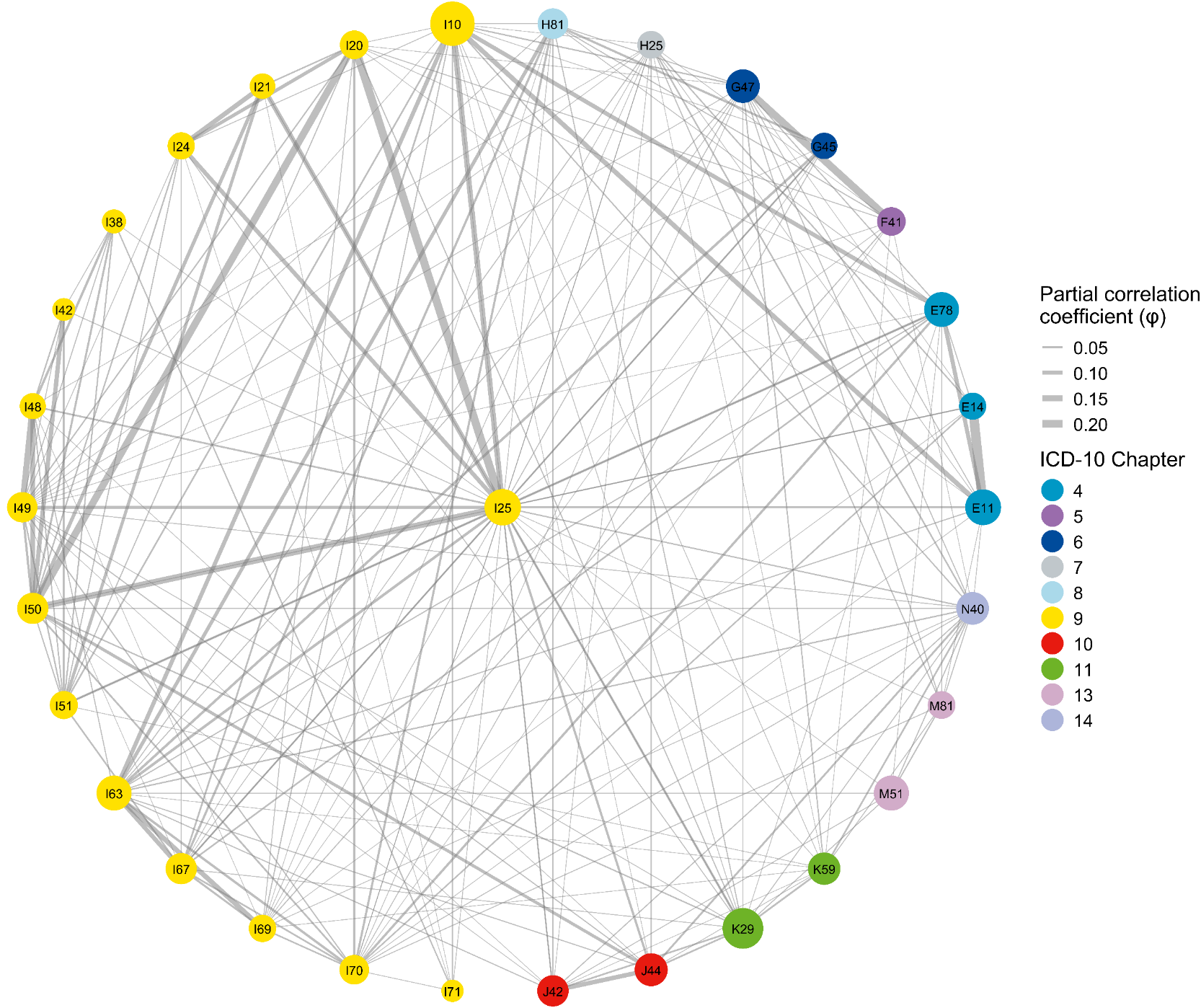

S8O

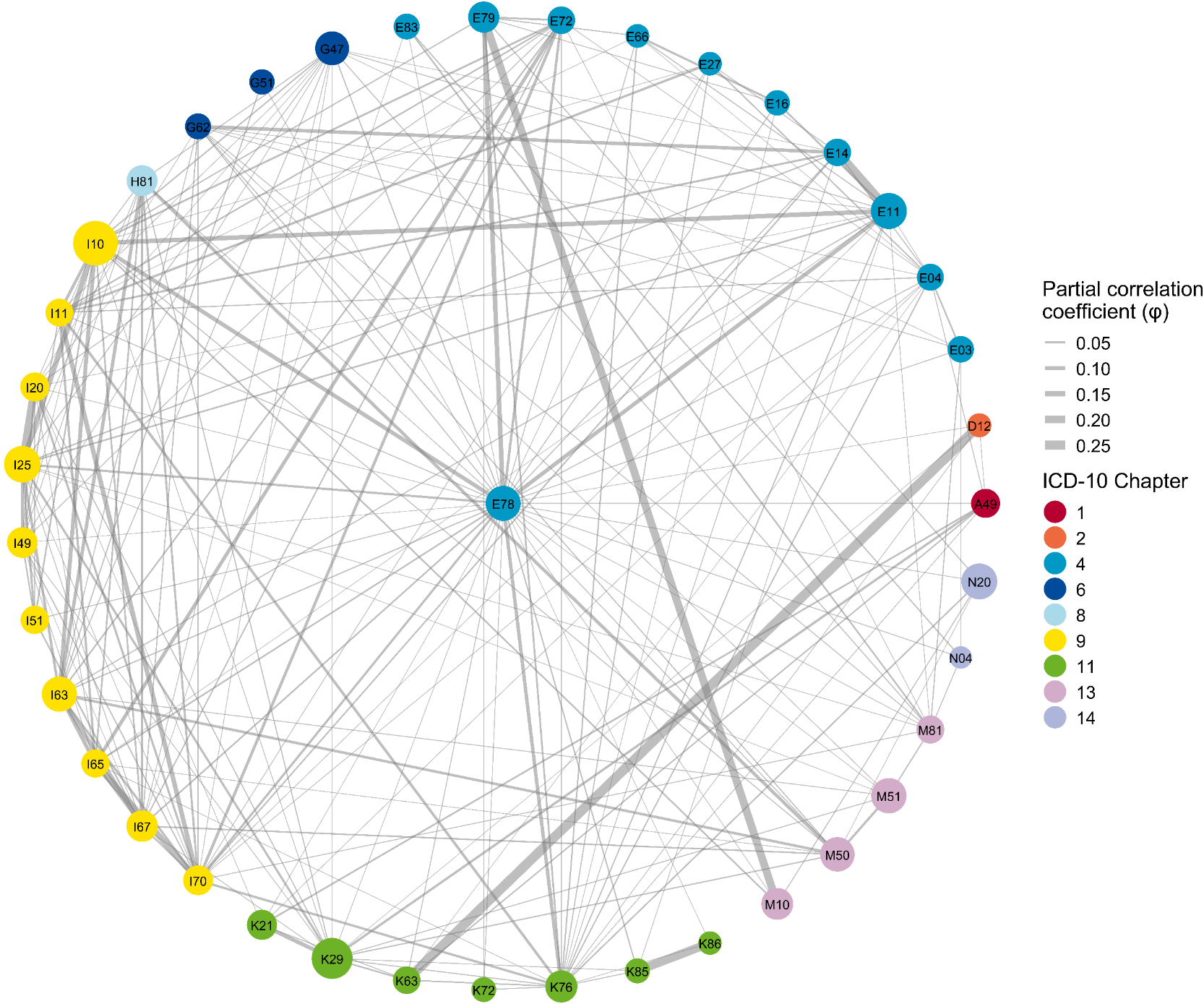

S8P

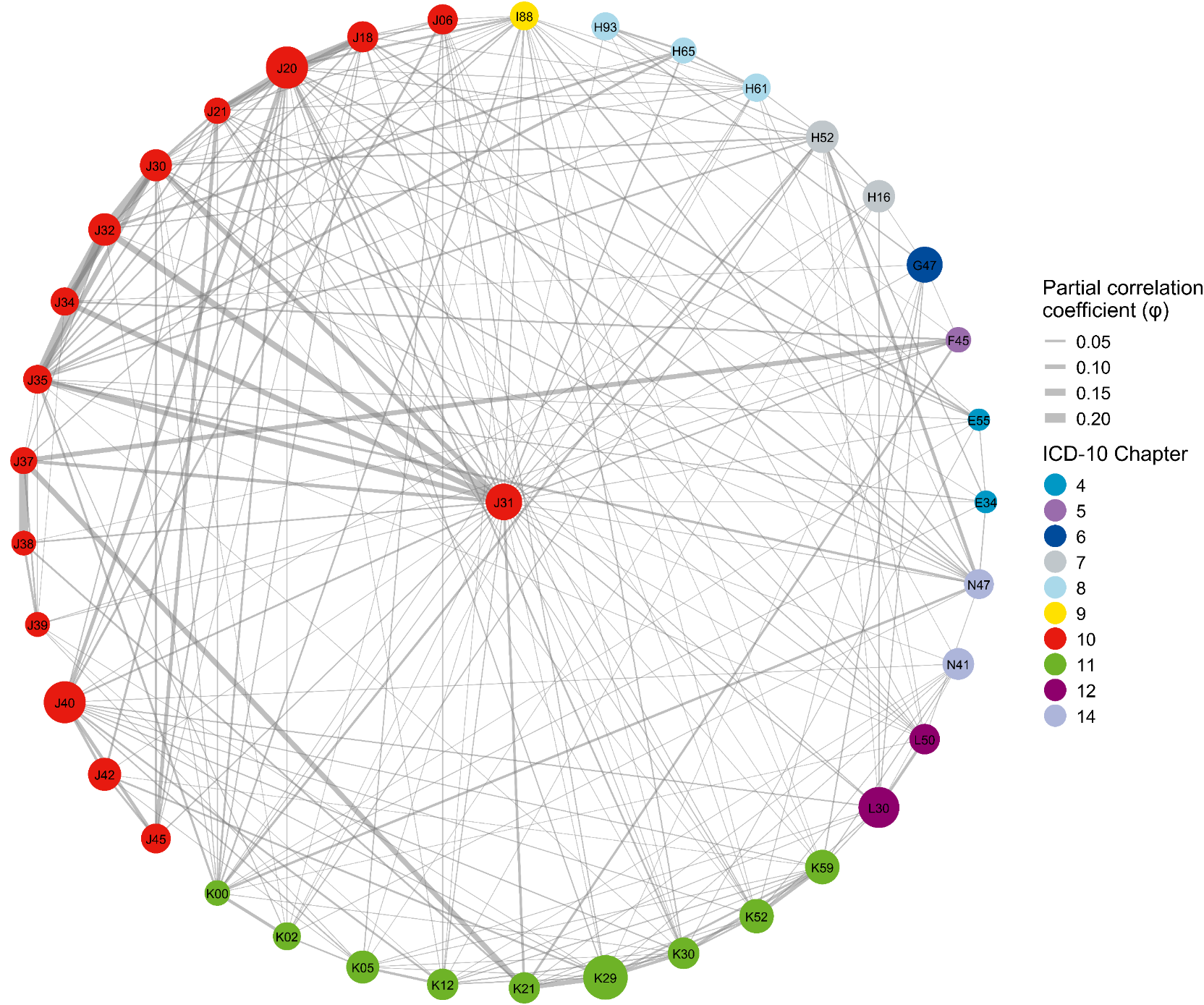

S8Q

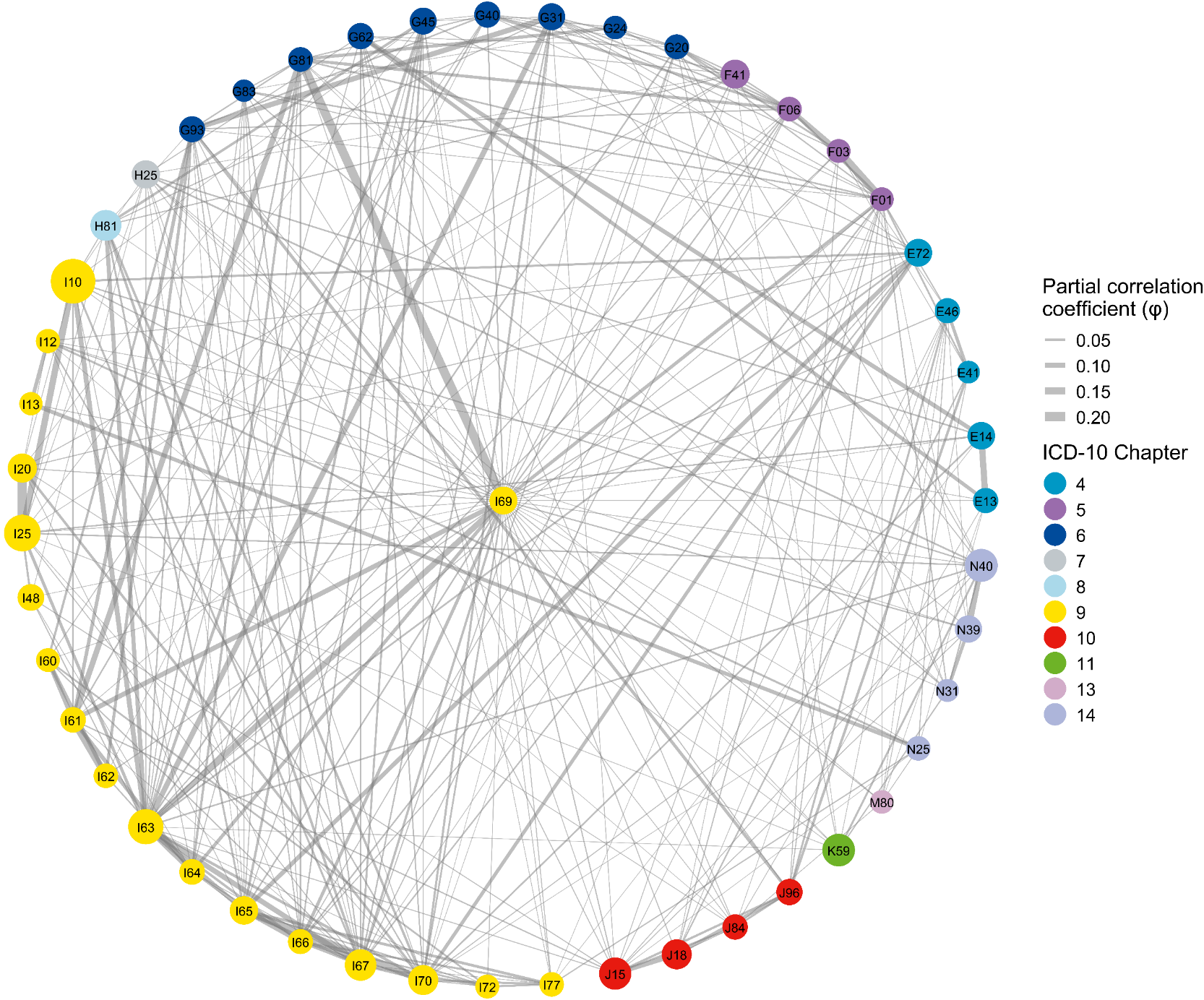

S8R

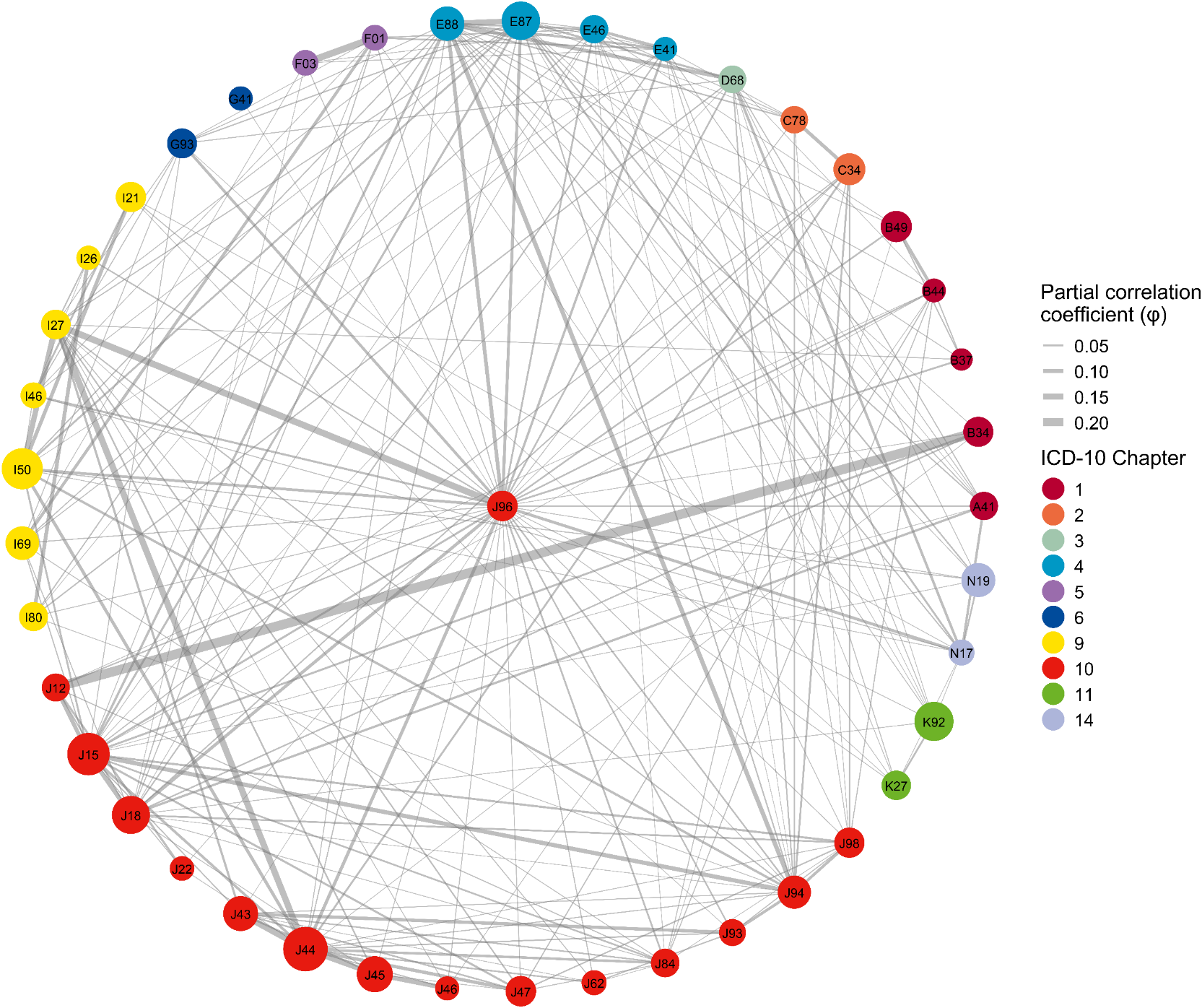

S8S

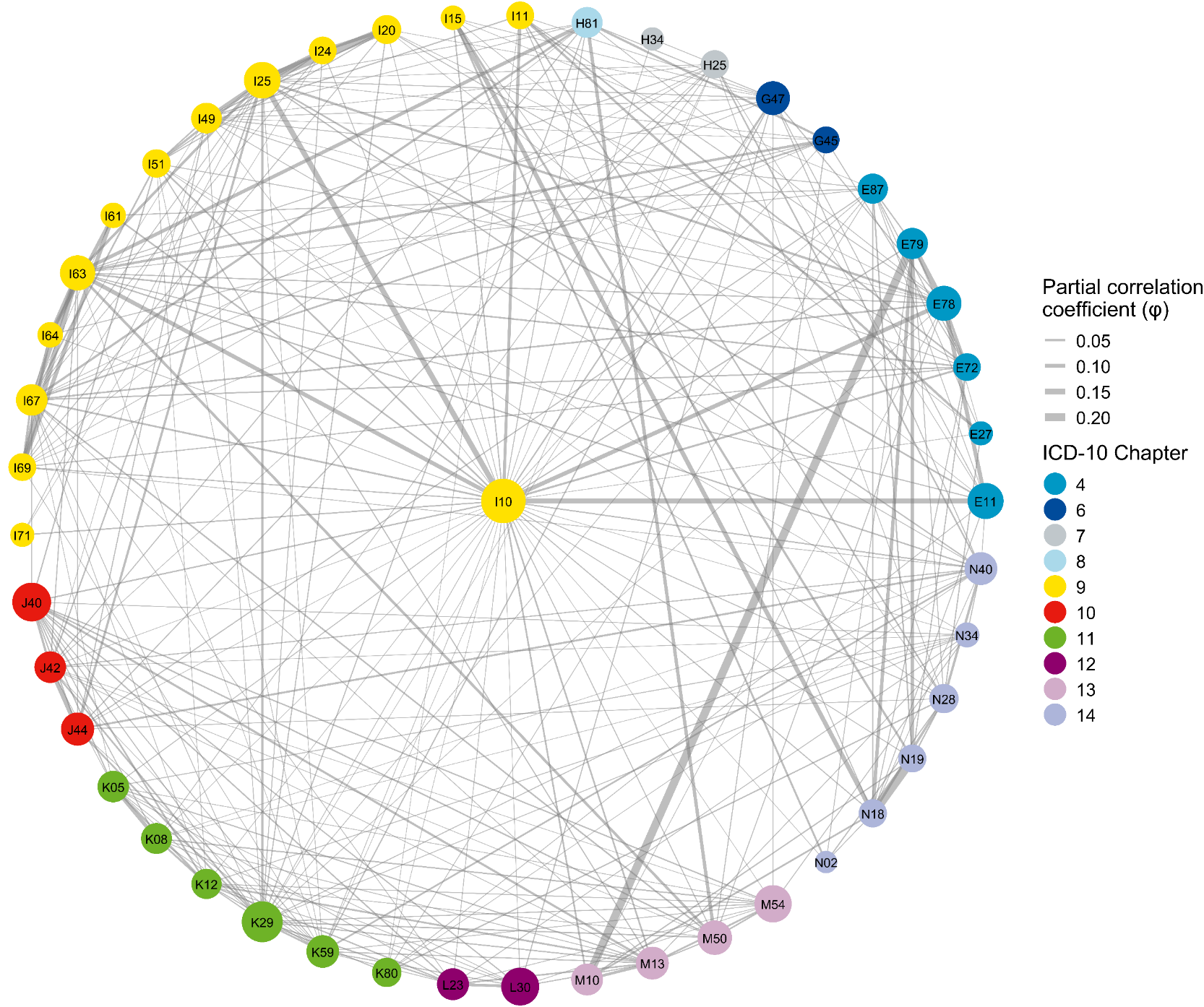

S8T

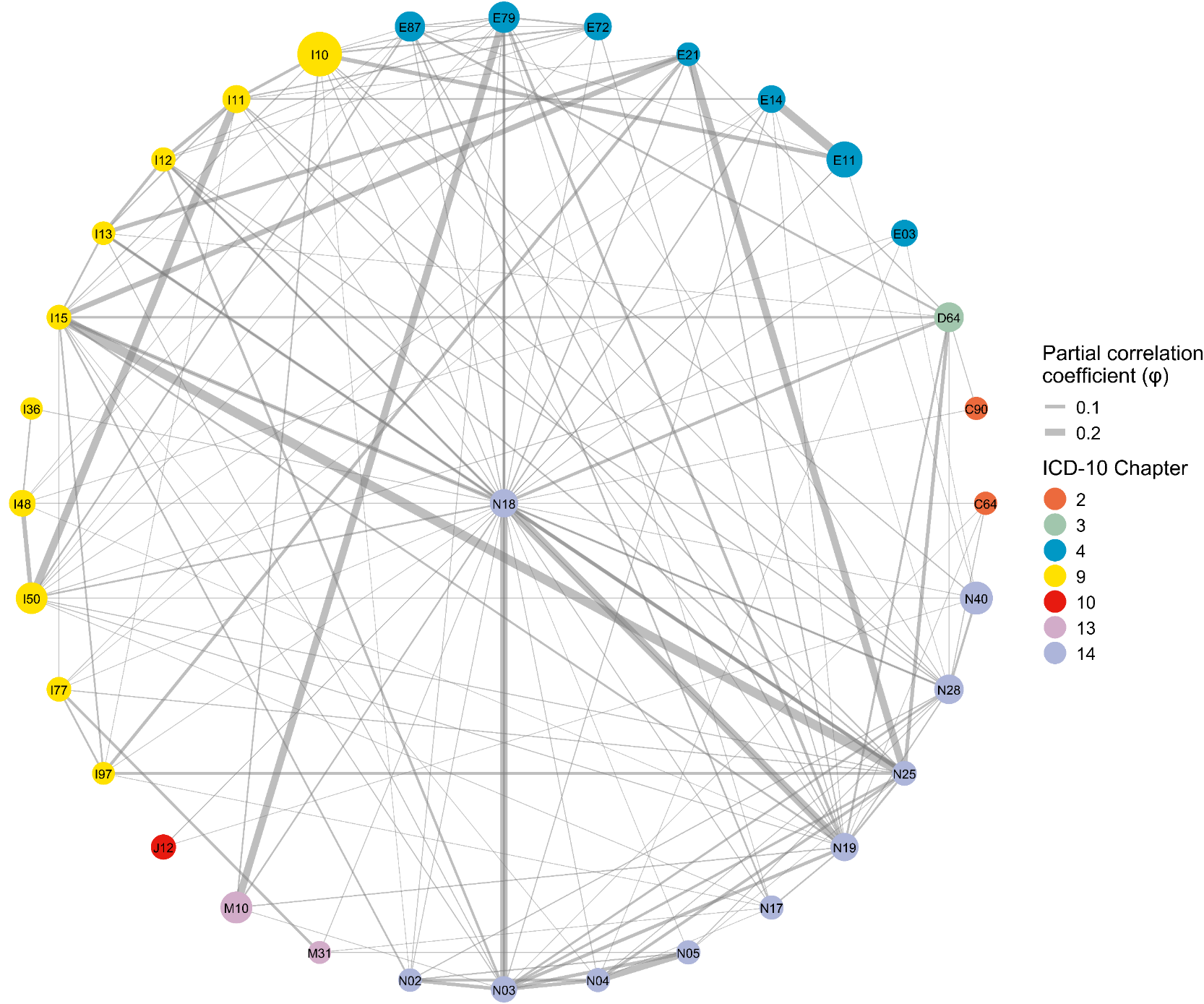

Supplementary Table S25. Descriptions of the top 50 most prevalent ICD-10 codes appearing in study figures and tables.

| Code | Prevalence | Description | Chapter | Chapter description |
| --- | --- | --- | --- | --- |
| I10 | 0.2007 | Essential (primary) hypertension | IX | Diseases of the circulatory system |
| K29 | 0.1588 | Gastritis and duodenitis | XI | Diseases of the digestive system |
| J20 | 0.1217 | Acute bronchitis | X | Diseases of the respiratory system |
| J40 | 0.1210 | Bronchitis, not specified as acute or chronic | X | Diseases of the respiratory system |
| L30 | 0.1137 | Other dermatitis | XII | Diseases of the skin and subcutaneous tissue |
| M54 | 0.0995 | Dorsalgia | XIII | Diseases of the musculoskeletal system and connective tissue |
| I25 | 0.0847 | Chronic ischaemic heart disease | IX | Diseases of the circulatory system |
| M50 | 0.0791 | Cervical disc disorders | XIII | Diseases of the musculoskeletal system and connective tissue |
| M51 | 0.0781 | Other intervertebral disc disorders | XIII | Diseases of the musculoskeletal system and connective tissue |
| E11 | 0.0762 | Type 2 diabetes mellitus | IV | Endocrine, nutritional and metabolic diseases |
| M25 | 0.0752 | Other joint disorders, not elsewhere classified | XIII | Diseases of the musculoskeletal system and connective tissue |
| I63 | 0.0714 | Cerebral infarction | IX | Diseases of the circulatory system |
| E78 | 0.0707 | Disorders of lipoprotein metabolism and other lipidaemias | IV | Endocrine, nutritional and metabolic diseases |
| G47 | 0.0685 | Sleep disorders | VI | Diseases of the nervous system |
| J31 | 0.0642 | Chronic rhinitis, nasopharyngitis and pharyngitis | X | Diseases of the respiratory system |
| N20 | 0.0575 | Calculus of kidney and ureter | XIV | Diseases of the genitourinary system |
| M13 | 0.0514 | Other arthritis | XIII | Diseases of the musculoskeletal system and connective tissue |
| K59 | 0.0478 | Other functional intestinal disorders | XI | Diseases of the digestive system |
| L23 | 0.0451 | Allergic contact dermatitis | XII | Diseases of the skin and subcutaneous tissue |
| K52 | 0.0442 | Other noninfective gastroenteritis and colitis | XI | Diseases of the digestive system |
| N92 | 0.0436 | Excessive, frequent and irregular menstruation | XIV | Diseases of the genitourinary system |
| H81 | 0.0423 | Disorders of vestibular function | VIII | Diseases of the ear and mastoid process |
| H16 | 0.0397 | Keratitis | VII | Diseases of the eye and adnexa |
| I67 | 0.0395 | Other cerebrovascular diseases | IX | Diseases of the circulatory system |
| J15 | 0.0389 | Bacterial pneumonia, not elsewhere classified | X | Diseases of the respiratory system |
| M79 | 0.0374 | Other soft tissue disorders, not elsewhere classified | XIII | Diseases of the musculoskeletal system and connective tissue |
| J44 | 0.0362 | Other chronic obstructive pulmonary disease | X | Diseases of the respiratory system |
| K76 | 0.0358 | Other diseases of liver | XI | Diseases of the digestive system |
| K05 | 0.0358 | Gingivitis and periodontal diseases | XI | Diseases of the digestive system |
| H52 | 0.0355 | Disorders of refraction and accommodation | VII | Diseases of the eye and adnexa |
| N93 | 0.0341 | Other abnormal uterine and vaginal bleeding | XIV | Diseases of the genitourinary system |
| I50 | 0.0337 | Heart failure | IX | Diseases of the circulatory system |
| D64 | 0.0336 | Other anaemias | III | Diseases of the blood and blood-forming organs and certain disorders involving the immune mechanism |
| J42 | 0.0333 | Unspecified chronic bronchitis | X | Diseases of the respiratory system |
| K08 | 0.0332 | Other disorders of teeth and supporting structures | XI | Diseases of the digestive system |
| J32 | 0.032 | Chronic sinusitis | X | Diseases of the respiratory system |
| I49 | 0.0311 | Other cardiac arrhythmias | IX | Diseases of the circulatory system |
| J30 | 0.0299 | Vasomotor and allergic rhinitis | X | Diseases of the respiratory system |
| K64 | 0.0294 | Haemorrhoids and perianal venous thrombosis | XI | Diseases of the digestive system |
| K30 | 0.0294 | Functional dyspepsia | XI | Diseases of the digestive system |
| N62 | 0.0292 | Hypertrophy of breast | XIV | Diseases of the genitourinary system |
| K21 | 0.0291 | Gastro-oesophageal reflux disease | XI | Diseases of the digestive system |
| K12 | 0.0288 | Stomatitis and related lesions | XI | Diseases of the digestive system |
| M06 | 0.0278 | Other rheumatoid arthritis | XIII | Diseases of the musculoskeletal system and connective tissue |
| L50 | 0.0269 | Urticaria | XII | Diseases of the skin and subcutaneous tissue |
| E87 | 0.0267 | Other disorders of fluid, electrolyte and acid-base balance | IV | Endocrine, nutritional and metabolic diseases |
| K92 | 0.0264 | Other diseases of digestive system | XI | Diseases of the digestive system |
| N72 | 0.0262 | Inflammatory disease of cervix uteri | XIV | Diseases of the genitourinary system |
| M75 | 0.0255 | Shoulder lesions | XIII | Diseases of the musculoskeletal system and connective tissue |
| K80 | 0.0251 | Cholelithiasis | XI | Diseases of the digestive system |

Note: The table lists the top 50 most prevalent three-character ICD-10 codes that appear in the manuscript's figures, along with their full descriptions and chapter/block information. The complete list of all codes used in the study is available for download at our GitHub repository: https://github.com/PUMCWh/ICD-10-info/blob/main/icd10_code_list_full.xlsx
